# GA4GH Phenopacket-Driven Characterization of Genotype-Phenotype Correlations in Mendelian Disorders

**DOI:** 10.1101/2025.03.05.25323315

**Authors:** Lauren Rekerle, Daniel Danis, Filip Rehburg, Adam SL Graefe, Viktor Bily, Andrés Caballero-Oteyza, Pilar Cacheiro, Leonardo Chimirri, Jessica X Chong, Evan Connelly, Bert BA de Vries, Alexander JM Dingemans, Michael H Duyzend, Tomas Freiberger, Petra Gehle, Tudor Groza, Peter Hansen, Julius O.B. Jacobsen, Adam Klocperk, Markus S Ladewig, Michael I Love, Allison J Marcello, Alexander Mordhorst, Monica C Munoz-Torres, Justin Reese, Catharina Schütz, Damian Smedley, Timmy Strauss, Ondrej Vladyka, David Zocche, Sylvia Thun, Christopher J Mungall, Melissa A Haendel, Peter N Robinson

## Abstract

Comprehensively characterizing genotype-phenotype correlations (GPCs) in Mendelian disease would create new opportunities for improving clinical management and understanding disease biology. However, heterogeneous approaches to data sharing, reuse, and analysis have hindered progress in the field. We developed Genotype Phenotype Evaluation of Statistical Association (GPSEA), a software package that leverages the Global Alliance for Genomics and Health (GA4GH) Phenopacket Schema to represent case-level clinical and genetic data about individuals. GPSEA applies an independent filtering strategy to boost statistical power to detect categorical GPCs represented by Human Phenotype Ontology terms. GPSEA additionally enables visualization and analysis of continuous phenotypes, clinical severity scores, and survival data such as age of onset of disease or clinical manifestations. We applied GPSEA to 85 cohorts with 6613 previously published individuals with variants in one of 80 genes associated with 122 Mendelian diseases and identified 225 significant GPCs, with 48 cohorts having at least one statistically significant GPC. These results highlight the power of standardized representations of clinical data for scalable discovery of GPCs in Mendelian disease.

## Introduction

There are a huge number of clinical manifestations of human disease, and even individuals with the same clinical diagnosis may present with different combinations of phenotypic abnormalities, ages of onset of these abnormalities, and degrees of clinical severity. A key question for genomic precision medicine is how specific genetic variants influence clinical phenotype. The correlation between genotype (the type of variant or variants present at a given location) and phenotype (presence or absence of medically relevant observable traits) is defined as an above-chance probability of an association between the two, an association termed genotype-phenotype correlation (GPC).^1^ Commonly, even individuals with an identical pathogenic variant may display variable findings, so GPCs are rarely absolute. Instead, GPCs usually signify a higher frequency of a feature in the presence of a certain genotype, or an earlier age of onset of the disease or a disease feature, or in some cases earlier mortality. For instance, a specific in-frame deletion of codon 992 of the *NF1* gene is associated with a milder phenotype characterized by café-au-lait spots and skinfold freckling with the absence of cutaneous and visible plexiform neurofibromas, whereas individuals with missense mutations affecting any of the five codons 844–848 have a more severe phenotype characterized by a high prevalence of plexiform neurofibromas, optic pathway gliomas, malignant neoplasms, and skeletal abnormalities.^2^ A core paradigm in precision genomic medicine is to match therapeutic interventions and other forms of clinical care to the pathomechanism of disease and where appropriate to specific genetic variants. Although this paradigm has been extremely successful in oncology, where targeted therapies are applied to treat cancer types if a certain genetic variant is identified, this approach has been less successful in Mendelian disease.^3^ Historically, it has been difficult to perform GPCs for rare Mendelian diseases because the rarity of diseases implies it is generally difficult or impossible to recruit cohorts large enough to achieve statistical power.

The Human Phenotype Ontology (HPO) is a comprehensive bioinformatic resource for the analysis of human diseases and phenotypes, offering a computational bridge between genome biology and clinical medicine, and is used internationally for analysis and exchange of phenotype data in rare disease (RD) medicine.^4–6^ A growing number of published works leverage HPO-encoded data for GPC analysis.^7–21^ However, there are challenges to using HPO for GPC because of the need to propagate annotations up the hierarchy of the HPO and to include only explicitly observed or excluded HPO terms in categorical testing, both operations that are not natively supported by standard spreadsheet tools or bioinformatics packages. Additionally, it is desirable to integrate the analysis with other kinds of clinical data including numerical measurements, age of onset or mortality, and severity scoring. In addition, the community has lacked a common schema for representing individual (i.e. case-level) clinical trajectories with these and other attributes. Additionally, heterogeneous approaches to data sharing, reuse, and analysis have software packages and data repositories that support GPC analysis. The Global Alliance for Genomics and Health (GA4GH) is an organization developing a suite of coordinated standards for genomics. The GA4GH Phenopacket Schema is a standard for sharing disease and phenotype information characterizing an individual person or biosample that addresses the challenge of documenting case-level phenotypic information.^22–25^ Here, we present GPSEA (Genotype-Phenotype Statistical Evaluation of Associations). GPSEA leverages case-level phenopackets, characterizing an individual person or biosample and linking the individual to detailed phenotypic descriptions, genetic information, diagnoses, and treatments.^22^ GPSEA automates the process of visualizing and performing GPC analysis. We applied the software to 85 cohorts, 48 (56%) of which had at least one statistically significant GPC (there were a total of 225 statistically significant results). We show the power of utilizing case-level patient characterization and discuss future utility in differential diagnostics and precision medicine.

## Results

### Characterizing genotype-phenotype correlations with GPSEA

GPSEA is an end-to-end software framework for exploring and visualizing the cohorts and for characterizing GPCs. GPSEA is a Python package designed to be used in a Jupyter notebook, but the analysis functions can also be used as a programming library. The GPSEA framework enables testing of existing hypotheses about the disease or gene in question or generation of new hypotheses based on salient aspects of the investigated cohort depicted by the tables and visualizations. Four main classes of statistical tests are supported for categorical phenotypic traits (i.e., observed vs. excluded HPO terms or disease diagnoses), numerical values (e.g. laboratory test results), phenotype scores, and survival data (i.e., mortality, disease onset or onset of a specific HPO term). GPSEA formats the results into tables and figures suitable for processing in bioinformatic pipelines or interactive exploration within the Python data science environment (**Fig. 1**). We tested GPSEA on 85 cohorts, covering 80 genes and 122 diseases. We first explain the algorithmic approaches to setting up GPC testing and then present an overview of 225 significant correlations identified in the cohorts.

**Fig. 1:**
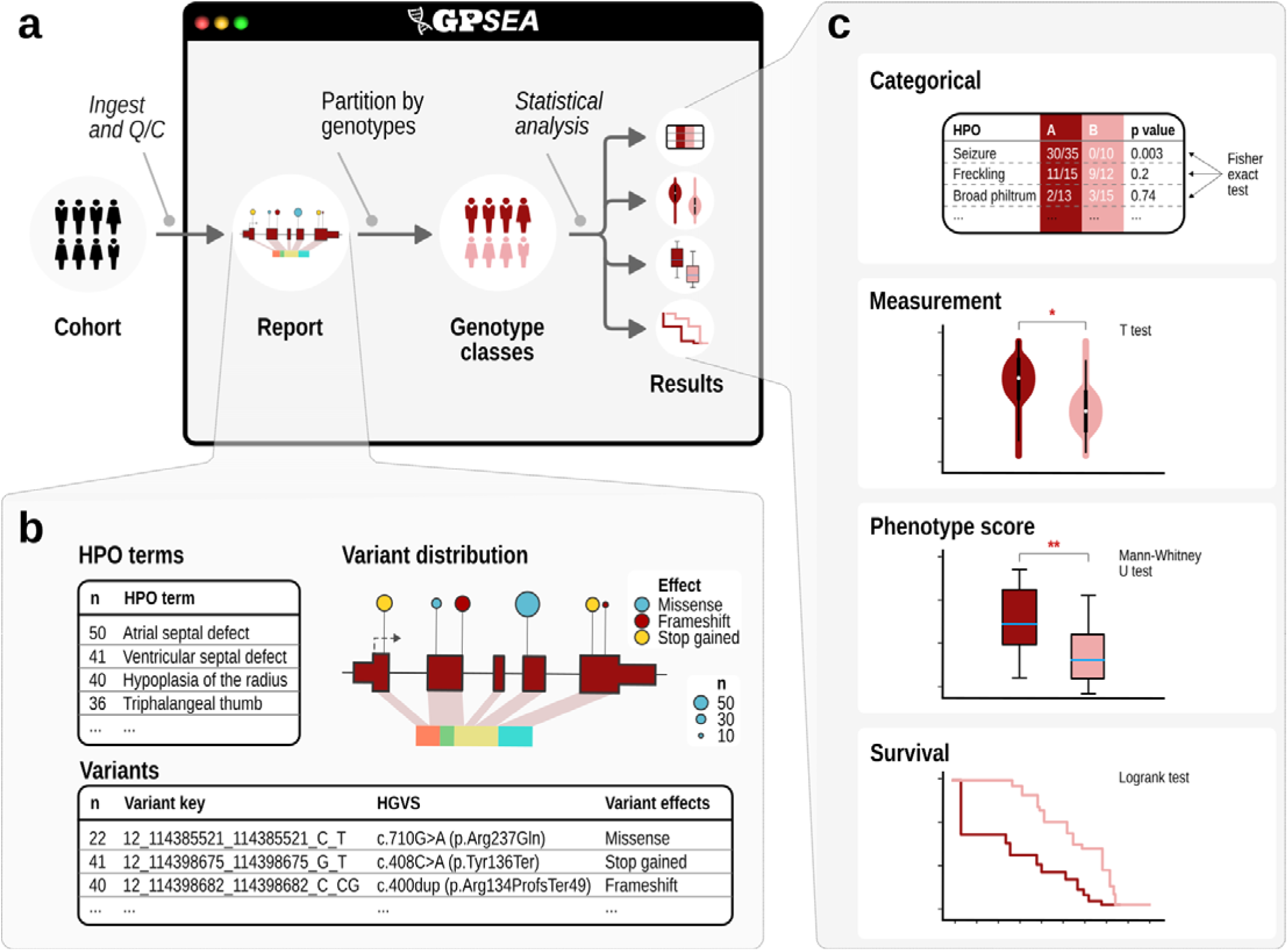
Schematic overview of GPSEA workflow. **a) Overview**. GPSEA is a Python package designed to work well in Jupyter notebooks. GPSEA takes a collection of GA4GH phenopackets as input, performs quality assessment and visualizes the salient characteristics of the cohort; genotype classes are defined (Figure 2); and one of four classes of statistical test is performed for each hypothesis the user decides to test. **b) Visualize data and formulate hypotheses**. GPSEA displays tables with the distribution of phenotypic abnormalities, disease diagnoses, variants, and other information, and presents a cartoon with the distribution of variants across the protein. This information intends to help users formulate hypotheses about genotype-phenotype correlations (GPCs). **c) Statistical testing**. GPSEA offers four main ways of testing phenotypes (See text for details and Figure 4 for examples).

### Partitioning the cohort according to genotypes

The GPC analysis starts with defining one or more hypotheses. Ideally, decisions regarding the analysis structure will be based on prior hypotheses about the disease or gene in question. Alternatively or additionally, GPSEA helps users generate hypotheses based on the tables and visualizations. For instance, if GPSEA shows that roughly 50% of the variants observed in a cohort are missense, and the other 50% are truncation or presumed loss of function (LoF) variants, users may choose to analyze whether missense are associated with significantly different phenotypes than LoF variants. Other GPSEA visualizations may help to formulate hypotheses about commonly occurring variants, protein domains, exons, or other classes of variation. Once a hypothesis has been conceived, it must be encoded into a genotype classifier, i.e., a GPSEA component that assigns each cohort member into one of the (typically two) classes.

Variant predicates are key building blocks for classification based on genomic variants. GPSEA offers predicates for specific variants, for variant effect categories such as missense and stop gained variants, specific exons, protein regions, types of structural variant, and others. Predicates can be combined using Boolean algebra to create more expressive predicates. The framework provides five genotype classifiers that are used to divide the cohort into groups for statistical testing. The mono- and biallelic classifiers use variant predicates to select the variants of interest and assign individuals to genotype classes. The sex classifier investigates differences between males and females. The diagnosis classifier tests for differences in the phenotypic spectrum of different diseases (e.g. Loeys-Dietz syndrome 1 OMIM:609192 vs. Loeys-Dietz syndrome 2 OMIM:610168). The allele count classifier takes a variant predicate to select the variants of interest and classify the individuals according to the number of variant alleles. For instance, monoallelic and biallelic variants in *EHZ1* cause dominant and recessive neurodevelopmental disorders;^26^ with the allele count classifier, the distribution of phenotypic features can be compared between individuals with monoallelic and biallelic variants (**Fig. 2**; **Supplementary Table S1**).

**Fig. 2:**
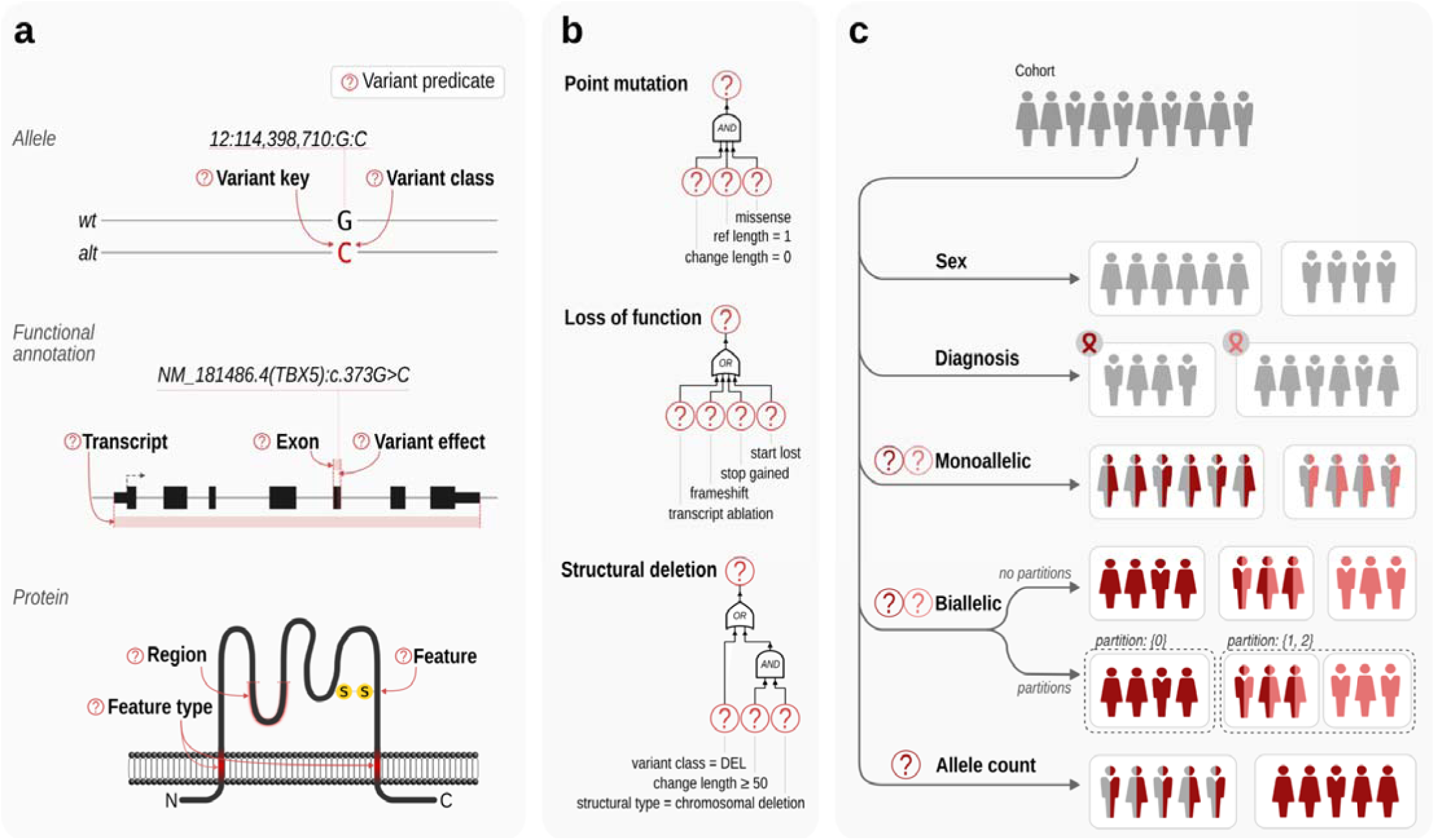
Variant predicates and genotype classifiers. **a, Variant predicate tests**. GPSEA provides predicate functions that test if a variant meets a criterion from one of three evidence groups: allele, functional annotation or protein. For instance, the predicate checks if the variant is a deletion, if it overlaps with a specific exon or with a protein region of interest. **b, Boolean algebra**. Variant predicates can be combined using AND, OR, and NOT operators of Boolean algebra to test complex criteria. For instance, a predicate for a point mutation can be formulated as a “missense mutation affecting one reference base and change length of z (no sequence loss or gain). A predicate for a loss-of-function mutation can be defined as a mutation leading to a ro” transcript ablation, frameshift, introduction of a premature stop codon or the start codon loss. A predicate for a structural deletion can test if the variant is either an imprecise chromosomal deletion or a deletion involving 50 or more base pairs (or other thresholds) ^27^. **c, Genotype classifiers**. Each classifier splits a cohort into two or more classes to enable genotype-phenotype comparisons. GPSEA ships with five built-in classifiers to classify the cohort members using their sex, diagnosis, a fixed count of alleles of different types (Monoallelic and Biallelic), or by a different allele count of the same type (Allele count).

### Analyzing the cohort according to phenotypes

GPSEA offers four major tests for different kinds of clinical data. The categorical test is designed to be used for observations of HPO terms (observed/excluded) or disease diagnoses. Numerical values such as laboratory measurements can be analyzed with a t test. Phenotype scores can be derived as a proxy of clinical severity and are analyzed by a Mann-Whitney U test. Finally, survival analysis can be performed for age of disease onset, onset of a phenotypic feature (HPO term), or death. The following sections explain the approach. The examples are taken from the 85 cohorts (Overview in **Supplemental Tables S2-S10**; Result summary for each cohort in **Supplemental Figures S1-S86**; Source code for analyses: See online resources).

#### Categorical association

The Fisher exact test (FET) calculates the exact probability for observing as extreme a contingency table for the relationship between two categorical variables, if in fact they are independent. In our implementation, the two categorical variables are the genotype and the phenotype. For instance, the individuals of the cohort may be divided according to whether or not they have a stop-gained (nonsense) variant and according to whether or not they have *Strabismus* (HP:0000486).

#### Independent Filtering for Human Phenotype Ontology (IF-HPO)

Larger cohorts may include several hundreds of HPO terms. Even though many published articles on GPC analysis do not apply a multiple-testing correction (MTC) to the tests, we feel it is appropriate to do so unless users have a well defined hypothesis prior to performing the analysis. In the cohorts analyzed here, up to hundreds of HPO terms are used, and so MTC can result in low statistical power. In high-dimensional data such as analysis of mRNA expression in cohorts, a two-stage approach termed independent filtering prefilters hypotheses (*e.g.* expression differences per gene) by a criterion independent of the test statistic under the null hypothesis, before testing any hypotheses, to reduce the number of the hypotheses tested at stage two, leading to milder MTC effect and thereby increased power.^28^ We developed an analogous approach, IF-HPO, to reduce the testing burden before MTC is applied (**Fig. 3**). This rule-based approach leverages the hierarchical structure of the HPO to avoid unnecessary tests and the tests that are unlikely to reveal an interesting result. IF-HPO reduces the total number of tested terms by over ten-fold in the cohorts analyzed here (before filtering: mean 304, median 277, min 45, max 967; following independent filtering: mean 40, median 28, min 1, max 225). While any such heuristic has its tradeoffs and it is possible that some significant and interesting results are removed, the IF-HPO procedure provides a substantial boost in statistical power for the remaining terms.

**Fig. 3:**
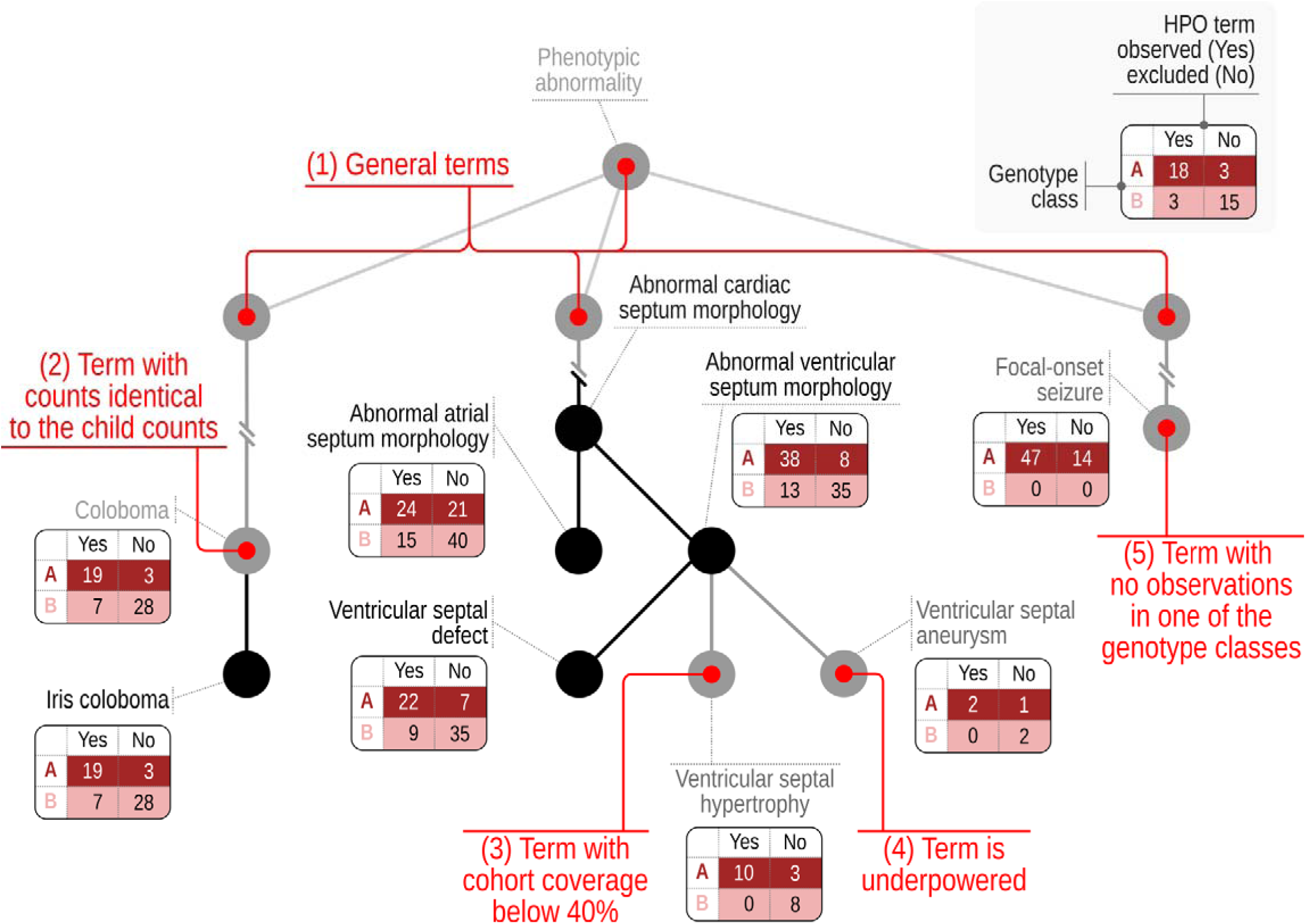
Independent-Filtering for Human Phenotype Ontology. Independent filtering for HPO (IF-HPO) removes hypotheses (here, HPO terms) by criteria independent of the tes statistic to reduce the multiple testing burden and boost power. The HPO has a hierarchical structure going from general to specific terms. (1) IF-HPO does not test the top two levels of the HPO under the *Phenotypic abnormality* root or the terms that are not descendants of the *Phenotypic abnormality* under the assumption that more specific terms are of higher medical and scientific interest and the signal is likely to be driven by a more specific clinical manifestation. (2) Terms are not tested if they have the exact same counts as one of their child terms, because in this case the annotations of the parent term are derived entirely from those of the child term by the true path rule. (3) Terms are not tested if the coverage is less than 40% of the entire cohort (assuming a cohort of 100 individuals in the figure), under the assumption that the result would not be representative for the cohort. (4) Terms are not tested if the total count is below a threshold for reaching the nominal statistical power. (5) Finally, terms are not tested if one of the genotype classes has neither present nor excluded observations. Fig. 4b shows an example of applying categorical analysis to test for associations of variants at residue 1830^Arg^. Nine HPO terms were found to have a significantly lower or higher frequency in individuals with variants at this position as compared to other variants in the *NF1* gene (Additional results for *NF1* in **Supplemental Figure S42;** Summary of all significant categorical test results in **Supplemental Table S2**).

Alternatively, users can choose to test specific HPO terms if there is a prior hypothesis, or to test all terms. Standard MTC is applied to the tests performed following IF-HPO. By default, GPSEA applies the Benjamini Hochberg method^29^ (10 other standard MTC approaches are available).

#### Phenotype scores

Phenotype scores have been developed for some diseases to provide a semi-objective assessment of disease severity. Many such scores count the total number of observed phenotypic features from a list of stipulated terms. Other scores involve more complicated systems that use boolean logic or thresholding. An example of the first score is provided in the analysis of Atrophin domain variants in the *RERE* gene, which were previously found to be significantly associated with higher scores defined by the counts of structural defects of the brain, eye, heart, kidney, and sensorineural hearing loss.^30^ GPSEA provides Counting scorer that allows users to indicate relevant HPO terms. The hierarchical structure of the HPO is used to count annotations to the term itself or any of its descendents; for each of the terms, a count of 1 is given if one or more such annotations were found; otherwise, a count of zero is assigned for the term. The phenotype score thus ranges from 0 if no relevant abnormalities were recorded to the total count of specified HPO terms if an abnormality was found in all items. For instance, one of the terms was *Abnormal brain morphology* (HP:0012443). Therefore, one point would be given if the individual was annotated to any of the descendent terms, for instance, *Agenesis of corpus callosum* (HP:0001274). There was a significantly higher severity score for variants located in the Atrophin domain of *RERE* (**Fig. 4c; Supplemental Fig. S49**).

**Fig 4:**
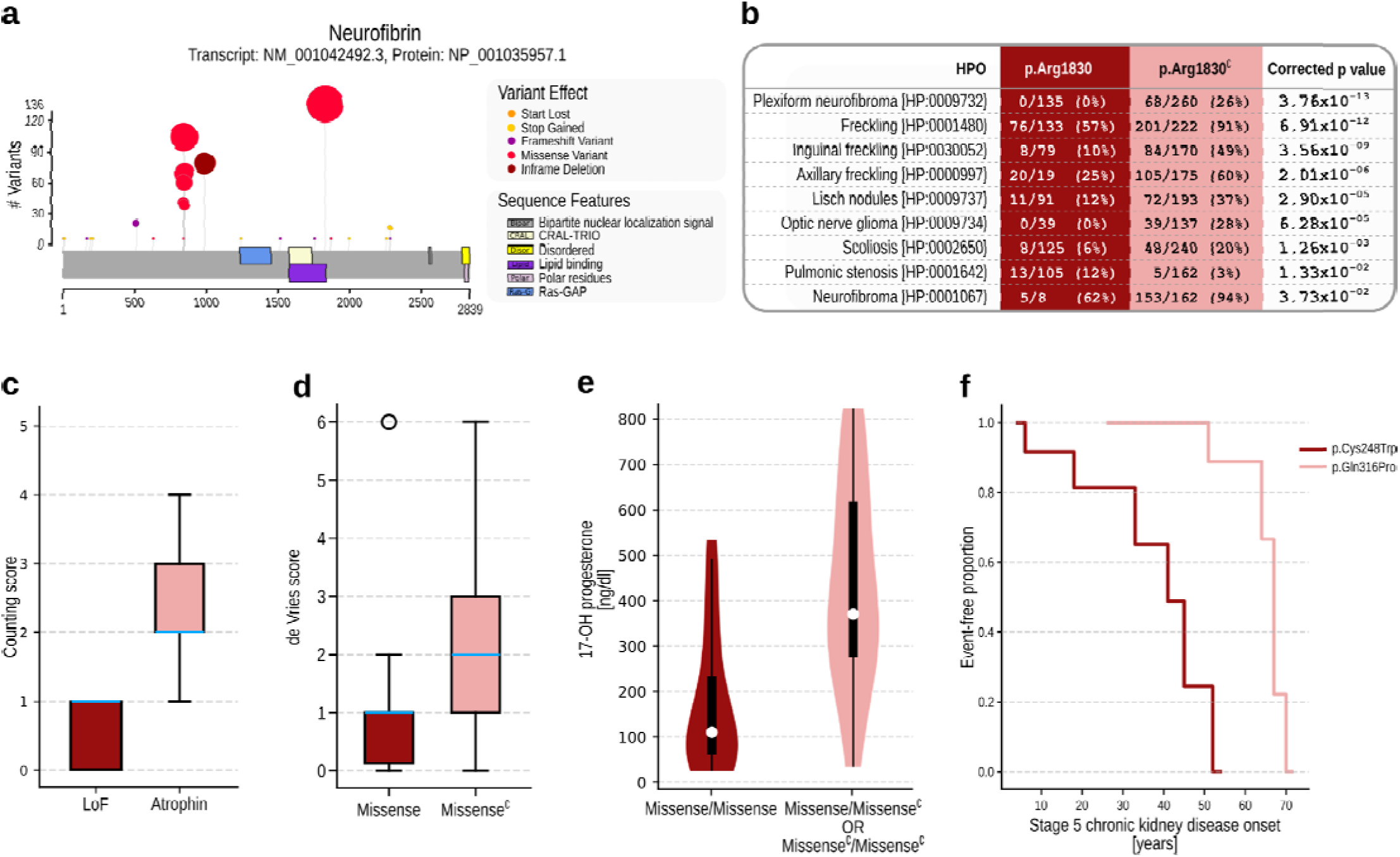
GPC Analysis. This figure shows excerpted results from five example analyses. **a) Visualization.** GPSEA generates a cartoon showing the location and frequency of variants in protein sequences. The following panels show examples of statistically significant GPCs identified by GPSEA. **b) Categorical analysis.** Several phenotypic abnormalities (HPO terms) such as neurofibromas, freckling, Lisch nodules, optic nerve glioma, and scoliosis are significantly less frequent in individuals with neurofibromatosis type 1 due to variants located at the arginine residue at position 1830 of neurofibromin isoform 1 than in those with different mutations (Fisher exact test, IF-HPO, Benjamini-Hochberg correction). Pulmonary stenosis is, however, observed more often in those with Arg1830 mutation. **c) Severity score**. A box plot with counts of abnormalities in 5 organ systems in the individuals with mutations in *RERE* showing the association of the mutations in the Atrophin domain with abnormalities in multiple organ systems;^30^ (Mann-Whitney U test, p=1.44 x 10^-3^). The boxes represent the Q1-Q3 range and the whiskers extend to the farthest score lying within 1.5x the interquartile range. The blue line denotes the median score. **d) de Vries score**. Box plots representing the association of the de Vries phenotype score ^13^ and missense variants in *CHD8* (Mann-Whitney U test, p=8.99 x 10^-4^) **e) Continuous phenotypes.** Association of *CYP21A2* genotype (homozygous missense vs.other) with concentration of 17-OH progesterone (t-test, p=7.91 x 10^-6^). **f) Survival analysis**. Comparison of the onset of *Stage 5 chronic kidney disease* (HP:0003774) in individuals with *UMOD* mutations showing a significantly earlier onset of the disease in the individuals with NP_000491.4:p.(Cys248Trp) mutation than in those with p.(Gln316Pro) (Logrank test, p=4.1 x 10-4). Missense^∁^: set complement of “missense”, i.e., any mutation that is *not* missense, LoF: loss-of-function.

Other scores have been developed with more involved rules. For instance, the de Vries score was developed as a relatively simple phenotypic severity score for individuals with intellectual disability in which points are given for (severity of) intellectual disability, growth abnormalities (prenatal and postnatal), facial dysmorphisms, non-facial dysmorphisms, and other congenital anomalies.^13^ We developed a modified version of this score that uses the structure of the HPO to roll up specific terms. Using this, we identified a significantly lower score (corresponding to milder clinical manifestations) in individuals with *CHD8* missense variants than those with other *CHD8* variants, similar to the original application of the score to *CHD8* (**Fig. 4d**).^13^ We also applied the score to other cohorts; a similar significant association of missense variants with lower scores was identified for *CTCF* (**Supplemental Fig. S15;** Summary of all phenotype score results in **Supplemental Table S3**).

#### Numerical values

HPO terms are categorical, and are not designed to capture continuous (numerical) values. Instead, the GA4GH Phenopacket Schema has a measurement element that can be used to represent the results of laboratory tests for analytes such as enzyme activity or metabolite concentrations. GPSEA can test association of numerical data, such as metabolite levels or enzyme activities, with genotype classes. For example, in 21 hydroxylase deficiency, it is assumed that the mildest mutation determines the phenotype in compound heterozygotes, and that missense variants have a less severe effect on enzyme activity than do other variants such as truncation or ablation variants.^31^ Using GPSEA, we applied a t-test and observed significantly lower 17-OH-progesterone levels (which are known to increase with reduced 21 hydroxylase activity) in the individuals with two missense alleles (**Fig. 4e; Supplemental Fig. S16**; Summary of all t test results in **Supplemental Table S4**).

#### Survival analysis

GPSEA can perform survival analysis to assess associations between genotype classes and mortality, disease onset, or onset of a specific phenotypic abnormality such as *Stage 5 chronic kidney disease* (HP:0003774). The data is plotted as a Kaplan Meier curve, and a Logrank test is applied to assess statistical significance (**Figure 4f; Supplemental Figure S81**). This requires that the phenopackets have information about the ages of onset or mortality; because this information was not available in most of the publications curated for this project, survival analysis was performed only for a subset of cohorts. The analysis leverages the ontological structure of the HPO to rollup annotation from descendent terms, similar to the procedure for categorical analysis. If we are testing for onset of *Seizure* (HP:0001250), and an individual was noted to have both *Tonic seizure* (HP:0032792) and *Generalized myoclonic seizure* (HP:0002123), then the youngest age of onset for the latter two terms is chosen (Summary of all survival analysis results in **Supplemental Tables S5-S7**).

#### Analysis by disease diagnosis

GPSEA also allows users to search for HPO terms that are different between two diseases. For instance, 8/25 (32%) individuals with Kabuki syndrome 1 (OMIM:147920) displayed Feeding difficulties [HP:0011968], whereas 55/63 (87%) individuals with Kabuki syndrome 2 (OMIM:300867) did (p=2.1 x 10^-5^, Fisher exact test, IF-HPO, Benjamini-Hochberg correction). A total of 16 significant findings were observed (**Supplemental Fig. S28**; Summary of all disease analysis results in **Supplemental Table S8**).

#### Analysis for sex differences

A categorical analysis can be performed for association between phenotypic features and sex (male, female). Tests were performed in 44 cohorts, and one significant difference was identified in the cohort for Kabuki syndrome type 2, in which 14/18 (78%) males were annotated to the HPO term *Intellectual disability, severe* (HP:0010864), but only 7/25 (28%) females (p=1.94 x 10^-3^; FET, BH) (**Supplemental Table S9**).

### GPCs are common in Mendelian disease

We analyzed 85 cohorts with 6613 individuals (median 49 per cohort, range: 16-462) with 122 Mendelian diseases. Each individual was encoded as a phenopacket with information about the disease diagnosis, phenotypic abnormalities (HPO terms), and where available age of onset of the disease and of individual features, age of death, and in some cases numerical laboratory test results. GPSEA analysis was applied to each of the cohorts. Existing knowledge about GPCs related to the gene or disease of interest was sought in PubMed (Methods) and if possible, an analysis was performed in GPSEA to reproduce a similar result using the cohorts available. Alternatively or additionally, GPSEA visualizations were consulted to generate hypotheses about testable GPCs for common variant categories (missense, nonsense, etc.), common variants, exons, protein domains, or regions. If relevant information was available about onset or mortality, survival analysis was performed. In some cases, phenotype severity scores were applied or numerical analyses were performed. A total of 241 significant correlations were identified. In some cases, our choice of hypotheses to test was guided by previously published results; we searched the literature for published GPCs for all of the cohorts (Methods). We did not identify even a single publication for which the data and analysis script were made available in a way that would allow the original analysis to be replicated. Additionally, we attempted to curate data from all available publications for each gene or disease being analyzed, and so had different cohorts and a different methodology. Nevertheless, we assessed whether results are similar to previously published ones. Some of our cohorts involved comparison of diseases with well known phenotypic differences; for instance, we compared Spastic paraplegia 78 (SPG78) and Kufor-Rakeb syndrome (KRS), both of which are caused by variants in *ATP13A2*, and showed a significantly higher frequency of *Parkinsonism* (HP:0001300) and *Bradykinesia* (HP:0002067). Although statistical tests are rarely conducted to characterize allelic diseases in this way, we regard such differences as well known and record them as previously published in the literature for the purposes of **Table 1**. Other differences, such as a higher prevalence of *Intrauterine growth retardation* (HP:0001511) in Cornelia de Lange syndrome 1 (37/45; 82%) (*NIPBL*), as compared to Cornelia de Lange syndrome 6 (2/9; 22%; p=0.031, Fisher Exact Test, BH) could not be identified in previous literature. Significant results were identified for 48 cohorts. We identified previously published GPCs for 31 of these cohorts, many of which overlapped with our findings (references and detailed analysis in **Supplemental Fig. S1-S86**). 52 significant findings in the remaining 16 cohorts represent candidate GPCs that should be validated by independent studies on validation cohorts (**Table 1**; references for previously published findings in **Supplemental Fig. S1-S86**).

**Table 1.** Summary of tests performed according to type of test. The table provides a summary of the results on the 85 cohorts tested. The table is arranged according to the types of statistical tests offered by GPSEA. **Categorical analysis**: Association of genotypes with phenotypes by Fisher exact test. **t test**: Test of means of continuous values by Student’s t test. **HPO onset**: Logrank test for association of genotypes with age of onset of a phenotypic abnormality represented by an HPO term. **Disease onset**: Logrank test for association of genotypes with age of onset of a disease.

| Statistical procedure | Cohorts tested | Tests performed | Significant tests |
| --- | --- | --- | --- |
| Categorical analysis | 83 | 8950 | 205 |
| t test | 2 | 3 | 3 |
| HPO onset | 4 | 6 | 3 |
| Disease onset | 4 | 10 | 6 |
| Mortality | 4 | 3 | 1 |
| Phenotype scores | 4 | 9 | 7 |
| Disease diagnosis | 6 | 266 | 15 |
| Sex differences | 44 | 1979 | 1 |
| **Total** | **85(\*)** | **11226** | **241** |

**Mortality**: Logrank test for association of genotypes with age of death. **Phenotype scores**: Mann-Whitney U test for association of genotypes with magnitude of a phenotype severity score. Multiple testing correction was applied to the categorical tests (Benjamini-Hochberg method), following the Independent Filtering procedure (IF-HPO). No multiple testing correction was applied to the remaining tests, which were considered to represent distinct hypotheses (Detailed results shown in **Supplemental Table S10**). (*)Multiple statistical procedure types were performed for some cohorts.

#### Distribution of phenotypic features with significant GPCs

We analyzed the distribution of HPO terms for which significant GPCs were identified by identifying the top-level term (direct child of *Phenotypic abnormality*, HP:0000118). The distribution of terms was significantly different from what one would expect based on the counts of all terms in the Phenotypic abnormality subhierarchy of the HPO (Exact multinomial test, p=2.39 x 10^-34^). The largest differences were observed for *Abnormality of the nervous system* (HP:0000707; expected 10.8%, observed 22.2%), *Abnormality of metabolism/homeostasis* (HP:0001939; expected 9.5%, observed 1.2%, and *Neoplasm* (HP:0002664; expected 2.8%, observed 7.8%). This raises the possibility that phenotypic features in different organ systems may have a differential tendency to display GPCs, although our observation may also be the result of an ascertainment or other bias (**Supplemental Table S11**).

## Discussion

Precision genomic medicine is an emerging medical discipline that aims to apply genomic information for prediction, prevention for early diagnosis or tailored treatment in order to improve clinical care. Although precision approaches have been applied successfully to some Mendelian diseases such as cystic fibrosis,^32^ our understanding of diseases subtypes and GPCs is limited for the vast majority of the roughly 7000 characterized rare Mendelian diseases. Understanding GPCs can contribute towards understanding disease pathophysiology and stratified clinical management. A barrier has been the lack of standardized data exchange and analysis schemas, which means that it is difficult to combine data from multiple sources and to the fact that scripts or program code needs to be created anew for each project. The GA4GH Phenopacket Schema was released in 2022 and approved by the International Standards Organization (ISO 4454:2022) as a standard for sharing clinical and genomic information about an individual. Each phenopacket is a computational representation of the clinical trajectory of one individual and can contain data about phenotypic descriptions, numerical measurements, genetic information, diagnoses, and treatments. A phenopacket can be used for data exchange and as a computational model for software that supports phenotype-driven genomic diagnostics and for algorithms that facilitate patient classification and stratification.^22^ Therefore, when phenopackets serve as a unifying standard across projects, locations, and registries, they enable machine-readability and reusability for multiple analyses while offering precise, ontology-based semantics and adherence to the FAIR data principles. The Phenopacket Schema thus enables software such as GPSEA to be used for any relevant dataset that is available as or can be transformed into phenopackets.

As more and more data in human genetics becomes available with HPO annotations, new challenges arise for analysis because of the ontological structure of the HPO. Analysis software needs to “roll up” annotations; for instance, if an individual is annotated to *Nuclear cataract* (HP:0100018), it is always true that the individual also has the manifestation described by the parent term *Zonular cataract* (HP:0010920) and the grandparent term *Cataract* (HP:0000518), and so on (this is termed the “true path rule”). However, HPO annotation is performed to the most specific level, and typical statistical software used to work with data frames is not able to perform the rolling up that is needed for correct analysis. Another challenge when using ontologies for analysis relates to the redundancies inherent in the hierarchical structure.^33^ GPSEA addresses both challenges by preparing data for analysis using the true path rule and minimizing redundancy and multiple-testing burden by the IF-HPO procedure.

Many publications on GPC analysis either do not provide raw data, or provide a summary of data in the supplement as an Excel file or related format. Rarely is analysis code provided to reproduce the results, and since the formats used in the supplemental files are diverse, it requires a substantial amount of work to prepare them for statistical analysis. The approach we have presented here makes the investigation of GPCs FAIR. The entirety of the data used for the analysis is freely available in Phenopacket Store,^23^ and all code used for analysis is available in the GitHub repository (one notebook is provided for each cohort). An additional advantage of this approach is that results for new cohorts can be assessed in comparison to a body of previous results using the same software.

For this project, we used data derived from published cohorts for our analysis. The cohorts we present here were all derived from published case and cohort reports. In general, published cohorts do not contain comprehensive clinical information, but instead present features deemed most relevant or important by the authors. Furthermore, phenotype is not solely determined by genetic factors; environmental and lifestyle influences can interact with allelic variability and modifier genes to further shape phenotype. For this reason, and also because of potential publication biases that may lead to an overestimation of clinical severity,^34^ the results about specific GPCs presented here should be regarded as hypotheses that will require confirmation in independent studies.

Allelic variability is only one of many factors that determine phenotype. However, phenotypic severity or penetrance can be influenced by the genotype at another locus, which is referred to as a modifier gene.^35^ Indeed, in hereditary breast cancer, polygenic risk scores (PRSs) for ovarian cancer are associated with penetrance of ovarian cancer in *BRCA1*/*BRCA2* variant carriers.^36^ GPSEA could be easily extended to evaluate the effects of PRSs or variants in modifier genes, but the main challenge will be in the collection of comprehensive clinical and genomic data. Prospectively capturing clinical data using a common data model could be beneficial.^37^

GPSEA serves as a foundational tool, enabling correlation studies to be conducted at any scale, within any setup, and for any hypothesis. For instance, we envision that GPSEA can be used for comprehensive research databases with more balanced datasets, whether or not the raw data can be shared publicly. Wide community adoption of the Phenopacket Schema and application of consistent practices for recording and reporting phenotypic features in publications and databases would accelerate characterization of GPCs across the Mendeliome and thereby contribute to our knowledge of the natural history of rare diseases.

## Methods

### Input data

Genotypic and phenotypic data about individuals with rare Mendelian disease were derived from the Phenopacket Store repository.^23^ Version 0.1.24 of Phenopacket Store includes 8182 phenopackets representing 491 Mendelian and chromosomal diseases associated with 457 genes and 4469 unique pathogenic alleles curated from 1227 different publications. The phenopackets are structured representations of data comprising age of onset and age at last examination, vital status, genotype of the variant(s) deemed to be causal, disease diagnosis, and HPO terms representing the clinical manifestations of the disease. Where available in the original publication, the age of onset of the clinical manifestations is indicated. Missing data were not imputed.

For each GPC analysis, a cohort was defined based on the gene or disease. The analysis code for each cohort is available in the project GitHub repository (Code availability). The phenopackets used for analysis are automatically imported from Phenopacket Store by each cohort notebook. If desired, the phenopackets can also be obtained directly from Phenopacket Store (Data availability).

### GPSEA

GPSEA is a Python package for streamlining GPC analysis. For the analysis described here, version 0.9.7 was used. GPSEA enables a stepwise workflow to characterize GPCs. (1) A collection of phenopackets is loaded into a cohort and a report with basic descriptive statistics is displayed. The report comprises the number of individuals, the distribution of sex and age, as well as tables with the most commonly annotated HPO terms, diseases, and associated genes. The variants are summarized according to their frequency and the predicted effect on the clinically relevant transcript, including a graphic with the location and frequencies of all non-structural variants. (2) The user can generate hypotheses about the GPC analyses that are most likely to be fruitful. For instance, if roughly half of the variants are missense, then it might make sense to test whether missense variants are associated with different clinical manifestations as compared to other variants. (3) The analysis is configured with respect to multiple testing correction and other parameters. (4) A hypothesis is expressed with GPSEA’s variant predicates and genotype classifiers to define a partitioning of the cohort by genotype for testing with four statistical approaches. (5) The analysis results are presented as figures and tables. The data used to perform the statistical tests can be exported as data frames for additional analysis. A detailed tutorial is available online (Code availability).

### Statistical tests

GPSEA provides four main statistical tests, each of which can be combined with predicates to test a wide variety of GPCs. For each test performed, GPSEA checks whether data is available for each individual (observed or excluded in the case of HPO term-based tests, numerical measurement results for the t-test, and duration until event for survival analysis), and omits a data point if either phenotypic or genotypic information is not available or applicable. Associations with p value less than 0.05 were considered to be statistically significant.

### Dichotomous (qualitative) phenotypes

The Fisher exact test (FET) calculates the exact probability value for the relationship between two dichotomous variables. In our implementation, the two dichotomous variables are the genotype and the phenotype. For instance, the individuals of the cohort may be divided according to whether they have a missense or a stop gained (nonsense) variant and according to whether or not they have *Strabismus* (HP:0000486):

|  | Missense | Stop gained |
| --- | --- | --- |
| HP:0000486 observed | 17 | 6 |
| HP:0000486 excluded | 1 | 19 |

In this example, a FET is used to assess the association of *Strabismus* with missense variants compared to stop-gained (nonsense) variants. A p value of 5.43 x 10^-6^ is obtained, meaning there is a significant difference between the groups.

### Multiple testing correction

For the FET procedure, GPSEA can perform one FET test for each HPO term identified in the cohort. However, in some cohorts, up to hundreds of terms are identified. GPSEA offers two approaches towards controlling the type I error rate, i.e., the probability of rejecting the null hypothesis when it is true. Firstly, 11 classical multiple testing correction procedures are offered including Bonferroni and Benjamini-Hochberg (BH). A MTC procedure (by default BH) is applied to each test result and the adjusted p-value is reported.

The multiple testing burden can also be reduced by selecting only a subset of terms to test. If the user has a hypothesis about which HPO terms are involved in a GPC, then GPSEA can be instructed to test only this term (or subset of terms).

### Independent Filtering for Human Phenotype Ontology: IF-HPO

Additionally, we developed the IF-HPO procedure that applies a series of heuristics to select terms to test. The procedure was inspired by analogous strategies used in functional genomics that consist in filtering by a variable that is independent of the test statistic under the null hypothesis. By reducing the number of tests in this way, we maximize power for differential testing, while preserving Type I error control.^28^ (1) Skipping “general” level terms. All the direct children of the root phenotype term *Phenotypic abnormality* are skipped, because of the assumption that if there is a valid signal, it will derive from one of the more specific descendents. For instance, *Abnormality of the nervous system* (HP:0000707) is a child of *Phenotypic abnormality*, and this assumption implies that if there is a signal from the nervous system, it will lead to at least one of the descendants of *Abnormality of the nervous system* being significant. (2) Skip terms if all counts are identical to counts for a child term. Let’s say a term such as *Posterior polar cataract* (HP:0001115) was observed in 7 of 11 individuals with MISSENSE variants and in 3 of 8 individuals with NONSENSE variants. If we find the same patient counts (7 of 11 and 3 of 8) in the parent term *Polar cataract* (HP:0010696), then we choose to not test the parent term. This is because the more specific an HPO term is, the more information it has (the more interesting the correlation would be if it exists), and the result of the Fisher Exact test for Polar cataract would be exactly the same as for Posterior polar cataract. (3) We skip terms that occur in less than a certain proportion of cohort members (default 0.4), because even if a correlation is identified, it is unlikely to be of great interest because the phenotype in question occurs rarely. (4) If the individuals are binned into 2 genotype groups and 2 phenotype groups (2x2) and the total count of individuals is less than 7, or into 3 genotype groups and 2 phenotype groups (3x2) and the total count of individuals is less than 6, then there is a lack even of the nominal statistical power and the counts can never be significant. (5) Skip terms if there are no HPO observations in a genotype class. If one of the genotype classes has neither observed nor excluded observations for an HPO term, skip it. This situation suggests that the data are not sufficiently rich to confidently perform a test.

### Phenotype scores

It is hard to define an objective measure for the clinical severity of disease. Some published studies use the total count of features from a defined set as a proxy for severity. For instance, Jordan et al. (2018)^30^ found that the total number of structural defects of the brain, eye, heart, and kidney and sensorineural hearing loss seen in individuals with point mutations in the Atrophin domain of the *RERE* gene is significantly higher than expected based on the number of similar defects seen in individuals with putative loss-of-function variants. Since there are five potential defects, each individual has a count ranging between 0 and 5. The authors regarded higher counts as representative of a severe clinical presentation.^30^

GPSEA performs a Mann-Whitney U Test (a.k.a. Wilcoxon Rank-Sum Test) to compare the distribution of such counts between genotype classes. This is a non-parametric test that compares the class medians to determine if they come from the same distribution.

A set of HPO terms that define the severity score is entered. GPSEA increments the total count by one for each of the terms (or more specific descendent terms) to which an individual is annotated in the phenopacket.

If multiple HPO terms are found related to one of the specified terms then only one count is incremented (e.g., if both *Ventricular septal defect* (HP:0001629) and *Atrial septal defect* (HP:0001631) are identified then a score of 1 and not 2 is entered for *Abnormal heart morphology* (HP:0001627)).

The de Vries score is a simple phenotypic severity score for individuals with intellectual disability in which points are given for (severity of) intellectual disability, growth abnormalities (prenatal and postnatal), facial dysmorphisms, non-facial dysmorphisms, and other congenital anomalies.^13,38^ Our implementation of the de Vries score leverages the hierarchical structure of the HPO to include more specific descendants of phenotypic abnormalities included in the original score. For instance, *Disproportionate short stature* (HP:0003498) would be counted for *Short stature* (HP:0004322)).

GPSEA can also make use of user-defined functions to support a plethora of scoring schemes used in different clinical domains. A scoring function is required to “condense” the phenotype of the individual into a numeric score, or return “not a number” (NaN) value, if the individual should be excluded from the analysis. We provide examples for using user-defined functions as well as defining custom phenotype scorers in GPSEA documentation.

### Genotype-specific survival analysis

We may wish to compare the genotype classes with respect to the time point of a specific event, such as age of onset, age at death, or age at onset of a specified phenotypic feature such as kidney failure. To do this, GPSEA tabulates the age of the event (if the event was observed) or whether the individual was still alive (based on the time of last evaluation) without the event having occurred - that is, the survival time was right censored. The Logrank test is used to test the null hypothesis that there is no difference between the populations in the probability of an event at any time point.^39^

### Student’s t test for numerical values

GPSEA performs an unpaired and two-sided t-test to compare the means of the two groups defined by the genotype classifier. GPSEA expects numerical data for this test to be made available as measurement elements in the GA4GH Phenopacket Schema. GPSEA does not stipulate any specific ontology to represent the measurements, but in our examples we use LOINC codes to denote the assay and UCUM codes to represent units. GPSEA does not apply multiple-testing correction to these results, and users need to perform one analysis for each measurement to be tested.

### Variant predicates

GPSEA has flexible predicates (functions that return either True or False based on the input) that can be used to partition the cohort into (usually) two groups of individuals. The predicates can be combined using “AND”, “OR”, and “NOT” operators of Boolean algebra logic to test complex conditions. GPSEA offers predicates for specific variants, for variant effect categories such as missense and stop gained variants, specific exons, protein regions, types of structural variant, and others (**Supplemental Table S1**). Besides the off-the-shelf predicates, custom predicates for testing arbitrary variant properties can be designed.

### Genotype classifiers

GPSEA has five classifiers. The monoallelic classifiers can be used to investigate autosomal dominant (heterozygous variants) and X-chromosomal diseases (hemizygous variants; if desired cohorts of males and females can be analyzed, in which case the monoallelic classifier would identify hemizygous variants in males and heterozygous variants in females). For instance, a monoallelic classifier might partition individuals according to whether they have a heterozygous missense variant (group A) or not (group B); another classifier might partition individuals according to whether they have a heterozygous missense variant (group A) or a heterozygous structural variant (group B). In some cases, a genotype classifier might omit certain individuals; for instance, in the previous example, the classifier would omit any individual who does not have either a missense or a structural variant, or any individuals with homozygous or compound heterozygous genotypes for missense or structural variants. The biallelic classifier is designed for autosomal recessive conditions; it is possible to test three genotypes (e.g., AA, AB, BB, where A refers to a genotype such as “stop-gained variant” and B refers to other variants), which is a 3x2 contingency table that can be analyzed by FET, or to form two groups (e.g., AA and AB vs. BB, i.e., one or two stop-gained alleles vs. no stop-gained allele). The online documentation shows how to define partitions to perform these tests. Any of the variant predicates can be used together with the monoallelic and biallelic genotype classifiers.

The disease classifier is designed to assay differences between the phenotypic features of two different diseases. For instance, in our cohort we test for differences between Loeys-Dietz syndrome 1 and Loeys-Dietz syndrome 3 as well as between autosomal recessive and dominant forms of Robinow syndrome. The sex classifier is designed to test differences between the phenotypic features observed in the males and females of a cohort. Individuals with unknown or unspecified sex are ignored by this classifier. The allele count classifier is designed to assay differences between individuals with one and two variant alleles in the same gene. For instance, in our cohort we test whether individuals with monoallelic and biallelic variants in *EHZ1* have distinct phenotypic profiles. This disease, sex, and allele count classifiers do not take genotype into account.

### Visualization

GPSEA visualizes variants against the background of the protein domain structure. To do so, it leverages the UniProt^40^ API to retrieve information about protein domains. It is also possible to manually construct a dataframe with information about protein domains in cases where the UniProt API fails or does not contain information about a domain of interest. GPSEA then extracts information about all variants found in the cohort and plots each variant as a “lollipop” whose height and size reflect the number of times the variant was found in the cohort and color represents the functional effect predicted for the transcript of interest. Protein domains are depicted as colored boxes. Currently, GPSEA does not display non-coding or structural variants.

### Cohorts

A total of 85 cohorts were chosen for GPSEA analysis from version 0.1.24 of Phenopacket Store.^23^ The cohorts had a mean of 77.8 individuals (median 49, minimum 16, maximum 462). The cohorts comprised information in 6613 individuals. Information on the sex of participants was available for 82.8% of these individuals, with 53% being male and 47% female.

To use GPSEA with new cohorts, it will be necessary to convert data to GA4GH Phenopacket Schema format. Several software tools are available to streamline this process.^23,24^

### Search for previously published genotype phenotype correlations

For each of the gene-specific cohorts, we searched for publications that described genotype phenotype correlations in PubMed. Each search was designed as {Disease name synonyms} AND {gene/variant synonyms} AND {genotype-phenotype correlation}. The following is an example for Loeys-Dietz syndrome type 3.

(“Loeys-Dietz syndrome type 3” OR LDS3 OR "Loeys-Dietz syndrome 3") AND (SMAD3 OR variant OR mutation) AND

("genotype phenotype correlation" OR "phenotype genotype correlation")

Additionally, the relevant entries from Online Mendelian Inheritance in Man (OMIM)^41^ were consulted as were the publications used for curation.

## Acknowledgements

This work was supported by grants from the the National Human Genome Research Institute (A phenomics-first resource for interpretation of variants; 5RM1HG010860 and The Human Phenotype Ontology: Accelerating Computational Integration of Clinical Data for Genomics; 5U24HG011449). J.X.C. and A.J.M were supported by 1R35HG011297. P.N.R. was supported by a Professorship of the Alexander von Humboldt Foundation. AK and OV were supported by the grant NU23-05-00097 issued by the Czech Health Research Council, Ministry of Health of the Czech Republic.

## Author contributions

P.N.R. and D.D. conceived and designed the project and methodology. L.R. and D.D. wrote the Python code with contributions from P.N.R., J.R. and F.R.. P.N.R. developed the statistical modeling with input from M.I.L.. The cohorts were analyzed by L.R., A.S.L.G, V.B., A.C.-O., P.C., L.C., J.X.C., E.C., A.J.M.D., B.B.A.V., M.H.D., P.G., P.H., A.K., M.S.L., A.M., A.J.M., J.R., C.S, T.S, O.V., D.Z. and P.N.R. using the GPSEA software. M.H., T.G., J.O.B.J., C.J.M., M-M.-T., S.T. and D.S. advised about the use and creation of phenopackets. D.D. and P.N.R. wrote the manuscript, and all authors reviewed and approved the final version.

## Competing interests

The authors declare no competing interests

## Data availability

No original data was generated for this analysis. The phenopackets used are available from the Phenopacket Store (https://github.com/monarch-initiative/phenopacket-store).

## Code availability

GPSEA is available at https://github.com/P2GX/gpsea under an MIT license. Documentation and a tutorial are provided at https://p2gx.github.io/gpsea/stable/.

The GPSEA case studies (gpsea-cs) repository provides one Jupyter notebook for each of the cohorts analyzed in this work, and is available at https://github.com/P2GX/gpsea-cs. This repository additionally contains code we used to generate the supplemental Figures and Tables that is not needed by new users of GPSEA.

## Notes

### Competing Interest Statement

The authors have declared no competing interest.

### Author Declarations

Genotypic and phenotypic data about individuals with rare Mendelian disease were derived from the Phenopacket Store repository.23 Version 0.1.24 of Phenopacket Store includes 8182 phenopackets representing 491 Mendelian and chromosomal diseases associated with 457 genes and 4469 unique pathogenic alleles curated from 1227 different publications. The data is available from https://github.com/monarch-initiative/phenopacket-store

