## Supplementary material for "GA4GH Phenopacket-Driven Characterization of Genotype-Phenotype Correlations in Mendelian Disorders": Online supplement

| Name | Match criterion | Example |
|---|---|---|
| variant key | Specific variant | 1_8364773_8364773_A_AC (NM_001042681.2:c.1512dup) |
| variant effect | Leads to the target effect | START_LOST |
| variant class | Variant has the target type | SNV, DEL, DUP, INS, INV |
| gene | Variant affects gene | Affects *RERE* |
| transcript | Variant affects transcript | Overlaps NM_12345.01 |
| exon | Variant overlaps exon | Located in exon 3 |
| region | Variant overlaps region on protein | Located between amino acid residues 1 to 77 |
| is large imprecise sv | Variant is structural variant without exact breakpoint coordinates | – |
| is structural variant | The variant affects at least n base pairs (n=50 by default) or is a large imprecise SV or a translocation | – |
| structural type | The target ontology class for a variant with imprecise/unknown breakpoints | Chromosomal deletion (SO:1000029) |
| is structural deletion | Variant is a Chromosomal deletion (SO:1000029) or an SV with known breakpoints that deletes at least a $n$ base pairs (`n=50bp` by default) | – |
| ref length | Length of the reference sequence is above, below, or (not) equal to $n$ bases | $n > 50$ for variants affecting more than 50 bp |
| change length | Change of length between the REF/ALT is above, below, or (not) equal to `n` bases | $n \leq -50$ for testing if a variant removes at least 50 base pairs |
| protein feature type | Variant affects protein feature type | DOMAIN |
| protein feature | Variant region overlaps a protein feature | Overlaps with "Bipartite nuclear localization signal" |
| allof | Logical AND for two or more predicates to test if *all* predicates match | missense AND exon 7 |
| anyof | Logical OR for two or more predicates to test if *any* predicate matches | START_LOST OR STOP_GAINED |
| not | Logical NOT | NOT(missense) |

**Table S1: Variant predicates.** Version 0.9.7 of GPSEA offers the variant predicates shown in this table. Each predicate evaluates a variant and returns `True` or `False`.

**Table S2:** Fischer exact test for association between genotypes and phenotypic features. ∁: set complement of a variant predicate. E.g. ∁ for a "missense" predicate includes any mutation that is *not* missense.

| Cohort | HPO | Genotype A | | Genotype B | | p-val | adj. p |
|---|---|---|---|---|---|---|---|
| BRD4 | Intrauterine growth retardation [HP:0001511] | NIPBL | 37/45 (82%) | BRD4 | 2/9 (22%) | $9.4 \times 10^{-04}$ | 0.032 |
| EHMT1 | Attention deficit hyperactivity disorder [HP:0007018] | N Term Frameshift | 4/8 (50%) | ∁ | 3/96 (3%) | $4.8 \times 10^{-04}$ | 0.029 |
| FBN1 | Disproportionate tall stature [HP:0001519] | TB domain | 2/40 (5%) | cbEGF | 12/43 (28%) | 0.007 | 0.029 |
| FBN1 | Ectopia lentis [HP:0001083] | TB domain | 9/18 (50%) | cbEGF | 48/59 (81%) | 0.013 | 0.038 |
| FBN1 | Mitral valve prolapse [HP:0001634] | TB domain | 1/28 (4%) | cbEGF | 13/47 (28%) | 0.012 | 0.038 |
| FBN1 | Proportionate short stature [HP:0003508] | TB domain | 20/36 (56%) | cbEGF | 0/24 (0%) | $2.3 \times 10^{-06}$ | $2.3 \times 10^{-05}$ |
| FBN1 | Severe short stature [HP:0003510] | TB domain | 15/36 (42%) | cbEGF | 0/24 (0%) | $1.4 \times 10^{-04}$ | $9.0 \times 10^{-04}$ |
| FBN1 | Short stature [HP:0004322] | TB domain | 23/39 (59%) | cbEGF | 0/24 (0%) | $4.4 \times 10^{-07}$ | $8.8 \times 10^{-06}$ |
| FBN1 | Tall stature [HP:0000098] | TB domain | 7/38 (18%) | cbEGF | 21/40 (52%) | 0.002 | 0.011 |
| FBN1 | Arachnodactyly [HP:0001166] | exon 37 | 0/8 (0%) | ∁ | 52/95 (55%) | 0.003 | 0.019 |
| FBN1 | Ectopia lentis [HP:0001083] | exon 37 | 1/9 (11%) | ∁ | 78/100 (78%) | $1.1 \times 10^{-04}$ | $1.0 \times 10^{-03}$ |
| FBN1 | Stiff skin [HP:0030053] | exon 37 | 8/9 (89%) | ∁ | 0/50 (0%) | $4.1 \times 10^{-09}$ | $8.5 \times 10^{-08}$ |
| FBN1 | Hyperextensibility of the finger joints [HP:0001187] | fs last two | 2/2 (100%) | ∁ | 1/90 (1%) | $7.2 \times 10^{-04}$ | 0.012 |
| FBN1 | Arachnodactyly [HP:0001166] | missense | 34/81 (42%) | ∁ | 18/22 (82%) | $1.0 \times 10^{-03}$ | 0.026 |
| FBN1 | Hyperextensibility of the finger joints [HP:0001187] | missense | 0/78 (0%) | ∁ | 3/14 (21%) | 0.003 | 0.026 |
| FBN1 | Thoracic aortic aneurysm [HP:0012727] | missense | 25/64 (39%) | ∁ | 12/14 (86%) | 0.002 | 0.026 |
| FBXL4 | Feeding difficulties [HP:0011968] | missense/ ∁ OR ∁/ ∁ | 23/24 (96%) | missense/ missense | 13/27 (48%) | $1.7 \times 10^{-04}$ | 0.010 |
| FGD1 | Broad foot [HP:0001769] | missense | 1/7 (14%) | ∁ | 14/15 (93%) | $6.2 \times 10^{-04}$ | 0.032 |
| GLI3 | Anal atresia [HP:0002023] | Fs in mid region or splice | 7/25 (28%) | ∁ | 3/57 (5%) | 0.007 | 0.036 |
| GLI3 | Nail dysplasia [HP:0002164] | Fs in mid region or splice | 7/20 (35%) | ∁ | 2/54 (4%) | $1.0 \times 10^{-03}$ | 0.009 |
| GLI3 | Preaxial foot polydactyly [HP:0001841] | Fs in mid region or splice | 5/25 (20%) | ∁ | 32/57 (56%) | 0.003 | 0.021 |
| GLI3 | Y-shaped metacarpals [HP:0006042] | Fs in mid region or splice | 11/23 (48%) | ∁ | 4/56 (7%) | $9.9 \times 10^{-05}$ | $1.0 \times 10^{-03}$ |
| GLI3 | Y-shaped metatarsals [HP:0010567] | Fs in mid region or splice | 11/19 (58%) | ∁ | 4/50 (8%) | $3.2 \times 10^{-05}$ | $7.8 \times 10^{-04}$ |
| GLI3 | Anal atresia [HP:0002023] | Truncating variants in Exon 15 | 8/28 (29%) | ∁ | 2/54 (4%) | 0.002 | 0.017 |

Continued on next page

**Table S2 – continued from previous page**

| Cohort | HPO | Genotype A | | Genotype B | | p-val | adj. p |
|---|---|---|---|---|---|---|---|
| GLI3 | Macrocephaly [HP:0000256] | Truncating variants in Exon 15 | 13/16 (81%) | C | 15/42 (36%) | 0.003 | 0.017 |
| GLI3 | Postaxial foot polydactyly [HP:0001830] | Truncating variants in Exon 15 | 9/20 (45%) | C | 2/35 (6%) | $8.9 \times 10^{-04}$ | 0.011 |
| GLI3 | Preaxial foot polydactyly [HP:0001841] | Truncating variants in Exon 15 | 7/28 (25%) | C | 30/54 (56%) | 0.010 | 0.050 |
| GLI3 | Syndactyly [HP:0001159] | Truncating variants in Exon 15 | 5/17 (29%) | C | 33/38 (87%) | $4.8 \times 10^{-05}$ | $1.0 \times 10^{-03}$ |
| GLI3 | Macrocephaly [HP:0000256] | Variants in C-terminal third | 13/15 (87%) | C | 15/43 (35%) | $7.3 \times 10^{-04}$ | 0.004 |
| GLI3 | Postaxial foot polydactyly [HP:0001830] | Variants in C-terminal third | 9/16 (56%) | C | 2/39 (5%) | $7.3 \times 10^{-05}$ | 0.002 |
| GLI3 | Postaxial hand polydactyly [HP:0001162] | Variants in C-terminal third | 15/16 (94%) | C | 21/49 (43%) | $3.4 \times 10^{-04}$ | 0.003 |
| GLI3 | Syndactyly [HP:0001159] | Variants in C-terminal third | 5/16 (31%) | C | 33/39 (85%) | $2.2 \times 10^{-04}$ | 0.003 |
| IKZF1 | Recurrent pneumonia [HP:0006532] | p.Asn159Ser | 7/9 (78%) | C | 7/39 (18%) | $1.0 \times 10^{-03}$ | 0.035 |
| IKZF1 | T-cell acute lymphoblastic leukemias [HP:0006727] | p.Asn159Ser | 3/13 (23%) | C | 0/69 (0%) | 0.003 | 0.047 |
| ITPR1 | Aniridia [HP:0000526] | GS Hotspot | 14/20 (70%) | C | 6/48 (12%) | $6.1 \times 10^{-06}$ | $1.6 \times 10^{-04}$ |
| ITPR1 | Delayed ability to sit [HP:0025336] | GS Hotspot | 13/13 (100%) | C | 40/68 (59%) | 0.003 | 0.030 |
| ITPR1 | Delayed ability to walk [HP:0031936] | GS Hotspot | 14/14 (100%) | C | 40/67 (60%) | 0.003 | 0.030 |
| ITPR1 | Delayed gross motor development [HP:0002194] | GS Hotspot | 18/18 (100%) | C | 57/83 (69%) | 0.005 | 0.032 |
| ITPR1 | Motor delay [HP:0001270] | GS Hotspot | 19/19 (100%) | C | 68/94 (72%) | 0.006 | 0.032 |
| ITPR1 | Aniridia [HP:0000526] | IP3 binding | 0/14 (0%) | C | 20/54 (37%) | 0.007 | 0.024 |
| ITPR1 | Delayed ability to sit [HP:0025336] | IP3 binding | 11/11 (100%) | C | 42/70 (60%) | 0.013 | 0.043 |
| ITPR1 | Delayed ability to walk [HP:0031936] | IP3 binding | 13/13 (100%) | C | 41/68 (60%) | 0.004 | 0.019 |
| ITPR1 | Delayed gross motor development [HP:0002194] | IP3 binding | 18/18 (100%) | C | 57/83 (69%) | 0.005 | 0.024 |
| ITPR1 | Delayed speech and language development [HP:0000750] | IP3 binding | 13/13 (100%) | C | 37/63 (59%) | 0.003 | 0.019 |
| ITPR1 | Motor delay [HP:0001270] | IP3 binding | 24/24 (100%) | C | 63/89 (71%) | 0.002 | 0.019 |
| ITPR1 | Neurodevelopmental delay [HP:0012758] | IP3 binding | 29/29 (100%) | C | 70/90 (78%) | 0.003 | 0.019 |
| ITPR1 | Nystagmus [HP:0000639] | IP3 binding | 25/26 (96%) | C | 53/78 (68%) | 0.003 | 0.019 |
| ITPR1 | Delayed ability to sit [HP:0025336] | SV Deletion | 0/19 (0%) | C | 53/62 (85%) | $4.6 \times 10^{-12}$ | $1.7 \times 10^{-11}$ |

Continued on next page

Table S2 – continued from previous page

| Cohort | HPO | Genotype A | | Genotype B | | p-val | adj. p |
|---|---|---|---|---|---|---|---|
| ITPR1 | Delayed ability to walk [HP:0031936] | SV Deletion | 0/19 (0%) | C | 54/62 (87%) | $1.5 \times 10^{-12}$ | $7.6 \times 10^{-12}$ |
| ITPR1 | Delayed gross motor development [HP:0002194] | SV Deletion | 0/19 (0%) | C | 75/82 (91%) | $4.0 \times 10^{-15}$ | $3.0 \times 10^{-14}$ |
| ITPR1 | Delayed speech and language development [HP:0000750] | SV Deletion | 0/19 (0%) | C | 50/57 (88%) | $1.7 \times 10^{-12}$ | $7.6 \times 10^{-12}$ |
| ITPR1 | Global developmental delay [HP:0001263] | SV Deletion | 0/19 (0%) | C | 48/57 (84%) | $1.8 \times 10^{-11}$ | $5.7 \times 10^{-11}$ |
| ITPR1 | Motor delay [HP:0001270] | SV Deletion | 0/19 (0%) | C | 87/94 (93%) | $3.9 \times 10^{-16}$ | $4.3 \times 10^{-15}$ |
| ITPR1 | Neurodevelopmental delay [HP:0012758] | SV Deletion | 0/19 (0%) | C | 99/100 (99%) | $4.1 \times 10^{-21}$ | $9.0 \times 10^{-20}$ |
| ITPR1 | Nystagmus [HP:0000639] | SV Deletion | 17/17 (100%) | C | 61/87 (70%) | 0.006 | 0.016 |
| KCNH5 | Epileptic encephalopathy [HP:0200134] | Arg327His | 15/15 (100%) | Arg333His | 0/3 (0%) | $1.0 \times 10^{-03}$ | 0.032 |
| KDM6A | Pulmonic stenosis [HP:0001642] | p.Asn891ValfsTer22 | [illegible] (100%) | C | 0/59 (0%) | $5.5 \times 10^{-04}$ | 0.016 |
| LMNA | Distal muscle weakness [HP:0002460] | Gly608= | 0/15 (0%) | C | 36/97 (37%) | 0.002 | 0.004 |
| LMNA | Elevated hemoglobin A1c [HP:0040217] | Gly608= | 0/15 (0%) | C | 79/110 (72%) | $5.7 \times 10^{-08}$ | $6.8 \times 10^{-07}$ |
| LMNA | Limb muscle weakness [HP:0003690] | Gly608= | 0/15 (0%) | C | 38/99 (38%) | 0.002 | 0.004 |
| LMNA | Lipodystrophy [HP:0009125] | Gly608= | 15/15 (100%) | C | 127/236 (54%) | $1.9 \times 10^{-04}$ | $7.5 \times 10^{-04}$ |
| LMNA | Muscle weakness [HP:0001324] | Gly608= | 0/15 (0%) | C | 63/124 (51%) | $6.1 \times 10^{-05}$ | $3.6 \times 10^{-04}$ |
| LMNA | Proximal muscle weakness [HP:0003701] | Gly608= | 0/15 (0%) | C | 53/114 (46%) | $3.6 \times 10^{-04}$ | $1.0 \times 10^{-03}$ |
| LMNA | Proximal muscle weakness in upper limbs [HP:0008997] | Gly608= | 0/15 (0%) | C | 35/102 (34%) | 0.005 | 0.007 |
| LMNA | Upper limb muscle weakness [HP:0003484] | Gly608= | 0/15 (0%) | C | 35/96 (36%) | 0.003 | 0.004 |
| LMNA | Achilles tendon contracture [HP:0001771] | Upstream Tail (1-383) | 17/81 (21%) | C | 23/34 (68%) | $3.4 \times 10^{-06}$ | $1.4 \times 10^{-05}$ |
| LMNA | Atrioventricular block [HP:0001678] | Upstream Tail (1-383) | 17/35 (49%) | C | 8/117 (7%) | $1.2 \times 10^{-07}$ | $1.5 \times 10^{-06}$ |
| LMNA | Dilated cardiomyopathy [HP:0001644] | Upstream Tail (1-383) | 35/69 (51%) | C | 5/103 (5%) | $2.2 \times 10^{-12}$ | $4.2 \times 10^{-11}$ |
| LMNA | Elbow contracture [HP:0034391] | Upstream Tail (1-383) | 17/79 (22%) | C | 22/30 (73%) | $9.6 \times 10^{-07}$ | $7.3 \times 10^{-06}$ |
| LMNA | First degree atrioventricular block [HP:0011705] | Upstream Tail (1-383) | 7/62 (11%) | C | 2/116 (2%) | 0.009 | 0.032 |
| LMNA | Foot joint contracture [HP:0008366] | Upstream Tail (1-383) | 17/77 (22%) | C | 23/31 (74%) | $6.7 \times 10^{-07}$ | $6.4 \times 10^{-06}$ |
| LMNA | Hip contracture [HP:0003273] | Upstream Tail (1-383) | 4/79 (5%) | C | 9/30 (30%) | $1.0 \times 10^{-03}$ | 0.004 |
| LMNA | Limb joint contracture [HP:0003121] | Upstream Tail (1-383) | 19/79 (24%) | C | 23/31 (74%) | $2.1 \times 10^{-06}$ | $9.8 \times 10^{-06}$ |

Continued on next page

Table S2 – continued from previous page

| Cohort | HPO | Genotype A | | Genotype B | | p-val | adj. p |
|---|---|---|---|---|---|---|---|
| LMNA | Lipodystrophy [HP:0009125] | Upstream Tail (1-383) | 9/88 (10%) | C | 133/163 (82%) | $1.7 \times 10^{-29}$ | $6.5 \times 10^{-28}$ |
| LMNA | Lower-limb joint contracture [HP:0005750] | Upstream Tail (1-383) | 18/78 (23%) | C | 23/31 (74%) | $1.5 \times 10^{-06}$ | $8.4 \times 10^{-06}$ |
| LMNA | Second degree atrioventricular block [HP:0011706] | Upstream Tail (1-383) | 6/67 (9%) | C | 1/116 (1%) | 0.010 | 0.032 |
| LMNA | Upper-limb joint contracture [HP:0100360] | Upstream Tail (1-383) | 17/77 (22%) | C | 22/30 (73%) | $1.4 \times 10^{-06}$ | $8.4 \times 10^{-06}$ |
| LMNA | Achilles tendon contracture [HP:0001771] | Upstream of NLS | 21/85 (25%) | C | 19/30 (63%) | $2.7 \times 10^{-04}$ | $1.0 \times 10^{-03}$ |
| LMNA | Atrioventricular block [HP:0001678] | Upstream of NLS | 17/36 (47%) | C | 8/116 (7%) | $2.1 \times 10^{-07}$ | $2.7 \times 10^{-06}$ |
| LMNA | Dilated cardiomyopathy [HP:0001644] | Upstream of NLS | 35/70 (50%) | C | 5/102 (5%) | $4.2 \times 10^{-12}$ | $8.0 \times 10^{-11}$ |
| LMNA | Elbow contracture [HP:0034391] | Upstream of NLS | 21/83 (25%) | C | 18/26 (69%) | $1.0 \times 10^{-04}$ | $6.6 \times 10^{-04}$ |
| LMNA | First degree atrioventricular block [HP:0011705] | Upstream of NLS | 7/63 (11%) | C | 2/115 (2%) | 0.010 | 0.033 |
| LMNA | Foot joint contracture [HP:0008366] | Upstream of NLS | 21/81 (26%) | C | 19/27 (70%) | $6.2 \times 10^{-05}$ | $5.9 \times 10^{-04}$ |
| LMNA | Hip contracture [HP:0003273] | Upstream of NLS | 4/83 (5%) | C | 9/26 (35%) | $2.7 \times 10^{-04}$ | $1.0 \times 10^{-03}$ |
| LMNA | Limb joint contracture [HP:0003121] | Upstream of NLS | 23/83 (28%) | C | 19/27 (70%) | $1.7 \times 10^{-04}$ | $8.1 \times 10^{-04}$ |
| LMNA | Lipodystrophy [HP:0009125] | Upstream of NLS | 10/93 (11%) | C | 132/158 (84%) | $1.7 \times 10^{-31}$ | $6.6 \times 10^{-30}$ |
| LMNA | Lower-limb joint contracture [HP:0005750] | Upstream of NLS | 22/82 (27%) | C | 19/27 (70%) | $8.2 \times 10^{-05}$ | $6.2 \times 10^{-04}$ |
| LMNA | Pancreatitis [HP:0001733] | Upstream of NLS | 4/7 (57%) | C | 14/110 (13%) | 0.011 | 0.033 |
| LMNA | Second degree atrioventricular block [HP:0011706] | Upstream of NLS | 6/68 (9%) | C | 1/115 (1%) | 0.011 | 0.033 |
| LMNA | Upper-limb joint contracture [HP:0100360] | Upstream of NLS | 21/81 (26%) | C | 18/26 (69%) | $1.2 \times 10^{-04}$ | $6.7 \times 10^{-04}$ |
| LMNA | Elevated hemoglobin A1c [HP:0040217] | missense | 73/101 (72%) | C | 6/24 (25%) | $3.1 \times 10^{-05}$ | $5.9 \times 10^{-04}$ |
| LMNA | Loss of truncal subcutaneous adipose tissue [HP:0009002] | missense | 104/104 (100%) | C | 4/11 (36%) | $7.5 \times 10^{-09}$ | $2.9 \times 10^{-07}$ |
| MPV17 | Peripheral axonal neuropathy [HP:0003477] | Pro98Leu/ Pro98Leu | 3/3 (100%) | C/C OR Pro98Leu/C | 1/22 (5%) | 0.002 | 0.037 |
| NBAS | Decreased circulating IgG concentration [HP:0004315] | missense/ missense | 1/12 (8%) | C/ C OR missense/ C | 15/23 (65%) | 0.002 | 0.035 |
| NF1 | Axillary freckling [HP:0000997] | Leu847Pro | 50/61 (82%) | C | 75/193 (39%) | $3.5 \times 10^{-09}$ | $7.8 \times 10^{-08}$ |
| NF1 | Freckling [HP:0001480] | Leu847Pro | 54/54 (100%) | C | 223/301 (74%) | $5.3 \times 10^{-07}$ | $5.8 \times 10^{-06}$ |
| NF1 | Inguinal freckling [HP:0030052] | Leu847Pro | 34/60 (57%) | C | 58/189 (31%) | $3.9 \times 10^{-04}$ | 0.002 |

Continued on next page

**Table S2 – continued from previous page**

| Cohort | HPO | Genotype A | | Genotype B | | p-val | adj. p |
|---|---|---|---|---|---|---|---|
| NF1 | Lisch nodules [HP:0009737] | Leu847Pro | 22/42 (52%) | C | 61/242 (25%) | $7.5 \times 10^{-04}$ | 0.003 |
| NF1 | Optic nerve glioma [HP:0009734] | Leu847Pro | 15/33 (45%) | C | 24/143 (17%) | $8.6 \times 10^{-04}$ | 0.003 |
| NF1 | Plexiform neurofibroma [HP:0009732] | Leu847Pro | 24/66 (36%) | C | 44/329 (13%) | $4.6 \times 10^{-05}$ | $3.4 \times 10^{-04}$ |
| NF1 | Axillary freckling [HP:0000997] | Met992del | 0/14 (0%) | C | 125/240 (52%) | $8.6 \times 10^{-05}$ | $1.0 \times 10^{-03}$ |
| NF1 | Inguinal freckling [HP:0030052] | Met992del | 0/14 (0%) | C | 92/235 (39%) | 0.003 | 0.008 |
| NF1 | Lipoma [HP:0012032] | Met992del | 5/44 (11%) | C | 4/284 (1%) | 0.003 | 0.008 |
| NF1 | Lisch nodules [HP:0009737] | Met992del | 3/36 (8%) | C | 80/248 (32%) | 0.003 | 0.008 |
| NF1 | Plexiform neurofibroma [HP:0009732] | Met992del | 0/44 (0%) | C | 68/351 (19%) | $2.0 \times 10^{-04}$ | $1.0 \times 10^{-03}$ |
| NF1 | Scoliosis [HP:0002650] | SV | 18/57 (32%) | C | 38/308 (12%) | $9.7 \times 10^{-04}$ | 0.017 |
| NF1 | Axillary freckling [HP:0000997] | p.Arg1830 | 20/79 (25%) | C | 105/175 (60%) | $3.7 \times 10^{-07}$ | $2.0 \times 10^{-06}$ |
| NF1 | Freckling [HP:0001480] | p.Arg1830 | 76/133 (57%) | C | 201/222 (91%) | $6.3 \times 10^{-13}$ | $6.9 \times 10^{-12}$ |
| NF1 | Inguinal freckling [HP:0030052] | p.Arg1830 | 8/79 (10%) | C | 84/170 (49%) | $4.9 \times 10^{-10}$ | $3.6 \times 10^{-09}$ |
| NF1 | Lisch nodules [HP:0009737] | p.Arg1830 | 11/91 (12%) | C | 72/193 (37%) | $6.6 \times 10^{-06}$ | $2.9 \times 10^{-05}$ |
| NF1 | Neurofibroma [HP:0001067] | p.Arg1830 | 5/8 (62%) | C | 153/162 (94%) | 0.012 | 0.030 |
| NF1 | Optic nerve glioma [HP:0009734] | p.Arg1830 | 0/39 (0%) | C | 39/137 (28%) | $1.7 \times 10^{-05}$ | $6.3 \times 10^{-05}$ |
| NF1 | Plexiform neurofibroma [HP:0009732] | p.Arg1830 | 0/135 (0%) | C | 68/260 (26%) | $1.7 \times 10^{-14}$ | $3.8 \times 10^{-13}$ |
| NF1 | Pulmonic stenosis [HP:0001642] | p.Arg1830 | 13/105 (12%) | C | 5/162 (3%) | 0.005 | 0.013 |
| NF1 | Scoliosis [HP:0002650] | p.Arg1830 | 8/125 (6%) | C | 48/240 (20%) | $4.0 \times 10^{-04}$ | $1.0 \times 10^{-03}$ |
| PTPN11 | Hypertelorism [HP:0000316] | missense | 37/41 (90%) | C | 0/12 (0%) | $6.8 \times 10^{-09}$ | $2.7 \times 10^{-08}$ |
| PTPN11 | Intellectual disability, mild [HP:0001256] | missense | 8/23 (35%) | C | 0/12 (0%) | 0.032 | 0.032 |
| PTPN11 | Pulmonic stenosis [HP:0001642] | missense | 18/34 (53%) | C | 0/12 (0%) | $1.0 \times 10^{-03}$ | 0.002 |
| PTPN11 | Webbed neck [HP:0000465] | missense | 15/20 (75%) | C | 0/12 (0%) | $2.9 \times 10^{-05}$ | $5.9 \times 10^{-05}$ |
| RPGRIP1 | Eye poking [HP:0001483] | 1107del/1107del OR 1107del/C | 16/16 (100%) | C/C | 19/41 (46%) | $1.3 \times 10^{-04}$ | 0.002 |
| SAMD9L | Neutropenia [HP:0001875] | Arg986Cys | 7/9 (78%) | Ser626Leu | 0/9 (0%) | 0.002 | 0.008 |
| SAMD9L | Pancytopenia [HP:0001876] | Arg986Cys | 4/6 (67%) | Ser626Leu | 0/9 (0%) | 0.011 | 0.026 |
| SAMD9L | Thrombocytopenia [HP:0001873] | Arg986Cys | 7/9 (78%) | Ser626Leu | 0/9 (0%) | 0.002 | 0.008 |
| SATB2 | Cleft palate [HP:0000175] | missense | 11/49 (22%) | C | 59/105 (56%) | $1.1 \times 10^{-04}$ | 0.002 |
| SCN2A | Intellectual disability [HP:0001249] | I repeat | 21/42 (50%) | C | 157/190 (83%) | $2.6 \times 10^{-05}$ | $4.2 \times 10^{-04}$ |

Continued on next page

**Table S2 – continued from previous page**

| Cohort | HPO | Genotype A | | Genotype B | | p-val | adj. p |
|---|---|---|---|---|---|---|---|
| SCN2A | Neurodevelopmental abnormality [HP:0012759] | I repeat | 48/65 (74%) | C | 198/218 (91%) | $1.0 \times 10^{-03}$ | 0.009 |
| SCN2A | Autism [HP:0000717] | missense | 59/146 (40%) | C | 33/43 (77%) | $2.7 \times 10^{-05}$ | $8.6 \times 10^{-05}$ |
| SCN2A | Focal-onset seizure [HP:0007359] | missense | 141/170 (83%) | C | 8/33 (24%) | $8.7 \times 10^{-11}$ | $6.9 \times 10^{-10}$ |
| SCN2A | Generalized-onset seizure [HP:0002197] | missense | 104/133 (78%) | C | 6/31 (19%) | $1.4 \times 10^{-09}$ | $5.7 \times 10^{-09}$ |
| SCN2A | Intellectual disability [HP:0001249] | missense | 144/198 (73%) | C | 34/34 (100%) | $9.4 \times 10^{-05}$ | $2.5 \times 10^{-04}$ |
| SCN2A | Motor seizure [HP:0020219] | missense | 146/175 (83%) | C | 6/31 (19%) | $4.5 \times 10^{-12}$ | $7.2 \times 10^{-11}$ |
| SCN2A | Neurodevelopmental abnormality [HP:0012759] | missense | 201/238 (84%) | C | 45/45 (100%) | $1.0 \times 10^{-03}$ | 0.003 |
| SCN2A | Seizure [HP:0001250] | missense | 298/327 (91%) | C | 28/53 (53%) | $1.6 \times 10^{-10}$ | $8.4 \times 10^{-10}$ |
| SCO2 | Hypertrophic cardiomyopathy [HP:0001639] | p.Glu140Lys/ p.Glu140Lys | 2/6 (33%) | C/C OR p.Glu140Lys/C | 13/13 (100%) | 0.004 | 0.027 |
| SETD2 | Delayed ability to walk [HP:0031936] | p.Arg1740Trp | 8/8 (100%) | C | 1/10 (10%) | $4.1 \times 10^{-04}$ | 0.003 |
| SETD2 | Hypertelorism [HP:0000316] | p.Arg1740Trp | 11/11 (100%) | C | 5/23 (22%) | $1.5 \times 10^{-05}$ | $2.6 \times 10^{-04}$ |
| SETD2 | Macrocephaly [HP:0000256] | p.Arg1740Trp | 0/11 (0%) | C | 19/28 (68%) | $1.5 \times 10^{-04}$ | 0.002 |
| SETD2 | Scoliosis [HP:0002650] | p.Arg1740Trp | 6/6 (100%) | C | 2/14 (14%) | $7.2 \times 10^{-04}$ | 0.005 |
| SETD2 | Severe global developmental delay [HP:0011344] | p.Arg1740Trp | 9/9 (100%) | C | 0/12 (0%) | $3.4 \times 10^{-06}$ | $1.2 \times 10^{-04}$ |
| SETD2 | Ventriculomegaly [HP:0002119] | p.Arg1740Trp | 4/4 (100%) | C | 2/17 (12%) | 0.003 | 0.012 |
| SETD2 | Wide nasal bridge [HP:0000431] | p.Arg1740Trp | 9/9 (100%) | C | 2/9 (22%) | 0.002 | 0.012 |
| SETD2 | Macrocephaly [HP:0000256] | missense | 4/24 (17%) | C | 15/15 (100%) | $1.5 \times 10^{-07}$ | $6.3 \times 10^{-06}$ |
| SMAD3 | Osteoarthritis [HP:0002758] | p.Arg287Trp | 19/19 (100%) | C | 7/19 (37%) | $3.7 \times 10^{-05}$ | $8.6 \times 10^{-04}$ |
| SMARCB1 | Atypical teratoid/rhabdoid tumor [HP:0034401] | SV | 8/9 (89%) | C | 2/19 (11%) | $1.2 \times 10^{-04}$ | $5.9 \times 10^{-04}$ |
| SMARCB1 | Embryonal neoplasm [HP:0002898] | SV | 8/8 (100%) | C | 2/19 (11%) | $2.0 \times 10^{-05}$ | $1.5 \times 10^{-04}$ |
| SMARCB1 | Neoplasm by anatomical site [HP:0011793] | SV | 3/3 (100%) | C | 3/20 (15%) | 0.011 | 0.028 |
| SMARCB1 | Neoplasm by histology [HP:0011792] | SV | 11/11 (100%) | C | 4/21 (19%) | $1.1 \times 10^{-05}$ | $1.5 \times 10^{-04}$ |
| SMARCB1 | Neuroepithelial neoplasm [HP:0030063] | SV | 2/2 (100%) | C | 0/17 (0%) | 0.006 | 0.018 |
| SMARCB1 | Rhabdoid tumor [HP:0034557] | SV | 4/4 (100%) | C | 2/19 (11%) | 0.002 | 0.006 |
| SMARCC2 | Intellectual disability [HP:0001249] | c.3222del | 1/6 (17%) | C | 49/52 (94%) | $7.0 \times 10^{-05}$ | 0.006 |
| SPTAN1 | Appendicular spasticity [HP:0034353] | Arg19Trp | 21/21 (100%) | C | 2/15 (13%) | $4.5 \times 10^{-08}$ | $1.9 \times 10^{-07}$ |

Continued on next page

Table S2 – continued from previous page

| Cohort | HPO | Genotype A | | Genotype B | | p-val | adj. p |
|---|---|---|---|---|---|---|---|
| SPTAN1 | Distal lower limb muscle weakness [HP:0009053] | Arg19Trp | 8/15 (53%) | C | 1/20 (5%) | 0.002 | 0.004 |
| SPTAN1 | Epileptic spasm [HP:0011097] | Arg19Trp | 0/19 (0%) | C | 15/20 (75%) | $7.7 \times 10^{-07}$ | $2.7 \times 10^{-06}$ |
| SPTAN1 | Infantile spasms [HP:0012469] | Arg19Trp | 0/19 (0%) | C | 15/41 (37%) | $1.0 \times 10^{-03}$ | 0.003 |
| SPTAN1 | Intellectual disability [HP:0001249] | Arg19Trp | 0/21 (0%) | C | 27/34 (79%) | $1.8 \times 10^{-09}$ | $1.5 \times 10^{-08}$ |
| SPTAN1 | Lower limb muscle weakness [HP:0007340] | Arg19Trp | 14/14 (100%) | C | 17/30 (57%) | 0.003 | 0.007 |
| SPTAN1 | Lower limb spasticity [HP:0002061] | Arg19Trp | 21/21 (100%) | C | 0/13 (0%) | $1.1 \times 10^{-09}$ | $1.4 \times 10^{-08}$ |
| SPTAN1 | Microcephaly [HP:0000252] | Arg19Trp | 0/21 (0%) | C | 19/45 (42%) | $2.4 \times 10^{-04}$ | $6.1 \times 10^{-04}$ |
| SPTAN1 | Motor axonal neuropathy [HP:0007002] | Arg19Trp | 0/16 (0%) | C | 12/19 (63%) | $6.3 \times 10^{-05}$ | $1.7 \times 10^{-04}$ |
| SPTAN1 | Motor seizure [HP:0020219] | Arg19Trp | 0/19 (0%) | C | 20/25 (80%) | $3.3 \times 10^{-08}$ | $1.7 \times 10^{-07}$ |
| SPTAN1 | Peripheral axonal neuropathy [HP:0003477] | Arg19Trp | 4/20 (20%) | C | 12/19 (63%) | 0.010 | 0.017 |
| SPTAN1 | Seizure [HP:0001250] | Arg19Trp | 2/21 (10%) | C | 35/40 (88%) | $2.4 \times 10^{-09}$ | $1.5 \times 10^{-08}$ |
| SPTAN1 | Spastic paraplegia [HP:0001258] | Arg19Trp | 21/21 (100%) | C | 0/20 (0%) | $3.7 \times 10^{-12}$ | $9.3 \times 10^{-11}$ |
| SPTAN1 | Spasticity [HP:0001257] | Arg19Trp | 21/21 (100%) | C | 5/18 (28%) | $1.0 \times 10^{-06}$ | $3.3 \times 10^{-06}$ |
| SPTAN1 | Appendicular spasticity [HP:0034353] | missense | 21/22 (95%) | C | 2/14 (14%) | $8.7 \times 10^{-07}$ | $8.1 \times 10^{-06}$ |
| SPTAN1 | Distal lower limb muscle weakness [HP:0009053] | missense | 9/19 (47%) | C | 0/16 (0%) | $1.0 \times 10^{-03}$ | 0.004 |
| SPTAN1 | Epileptic spasm [HP:0011097] | missense | 2/22 (9%) | C | 13/17 (76%) | $2.9 \times 10^{-05}$ | $1.4 \times 10^{-04}$ |
| SPTAN1 | Infantile spasms [HP:0012469] | missense | 2/29 (7%) | C | 13/31 (42%) | 0.002 | 0.005 |
| SPTAN1 | Intellectual disability [HP:0001249] | missense | 9/32 (28%) | C | 18/23 (78%) | $3.4 \times 10^{-04}$ | $1.0 \times 10^{-03}$ |
| SPTAN1 | Lower limb muscle weakness [HP:0007340] | missense | 15/16 (94%) | C | 16/28 (57%) | 0.015 | 0.030 |
| SPTAN1 | Lower limb spasticity [HP:0002061] | missense | 21/22 (95%) | C | 0/12 (0%) | $2.4 \times 10^{-08}$ | $6.6 \times 10^{-07}$ |
| SPTAN1 | Microcephaly [HP:0000252] | missense | 4/34 (12%) | C | 15/32 (47%) | 0.002 | 0.005 |
| SPTAN1 | Motor axonal neuropathy [HP:0007002] | missense | 0/19 (0%) | C | 12/16 (75%) | $2.2 \times 10^{-06}$ | $1.5 \times 10^{-05}$ |
| SPTAN1 | Motor seizure [HP:0020219] | missense | 7/27 (26%) | C | 13/17 (76%) | 0.002 | 0.004 |
| SPTAN1 | Peripheral axonal neuropathy [HP:0003477] | missense | 4/23 (17%) | C | 12/16 (75%) | $6.7 \times 10^{-04}$ | 0.002 |
| SPTAN1 | Seizure [HP:0001250] | missense | 13/33 (39%) | C | 24/28 (86%) | $2.5 \times 10^{-04}$ | $9.9 \times 10^{-04}$ |
| SPTAN1 | Spastic paraplegia [HP:0001258] | missense | 21/25 (84%) | C | 0/16 (0%) | $4.7 \times 10^{-08}$ | $6.6 \times 10^{-07}$ |

Continued on next page

**Table S2 – continued from previous page**

| Cohort | HPO | Genotype A | | Genotype B | | p-val | adj. p |
|---|---|---|---|---|---|---|---|
| SPTAN1 | Spasticity [HP:0001257] | missense | 22/23 (96%) | ∁ | 4/16 (25%) | $5.2 \times 10^{-06}$ | $2.9 \times 10^{-05}$ |
| SPTAN1 | Appendicular spasticity [HP:0034353] | truncating | 0/8 (0%) | ∁ | 23/28 (82%) | $4.3 \times 10^{-05}$ | $2.7 \times 10^{-04}$ |
| SPTAN1 | Hypotonia [HP:0001252] | truncating | 1/11 (9%) | ∁ | 15/24 (62%) | 0.004 | 0.015 |
| SPTAN1 | Lower limb spasticity [HP:0002061] | truncating | 0/8 (0%) | ∁ | 21/26 (81%) | $7.1 \times 10^{-05}$ | $3.5 \times 10^{-04}$ |
| SPTAN1 | Motor axonal neuropathy [HP:0007002] | truncating | 12/12 (100%) | ∁ | 0/23 (0%) | $1.2 \times 10^{-09}$ | $3.0 \times 10^{-08}$ |
| SPTAN1 | Peripheral axonal neuropathy [HP:0003477] | truncating | 12/12 (100%) | ∁ | 4/27 (15%) | $4.6 \times 10^{-07}$ | $5.8 \times 10^{-06}$ |
| SPTAN1 | Spastic paraplegia [HP:0001258] | truncating | 0/8 (0%) | ∁ | 21/33 (64%) | $1.0 \times 10^{-03}$ | 0.005 |
| SPTAN1 | Spasticity [HP:0001257] | truncating | 0/8 (0%) | ∁ | 26/31 (84%) | $2.1 \times 10^{-05}$ | $1.7 \times 10^{-04}$ |
| SUOX | Microcephaly [HP:0000252] | homodimerization/homodimerization OR homodimerization/∁ | 0/9 (0%) | ∁/∁ | 10/12 (83%) | $2.2 \times 10^{-04}$ | 0.003 |
| TBCK | Developmental regression [HP:0002376] | R126*/R126* | 9/12 (75%) | R126*/∁ OR ∁/∁ | 2/22 (9%) | $1.8 \times 10^{-04}$ | 0.003 |
| TBCK | Macroglossia [HP:0000158] | R126*/R126* | 11/12 (92%) | R126*/∁ OR ∁/∁ | 3/22 (14%) | $1.3 \times 10^{-05}$ | $4.3 \times 10^{-04}$ |
| TBX1 | Global developmental delay [HP:0001263] | Tyr418PhefsTer42 | 5/5 (100%) | ∁ | 3/20 (15%) | $1.0 \times 10^{-03}$ | 0.023 |
| TBX1 | Narrow nose [HP:0000460] | Tyr418PhefsTer42 | 5/5 (100%) | ∁ | 0/6 (0%) | 0.002 | 0.024 |
| TBX5 | Upper limb phocomelia [HP:0009813] | Arg237Gln | 7/22 (32%) | ∁ | 3/131 (2%) | $4.6 \times 10^{-05}$ | $7.3 \times 10^{-04}$ |
| TBX5 | Ventricular septal defect [HP:0001629] | Arg237Gln | 0/17 (0%) | ∁ | 61/73 (84%) | $5.6 \times 10^{-11}$ | $1.8 \times 10^{-09}$ |
| TBX5 | Ventricular septal defect [HP:0001629] | missense | 31/60 (52%) | ∁ | 30/30 (100%) | $4.6 \times 10^{-07}$ | $1.5 \times 10^{-05}$ |
| TGFBR1 | Self-healing squamous epithelioma [HP:0034720] | Gly52Arg | 7/7 (100%) | ∁ | 11/33 (33%) | 0.002 | 0.012 |
| TGFBR1 | Arterial tortuosity [HP:0005116] | MSSE var | 1/19 (5%) | ∁ | 8/16 (50%) | 0.005 | 0.026 |
| TGFBR1 | Hypertelorism [HP:0000316] | MSSE var | 0/18 (0%) | ∁ | 15/19 (79%) | $5.0 \times 10^{-07}$ | $4.0 \times 10^{-06}$ |
| TGFBR1 | Self-healing squamous epithelioma [HP:0034720] | MSSE var | 18/19 (95%) | ∁ | 0/21 (0%) | $1.7 \times 10^{-10}$ | $2.7 \times 10^{-09}$ |
| UMOD | Hyperuricemia [HP:0002149] | EGF | 14/32 (44%) | ∁ | 50/57 (88%) | $1.8 \times 10^{-05}$ | $1.1 \times 10^{-04}$ |
| UMOD | Hyperuricemia [HP:0002149] | cysteine | 38/41 (93%) | ∁ | 26/48 (54%) | $4.5 \times 10^{-05}$ | $2.7 \times 10^{-04}$ |
| WWOX | Bilateral tonic-clonic seizure with focal onset [HP:0007334] | MAPT Interaction/MAPT Interaction OR MAPT Interaction/∁ | 0/18 (0%) | ∁/∁ | 7/9 (78%) | $4.1 \times 10^{-05}$ | 0.002 |
| ZFX | Hyperparathyroidism [HP:0000843] | missense | 7/9 (78%) | ∁ | 0/5 (0%) | 0.021 | 0.021 |

Continued on next page

**Table S2 – continued from previous page**

| Cohort | HPO | Genotype A | | Genotype B | | p-val | adj. p |
|---|---|---|---|---|---|---|---|
| ZMYM3 | Cupped ear [HP:0000378] | R441 | 7/10 (70%) | C | 1/23 (4%) | $2.0 \times 10^{-04}$ | 0.012 |

| cohort | genotype (A) | genotype (B) | Scorer | p-val | xrefs |
|---|---|---|---|---|---|
| ANKRD11 | FEMALE | MALE | HPO Group Count | 0.007 | - |
| ANKRD11 | SV | C | HPO Group Count | $2.64 \times 10^{-4}$ | [1] |
| CHD8 | FEMALE | MALE | De Vries Score | 0.006 | - |
| CHD8 | missense | C | De Vries Score | $8.99 \times 10^{-04}$ | - |
| CTCF | missense | C | De Vries Score | 0.009 | - |
| LMNA | Upstream of NLS | C | HPO Group Count | $1.83 \times 10^{-16}$ | - |
| RERE | LoF | Atrophin | HPO Group Count | 0.001 | - |

**Table S3:** Phenotype severity scores. Mann-Whitney U tests performed using GPSEA to assess the association between a genotype and the total value of a phenotype severity score. The references in the xrefs column show previous publications that have presented similar findings. HPO Group Count scorer assigns a phenotype score that is equivalent to the count of present phenotypes that are either an exact match to the query terms or their descendants. DeVries scorer is an adaption of the DeVries score [2] using HPO. C: set complement of a variant predicate. See Table S2 for a definition.

| cohort | genotype (A) | genotype (B) | Outcome Variable | p-val | xrefs |
|---|---|---|---|---|---|
| ACADM | K329Q: 1/1 | $1/\complement$ OR $\complement/\complement$ | MCAD Activity% [LOINC:74892-1] | $6.1 \times 10^{-10}$ | [3] |
| ACADM | Y67H: 1/1 OR $1/\complement$ | $\complement/\complement$ | MCAD Activity% [LOINC:74892-1] | $2.0 \times 10^{-05}$ | [3] |
| CYP21A2 | missense: $1/\complement$ OR $\complement/\complement$ | 1/1 | 17-Hydroxyprogesterone [LOINC:1668-3] | $7.9 \times 10^{-06}$ | - |

**Table S4: Student t-tests performed using GPSEA**. $\complement$: set complement of a variant predicate. See Table S2 for a definition. Citations in the xrefs column show previous publications that have presented similar findings.

| cohort | genotype (A) | genotype (B) | HPO Term | p-val | xrefs |
|---|---|---|---|---|---|
| AIRE | R257*/R257* OR R257*/C | C/C | Survival analysis: Chronic mucocutaneous candidiasis | 0.019 | - |
| CLDN16 | missense/missense OR missense/C | C/C | Survival analysis: Stage 5 chronic kidney disease | 0.034 | - |
| UMOD | Cys248Trp | Gln316Pro | Survival analysis: Stage 5 chronic kidney disease | $4.1 \times 10^{-04}$ | - |

**Table S5: Age of onset of phenotypic abnormality**. Logrank tests performed using GPSEA to assess association between a genotype and the age of onset of a phenotypic feature represented by an HPO term. C: set complement of a variant predicate. See Table S2 for a definition. Citations in the xrefs column show previous publications that have presented similar findings.

| cohort | genotype (A) | genotype (B) | Disease onset | p-val | xrefs |
|---|---|---|---|---|---|
| CNTNAP2 | 1 allele | 2 alleles | Compute time until OMIM:610042 onset | $6.2 \times 10^{-06}$ | - |
| FBXL4 | missense/C OR C/C | missense/missense | Compute time until OMIM:615471 onset | 0.031 | - |
| HMGCS2 | missense/missense OR missense/C | C/C | Compute time until OMIM:605911 onset | 0.038 | - |
| MPV17 | missense/missense OR missense/C | C/C | Compute time until OMIM:256810 onset | 0.002 | - |
| SETD2 | Missense | C | Compute time until OMIM:616831 onset | $8.5 \times 10^{-05}$ | - |
| SUOX | Missense/Missense OR Missense/C | C/C | Compute time until OMIM:272300 onset | $9.2 \times 10^{-06}$ | - |

**Table S6: Age of onset of disease**. Log rank tests performed using GPSEA to assess association between a genotype and the age of onset of a disease. 1/1, 1/C, C/C: See Table S2 for definitions. Citations in the xrefs column show previous publications that have presented similar findings. hdim: homodimerization; OMIM:620465: Epilepsy, early-onset, 3, with or without developmental delay; OMIM:248250: Hypomagnesemia 3, renal; OMIM:610042: Pitt-Hopkins like syndrome 1; OMIM:130050: Ehlers-Danlos syndrome, vascular type; OMIM:615471: Mitochondrial DNA depletion syndrome 13 (encephalomyopathic type); OMIM:605911: HMG-CoA synthase-2 deficiency; OMIM:256810: Mitochondrial DNA depletion syndrome 6 (hepatocerebral type); OMIM:604377: Mitochondrial complex IV deficiency, nuclear type 2; OMIM:616831: Luscan-Lumish syndrome; OMIM:272300: Sulfite oxidase deficiency.

| cohort | genotype (A) | genotype (B) | Disease | p-val | xrefs |
|---|---|---|---|---|---|
| MPV17 | Pro98Leu: 1/1 | Pro98Leu: 1/C or C/C | OMIM:256810 | 0.010 | - |

**Table S7: Age of death**. Log rank tests performed using GPSEA to assess association between a genotype and the age of death of individuals with a disease. 1/1, 1/C, C/C: See Table S2 for definitions. Citations in the xrefs column show previous publications that have presented similar findings. OMIM:615471: Mitochondrial DNA depletion syndrome 13 (encephalomyopathic type); OMIM:256810: Mitochondrial DNA depletion syndrome 6 (hepatocerebral type); OMIM:614800: Short stature, optic nerve atrophy, and Pelger-Huet anomaly.

| Cohort | HPO | disease A | | disease B | | p-val | adj. p |
|---|---|---|---|---|---|---|---|
| ATP13A2 | Bradykinesia [HP:0002067] | OMIM:606693 | 30/32 (94%) | OMIM:617225 | 4/10 (40%) | $9.2 \times 10^{-04}$ | 0.012 |
| ATP13A2 | Parkinsonism [HP:0001300] | OMIM:606693 | 28/28 (100%) | OMIM:617225 | 3/11 (27%) | $2.7 \times 10^{-06}$ | $7.2 \times 10^{-05}$ |
| Kabuki | Feeding difficulties [HP:0011968] | OMIM:147920 | 8/25 (32%) | OMIM:300867 | 55/63 (87%) | $7.4 \times 10^{-07}$ | $2.1 \times 10^{-05}$ |
| Kabuki | Motor delay [HP:0001270] | OMIM:147920 | 4/10 (40%) | OMIM:300867 | 58/61 (95%) | $1.0 \times 10^{-04}$ | $1.0 \times 10^{-03}$ |
| LDS 1 and 3 | Aortic aneurysm [HP:0004942] | OMIM:609192 | 11/11 (100%) | OMIM:613795 | 26/48 (54%) | 0.004 | 0.024 |
| LDS 1 and 3 | Hypertelorism [HP:0000316] | OMIM:609192 | 15/19 (79%) | OMIM:613795 | 13/35 (37%) | 0.004 | 0.024 |
| LDS 1 and 3 | Osteoarthritis [HP:0002758] | OMIM:609192 | 0/11 (0%) | OMIM:613795 | 26/38 (68%) | $4.6 \times 10^{-05}$ | $9.7 \times 10^{-04}$ |
| LDS 1 and 3 | Scoliosis [HP:0002650] | OMIM:609192 | 18/21 (86%) | OMIM:613795 | 20/43 (47%) | 0.003 | 0.024 |
| LDS 3 and 6 | Thoracic aortic aneurysm [HP:0012727] | OMIM:613795 | 0/22 (0%) | OMIM:619656 | 10/16 (62%) | $1.7 \times 10^{-05}$ | $3.2 \times 10^{-04}$ |
| RPGRIP1 | Nystagmus [HP:0000639] | OMIM:613826 | 64/66 (97%) | OMIM:608194 | 11/16 (69%) | 0.003 | 0.020 |
| RPGRIP1 | Very low visual acuity [HP:0032122] | OMIM:613826 | 35/39 (90%) | OMIM:608194 | 4/16 (25%) | $5.2 \times 10^{-06}$ | $7.8 \times 10^{-05}$ |
| Robinow | Cleft palate [HP:0000175] | OMIM:268310 | 0/17 (0%) | OMIM:616331 | 5/8 (62%) | $1.0 \times 10^{-03}$ | 0.028 |
| Robinow | Hearing impairment [HP:0000365] | OMIM:268310 | 3/22 (14%) | OMIM:616331 | 7/7 (100%) | $7.7 \times 10^{-05}$ | 0.003 |
| Robinow | Mesomelia [HP:0003027] | OMIM:268310 | 31/31 (100%) | OMIM:616331 | 10/15 (67%) | 0.002 | 0.043 |
| Robinow | Short stature [HP:0004322] | OMIM:268310 | 29/29 (100%) | OMIM:616331 | 3/11 (27%) | $2.2 \times 10^{-06}$ | $1.7 \times 10^{-04}$ |

**Table S8: Fischer exact test for association between disease diagnosis and phenotypic features.** `OMIM:606693`: Kufor-Rakeb syndrome (KRS); `OMIM:617225`: Spastic paraplegia 78, autosomal recessive (SPG78); `OMIM:147920`: Kabuki syndrome 1 (KABUK1); `OMIM:300867`: Kabuki syndrome 2 (KABUK2); `OMIM:609192`: Loeys-Dietz syndrome 1 (LDS1); `OMIM:613795`: Loeys-Dietz syndrome 3 (LDS3); `OMIM:619656`: Loeys-Dietz syndrome 6 (LDS6); `OMIM:613826`: Leber congenital amaurosis 6 (LCA6); `OMIM:608194`: Cone-rod dystrophy 13 (CORD13); `OMIM:268310`: Robinow syndrome, autosomal recessive 1 (RRS1); `OMIM:616331`: Robinow syndrome, autosomal dominant 2 (DRS2).

| cohort | HPO | genotype (A) | Counts (A) | genotype (B) | Counts (B) | p-val | adj. p |
|---|---|---|---|---|---|---|---|
| KDM6A | Intellectual disability, severe [HP:0010864] | FEMALE | 7/25 (28%) | MALE | 14/18 (78%) | 0.002 | 0.008 |

**Table S9: Fischer exact test for association between between phenotypic features and sex (male, female)**. Adj. p: p value adjusted with Benjamini-Hochberg method.

**Table S10:** Detailed results for Table 1 of the main manuscript. **Sig**: number of significant associations. **Categorical**: Association of genotypes with phenotypes (HPO terms) by Fisher exact test. **t test**: Test of means of continuous values by student t test. **HPO**: Logrank test for association of genotypes with age of onset of a phenotypic abnormality represented by an HPO term. **Disease**: Logrank test for association of genotypes with age of onset of a disease. **Mortality**: Logrank test for association of genotypes with age of death. **Score**: Mann Whitney U test for association of genotypes with magnitude of a phenotype severity score. Cohorts with no found significant associations are not reported in the table.

| Cohort | Sig | Categorical | t test | HPO | Disease | Mortality | Score |
|---|---|---|---|---|---|---|---|
| ACADM | 2 | - | 2 | - | - | - | - |
| AIRE | 1 | - | - | 1 | - | - | - |
| ANKRD11 | 2 | - | - | - | - | - | 2 |
| ATP13A2 | 2 | 0 | - | - | - | - | - |
| BRD4 | 1 | 1 | - | - | - | - | - |
| CHD8 | 2 | - | - | - | - | - | 2 |
| CLDN16 | 1 | - | - | 1 | - | - | - |
| CNTNAP2 | 1 | - | - | - | 1 | - | - |
| CTCF | 1 | - | - | - | - | - | 1 |
| CYP21A2 | 1 | - | 1 | - | - | - | - |
| EHMT1 | 1 | 1 | - | - | - | - | - |
| FBN1 | 14 | 14 | - | - | - | - | - |
| FBXL4 | 2 | 1 | - | - | 1 | - | - |
| FGD1 | 1 | 1 | - | - | - | - | - |
| GLI3 | 14 | 14 | - | - | - | - | - |
| HMGCS2 | 1 | - | - | - | 1 | - | - |
| IKZF1 | 2 | 2 | - | - | - | - | - |
| ITPR1 | 21 | 21 | - | - | - | - | - |
| Kabuki | 2 | 2 | - | - | - | - | - |
| KCNH5 | 1 | 1 | - | - | - | - | - |
| KDM6A | 2 | 2 | - | - | - | - | - |
| LDS 1 and 3 | 4 | 4 | - | - | - | - | - |
| LDS 3 and 6 | 1 | 1 | - | - | - | - | - |
| LMNA | 24 | 23 | - | - | - | - | 1 |
| MPV17 | 3 | 1 | - | - | 1 | 1 | - |
| NBAS | 1 | 1 | - | - | - | - | - |
| NF1 | 21 | 21 | - | - | - | - | - |
| PTPN11 | 4 | 4 | - | - | - | - | - |
| RERE | 1 | - | - | - | - | - | 1 |
| Robinow syndrome | 4 | 4 | - | - | - | - | - |
| RPGRIP1 | 3 | 3 | - | - | - | - | - |
| SAMD9L | 3 | 3 | - | - | - | - | - |
| SCN2A | 9 | 9 | - | - | - | - | - |
| SCO2 | 1 | 1 | - | - | - | - | - |
| SETD2 | 9 | 8 | - | - | 1 | - | - |

Continued on next page

**Table S10 – continued from previous page**

| Cohort | Sig | Categorical | t test | HPO | Disease | Mortality | Score |
|---|---|---|---|---|---|---|---|
| SMAD3 | 1 | 1 | - | - | - | - | - |
| SMARCB1 | 6 | 6 | - | - | - | - | - |
| SMARCC2 | 1 | 1 | - | - | - | - | - |
| SPTAN1 | 35 | 35 | - | - | - | - | - |
| SUOX | 2 | 1 | - | - | 1 | - | - |
| TBCK | 2 | 2 | - | - | - | - | - |
| TBX1 | 2 | 2 | - | - | - | - | - |
| TBX5 | 3 | 3 | - | - | - | - | - |
| TGFBR1 | 4 | 4 | - | - | - | - | - |
| UMOD | 3 | 2 | - | 1 | - | - | - |
| WWOX | 1 | 1 | - | - | - | - | - |
| ZFX | 1 | 1 | - | - | - | - | - |
| ZMYM3 | 1 | 1 | - | - | - | - | - |

| HPO | id | Count | Observed (%) | Expected (%) |
|---|---|---|---|---|
| Abnormality of the musculoskeletal system | HP:0033127 | 38 | 22.8% | 17.9% |
| Abnormality of limbs | HP:0040064 | 19 | 11.4% | 11.4% |
| Abnormality of the nervous system | HP:0000707 | 37 | 22.2% | 10.8% |
| Abnormality of metabolism/homeostasis | HP:0001939 | 2 | 1.2% | 9.5% |
| Abnormality of the genitourinary system | HP:0000119 | 0 | 0.0% | 6.5% |
| Abnormality of the cardiovascular system | HP:0001626 | 11 | 6.6% | 5.7% |
| Abnormality of head or neck | HP:0000152 | 8 | 4.8% | 5.7% |
| Abnormality of the immune system | HP:0002715 | 5 | 3.0% | 5.2% |
| Abnormality of the eye | HP:0000478 | 6 | 3.6% | 4.6% |
| Abnormality of the integument | HP:0001574 | 8 | 4.8% | 4.2% |
| Abnormality of blood and blood-forming tissues | HP:0001871 | 6 | 3.6% | 3.5% |
| Abnormality of the digestive system | HP:0025031 | 3 | 1.8% | 3.3% |
| Neoplasm | HP:0002664 | 13 | 7.8% | 2.8% |
| Abnormality of the respiratory system | HP:0002086 | 1 | 0.6% | 2.5% |
| Abnormality of the endocrine system | HP:0000818 | 1 | 0.6% | 1.8% |
| Abnormal cellular phenotype | HP:0025354 | 1 | 0.6% | 1.2% |
| Abnormality of the ear | HP:0000598 | 2 | 1.2% | 1.2% |
| Abnormality of prenatal development or birth | HP:0001197 | 0 | 0.0% | 0.9% |
| Constitutional symptom | HP:0025142 | 0 | 0.0% | 0.5% |
| Growth abnormality | HP:0001507 | 6 | 3.6% | 0.4% |
| Abnormality of the breast | HP:0000769 | 0 | 0.0% | 0.2% |
| Abnormality of the voice | HP:0001608 | 0 | 0.0% | 0.1% |
| Abnormality of the thoracic cavity | HP:0045027 | 0 | 0.0% | 0.0% |

**Table S11:** Distribution of significant Fisher exact test results according to top-level HPO term

### ABCB7

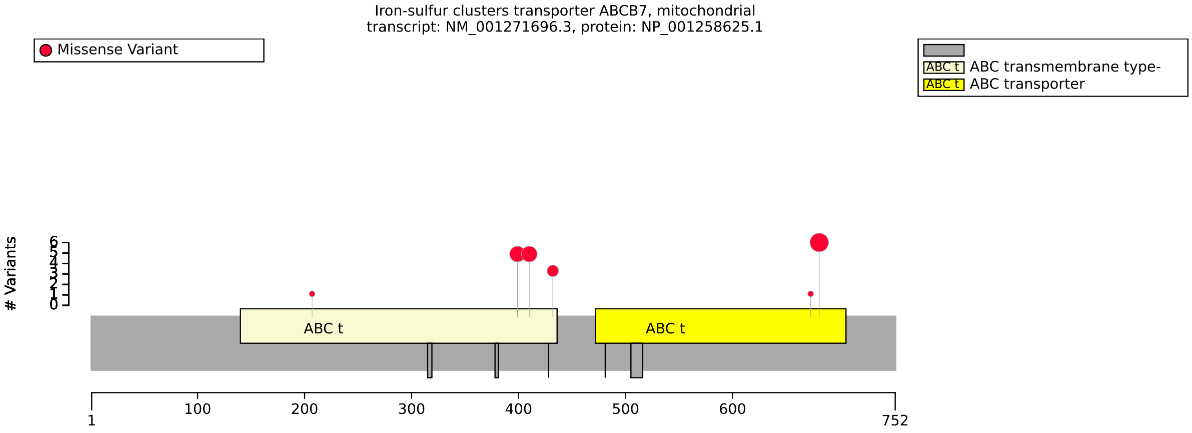


**(a)** Distribution of variants in ABCB7

| Genotype (A) | Genotype (B) | total tests performed | significant results |
|---|---|---|---|
| ABC transmembrane type-1 | Other region | 12 | 0 |
| p.Gly682Ser | Other variant | 10 | 0 |

**(b)** Fisher Exact Test performed to compare HPO annotation frequency with respect to variants located in the ABC transmembrane type-1 region and p.Gly682Ser.

**Figure S1:** The cohort comprised 18 individuals (0 females, 18 males). A total of 52 HPO terms were used to annotate the cohort. Disease diagnosis: Anemia, sideroblastic, and spinocerebellar ataxia (OMIM:301310). No statistically significant results identified. A total of 18 unique variant alleles were found in *ABCB7* (transcript: `NM_001271696.3`, protein id: `NP_001258625.1`).

### ACADM

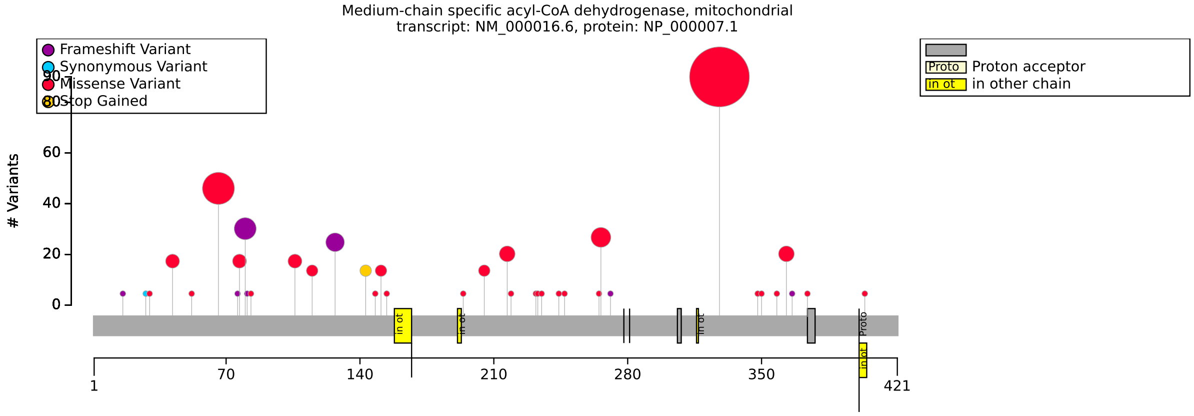


**(a)** Distribution of variants in ACADM

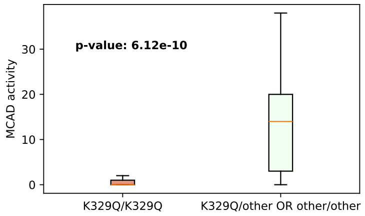


**(b)** Lys329Glu: t test for MCAD Activity (% normal; LOINC:74892-1): p=$6.12 \times 10^{-10}$.

| Description | Variable | Genotype (A) | Genotype (B) | p-value | ref |
|---|---|---|---|---|---|
| Value of MCAD Activity% [LOINC:74892-1] | LOINC:74892-1 | K329Q/K329Q | K329Q/other OR other/other | $6.12 \times 10^{-10}$ | [3] |

**(c)** t-test to compare K329Q/K329Q and K329Q/other OR other/other with respect to LOINC:74892-1. Mean MCAD activity for K329/K329: 0.52%, and for K329/other or other/other: 13.23%

| Description | Variable | Genotype (A) | Genotype (B) | p-value | xrefs |
|---|---|---|---|---|---|
| Value of MCAD Activity% [LOINC:74892-1] | LOINC:74892-1 | Y67H/Y67H OR Y67H/other | other/other | $2.01 \times 10^{-5}$ | [3] |

**(d)** t-test to compare Y67H/Y67H OR Y67H/other and other/other with respect to LOINC:74892-1. Mean MCAD activity for Y67H/Y67H: 18.60%, and for Y67H/other or other/other: 7.68%

**Figure S2:** The cohort comprised 115 individuals (0 females, 0 males, 115 with unknown sex). The cohort had data about medium chain Acyl-CoA dehydrogenase (MCAD), expressed as percentage of normal. The variant c.985G>A (p.Lys329Glu) is known to be severe, and the variant c.199T>C (p.Tyr67His) is known to be mild [3]. Disease diagnosis: Acyl-CoA dehydrogenase, medium chain, deficiency of (OMIM:201450). No statistically significant results identified. A total of 47 unique variant alleles were found in *ACADM* (transcript: `NM_000016.6`, protein id: `NP_000007.1`).

### ACBD6

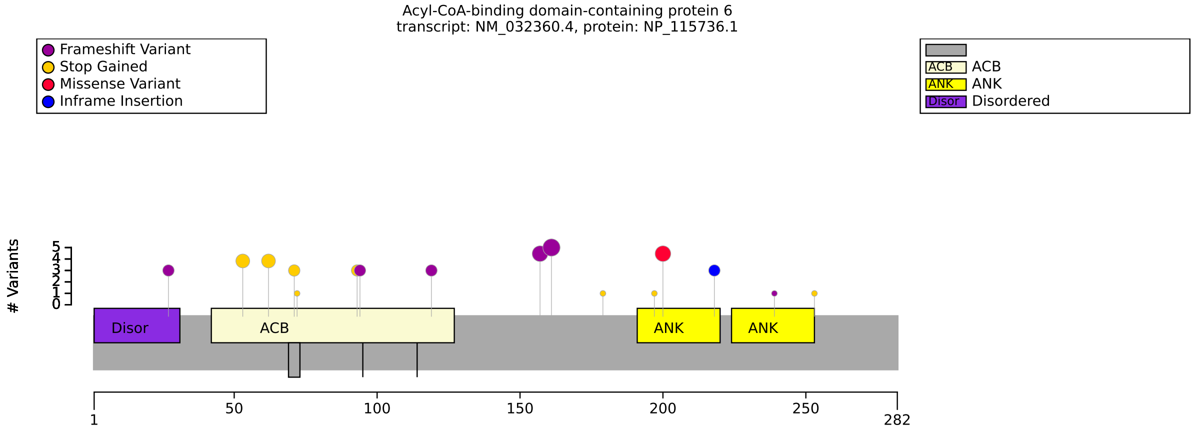


**(a)** Distribution of variants in ACBD6

| Genotype (A) | Genotype (B) | total tests performed | significant results |
|---|---|---|---|
| missense/missense OR missense/other | other/other | 99 | 0 |

**(b)** Fisher Exact Test performed to compare HPO annotation frequency with respect to missense/missense OR missense/other and other/other.

| Genotype (A) | Genotype (B) | total tests performed | significant results |
|---|---|---|---|
| ACB/ACB OR ACB/other | other/other | 99 | 0 |

**(c)** Fisher Exact Test performed to compare HPO annotation frequency with respect to ACB/ACB OR ACB/other and other/other.

| Genotype (A) | Genotype (B) | total tests performed | significant results |
|---|---|---|---|
| FEMALE | MALE | 99 | 0 |

**(d)** Fisher Exact Test performed to compare HPO annotation frequency with respect to FEMALE and MALE.

**Figure S3:** The cohort comprised 45 individuals (22 females, 23 males). A total of 79 HPO terms were used to annotate the cohort. Disease diagnosis: Neurodevelopmental disorder with progressive movement abnormalities (OMIM:620785). No statistically significant results identified. A total of 20 unique variant alleles were found in *ACBD6* (transcript: NM_032360.4, protein id: NP_115736.1).

### AIRE

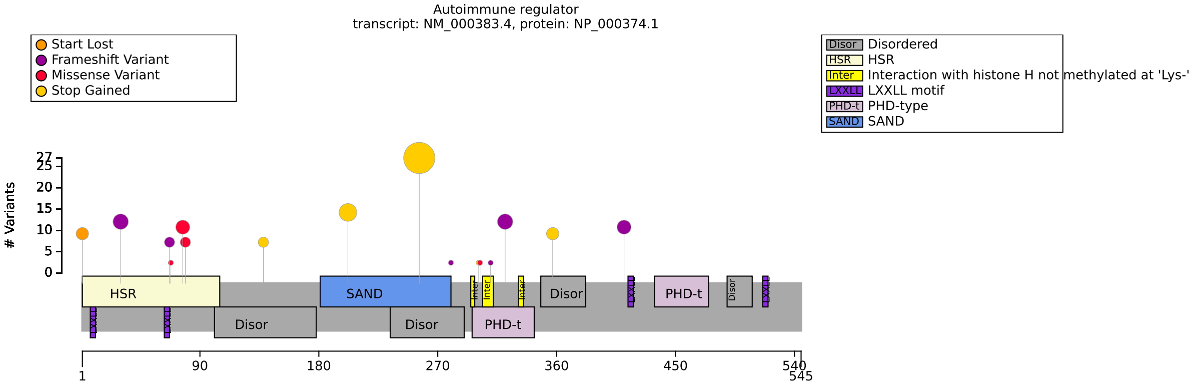


**(a)** Distribution of variants in AIRE

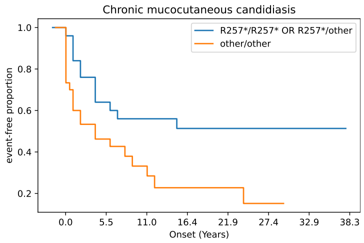


**(b)** log-rank p-value for Chronic mucocutaneous candidiasis (HP:0002728) and R357*/R357* or R357*/other versus other/other 0.0192

| Genotype (A) | Genotype (B) | total tests performed | significant results |
|---|---|---|---|
| R257*/R257* OR R257*/other | other/other | 18 | 0 |

**(c)** Fisher Exact Test performed to compare HPO annotation frequency with respect to R257*/R257* OR R257*/other and other/other.

| Description | Variable | Genotype (A) | Genotype (B) | p-value | xrefs |
|---|---|---|---|---|---|
| Survival analysis: Chronic mucocutaneous candidiasis | Onset of HP:0002728 | R257*/R257* OR R257*/other | other/other | 0.019 | [4] |

**(d)** Onset of Chronic mucocutaneous candidiasis (HP:0002728) to compare R257*/R257* OR R257*/other and other/other with respect to Onset of HP:0002728.

**Figure S4:** The cohort comprised 58 individuals (31 females, 27 males). 2 of these individuals were reported to be deceased. A total of 47 HPO terms were used to annotate the cohort. Disease diagnosis: Autoimmune polyendocrinopathy syndrome , type I, with or without reversible metaphyseal dysplasia (OMIM:240300). A higher prevalence of chronic mucocutanous candidiasis with the variant Arg357Ter than with other variants was reported previously [4]. We did not identify a significant difference in prevalence A total of 18 unique variant alleles were found in *AIRE* (transcript: `NM_000383.4`, protein id: `NP_000374.1`).

### ANKRD11

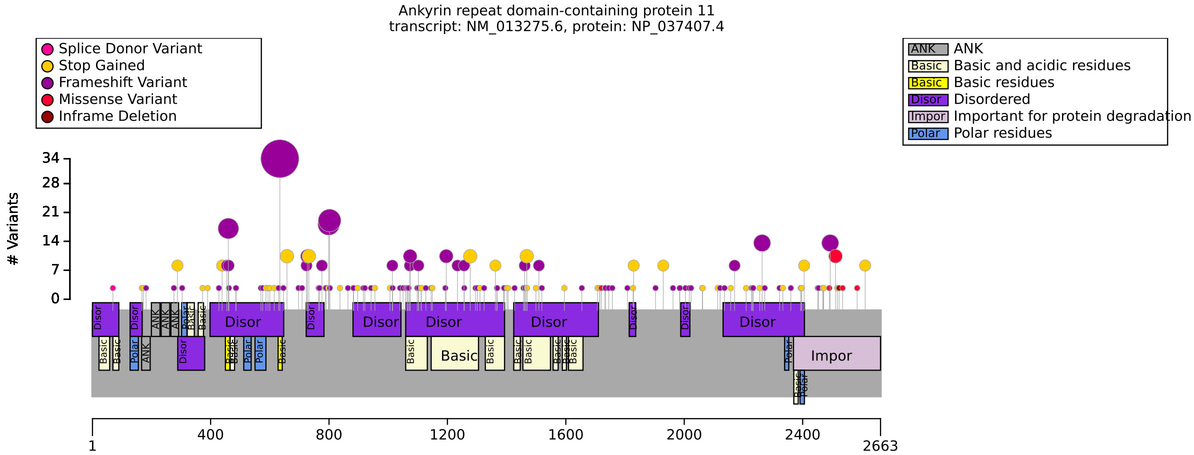


**(a)** Distribution of variants in ANKRD11

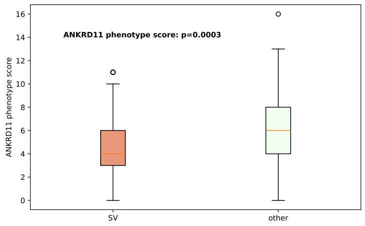


**(b)** ANKRD11 phenotypical score. Mean for structural variants: 4.62, and for other variants: 5.94

| Genotype (A) | Genotype (B) | total tests performed | significant results |
|---|---|---|---|
| SV | other | 23 | 0 |
| exon 9 | other | 23 | 0 |
| c.1903_1907del | other | 23 | 0 |

**(c)** Fisher Exact Test performed to compare HPO annotation frequency with respect to SV, exon 9, and c.1903_1907del.

| Description | Variable | Genotype (A) | Genotype (B) | p-value | xrefs |
|---|---|---|---|---|---|
| ANKRD11 phenotype score | HPO group count | SV | other | $2.64 \times 10^{-4}$ | [1] |
| ANKRD11 phenotype score | HPO group count | FEMALE | MALE | 0.0075 | - |

**(d)** AKKRD11 phenotype score for structural variants: 4.62; other variants: 5.94. male/female comparison: Female 5.28, Male: 6.15.

**Figure S5:** The cohort comprised 337 individuals (143 females, 175 males, 19 with unknown sex). A total of 48 HPO terms were used to annotate the cohort. Disease diagnosis: KBG syndrome (OMIM:148050). Validated results on ANKRD11 phenotypical score. Other results did not survive multiple testing correction. Awamleh et al. (2023) stated that no significant DNAm differences based on sex at the identified KBGS-specific signature sites [5]. We are aware of no other publication that mentions sex-specific differences in KBGS. A total of 163 unique variant alleles were found in *ANKRD11* (transcript: `NM_013275.6`, protein id: `NP_037407.4`).

### ASPM

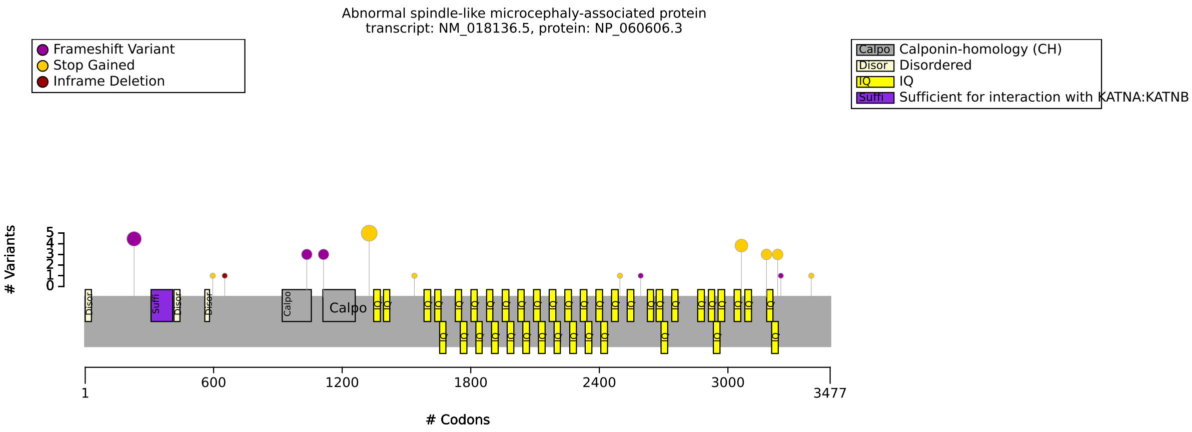


(**a**) Distribution of variants in ASPM

| Genotype (A) | Genotype (B) | total tests performed | significant results |
|---|---|---|---|
| N Term/N Term OR N Term/other | other/other | 14 | 0 |
| FEMALE | MALE | 14 | 0 |

(**b**) Fisher Exact Test performed to compare HPO annotation frequency with respect to N Term/N Term OR N Term/other and other/other, as well as male/female comparison.

**Figure S6:** The cohort comprised 22 individuals (14 females, 8 males). A total of 17 HPO terms were used to annotate the cohort. Disease diagnosis: Microcephaly 5, primary, autosomal recessive (OMIM:608716). No statistically significant results identified. A total of 15 unique variant alleles were found in *ASPM* (transcript: `NM_018136.5`, protein id: `NP_060606.3`).

### ATP6V0C

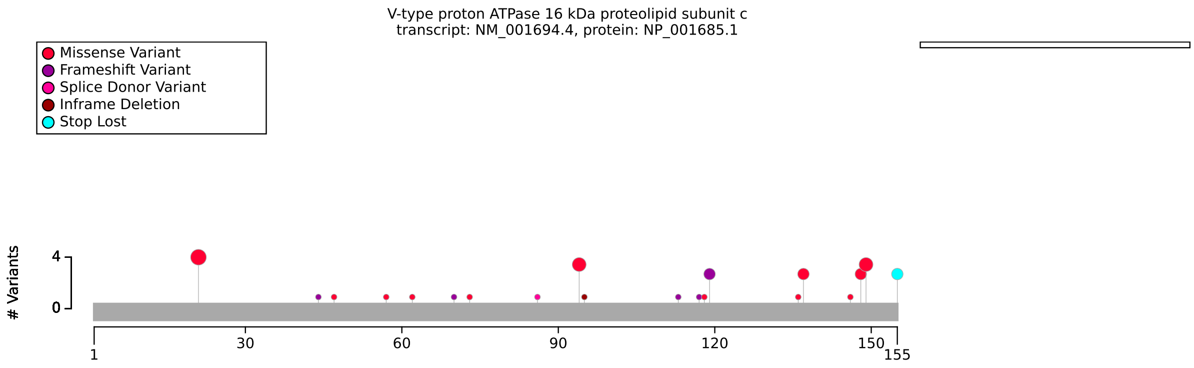


**(a)** Distribution of variants in ATP6V0C

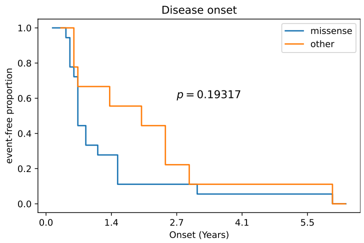


**(b)** Disease onset for ATP6V0C missense vs. other variants.

| Genotype (A) | Genotype (B) | total tests performed | significant results |
|---|---|---|---|
| 1-100 | 100+ | 80 | 0 |
| missense | other | 80 | 0 |
| FEMALE | MALE | 80 | 0 |

**(c)** Fisher Exact Test performed to compare HPO annotation frequency with respect to genotypes.

| Description | Variable | Genotype (A) | Genotype (B) | p-value | xrefs |
|---|---|---|---|---|---|
| Compute time until OMIM:620465 onset | Onset of OMIM:620465 | missense | other | 0.193 | - |

**(d)** Onset of OMIM:620465 to compare missense and other with respect to Onset of OMIM:620465.

**Figure S7:** The cohort comprised 31 individuals (12 females, 19 males). A total of 83 HPO terms were used to annotate the cohort. Disease diagnosis: Epilepsy, early-onset, 3, with or without developmental delay (OMIM:620465). to do. A total of 23 unique variant alleles were found in *ATP6V0C* (transcript: `NM_001694.4`, protein id: `NP_001685.1`).

### ATP13A2

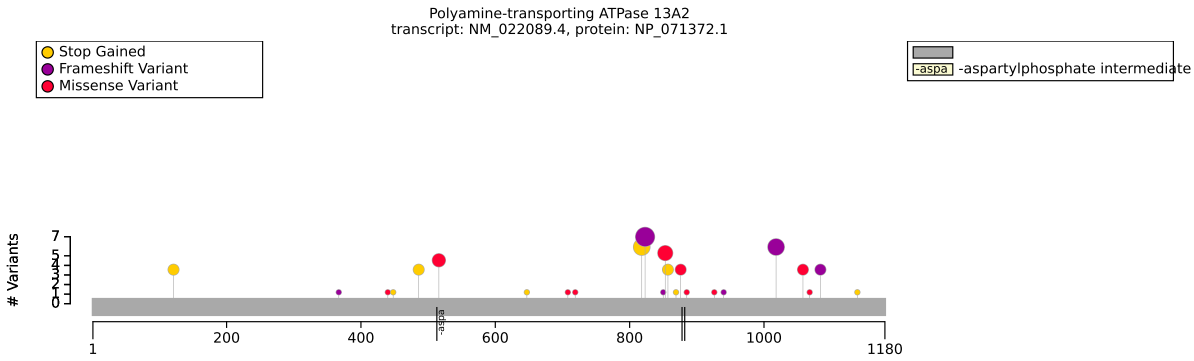


**(a)** Distribution of variants in ATP13A2

| HPO term | OMIM:606693 | OMIM:617225 | p-value | adj. p-value |
|---|---|---|---|---|
| Parkinsonism [HP:0001300] | 28/28 (100%) | 3/11 (27%) | $2.68 \times 10^{-6}$ | $7.78 \times 10^{-5}$ |
| Bradykinesia [HP:0002067] | 30/32 (94%) | 4/10 (40%) | $9.15 \times 10^{-4}$ | 0.013 |

**(b)** Fisher Exact Test performed to compare HPO annotation frequency with respect to Kufor-Rakeb syndrome (OMIM:606693) and Spastic paraplegia 78, autosomal recessive (OMIM:617225). Total of 29 tests were performed.

| Genotype (A) | Genotype (B) | total tests performed | significant results |
|---|---|---|---|
| missense/missense OR missense/other | other/other | 30 | 0 |

**(c)** Fisher Exact Test performed to compare HPO annotation frequency with respect to genotypes.

**Figure S8:** The cohort comprised 45 individuals (18 females, 27 males). A total of 71 HPO terms were used to annotate the cohort. Disease diagnoses: Kufor-Rakeb syndrome (OMIM:606693) (34 individuals), Spastic paraplegia 78, autosomal recessive (OMIM:617225) (11 individuals). As expected, the frequency of Parkinsonian manifestations is higher in Kufor-Rakeb syndrome, a rare autosomal recessive form of juvenile-onset atypical Parkinson disease. A total of 24 unique variant alleles were found in *ATP13A2* (transcript: `NM_022089.4`, protein id: `NP_071372.1`).

### BRD4

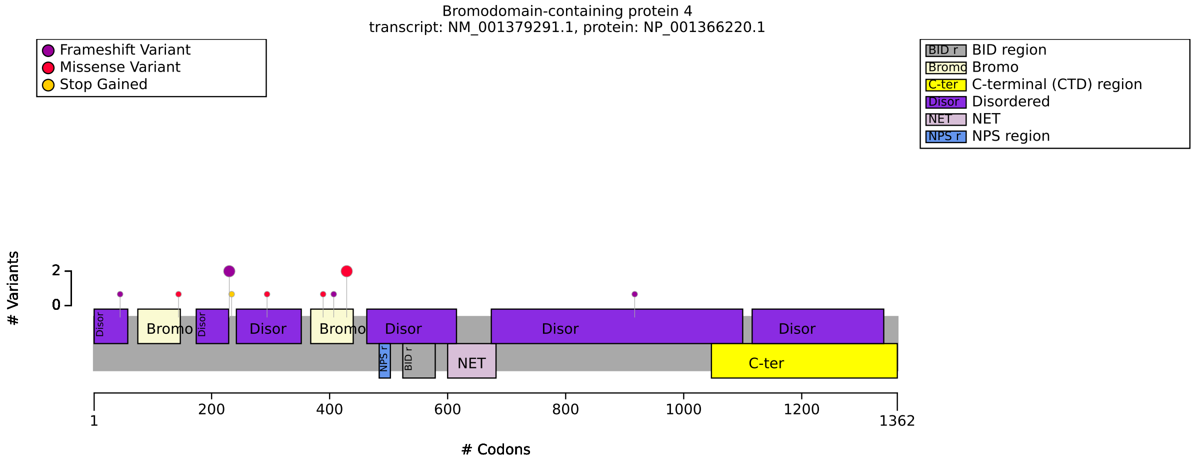


**(a)** Distribution of variants in BRD4

| HPO term | NIPBL | BRD4 | p-value | adj. p-value |
|---|---|---|---|---|
| Intrauterine growth retardation [HP:0001511] | 37/45 (82%) | 2/9 (22%) | $9.45 \times 10^{-4}$ | 0.032 |

**(b)** Fisher Exact Test performed to compare HPO annotation frequency with respect to NIPBL and BRD4. Total of 34 tests were performed.

| Genotype (A) | Genotype (B) | total tests performed | significant results |
|---|---|---|---|
| ablation | other | 27 | 0 |

**(c)** Fisher Exact Test performed to compare HPO annotation frequency with respect to genotypes.

**Figure S9:** The cohort comprised 18 individuals (8 females, 10 males). A total of 40 HPO terms were used to annotate the cohort. Disease diagnosis: Cornelia de Lange syndrome 6 (OMIM:620568). We compared phenotypic features of Cornelia de Lange syndrome 6 (BRD4) with those of Cornelia de Lange syndrome 1 (NIPBL). Intrauterine growth retardation was found to be significantly more common in Cornelia de Lange syndrome 1. A total of 10 unique variant alleles were found in *BRD4* (transcript: NM_001379291.1, protein id: NP_001366220.1).

### CHD8

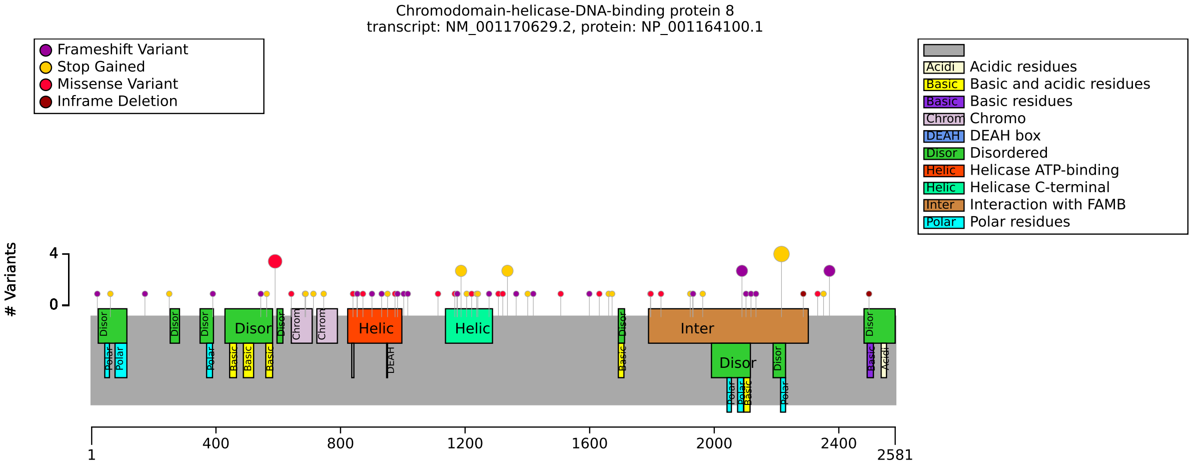


(**a**) Distribution of variants in CHD8

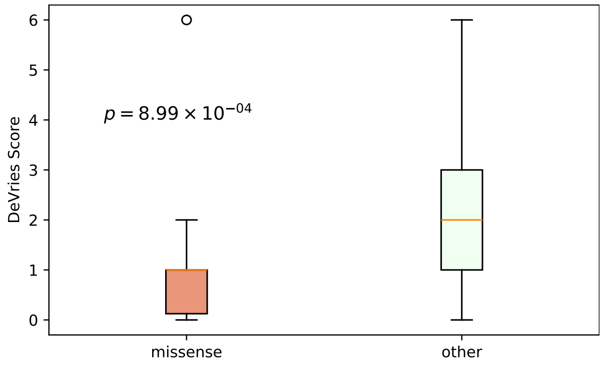


(**b**) De Vries Score to compare Missense and other variants.

| Genotype (A) | Genotype (B) | total tests performed | significant results |
|---|---|---|---|
| Missense | Not Missense | 36 | 0 |
| FEMALE | MALE | 36 | 0 |

(**c**) Fisher Exact Test performed to compare HPO annotation frequency with respect to genotypes.

| Description | Variable | Genotype (A) | Genotype (B) | p-value | xrefs |
|---|---|---|---|---|---|
| De Vries score | De Vries score | Missense | Not Missense | $8.99 \times 10^{-4}$ | [6] |
| De Vries score | De Vries score | FEMALE | MALE | 0.006 | - |

**(d)** De Vries Score to compare Missense and Other variants and for M/F sex differences.

**Figure S10:** The cohort comprised 79 individuals (26 females, 53 males). 1 of these individuals was reported to be deceased. A total of 81 HPO terms were used to annotate the cohort. Disease diagnosis: Intellectual developmental disorder with autism and macrocephaly (OMIM:615032). Dingemans et al. (2022) identified a correlation between the severity of the phenotypes (as measured by a phenotype severity score termed a DeVries test) and missense variants on the CHD8 gene, specifically that those with a missense variant were significantly less affected than other individuals [6]. A total of 70 unique variant alleles were found in *CHD8* (transcript: `NM_001170629.2`, protein id: `NP_001164100.1`).

### CLDN16

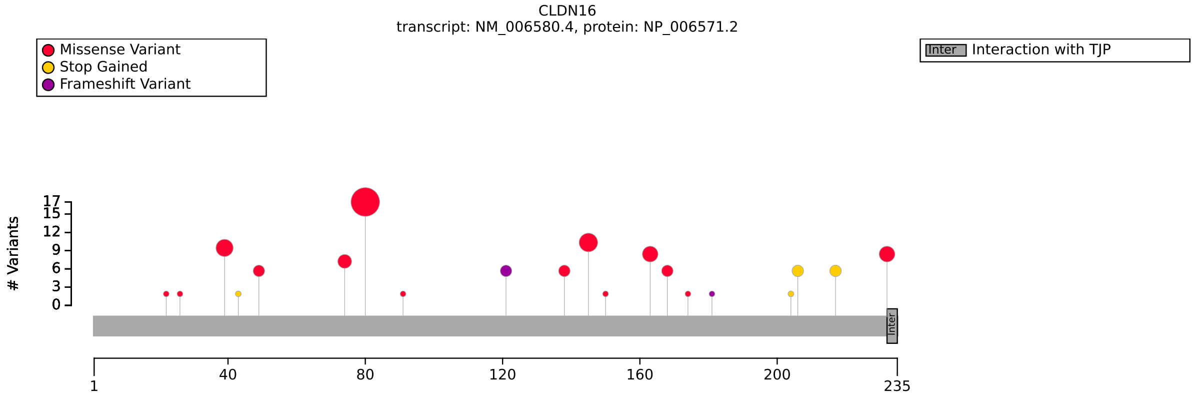


**(a)** Distribution of variants in CLDN16

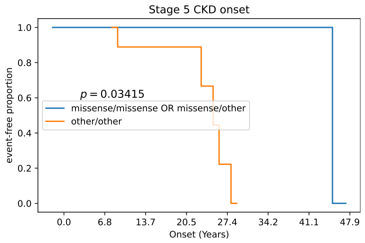


**(b)** Onset of stage 5 chronic kidney disease for CLDN16 missense vs. other variants. This result is similar to that of Konrad et al. (2008) [7].

| Genotype (A) | Genotype (B) | total tests performed | significant results |
|---|---|---|---|
| Leu81Phe/Leu81Phe | other/other OR Leu81Phe/other | 26 | 0 |
| missense/missense OR missense/other | other/other | 23 | 0 |
| FEMALE | MALE | 27 | 0 |

**(c)** Fisher Exact Test performed to compare HPO annotation frequency with respect to genotypes.

| Description | Variable | Genotype (A) | Genotype (B) | p-value | xrefs |
|---|---|---|---|---|---|
| Hypomagnesemia 3, renal (OMIM:248250) disease onset | Onset of OMIM:248250 | missense/missense OR missense/other | other/other | 0.333 | [7] |

**(d)** Onset of OMIM:248250 to compare missense/missense OR missense/other and other/other with respect to Onset of OMIM:248250.

| Description | Variable | Genotype (A) | Genotype (B) | p-value | xrefs |
|---|---|---|---|---|---|
| Survival analysis: Stage 5 chronic kidney disease | Onset of HP:0003774 | missense/missense OR missense/other | other/other | 0.034 | [7] |

**(e)** Onset of Stage 5 chronic kidney disease to compare missense/missense OR missense/other and other/other with respect to Onset of HP:0003774.

**Figure S11:** The cohort comprised 51 individuals (21 females, 29 males, 1 with unknown sex). A total of 53 HPO terms were used to annotate the cohort. Disease diagnosis: Hypomagnesemia 3, renal (OMIM:248250). to do. A total of 23 unique variant alleles were found in *CLDN16* (transcript: `NM_006580.4`, protein id: `NP_006571.2`).

### CNTNAP2

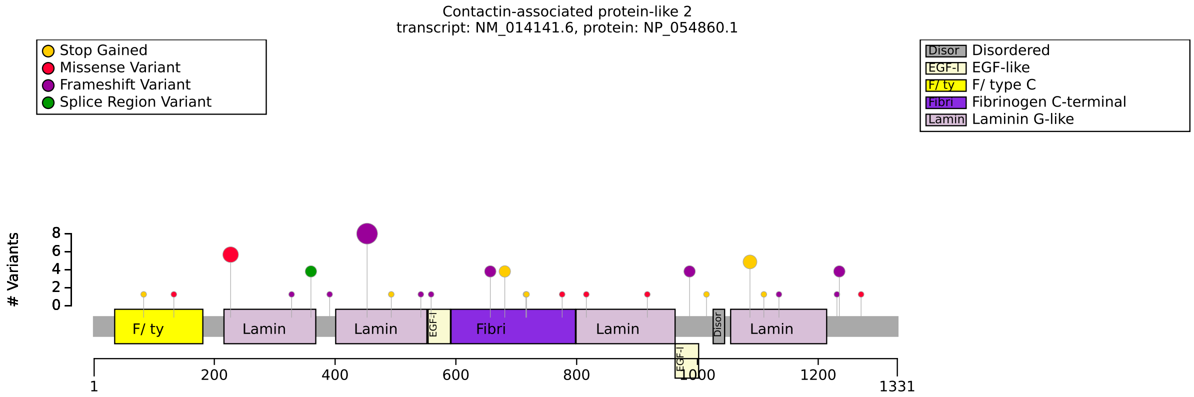


**(a)** Distribution of variants in CNTNAP2

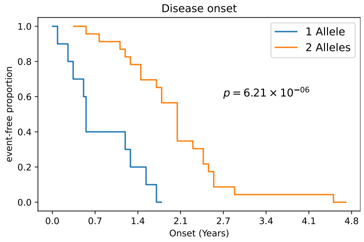


**(b)** Disease onset: Count of pathogenic CNTNAP2 alleles

| Genotype (A) | Genotype (B) | total tests performed | significant results |
|---|---|---|---|
| Ablation/Ablation | other/other OR Ablation/other | 46 | 0 |
| FEMALE | MALE | 48 | 0 |
| 1 allele | 2 alleles | 43 | 0 |

**(c)** Fisher Exact Test performed to compare HPO annotation frequency with respect to genotypes.

| Description | Variable | Genotype (A) | Genotype (B) | p-value | xrefs |
|---|---|---|---|---|---|
| Pitt-Hopkins like syndrome 1 (OMIM:610042) disease onset | Onset of OMIM:610042 | 1 allele | 2 alleles | $6.21 \times 10^{-6}$ | - |

**(d)** Onset of OMIM:610042 to compare 1 allele and 2 alleles with respect to Onset of OMIM:610042.

**Figure S12:** The cohort comprised 63 individuals (29 females, 32 males, 2 with unknown sex). 2 of these individuals were reported to be deceased. A total of 72 HPO terms were used to annotate the cohort. Disease diagnosis: Pitt-Hopkins like syndrome 1 (OMIM:610042). D'Onofrio et al. (2023) reported several GPCs, but the authors did not apply multiple-testing correction [8]. These authors did not perform survival analysis for age of onset of disease. A total of 30 unique variant alleles were found in *CNTNAP2* (transcript: `NM_014141.6`, protein id: `NP_054860.1`).

### COQ4

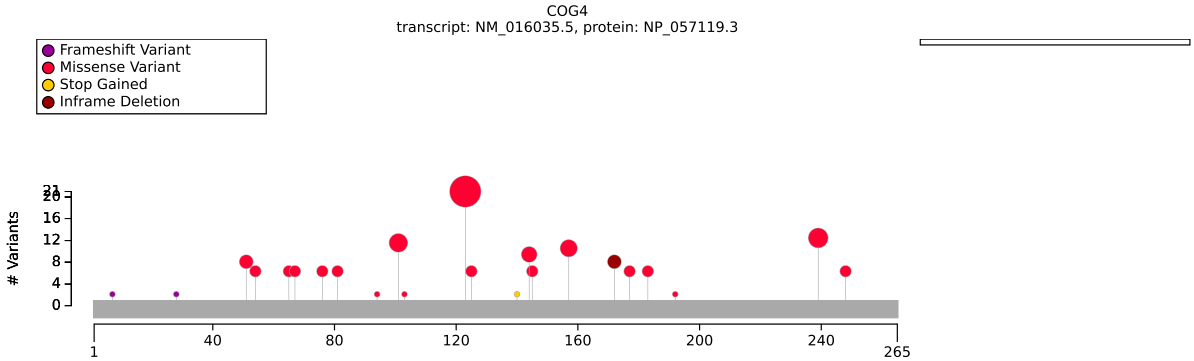


**(a)** Distribution of variants in COQ4

| Genotype (A) | Genotype (B) | total tests performed | significant results |
|---|---|---|---|
| missense/other OR missense/missense | other/other | 4 | 0 |
| G124S/other OR G124S/G124S | other/other | 13 | 0 |
| G124S/other OR G124S/G124S | other/other | 13 | 0 |
| Exon 1-4/other OR Exon 1-4/Exon 1-4 | other/other | 13 | 0 |

**(b)** Fisher Exact Test performed to compare HPO annotation frequency with respect to genotypes.

**Figure S13:** The cohort comprised 51 individuals (30 females, 21 males). 19 of these individuals were reported to be deceased. A total of 91 HPO terms were used to annotate the cohort. A previous work found that pathogenic COQ4 variants in exons 1-4 are associated with less life-threating presentations, late onset, responsiveness to CoQ10 therapy, and a relatively long lifespan, but correction for multiple testing was not performed [9]. Disease diagnoses: Coenzyme Q10 deficiency, primary, 7 (OMIM:616276) (35 individuals), Spastic ataxia 10, autosomal recessive (OMIM:620666) (16 individuals). to do. A total of 32 unique variant alleles were found in *COQ4* (transcript: `NM_016035.5`, protein id: `NP_057119.3`).

### COL3A1

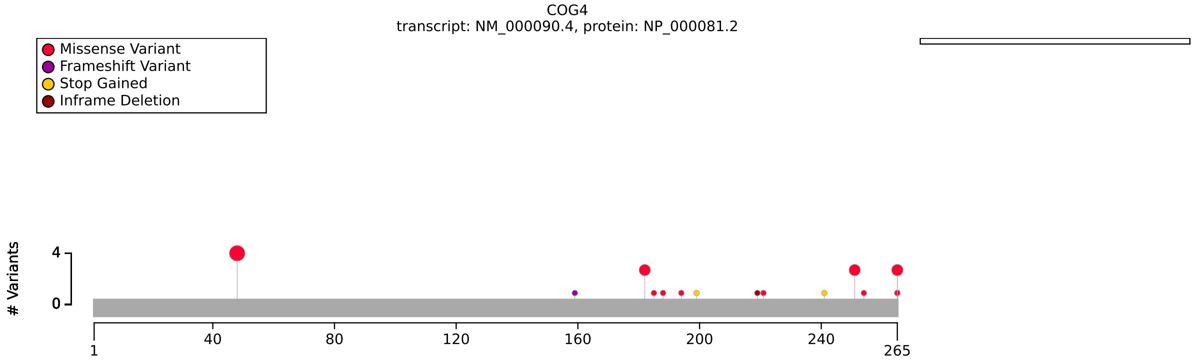


**(a)** Distribution of variants in COL3A1

| Genotype (A) | Genotype (B) | total tests performed | significant results |
|---|---|---|---|
| Missense | Other | 20 | 0 |
| Missense | Other | 20 | 0 |
| triple helix region | Other | 20 | 0 |
| OMIM:130050 | OMIM:618343 | 15 | 0 |
| Gly missense | Other | 20 | 0 |

**(b)** Fisher Exact Test performed to compare HPO annotation frequency with respect to genotypes.

| Description | Variable | Genotype (A) | Genotype (B) | p-value | xrefs |
|---|---|---|---|---|---|
| Age of OMIM:130050 onset | Onset of OMIM:130050 | missense | other | 0.317 | - |

**(c)** Onset of OMIM:130050 to compare missense and other with respect to Onset of OMIM:130050.

**Figure S14:** The cohort comprised 41 individuals (24 females, 17 males). A total of 43 HPO terms were used to annotate the cohort. Disease diagnoses: Ehlers-Danlos syndrome, vascular type (OMIM:130050) (35 individuals), Polymicrogyria with or without vascular-type EDS (OMIM:618343) (6 individuals). No significant GPC identified. Frank et al (2015) found that glycine missense variants were associated with a higher degree of severity in a cohort of 215 individuals. Primary data was not made available. A total of 38 unique variant alleles were found in *COL3A1* (transcript: `NM_000090.4`, protein id: `NP_000081.2`).

### CTCF

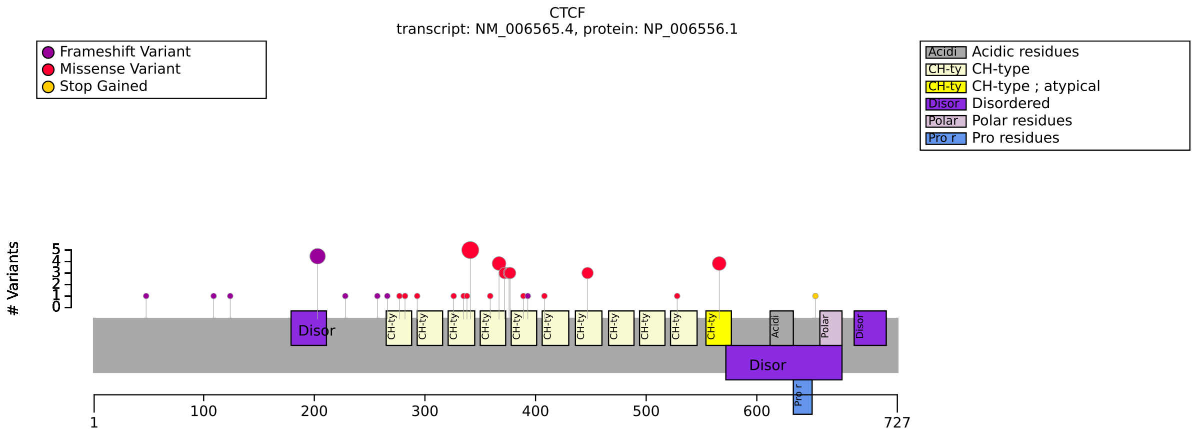


**(a)** Distribution of variants in CTCF

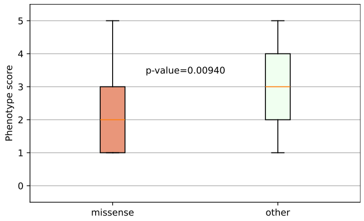


**(b)** DeVries score to compare CTCF missense vs other variants.

| Genotype (A) | Genotype (B) | total tests performed | significant results |
|---|---|---|---|
| missense | other | 27 | 0 |

**(c)** Fisher Exact Test performed to compare HPO annotation frequency with respect to genotypes.

| Description | Variable | Genotype (A) | Genotype (B) | p-value | xrefs |
|---|---|---|---|---|---|
| A phenotypic severity score for individuals with intellectual disability | De Vries score | missense | other | 0.009 | - |

**(d)** De Vries Score to compare missense and other with respect to De Vries score.

**Figure S15:** The cohort comprised 46 individuals (18 females, 28 males). A total of 110 HPO terms were used to annotate the cohort. Disease diagnosis: Intellectual developmental disorder, autosomal dominant 21 (OMIM:615502). Previously, a group did not observe a correlation between the location of the variants and the overall severity of various phenotypes, including the level of intellectual function, presence of congenital anomalies, or poor growth [10], but analysis using a score such as the DeVries score was not attempted. A total of 32 unique variant alleles were found in *CTCF* (transcript: `NM_006565.4`, protein id: `NP_006556.1`).

### CYP21A2

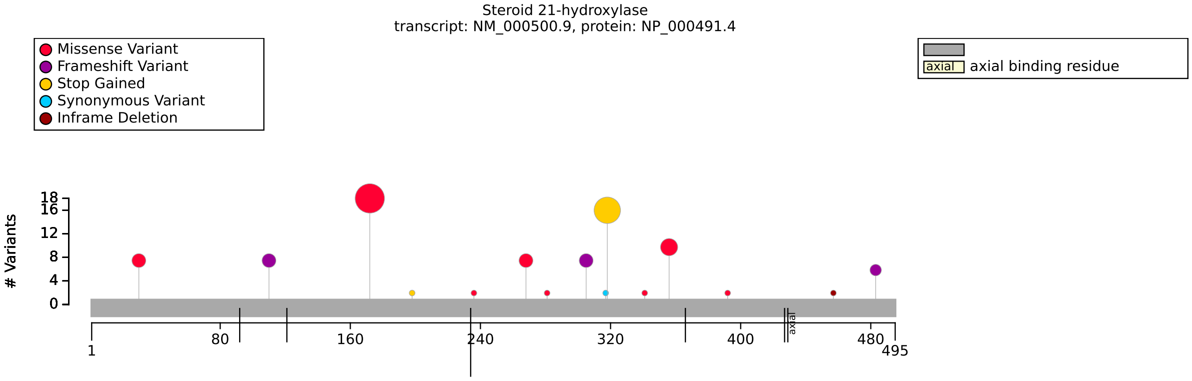


**(a)** Distribution of variants in CYP21A2

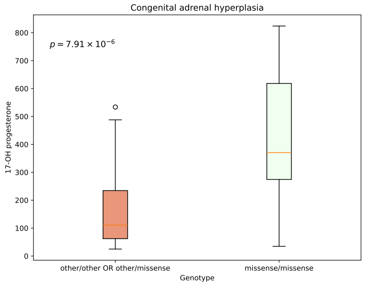


**(b)** t-test. 17-hydroxyprogesterone level. CYP21A2 missense variants are compared with other variants.

| Description | Variable | Genotype (A) | Genotype (B) | p-value | xrefs |
|---|---|---|---|---|---|
| 17-Hydroxyprogesterone [Mass/volume] in Serum or Plasma [LOINC:1668-3] | LOINC:1668-3 | other/other OR other/missense | missense/missense | $7.91 \times 10^{-6}$ | [11] |

**(c)** t-test to compare other/other OR other/missense and missense/missense with respect to LOINC:1668-3.

**Figure S16:** The cohort comprised 69 individuals (34 females, 35 males). A total of 27 HPO terms were used to annotate the cohort. Disease diagnosis: Adrenal hyperplasia, congenital, due to 21-hydroxylase deficiency (OMIM:201910). It is assumed that the mildest mutation determines the phenotype in compound heterozygotes. Several missense variants display the highest residual activities. High levels of 17-hydroxyprogesterone may be observed with 21-hydroxylase deficiency [11]. A total of 22 unique variant alleles were found in *CYP21A2* (transcript: `NM_000500.9`, protein id: `NP_000491.4`).

### EHMT1

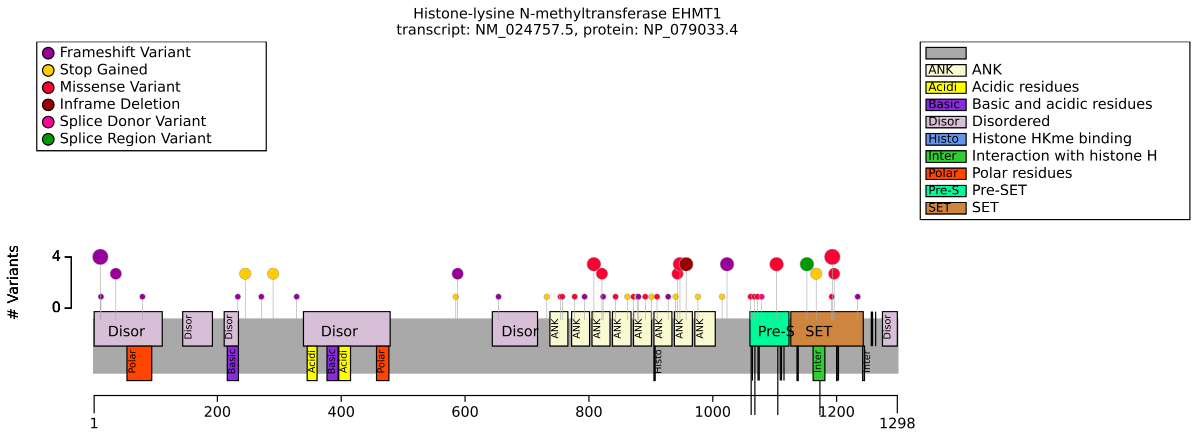


**(a)** Distribution of variants in EHMT1

| HPO term | N Term Frameshift | other | p-value | adj. p-value |
|---|---|---|---|---|
| Attention deficit hyperactivity disorder [HP:0007018] | 4/8 (50%) | 3/96 (3%) | $4.83 \times 10^{-4}$ | 0.029 |

**(b)** Fisher Exact Test performed to compare HPO annotation frequency with respect to N Term Frameshift and other. Total of 60 tests were performed.

| Genotype (A) | Genotype (B) | total tests performed | significant results |
|---|---|---|---|
| Missense | Other | 62 | 0 |
| FEMALE | MALE | 62 | 0 |
| ANKR | other | 62 | 0 |
| SET | other | 62 | 0 |

**(c)** Fisher Exact Test performed to compare HPO annotation frequency with respect to genotypes.

**Figure S17:** The cohort comprised 125 individuals (81 females, 44 males). A total of 60 HPO terms were used to annotate the cohort. Disease diagnosis: Kleefstra syndrome 1 (OMIM:610253). Rots et al. reported several correlations [12]. Multiple-testing correction was not performed. Frazier et al (2025) also reported significant correlations but the original data was not made available [13]. A total of 62 unique variant alleles were found in *EHMT1* (transcript: `NM_024757.5`, protein id: `NP_079033.4`).

### EZH1

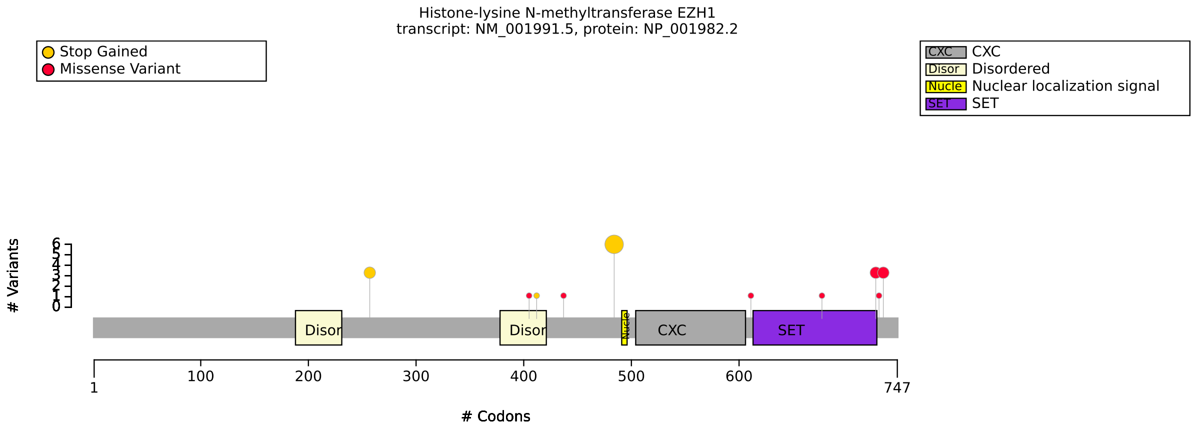


**(a)** Distribution of variants in EZH1

| Genotype (A) | Genotype (B) | total tests performed | significant results |
|---|---|---|---|
| 1 allele | 2 alleles | 33 | 0 |

**(b)** Fisher Exact Test performed to compare HPO annotation frequency with respect to monoallelic and biallelic pathogenic variants.

**Figure S18:** The cohort comprised 19 individuals (10 females, 8 males, 1 with unknown sex). A total of 106 HPO terms were used to annotate the cohort. Disease diagnosis: EZH1-related neurodevelopmental disorder (OMIM:601674). No statistically significant results identified. A total of 12 unique variant alleles were found in *EZH1* (transcript: `NM_001991.5`, protein id: `NP_001982.2`). In the original publication, it is stated that patients show neurodevelopmental delay with variable clinical presentations regardless of variant type and zygosity [14].

### FBN1

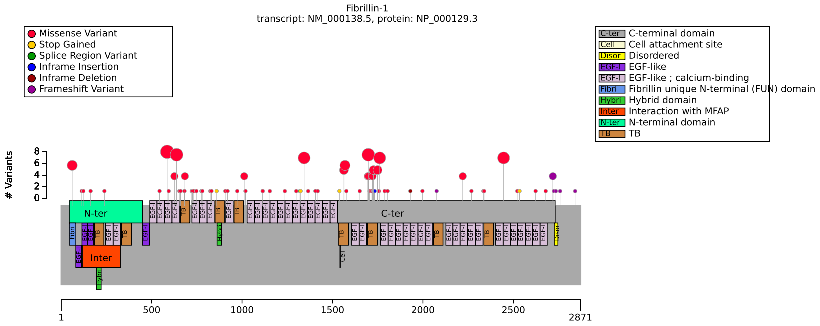


**(a)** Distribution of variants in FBN1

| HPO term | missense | other | p-value | adj. p-value |
|---|---|---|---|---|
| Arachnodactyly [HP:0001166] | 34/81 (42%) | 18/22 (82%) | 0.001 | 0.026 |
| Thoracic aortic aneurysm [HP:0012727] | 25/64 (39%) | 12/14 (86%) | 0.002 | 0.026 |
| Hyperextensibility of the finger joints [HP:0001187] | 0/78 (0%) | 3/14 (21%) | 0.003 | 0.026 |

**(b)** Fisher Exact Test: missense vs. other. Total of 27 tests were performed.

| HPO term | TB domain | cbEGF | p-value | adj. p-value |
|---|---|---|---|---|
| Ectopia lentis [HP:0001083] | 9/18 (50%) | 48/59 (81%) | 0.013 | 0.038 |
| Mitral valve prolapse [HP:0001634] | 1/28 (4%) | 13/47 (28%) | 0.012 | 0.038 |
| Disproportionate tall stature [HP:0001519] | 2/40 (5%) | 12/43 (28%) | 0.007 | 0.029 |
| Tall stature [HP:0000098] | 7/38 (18%) | 21/40 (52%) | 0.002 | 0.011 |
| Severe short stature [HP:0003510] | 15/36 (42%) | 0/24 (0%) | $1.36 \times 10^{-4}$ | $9.04 \times 10^{-4}$ |
| Proportionate short stature [HP:0003508] | 20/36 (56%) | 0/24 (0%) | $2.35 \times 10^{-6}$ | $2.35 \times 10^{-5}$ |
| Short stature [HP:0004322] | 23/39 (59%) | 0/24 (0%) | $4.39 \times 10^{-7}$ | $8.79 \times 10^{-6}$ |

**(c)** Fisher Exact Test: TB domain vs cbEGF. Total of 20 tests were performed.

| HPO term | exon 37 | other | p-value | adj. p-value |
|---|---|---|---|---|
| Arachnodactyly [HP:0001166] | 0/8 (0%) | 52/95 (55%) | 0.003 | 0.019 |
| Ectopia lentis [HP:0001083] | 1/9 (11%) | 78/100 (78%) | $1.12 \times 10^{-4}$ | 0.001 |
| Stiff skin [HP:0030053] | 8/9 (89%) | 0/50 (0%) | $4.06 \times 10^{-9}$ | $8.52 \times 10^{-8}$ |

**(d)** Fisher Exact Test: exon 37 vs. other. Total of 21 tests were performed.

| HPO term | fs last two | other | p-value | adj. p-value |
|---|---|---|---|---|
| Hyperextensibility of the finger joints [HP:0001187] | 2/2 (100%) | 1/90 (1%) | $7.17 \times 10^{-4}$ | 0.012 |

**(e)** Fisher Exact Test: Frameshift in last two exons vs other. Total of 12 tests were performed.

**Figure S19:** The cohort comprised 144 individuals (57 females, 55 males, 32 with unknown sex). 4 of these individuals were reported to be deceased. A total of 109 HPO terms were used to annotate the cohort. Numerous articles on genotype-phenotype correlations have been published [15, 16, 17], but the available data is said to be be imprecise and incomplete [18] and many studies only share aggregate data. Disease diagnoses: Marfan syndrome (n=51), Ectopia lentis, familial (n=44), Geleophysic dysplasia 2 (n=19), Acromicric dysplasia (n=13), Marfan lipodystrophy syndrome (OMIM:616914) (n=9), Stiff skin syndrome (n=8). A total of 94 unique variant alleles were found in *FBN1* (transcript: `NM_000138.5`, protein id: `NP_000129.3`).

### FBXL4

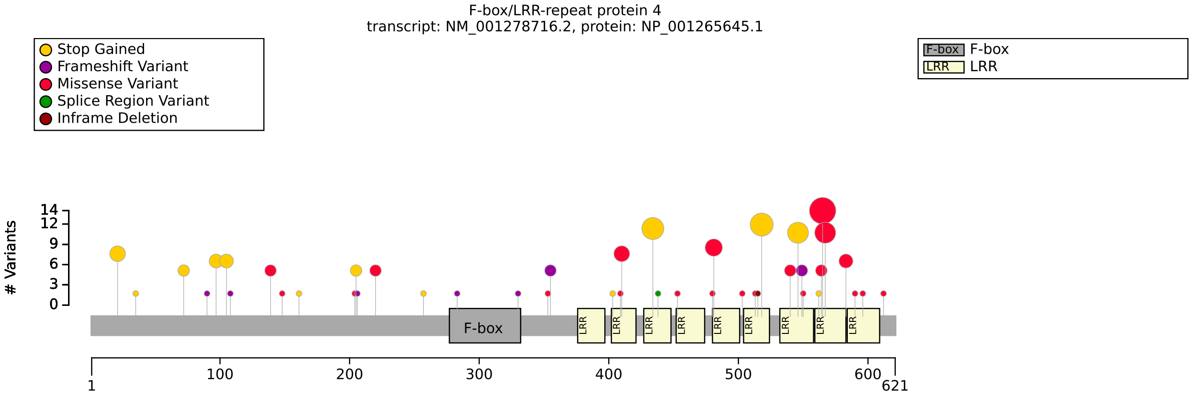


**(a)** Distribution of variants in FBXL4

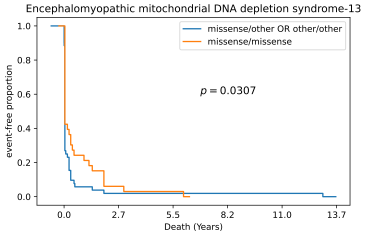


**(b)** Survival analysis.

| HPO term | missense/other OR other/other | missense/missense | p-value | adj. p-value |
|---|---|---|---|---|
| Feeding difficulties [HP:0011968] | 23/24 (96%) | 13/27 (48%) | $1.73 \times 10^{-4}$ | 0.010 |

**(c)** Fisher Exact Test performed to compare HPO annotation frequency with respect to missense/other OR other/other and missense/missense. Total of 55 tests were performed.

| Genotype (A) | Genotype (B) | total tests performed | significant results |
|---|---|---|---|
| LRR domain/LRR domain OR LRR domain/other | other/other | 55 | 0 |
| N-Terminal (1-276)/N-Terminal (1-276) OR N-Terminal (1-276)/other | other/other | 55 | 0 |
| FEMALE | MALE | 53 | 0 |

**(d)** Fisher Exact Test performed to compare HPO annotation frequency with respect to genotypes.

| Description | Variable | Genotype (A) | Genotype (B) | p-value | xrefs |
|---|---|---|---|---|---|
| Age of OMIM:615471 onset | Onset of OMIM:615471 | missense/other OR other/other | missense/missense | 0.031 | - |
| Age at postnatal death | Age of death | missense/other OR other/other | missense/missense | 0.080 | [19] |

**(e)** Survival analysis for Age of death and onset, comparing compare missense/other OR other/other and missense/missense.

**Figure S20:** The cohort comprised 95 individuals (37 females, 58 males). 26 of these individuals were reported to be deceased. A total of 91 HPO terms were used to annotate the cohort. Disease diagnosis: Mitochondrial DNA depletion syndrome 13 (encephalomyopathic type) (OMIM:615471). It was reported that pathological variants in FBXL4 indicated that genotypes with missense variants are frequently associated with longer survival [19]. Using the larger cohort analyzed here, mortality is not significantly associated with missense variants (p-value 0.08), but there was a significant association with age of onset (being later for missense variants) [19]. A total of 51 unique variant alleles were found in *FBXL4* (transcript: `NM_001278716.2`, protein id: `NP_001265645.1`).

### FBXO11

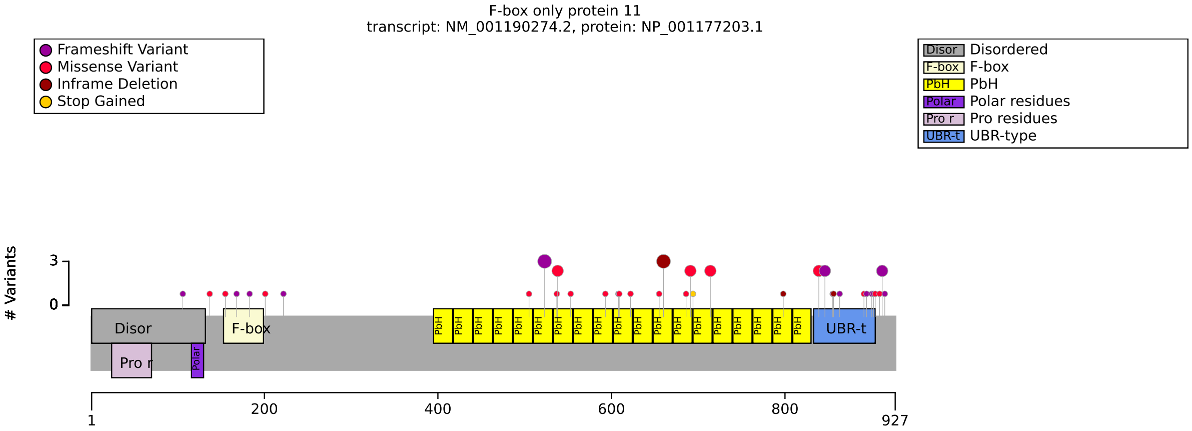


**(a)** Distribution of variants in FBXO11

| Genotype (A) | Genotype (B) | total tests performed | significant results |
|---|---|---|---|
| Missense | Other | 96 | 0 |
| FEMALE | MALE | 96 | 0 |

**(b)** Fisher Exact Test performed to compare HPO annotation frequency with respect to genotypes.

**Figure S21:** The cohort comprised 56 individuals (21 females, 35 males). A total of 128 HPO terms were used to annotate the cohort. Disease diagnosis: Intellectual developmental disorder with dysmorphic facies and behavioral abnormalities (OMIM:618089). No statistically significant results identified. A total of 43 unique variant alleles were found in *FBXO11* (transcript: `NM_001190274.2`, protein id: `NP_001177203.1`).

### FGD1

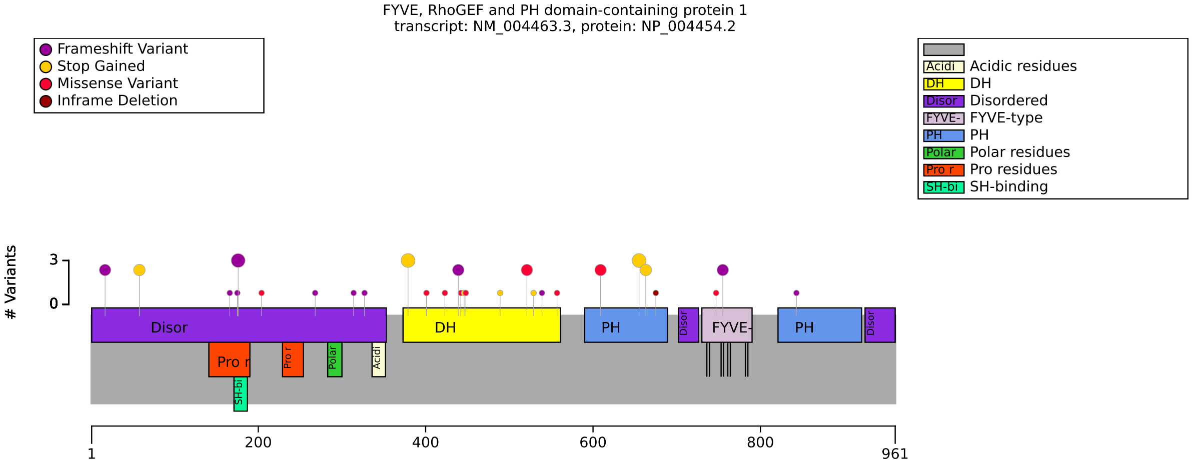


**(a)** Distribution of variants in FGD1

| HPO term | Missense | Other | p-value | adj. p-value |
|---|---|---|---|---|
| Broad foot [HP:0001769] | 1/7 (14%) | 14/15 (93%) | $6.22 \times 10^{-4}$ | 0.032 |

**(b)** Fisher Exact Test performed to compare HPO annotation frequency with respect to Missense and Other. Total of 52 tests were performed.

| Genotype (A) | Genotype (B) | total tests performed | significant results |
|---|---|---|---|
| DH domain | Other | 52 | 0 |

**(c)** Fisher Exact Test performed to compare HPO annotation frequency with respect to genotypes.

**Figure S22:** The cohort comprised 48 individuals (0 females, 48 males). A total of 73 HPO terms were used to annotate the cohort. Disease diagnosis: Aarskog-Scott syndrome (OMIM:305400). Li et al (2024) identified a number of correlations including a lower frequency of Deformity of foot (8/20) with missense than with ”drastic” variants (29/41); p=0.03. Multiple testing correction was not performed [20, 21]. A total of 35 unique variant alleles were found in *FGD1* (transcript: `NM_004463.3`, protein id: `NP_004454.2`).

### FZD5

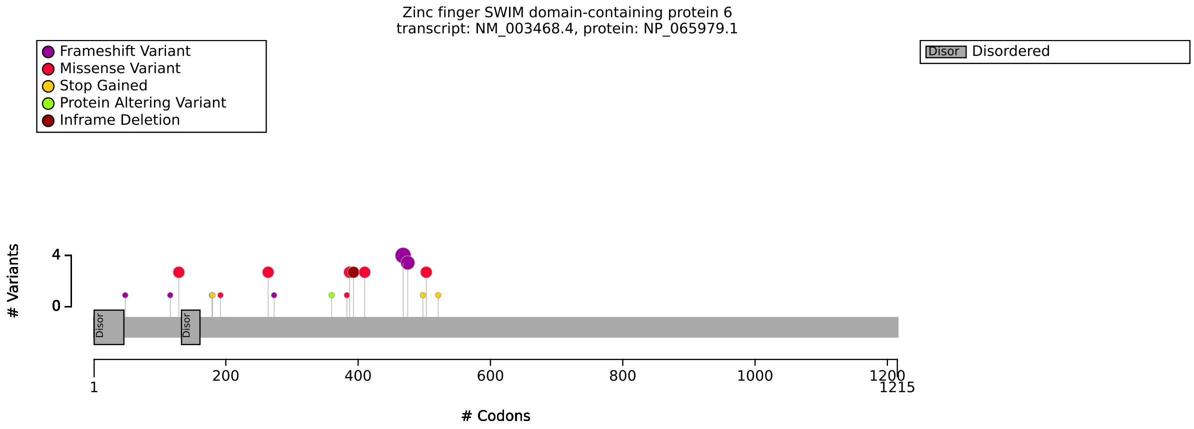


**(a)** Distribution of variants in FZD5

| Genotype (A) | Genotype (B) | total tests performed | significant results |
|---|---|---|---|
| missense | other | 18 | 0 |

**(b)** Fisher Exact Test performed to compare HPO annotation frequency with respect to genotypes.

**Figure S23:** The cohort comprised 29 individuals (11 females, 10 males, 8 with unknown sex). A total of 16 HPO terms were used to annotate the cohort. Disease diagnosis: Microphthalmia/coloboma 11 (OMIM:620731). No significant correlations identified. A total of 19 unique variant alleles were found in *FZD5* (transcript: `NM_003468.4`, protein id: `NP_065979.1`).

### GLI3

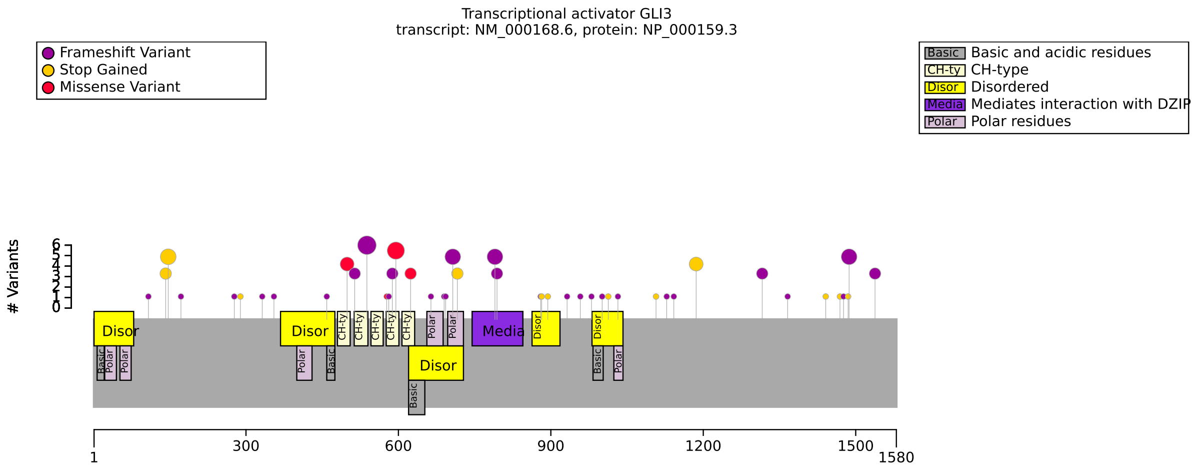


**(a)** Distribution of variants in GLI3

| HPO term | Fs in mid region or splice | other | p-value | adj. p-value |
|---|---|---|---|---|
| Preaxial foot polydactyly [HP:0001841] | 5/25 (20%) | 32/57 (56%) | 0.003 | 0.021 |
| Y-shaped metacarpals [HP:0006042] | 11/23 (48%) | 4/56 (7%) | $9.94 \times 10^{-5}$ | 0.001 |
| Y-shaped metatarsals [HP:0010567] | 11/19 (58%) | 4/50 (8%) | $3.25 \times 10^{-5}$ | $7.80 \times 10^{-4}$ |
| Nail dysplasia [HP:0002164] | 7/20 (35%) | 2/54 (4%) | 0.001 | 0.009 |
| Anal atresia [HP:0002023] | 7/25 (28%) | 3/57 (5%) | 0.007 | 0.036 |

**(b)** Fisher Exact Test performed to compare HPO annotation frequency with respect to Fs in mid region or splice and other. Total of 24 tests were performed.

| HPO term | Truncating variants in Exon 15 | other | p-value | adj. p-value |
|---|---|---|---|---|
| Macrocephaly [HP:0000256] | 13/16 (81%) | 15/42 (36%) | 0.003 | 0.017 |
| Preaxial foot polydactyly [HP:0001841] | 7/28 (25%) | 30/54 (56%) | 0.010 | 0.050 |
| Syndactyly [HP:0001159] | 5/17 (29%) | 33/38 (87%) | $4.82 \times 10^{-5}$ | 0.001 |
| Postaxial foot polydactyly [HP:0001830] | 9/20 (45%) | 2/35 (6%) | $8.91 \times 10^{-4}$ | 0.011 |
| Anal atresia [HP:0002023] | 8/28 (29%) | 2/54 (4%) | 0.002 | 0.017 |

**(c)** Fisher Exact Test performed to compare HPO annotation frequency with respect to Truncating variants in Exon 15 and other. Total of 24 tests were performed.

| HPO term | Variants in C-terminal third | other | p-value | adj. p-value |
|---|---|---|---|---|
| Macrocephaly [HP:0000256] | 13/15 (87%) | 15/43 (35%) | $7.31 \times 10^{-4}$ | 0.004 |
| Postaxial hand polydactyly [HP:0001162] | 15/16 (94%) | 21/49 (43%) | $3.40 \times 10^{-4}$ | 0.003 |
| Syndactyly [HP:0001159] | 5/16 (31%) | 33/39 (85%) | $2.25 \times 10^{-4}$ | 0.003 |
| Postaxial foot polydactyly [HP:0001830] | 9/16 (56%) | 2/39 (5%) | $7.35 \times 10^{-5}$ | 0.002 |

**(d)** Fisher Exact Test performed to compare HPO annotation frequency with respect to Variants in C-terminal third and other. Total of 24 tests were performed.

**Figure S24:** The cohort comprised 82 individuals (3 females, 7 males, 72 with unknown sex). A total of 34 HPO terms were used to annotate the cohort. Disease diagnoses: Greig cephalopolysyndactyly syndrome (OMIM:175700) (51 individuals), Pallister-Hall syndrome (OMIM:146510) (21 individuals), Polydactyly, postaxial, types A1 and B (OMIM:174200) (10 individuals). Genotype-Phenotype-Correlations in GLI3 have been extensive investigated, with findings being similar but not identical to those reported here [22, 23, 24, 25, 26, 27, 28]. A total of 49 unique variant alleles were found in *GLI3* (transcript: `NM_000168.6`, protein id: `NP_000159.3`).

### HMGCS2

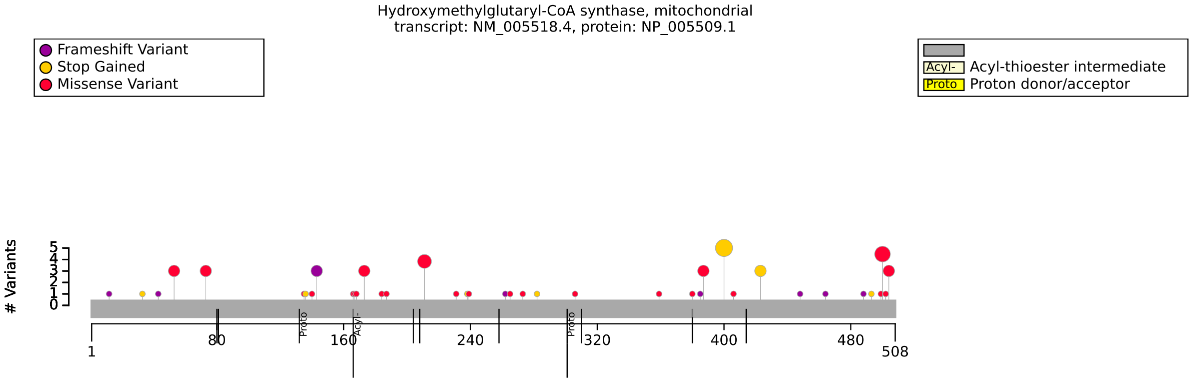


**(a)** Distribution of variants in HMGCS2

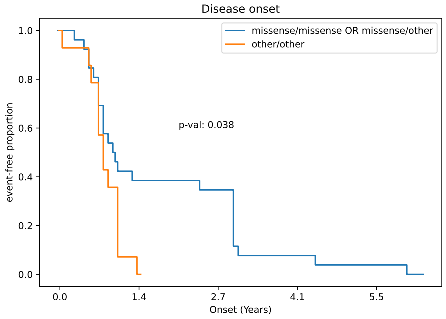


**(b)** Onset of HMGCS2 for missense/missense and missense/other vs. other/other variants.

| Genotype (A) | Genotype (B) | total tests performed | significant results |
|---|---|---|---|
| missense/missense OR missense/other | other/other | 19 | 0 |
| missense/missense OR missense/other | other/other | 19 | 0 |
| FEMALE | MALE | 17 | 0 |

**(c)** Fisher Exact Test performed to compare HPO annotation frequency with respect to genotypes.

| Description | Variable | Genotype (A) | Genotype (B) | p-value | xrefs |
|---|---|---|---|---|---|
| HMG-CoA synthase-2 deficiency (OMIM:605911) disease onset | Onset of OMIM:605911 | missense/missense OR missense/other | other/other | 0.038 | - |

**(d)** Onset of OMIM:605911 to compare missense/missense OR missense/other and other/other with respect to Onset of OMIM:605911.

**Figure S25:** The cohort comprised 40 individuals (14 females, 13 males, 13 with unknown sex). 1 of these individuals were reported to be deceased. A total of 44 HPO terms were used to annotate the cohort. Disease diagnosis: HMG-CoA synthase-2 deficiency (OMIM:605911). No previous publications with results on genotype-phenotype correlations in HMGCS2 were identified. A total of 46 unique variant alleles were found in *HMGCS2* (transcript: `NM_005518.4`, protein id: `NP_005509.1`).

### IKZF1

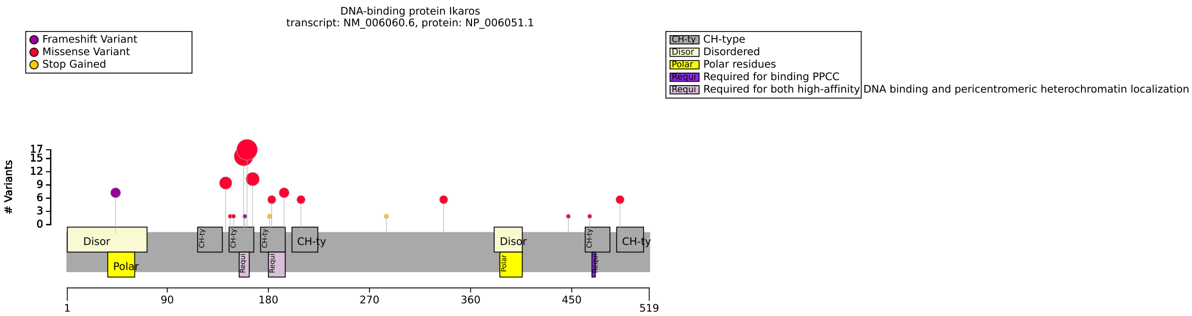


**(a)** Distribution of variants in IKZF1

| HPO term | p.Asn159Ser | Other variant | p-value | adj. p-value |
|---|---|---|---|---|
| Recurrent pneumonia [HP:0006532] | 7/9 (78%) | 7/39 (18%) | 0.001 | 0.035 |
| T-cell acute lymphoblastic leukemias [HP:0006727] | 3/13 (23%) | 0/69 (0%) | 0.003 | 0.047 |

**(b)** Fisher Exact Test performed to compare HPO annotation frequency with respect to p.Asn159Ser and Other variant. Total of 29 tests were performed.

| Genotype (A) | Genotype (B) | total tests performed | significant results |
|---|---|---|---|
| Missense | oher | 34 | 0 |

**(c)** Fisher Exact Test performed to compare HPO annotation frequency with respect to genotypes.

**Figure S26:** The cohort comprised 82 individuals (34 females, 37 males, 11 with unknown sex). 5 of these individuals were reported to be deceased. A total of 66 HPO terms were used to annotate the cohort. Disease diagnosis: Immunodeficiency, common variable, 13 (OMIM:616873). The variant p.Asn159Ser was functionally characterized to be dominant-negative and result in a combined-immunodeficiency phenotype that was distinct from other IKZF1 variants [29]. A total of 26 unique variant alleles were found in *IKZF1* (transcript: NM_006060.6, protein id: NP_006051.1).

### ITPR1

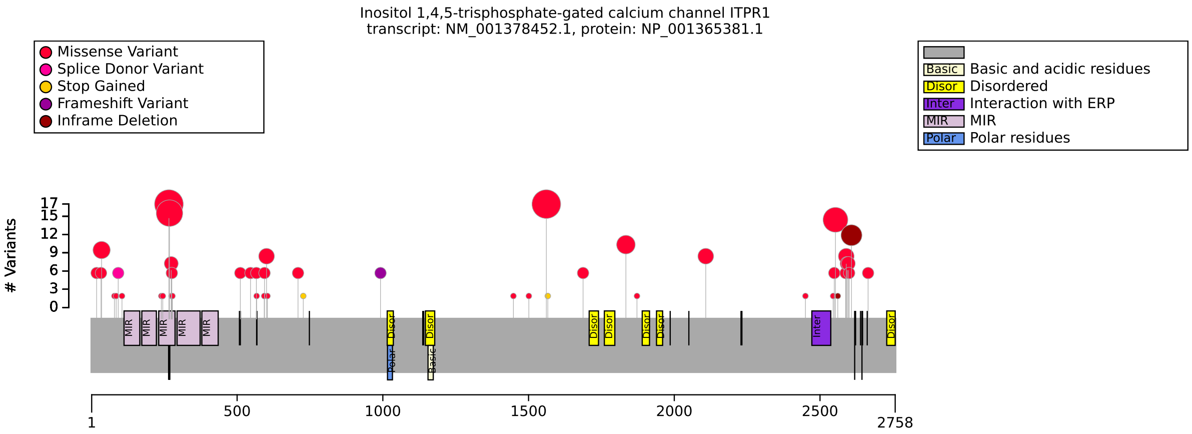


**(a)** Distribution of variants in ITPR1

| HPO term | IP3 binding | other | p-value | adj. p-value |
|---|---|---|---|---|
| Delayed speech and language development [HP:0000750] | 13/13 (100%) | 37/63 (59%) | 0.003 | 0.019 |
| Neurodevelopmental delay [HP:0012758] | 29/29 (100%) | 70/90 (78%) | 0.003 | 0.019 |
| Aniridia [HP:0000526] | 0/14 (0%) | 20/54 (37%) | 0.007 | 0.024 |
| Delayed gross motor development [HP:0002194] | 18/18 (100%) | 57/83 (69%) | 0.005 | 0.024 |
| Motor delay [HP:0001270] | 24/24 (100%) | 63/89 (71%) | 0.002 | 0.019 |
| Nystagmus [HP:0000639] | 25/26 (96%) | 53/78 (68%) | 0.003 | 0.019 |
| Delayed ability to sit [HP:0025336] | 11/11 (100%) | 42/70 (60%) | 0.013 | 0.043 |
| Delayed ability to walk [HP:0031936] | 13/13 (100%) | 41/68 (60%) | 0.004 | 0.019 |

**(b)** Fisher Exact Test performed to compare HPO annotation frequency with respect to IP3 binding and other. Total of 26 tests were performed.

| HPO term | SV Deletion | other | p-value | adj. p-value |
|---|---|---|---|---|
| Delayed speech and language development [HP:0000750] | 0/19 (0%) | 50/57 (88%) | $1.72 \times 10^{-12}$ | $7.57 \times 10^{-12}$ |
| Neurodevelopmental delay [HP:0012758] | 0/19 (0%) | 99/100 (99%) | $4.07 \times 10^{-21}$ | $8.96 \times 10^{-20}$ |
| Delayed gross motor development [HP:0002194] | 0/19 (0%) | 75/82 (91%) | $4.04 \times 10^{-15}$ | $2.96 \times 10^{-14}$ |
| Motor delay [HP:0001270] | 0/19 (0%) | 87/94 (93%) | $3.90 \times 10^{-16}$ | $4.29 \times 10^{-15}$ |
| Global developmental delay [HP:0001263] | 0/19 (0%) | 48/57 (84%) | $1.81 \times 10^{-11}$ | $5.68 \times 10^{-11}$ |
| Nystagmus [HP:0000639] | 17/17 (100%) | 61/87 (70%) | 0.006 | 0.016 |
| Delayed ability to sit [HP:0025336] | 0/19 (0%) | 53/62 (85%) | $4.56 \times 10^{-12}$ | $1.67 \times 10^{-11}$ |
| Delayed ability to walk [HP:0031936] | 0/19 (0%) | 54/62 (87%) | $1.47 \times 10^{-12}$ | $7.57 \times 10^{-12}$ |

**(c)** Fisher Exact Test performed to compare HPO annotation frequency with respect to SV Deletion and other. Total of 22 tests were performed.

| HPO term | GS Hotspot | other | p-value | adj. p-value |
|---|---|---|---|---|
| Aniridia [HP:0000526] | 14/20 (70%) | 6/48 (12%) | $6.13 \times 10^{-6}$ | $1.59 \times 10^{-4}$ |
| Delayed gross motor development [HP:0002194] | 18/18 (100%) | 57/83 (69%) | 0.005 | 0.032 |
| Motor delay [HP:0001270] | 19/19 (100%) | 68/94 (72%) | 0.006 | 0.032 |
| Delayed ability to sit [HP:0025336] | 13/13 (100%) | 40/68 (59%) | 0.003 | 0.030 |
| Delayed ability to walk [HP:0031936] | 14/14 (100%) | 40/67 (60%) | 0.003 | 0.030 |

**(d)** Fisher Exact Test performed to compare HPO annotation frequency with respect to GS Hotspot and other. Total of 25 tests were performed.

**Figure S27:** The cohort comprised 170 individuals (65 females, 41 males, 64 with unknown sex). 1 of these individuals was reported to be deceased. A Fisher exact test for male/female differences revealed no significant associations. A total of 196 HPO terms were used to annotate the cohort. Disease diagnoses: Spinocerebellar ataxia 29, congenital nonprogressive (OMIM:117360) (104 individuals), Gillespie syndrome (OMIM:206700) (39 individuals), Spinocerebellar ataxia 15 (OMIM:606658) (27 individuals). A total of 62 unique variant alleles were found in *ITPR1* (transcript: `NM_001378452.1`, protein id: `NP_001365381.1`).

### Kabuki

| HPO term | OMIM:147920 | OMIM:300867 | p-value | adj. p-value |
|---|---|---|---|---|
| Feeding difficulties [HP:0011968] | 8/25 (32%) | 55/63 (87%) | $7.41 \times 10^{-7}$ | $2.52 \times 10^{-5}$ |
| Motor delay [HP:0001270] | 4/10 (40%) | 58/61 (95%) | $1.04 \times 10^{-4}$ | 0.002 |

**(a)** Fisher Exact Test performed to compare HPO annotation frequency with respect to OMIM:147920 and OMIM:300867. Total of 34 tests were performed.

**Figure S28:** The cohort comprised 146 individuals (81 females, 65 males). 5 of these individuals were reported to be deceased. A total of 200 HPO terms were used to annotate the cohort. Disease diagnoses: Kabuki syndrome 2 (OMIM:300867) (81 individuals), Kabuki Syndrome 1 (OMIM:147920) (65 individuals). Previous studies have found that the phenotype of KS2 is different from that of KS1, for example, patients with KS1 type have a higher risk of typical facial features, short stature, and frequent infections compared to those with KS2 [30, 31, 32]. A total of 105 unique variant alleles were found in *n/a* (transcript: `n/a`, protein id: `n/a`).

### KCNH5

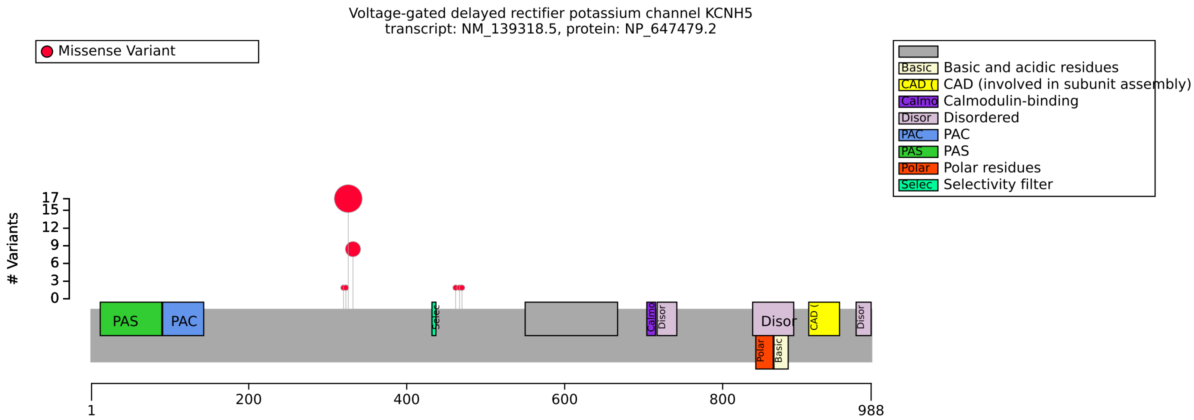


**(a)** Distribution of variants in KCNH5

| HPO term | Arg327His | Arg333His | p-value | adj. p-value |
|---|---|---|---|---|
| Epileptic encephalopathy [HP:0200134] | 15/15 (100%) | 0/3 (0%) | 0.001 | 0.032 |

**(b)** Fisher Exact Test performed to compare HPO annotation frequency with respect to Arg327His and Arg333His. Total of 26 tests were performed.

| Genotype (A) | Genotype (B) | total tests performed | significant results |
|---|---|---|---|
| FEMALE | MALE | 32 | 0 |

**(c)** Fisher Exact Test performed to compare HPO annotation frequency with respect to genotypes.

**Figure S29:** The cohort comprised 27 individuals (14 females, 13 males). A total of 28 HPO terms were used to annotate the cohort. Disease diagnosis: Developmental and epileptic encephalopathy 112 (OMIM:620537). Two recurrent variants have been reported, p.Arg327His and p.Arg333His, both of which are located in or near the functionally critical voltage-sensing or pore domains. Happ et al (2023) state that in their cohort, Individuals with the recurrent p.Arg333His variant had a self-limited drug-responsive focal or generalized epilepsy and normal intellect, whereas the recurrent p.Arg327His variant was associated with infantile-onset DEE [33]. A total of 8 unique variant alleles were found in *KCNH5* (transcript: `NM_139318.5`, protein id: `NP_647479.2`).

### KDM6A

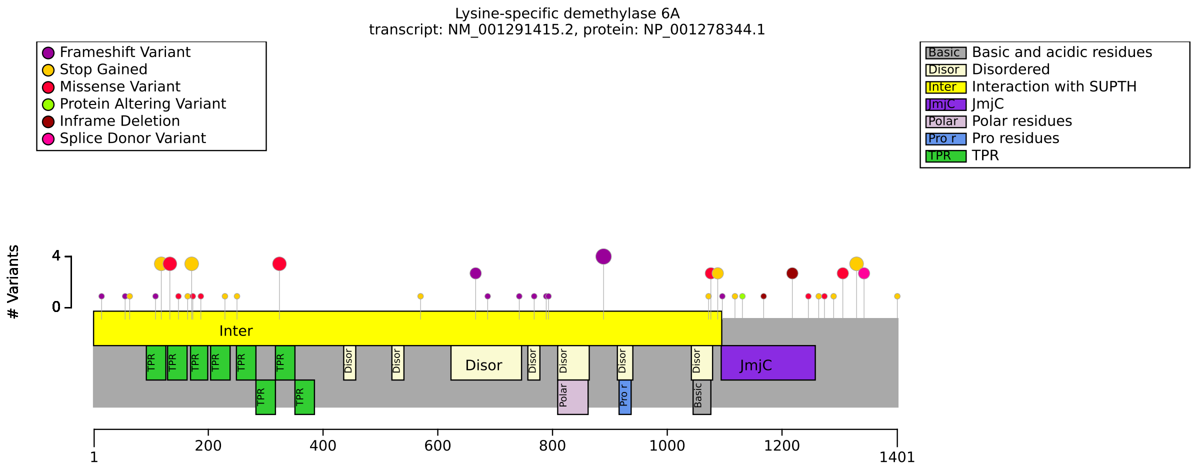


**(a)** Distribution of variants in KDM6A

| HPO term | FEMALE | MALE | p-value | adj. p-value |
|---|---|---|---|---|
| Intellectual disability, severe [HP:0010864] | 7/25 (28%) | 14/18 (78%) | 0.002 | 0.008 |

**(b)** Fisher Exact Test performed to compare HPO annotation frequency with respect to FEMALE and MALE. Total of 4 tests were performed. A similar sex difference was reported in ref. [34].

| HPO term | p.Asn891ValfsTer27 | Other | p-value | adj. p-value |
|---|---|---|---|---|
| Pulmonic stenosis [HP:0001642] | 2/2 (100%) | 0/59 (0%) | $5.46 \times 10^{-4}$ | 0.016 |

**(c)** Fisher Exact Test performed to compare HPO annotation frequency with respect to p.Asn891ValfsTer27 and Other. Total of 29 tests were performed.

| Genotype (A) | Genotype (B) | total tests performed | significant results |
|---|---|---|---|
| missense | other | 47 | 0 |
| SV | other | 47 | 0 |

**(d)** Fisher Exact Test performed to compare HPO annotation frequency with respect to genotypes.

**Figure S30:** The cohort comprised 81 individuals (46 females, 35 males). A total of 87 HPO terms were used to annotate the cohort. Disease diagnosis: Kabuki syndrome 2 (OMIM:300867). . A total of 53 unique variant alleles were found in *KDM6A* (transcript: `NM_001291415.2`, protein id: `NP_001278344.1`).

### KDM6B

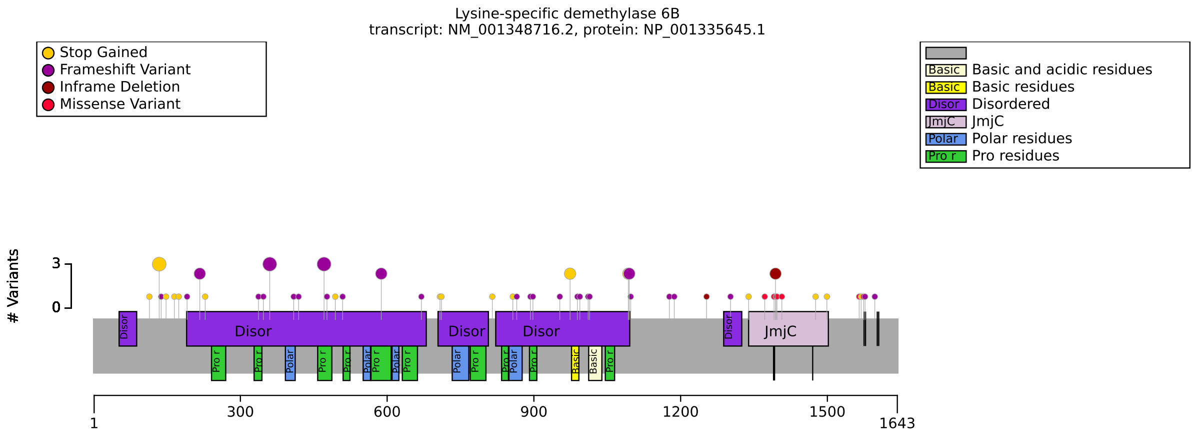


**(a)** Distribution of variants in KDM6B

| Genotype (A) | Genotype (B) | total tests performed | significant results |
|---|---|---|---|
| JmjC domain | Other | 54 | 0 |
| N term | C term | 54 | 0 |

**(b)** Fisher Exact Test performed to compare HPO annotation frequency with respect to genotypes.

**Figure S31:** The cohort comprised 73 individuals (19 females, 54 males). A total of 251 HPO terms were used to annotate the cohort. Disease diagnosis: Neurodevelopmental disorder with coarse facies and mild distal skeletal abnormalities (OMIM:618505). Stolerman et al (2016) identified no significant genotype-phenotype correlations [35] A total of 61 unique variant alleles were found in *KDM6B* (transcript: `NM_001348716.2`, protein id: `NP_001335645.1`).

### KMT2D

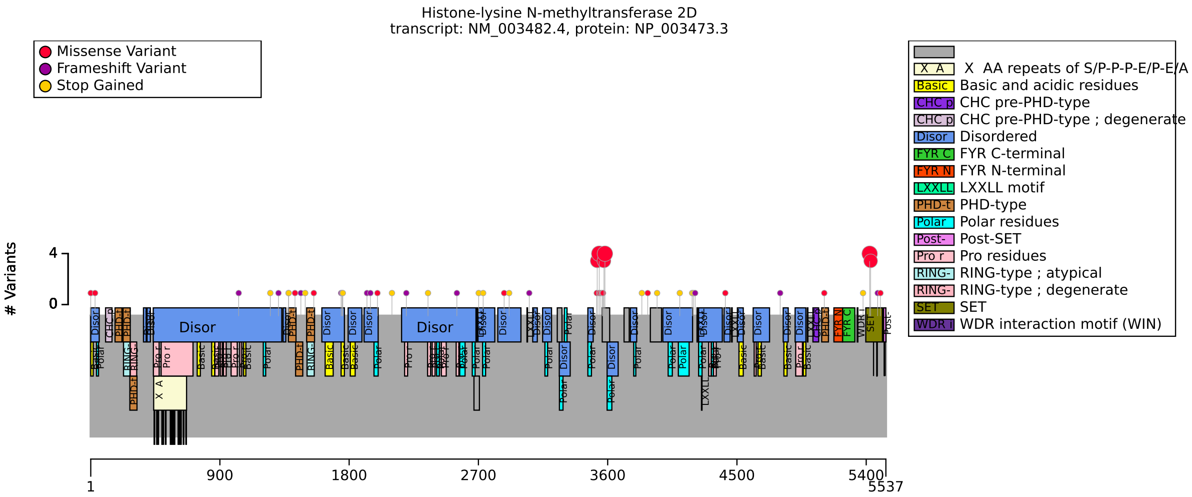


**(a)** Distribution of variants in KMT2D

| Genotype (A) | Genotype (B) | total tests performed | significant results |
|---|---|---|---|
| FEMALE | MALE | 42 | 0 |
| missense | other | 42 | 0 |
| n_term | other | 41 | 0 |
| p.Glu5425Lys | other | 36 | 0 |
| p.Leu3542Pro | Other variant | 8 | 0 |

**(b)** Fisher Exact Test performed to compare HPO annotation frequency with respect to genotypes.

**Figure S32:** The cohort comprised 65 individuals (35 females, 30 males). 5 of these individuals were reported to be deceased. A total of 151 HPO terms were used to annotate the cohort. Disease diagnosis: Kabuki Syndrome 1 (OMIM:147920). A total of 52 unique variant alleles were found in *KMT2D* (transcript: `NM_003482.4`, protein id: `NP_003473.3`).

### LDS 1 and 2

| Genotype (A) | Genotype (B) | total tests performed | significant results |
|---|---|---|---|
| OMIM:609192 | OMIM:610168 | 36 | 0 |

**(a)** Fisher Exact Test performed to compare HPO annotation frequency with respect to genotypes.

**Figure S33:** The cohort comprised 94 individuals (30 females, 46 males, 18 with unknown sex). 6 of these individuals were reported to be deceased. A total of 110 HPO terms were used to annotate the cohort. Disease diagnoses: Loeys-Dietz syndrome 2 (OMIM:610168) (53 individuals), Loeys-Dietz syndrome 1 (OMIM:609192) (23 individuals), Multiple self-healing squamous epithelioma, susceptibility to (OMIM:132800) (18 individuals). A total of 61 unique variant alleles were found.

### LDS 1 and 3

| HPO term | OMIM:609192 | OMIM:613795 | p-value | adj. p-value |
|---|---|---|---|---|
| Scoliosis [HP:0002650] | 18/21 (86%) | 20/43 (47%) | 0.003 | 0.027 |
| Hypertelorism [HP:0000316] | 15/19 (79%) | 13/35 (37%) | 0.004 | 0.027 |
| Aortic aneurysm [HP:0004942] | 11/11 (100%) | 26/48 (54%) | 0.004 | 0.027 |
| Osteoarthritis [HP:0002758] | 0/11 (0%) | 26/38 (68%) | $4.64 \times 10^{-5}$ | 0.001 |

**(a)** Fisher Exact Test performed to compare HPO annotation frequency with respect to OMIM:609192 and OMIM:613795. Total of 24 tests were performed.

**Figure S34:** The cohort comprised 90 individuals (29 females, 43 males, 18 with unknown sex). 2 of these individuals were reported to be deceased. A total of 89 HPO terms were used to annotate the cohort. Disease diagnoses: Loeys-Dietz syndrome 3 (OMIM:613795) (49 individuals), Loeys-Dietz syndrome 1 (OMIM:609192) (23 individuals), Multiple self-healing squamous epithelioma, susceptibility to (OMIM:132800) (18 individuals). A total of 37 unique variant alleles were found.

#### LDS 3 and 6

| HPO term | OMIM:613795 | OMIM:619656 | p-value | adj. p-value |
|---|---|---|---|---|
| Thoracic aortic aneurysm [HP:0012727] | 0/22 (0%) | 10/16 (62%) | $1.69 \times 10^{-5}$ | $4.23 \times 10^{-4}$ |

**(a)** Fisher Exact Test performed to compare HPO annotation frequency with respect to OMIM:613795 and OMIM:619656. Total of 25 tests were performed.

**Figure S35:** The cohort comprised 72 individuals (35 females, 35 males, 2 with unknown sex). A total of 98 HPO terms were used to annotate the cohort. Disease diagnoses: Loeys-Dietz syndrome 3 (OMIM:613795) (49 individuals), Loeys-Dietz syndrome 6 (OMIM:619656) (18 individuals), Congenital heart defects, multiple types, 8, with or without heterotaxy (OMIM:619657) (5 individuals). A total of 25 unique variant alleles were found.

### LMNA (Part 1 of 2)

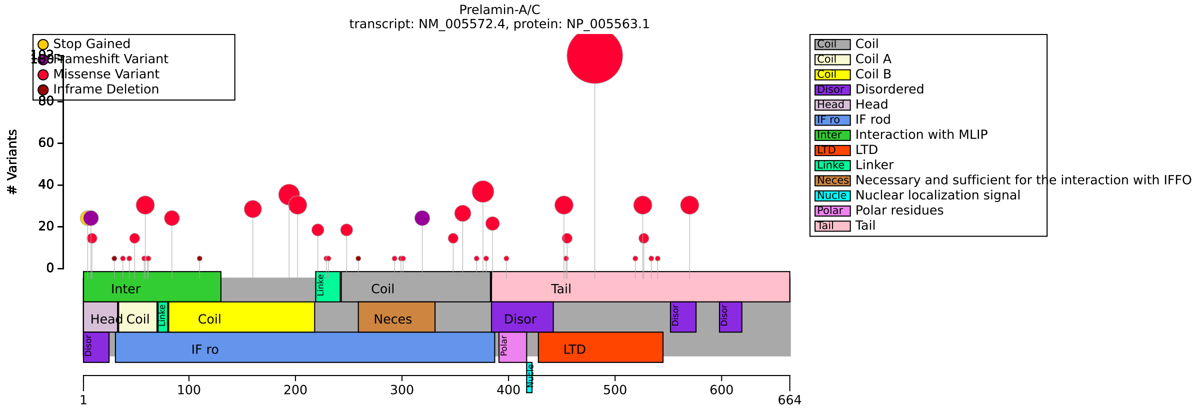


**(a)** Distribution of variants in LMNA

| HPO term | missense | other | p-value | adj. p-value |
|---|---|---|---|---|
| Loss of truncal subcutaneous adipose tissue [HP:0009002] | 104/104 (100%) | 4/11 (36%) | $7.53 \times 10^{-9}$ | $2.86 \times 10^{-7}$ |
| Elevated hemoglobin A1c [HP:0040217] | 73/101 (72%) | 6/24 (25%) | $3.11 \times 10^{-5}$ | $5.90 \times 10^{-4}$ |

**(b)** Fisher Exact Test performed to compare HPO annotation frequency with respect to missense and other. Total of 38 tests were performed.

| HPO term | Upstream of NLS | other | p-value | adj. p-value |
|---|---|---|---|---|
| Lipodystrophy [HP:0009125] | 10/93 (11%) | 132/158 (84%) | $1.73 \times 10^{-31}$ | $6.58 \times 10^{-30}$ |
| Pancreatitis [HP:0001733] | 4/7 (57%) | 14/110 (13%) | 0.011 | 0.033 |
| Dilated cardiomyopathy [HP:0001644] | 35/70 (50%) | 5/102 (5%) | $4.20 \times 10^{-12}$ | $7.98 \times 10^{-11}$ |
| Second degree atrioventricular block [HP:0011706] | 6/68 (9%) | 1/115 (1%) | 0.011 | 0.033 |
| Atrioventricular block [HP:0001678] | 17/36 (47%) | 8/116 (7%) | $2.11 \times 10^{-7}$ | $2.67 \times 10^{-6}$ |
| Achilles tendon contracture [HP:0001771] | 21/85 (25%) | 19/30 (63%) | $2.66 \times 10^{-4}$ | 0.001 |

**(c)** Fisher Exact Test: HPO annotation frequency and NLS Upstream vs other variants. 38 tests were performed in total.

**Figure S36:** See following page for caption

### LMNA (Part 2 of 2)

| HPO term | Upstream of NLS | other | p-value | adj. p-value |
|---|---|---|---|---|
| Foot joint contracture [HP:0008366] | 21/81 (26%) | 19/27 (70%) | $6.20 \times 10^{-5}$ | $5.89 \times 10^{-4}$ |
| Lower-limb joint contracture [HP:0005750] | 22/82 (27%) | 19/27 (70%) | $8.20 \times 10^{-5}$ | $6.23 \times 10^{-4}$ |
| Limb joint contracture [HP:0003121] | 23/83 (28%) | 19/27 (70%) | $1.71 \times 10^{-4}$ | $8.14 \times 10^{-4}$ |
| Elbow contracture [HP:0034391] | 21/83 (25%) | 18/26 (69%) | $1.04 \times 10^{-4}$ | $6.59 \times 10^{-4}$ |
| Upper-limb joint contracture [HP:0100360] | 21/81 (26%) | 18/26 (69%) | $1.23 \times 10^{-4}$ | $6.66 \times 10^{-4}$ |
| Hip contracture [HP:0003273] | 4/83 (5%) | 9/26 (35%) | $2.68 \times 10^{-4}$ | 0.001 |
| First degree atrioventricular block [HP:0011705] | 7/63 (11%) | 2/115 (2%) | 0.010 | 0.033 |

**(a)** Fisher Exact Test performed to compare HPO annotation frequency with respect to NLS Upstream and other. Total of 38 tests were performed.

| HPO term | Gly608= | Other | p-value | adj. p-value |
|---|---|---|---|---|
| Lipodystrophy [HP:0009125] | 15/15 (100%) | 127/236 (54%) | $1.87 \times 10^{-4}$ | $7.47 \times 10^{-4}$ |
| Elevated hemoglobin A1c [HP:0040217] | 0/15 (0%) | 79/110 (72%) | $5.65 \times 10^{-8}$ | $6.77 \times 10^{-7}$ |
| Muscle weakness [HP:0001324] | 0/15 (0%) | 63/124 (51%) | $6.07 \times 10^{-5}$ | $3.64 \times 10^{-4}$ |
| Proximal muscle weakness [HP:0003701] | 0/15 (0%) | 53/114 (46%) | $3.63 \times 10^{-4}$ | 0.001 |
| Distal muscle weakness [HP:0002460] | 0/15 (0%) | 36/97 (37%) | 0.002 | 0.004 |
| Proximal muscle weakness in upper limbs [HP:0008997] | 0/15 (0%) | 35/102 (34%) | 0.005 | 0.007 |
| Upper limb muscle weakness [HP:0003484] | 0/15 (0%) | 35/96 (36%) | 0.003 | 0.004 |
| Limb muscle weakness [HP:0003690] | 0/15 (0%) | 38/99 (38%) | 0.002 | 0.004 |

**(b)** Fisher Exact Test performed to compare HPO annotation frequency with respect to A and B. Total of 12 tests were performed.

| Description | Variable | Genotype (A) | Genotype (B) | p-value | xrefs |
|---|---|---|---|---|---|
| Phenotype score (see notebook) | HPO group count | NLS Upstream | other | $1.83 \times 10^{-16}$ | [36] |

**(c)** HPO Group Count to compare NLS Upstream and other with respect to HPO group count.

**Figure S37:** The cohort comprised 266 individuals (150 females, 99 males, 17 with unknown sex). 11 of these individuals were reported to be deceased. A total of 152 HPO terms were used to annotate the cohort. Disease diagnoses: Lipodystrophy, familial partial, type 2 (OMIM:151660) (127 individuals), Cardiomyopathy, dilated, 1A (OMIM:115200) (68 individuals), Emery-Dreifuss muscular dystrophy 2, autosomal dominant (OMIM:181350) (41 individuals), Hutchinson-Gilford progeria (OMIM:176670) (15 individuals), LMNA-related congenital muscular dystrophy (OMIM:613205) (15 individuals). Numerous LMNA genotype-phenotype correlations have been described. The literature is summarized in [37]. Cardiac involvement in multisystem laminopathies prevails with mutations upstream of the nuclear localisation signal [36]. A total of 55 unique variant alleles were found in *LMNA* (transcript: `NM_005572.4`, protein id: `NP_005563.1`).

### LZTR1

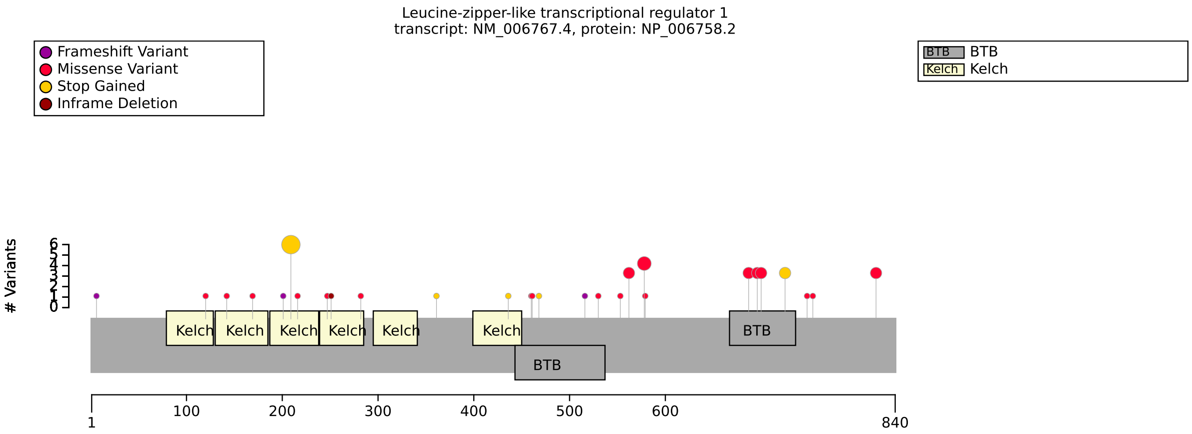


**(a)** Distribution of variants in LZTR1

| Genotype (A) | Genotype (B) | total tests performed | significant results |
|---|---|---|---|
| N Term/N Term OR N Term/Other | Other/Other | 49 | 0 |
| Missense/Missense OR Missense/Other | Other/Other | 49 | 0 |
| FEMALE | MALE | 54 | 0 |

**(b)** Fisher Exact Test performed to compare HPO annotation frequency with respect to genotypes.

**Figure S38:** The cohort comprised 38 individuals (18 females, 20 males). 3 of these individuals were reported to be deceased. A total of 96 HPO terms were used to annotate the cohort. Disease diagnosis: Noonan syndrome 2 (OMIM:605275). No significant association identified. A total of 38 unique variant alleles were found in *LZTR1* (transcript: `NM_006767.4`, protein id: `NP_006758.2`).

### MAPK8IP3

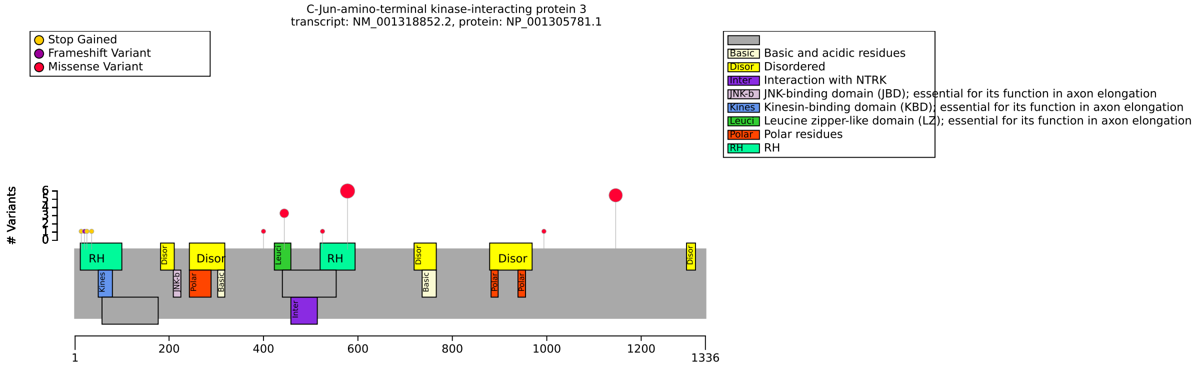


**(a)** Distribution of variants in MAPK8IP3

| Genotype (A) | Genotype (B) | total tests performed | significant results |
|---|---|---|---|
| N term | other | 68 | 0 |
| p.Arg579Cys | Other variant | 68 | 0 |
| RH2 | Other region | 68 | 0 |
| FEMALE | MALE | 68 | 0 |

**(b)** Fisher Exact Test performed to compare HPO annotation frequency with respect to genotypes.

**Figure S39:** The cohort comprised 20 individuals (9 females, 11 males). A total of 93 HPO terms were used to annotate the cohort. Disease diagnosis: Neurodevelopmental disorder with or without variable brain abnormalities (OMIM:618443). No significant association identified. A total of 10 unique variant alleles were found in *MAPK8IP3* (transcript: `NM_001318852.2`, protein id: `NP_001305781.1`).

### MPV17

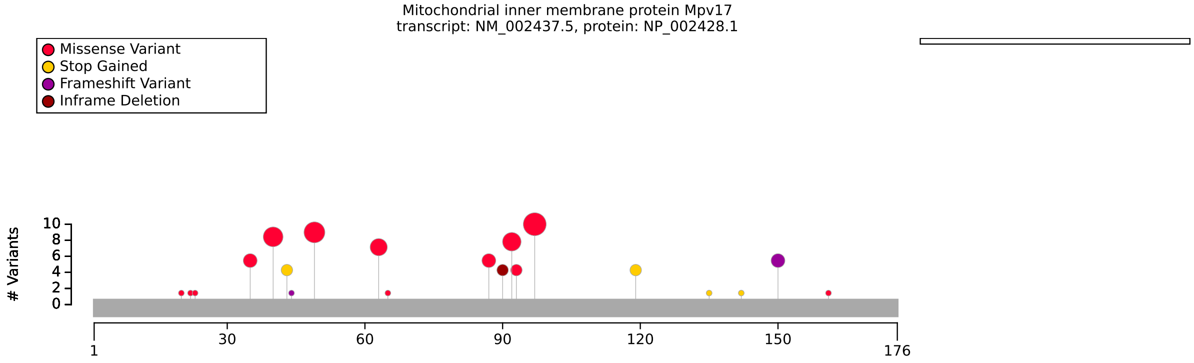


**(a)** Distribution of variants in MPV17

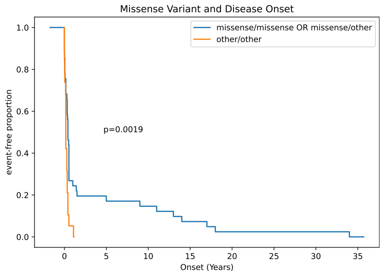


**(b)** MPV17 disease onset

| HPO term | Pro98Leu/Pro98Leu | other/other OR Pro98Leu/other | p-value | adj. p-value |
|---|---|---|---|---|
| Peripheral axonal neuropathy [HP:0003477] | 3/3 (100%) | 1/22 (5%) | 0.002 | 0.037 |

**(c)** Fisher Exact Test performed to compare HPO annotation frequency with respect to Pro98Leu/Pro98Leu and other/other OR Pro98Leu/other. Total of 21 tests were performed.

| Genotype (A) | Genotype (B) | total tests performed | significant results |
|---|---|---|---|
| missense/missense OR missense/other | other/other | 32 | 0 |
| Arg50Gln/Arg50Gln OR Arg50Gln/other | other/other | 29 | 0 |
| FEMALE | MALE | 32 | 0 |

**(d)** Fisher Exact Test performed to compare HPO annotation frequency with respect to genotypes.

| Description | Variable | Genotype (A) | Genotype (B) | p-value | xrefs |
|---|---|---|---|---|---|
| MTDPS6 (OMIM:256810) age at death | Age of death | Pro98Leu/Pro98Leu | other/other OR Pro98Leu/other | 0.010 | [38, 39] |

**(e)** Age of death to compare Pro98Leu/Pro98Leu and other/other OR Pro98Leu/other with respect to Age of death.

| Description | Variable | Genotype (A) | Genotype (B) | p-value | xrefs |
|---|---|---|---|---|---|
| MTDPS6 (OMIM:256810) disease onset | Onset of OMIM:256810 | missense/missense OR missense/other | other/other | 0.002 | - |

**(f)** Onset of OMIM:256810 to compare missense/missense OR missense/other and other/other with respect to Onset of OMIM:256810.

**Figure S40:** The cohort comprised 60 individuals (30 females, 30 males). 39 of these individuals were reported to be deceased. A total of 156 HPO terms were used to annotate the cohort. Disease diagnosis: Mitochondrial DNA depletion syndrome 6 (hepatocerebral type) (MTDPS6; OMIM:256810). No clear genotype-phenotype correlation exists. However, a trend for longer survival can be observed in individuals with biallelic pathogenic missense variants compared to individuals with biallelic null [38, 39]. A total of 31 unique variant alleles were found in *MPV17* (transcript: `NM_002437.5`, protein id: `NP_002428.1`).

### NBAS

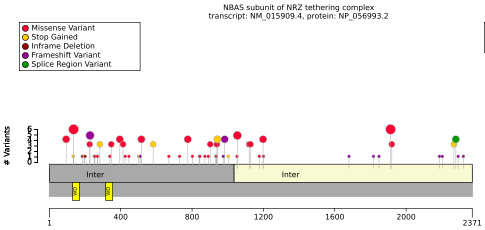


**(a)** Distribution of variants in NBAS

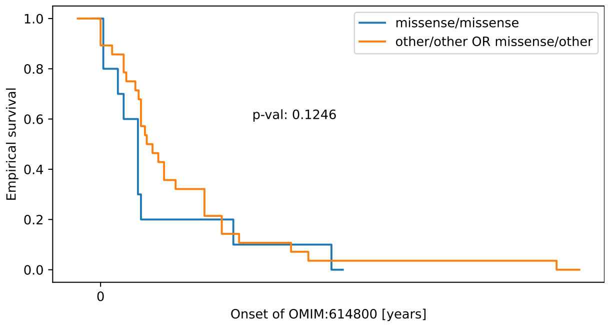


**(b)** NBAS missense variants vs. age at death.

| HPO term | missense/missense | other/other OR missense/other | p-value | adj. p-value |
|---|---|---|---|---|
| Decreased circulating IgG concentration [HP:0004315] | 1/12 (8%) | 15/23 (65%) | 0.002 | 0.035 |

**(c)** Fisher Exact Test performed to compare HPO annotation frequency with respect to missense/missense and other/other OR missense/other. Total of 22 tests were performed.

| Description | Variable | Genotype (A) | Genotype (B) | p-value | xrefs |
|---|---|---|---|---|---|
| Age of death | Age of death | missense/missense | other/other OR missense/other | 0.125 | - |

**(d)** Age of death to compare missense/missense and other/other OR missense/other with respect to Age of death.

**Figure S41:** The cohort comprised 67 individuals (19 females, 21 males, 27 with unknown sex). 9 of these individuals were reported to be deceased. A total of 134 HPO terms were used to annotate the cohort. Disease diagnosis: Short stature, optic nerve atrophy, and Pelger-Huet anomaly (OMIM:614800). It was reported that missense or in-frame deletions in the C-terminal region of the NBAS gene are associated with SOPH syndrome (614800), missense or in-frame deletions in the Sec30 domain of NBAS are associated with infantile liver failure syndrome type 2 (616483), while missense or in-frame deletions in the beta-propeller domain of NBAS are associated with a combined phenotype of multisystem involvement with acute liver failure [40]. In our dataset, we found an association with missense or in-frame deletions in the Sec30 with decreased circulating IgG. A similar finding was obtain for missense variants in general. Due to the small size of our cohort, this finding should be regarded as preliminary. A total of 74 unique variant alleles were found in *NBAS* (transcript: `NM_015909.4`, protein id: `NP_056993.2`).

# NF1

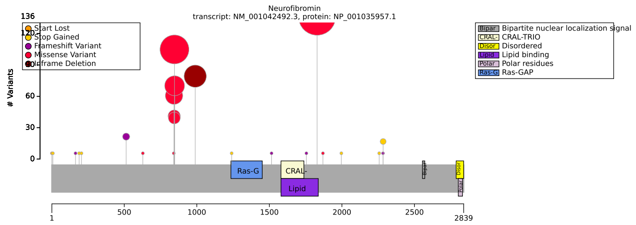


**(a)** Distribution of variants in NF1

| HPO term | p.Arg1830 | other | p-value | adj. p-value |
|---|---|---|---|---|
| Axillary freckling [HP:0000997] | 20/79 (25%) | 105/175 (60%) | $3.66 \times 10^{-7}$ | $2.01 \times 10^{-6}$ |
| Freckling [HP:0001480] | 76/133 (57%) | 201/222 (91%) | $6.28 \times 10^{-13}$ | $6.91 \times 10^{-12}$ |
| Plexiform neurofibroma [HP:0009732] | 0/135 (0%) | 68/260 (26%) | $1.71 \times 10^{-14}$ | $3.76 \times 10^{-13}$ |
| Neurofibroma [HP:0001067] | 5/8 (62%) | 153/162 (94%) | 0.012 | 0.030 |
| Scoliosis [HP:0002650] | 8/125 (6%) | 48/240 (20%) | $4.02 \times 10^{-4}$ | 0.001 |
| Lisch nodules [HP:0009737] | 11/91 (12%) | 72/193 (37%) | $6.58 \times 10^{-6}$ | $2.90 \times 10^{-5}$ |
| Inguinal freckling [HP:0030052] | 8/79 (10%) | 84/170 (49%) | $4.86 \times 10^{-10}$ | $3.56 \times 10^{-9}$ |
| Pulmonic stenosis [HP:0001642] | 13/105 (12%) | 5/162 (3%) | 0.005 | 0.013 |
| Optic nerve glioma [HP:0009734] | 0/39 (0%) | 39/137 (28%) | $1.71 \times 10^{-5}$ | $6.28 \times 10^{-5}$ |

**(b)** Fisher Exact Test performed to compare HPO annotation frequency with respect to p.Arg1830 and other. Total of 22 tests were performed.

| HPO term | Met992del | other | p-value | adj. p-value |
|---|---|---|---|---|
| Axillary freckling [HP:0000997] | 0/14 (0%) | 125/240 (52%) | $8.57 \times 10^{-5}$ | 0.001 |
| Plexiform neurofibroma [HP:0009732] | 0/44 (0%) | 68/351 (19%) | $2.05 \times 10^{-4}$ | 0.001 |
| Lisch nodules [HP:0009737] | 3/36 (8%) | 80/248 (32%) | 0.003 | 0.008 |
| Inguinal freckling [HP:0030052] | 0/14 (0%) | 92/235 (39%) | 0.003 | 0.008 |
| Lipoma [HP:0012032] | 5/44 (11%) | 4/284 (1%) | 0.003 | 0.008 |

**(c)** Fisher Exact Test performed to compare HPO annotation frequency with respect to Met992del and other. Total of 13 tests were performed.

| HPO term | Leu847Pro | other | p-value | adj. p-value |
|---|---|---|---|---|
| Axillary freckling [HP:0000997] | 50/61 (82%) | 75/193 (39%) | $3.55 \times 10^{-9}$ | $7.81 \times 10^{-8}$ |
| Freckling [HP:0001480] | 54/54 (100%) | 223/301 (74%) | $5.30 \times 10^{-7}$ | $5.83 \times 10^{-6}$ |
| Plexiform neurofibroma [HP:0009732] | 24/66 (36%) | 44/329 (13%) | $4.63 \times 10^{-5}$ | $3.39 \times 10^{-4}$ |
| Lisch nodules [HP:0009737] | 22/42 (52%) | 61/242 (25%) | $7.48 \times 10^{-4}$ | 0.003 |
| Inguinal freckling [HP:0030052] | 34/60 (57%) | 58/189 (31%) | $3.92 \times 10^{-4}$ | 0.002 |
| Optic nerve glioma [HP:0009734] | 15/33 (45%) | 24/143 (17%) | $8.57 \times 10^{-4}$ | 0.003 |

**(d)** Fisher Exact Test performed to compare HPO annotation frequency with respect to Leu847Pro and other. Total of 22 tests were performed.

| HPO term | SV | other | p-value | adj. p-value |
|---|---|---|---|---|
| Scoliosis [HP:0002650] | 18/57 (32%) | 38/308 (12%) | $9.71 \times 10^{-4}$ | 0.017 |

**(e)** Fisher Exact Test performed to compare HPO annotation frequency with respect to SV and other. Total of 17 tests were performed.

| Genotype (A) | Genotype (B) | total tests performed | significant results |
|---|---|---|---|
| Missense | other | 21 | 0 |

**(f)** Fisher Exact Test performed to compare HPO annotation frequency with respect to genotypes.

**Figure S42:** The cohort comprised 419 individuals (192 females, 169 males, 58 with unknown sex). A total of 36 HPO terms were used to annotate the cohort. Disease diagnosis: Neurofibromatosis, type 1 (OMIM:162200). A substantial body of literature exists about genotype-phenotype correlations. The correlations described in this notebook have been previously reported [41, 42, 43]. A total of 36 unique variant alleles were found in *NF1* (transcript: `NM_001042492.3`, protein id: `NP_001035957.1`).

### NIPBL

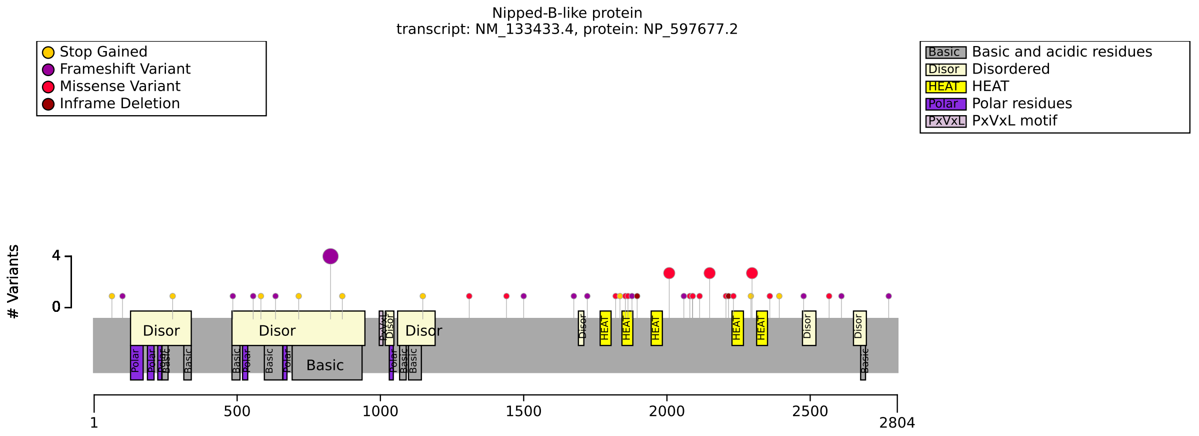


**(a)** Distribution of variants in NIPBL

| Genotype (A) | Genotype (B) | total tests performed | significant results |
|---|---|---|---|
| missense | other | 86 | 0 |
| disordered | other | 86 | 0 |
| FEMALE | MALE | 81 | 0 |

**(b)** Fisher Exact Test performed to compare HPO annotation frequency with respect to genotypes.

**Figure S43:** The cohort comprised 60 individuals (21 females, 31 males, 8 with unknown sex). A total of 90 HPO terms were used to annotate the cohort. Disease diagnosis: Cornelia de Lange syndrome 1 (OMIM:122470). No statistically significant genotype-phenotype correlation identified. A total of 50 unique variant alleles were found in *NIPBL* (transcript: `NM_133433.4`, protein id: `NP_597677.2`).

### NKX6-2

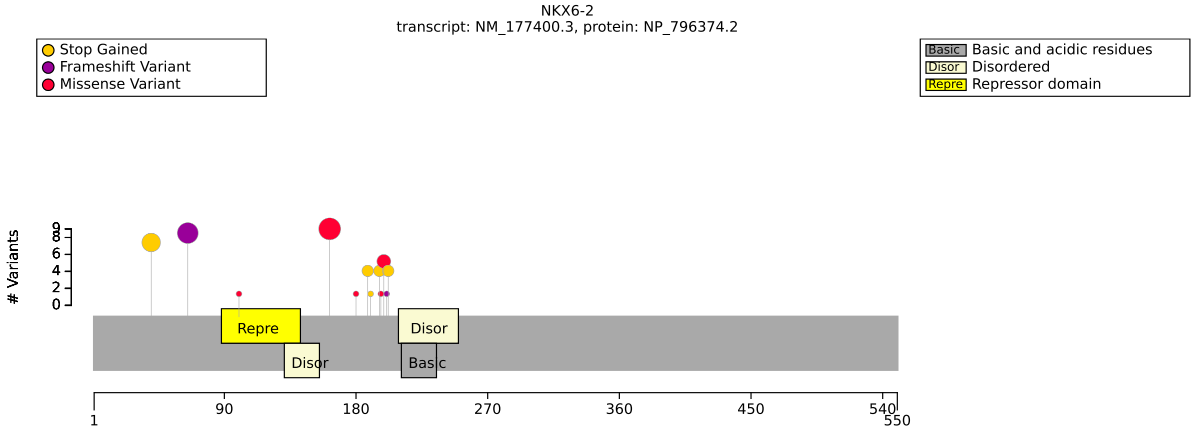


**(a)** Distribution of variants in NKX6-2

| Genotype (A) | Genotype (B) | total tests performed | significant results |
|---|---|---|---|
| missense/missense | other/other OR missense/other | 21 | 0 |
| L163V/L163V | L163V/other OR other/other | 21 | 0 |
| N term/N term OR N term/other | other/other | 21 | 0 |
| FEMALE | MALE | 21 | 0 |

**(b)** Fisher Exact Test performed to compare HPO annotation frequency with respect to genotypes.

**Figure S44:** The cohort comprised 33 individuals (15 females, 18 males). A total of 39 HPO terms were used to annotate the cohort. Disease diagnosis: Spastic ataxia 8, autosomal recessive, with hypomyelinating leukodystrophy (OMIM:617560). No statistically significant results identified. A total of 13 unique variant alleles were found in *NKX6-2* (transcript: `NM_177400.3`, protein id: `NP_796374.2`).

### PIGA

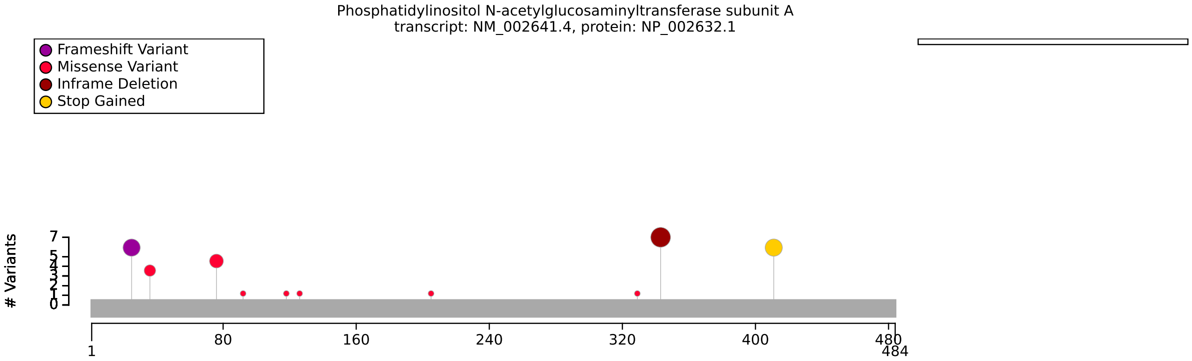


**(a)** Distribution of variants in PIGA

| Genotype (A) | Genotype (B) | total tests performed | significant results |
|---|---|---|---|
| missense | other | 23 | 0 |
| p.Leu344del | other | 20 | 0 |
| 1-100 | 100+ | 23 | 0 |

**(b)** Fisher Exact Test performed to compare HPO annotation frequency with respect to missense, p.Leu344del, and N-terminal (1-100) and other.

**Figure S45:** The cohort comprised 27 individuals (0 females, 27 males). A total of 188 HPO terms were used to annotate the cohort. Disease diagnoses: Multiple congenital anomalies-hypotonia-seizures syndrome 2 (OMIM:300868) (21 individuals), Neurodevelopmental disorder with epilepsy and hemochromatosis (OMIM:301072) (6 individuals). No statistically significant results identified. A total of 12 unique variant alleles were found in *PIGA* (transcript: `NM_002641.4`, protein id: `NP_002632.1`).

### POGZ

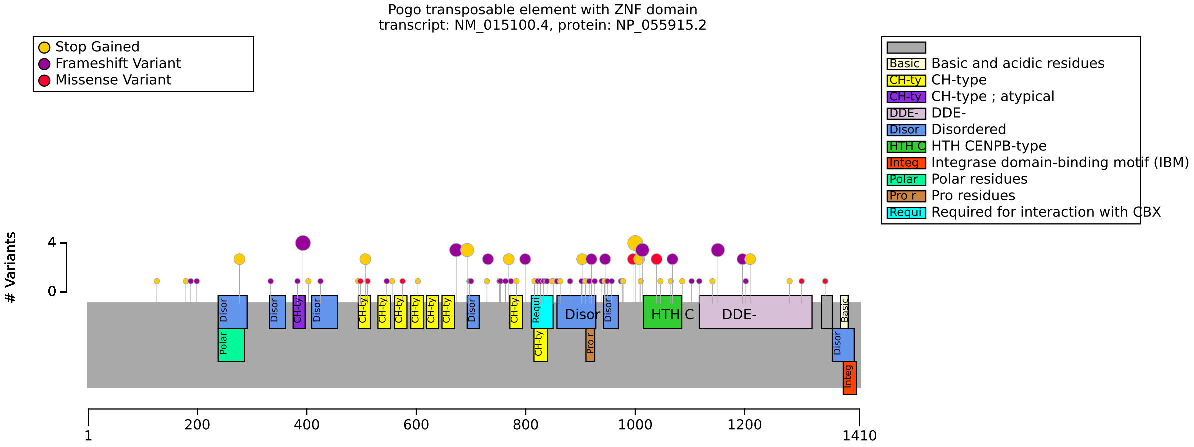


**(a)** Distribution of variants in POGZ

| Genotype (A) | Genotype (B) | total tests performed | significant results |
|---|---|---|---|
| Missense | Other | 24 | 0 |
| Frameshift | Other | 24 | 0 |

**(b)** Fisher Exact Test performed to compare HPO annotation frequency with respect to genotypes.

| Description | Variable | Genotype (A) | Genotype (B) | p-value | xrefs |
|---|---|---|---|---|---|
| A phenotypic severity score according to Nagy et al. 2022 | POGZ Severity Score | Missense | Other | 0.429 | - |

**(c)** POGZ Severity Score to compare Missense and Other with respect to POGZ Severity Score.

| Description | Variable | Genotype (A) | Genotype (B) | p-value | xrefs |
|---|---|---|---|---|---|
| A phenotypic severity score for individuals with intellectual disability | De Vries score | Missense | Other | 0.429 | - |

**(d)** De Vries Score to compare Missense and Other with respect to De Vries score.

**Figure S46:** The cohort comprised 117 individuals (54 females, 62 males, 1 with unknown sex). A total of 94 HPO terms were used to annotate the cohort. Disease diagnosis: White-Sutton syndrome (OMIM:616364). Negy et al. (2022) summarized data on 117 individuals with White-Sutton syndrome. They identified a correlation between a severity score and nonsense-mediated RNA decay (NMD). Missense variants were more often associated with mild phenotypes and truncating variants predicted to escape NMD presented with more severe phenotypes. Within this group, variants in the proline-rich region of the POGZ protein were associated with the most severe phenotypes). These authors did not apply multiple testing correction [44]. Our analysis did not identify a significant GPC. A total of 90 unique variant alleles were found in *POGZ* (transcript: `NM_015100.4`, protein id: `NP_055915.2`).

### PPP2R1A

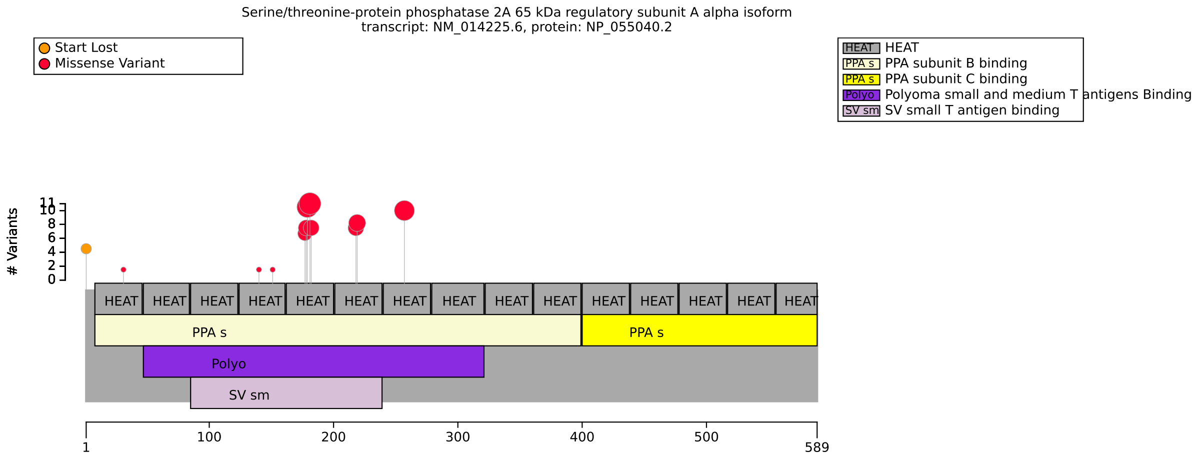


**(a)** Distribution of variants in PPP2R1A

| Genotype (A) | Genotype (B) | total tests performed | significant results |
|---|---|---|---|
| Absent in COSMIC | Present in COSMIC | 18 | 0 |
| Arg182Trp | other | 19 | 0 |
| SV40 small T antigen binding | other | 19 | 0 |

**(b)** Fisher Exact Test performed to compare HPO annotation frequency with respect to genotypes.

**Figure S47:** The cohort comprised 60 individuals (24 females, 32 males, 4 with unknown sex). A total of 52 HPO terms were used to annotate the cohort. Disease diagnosis: Houge-Janssen syndrome 2 (OMIM:616362). No statistically significant results identified. A total of 20 unique variant alleles were found in *PPP2R1A* (transcript: `NM_014225.6`, protein id: `NP_055040.2`).

### PTPN11

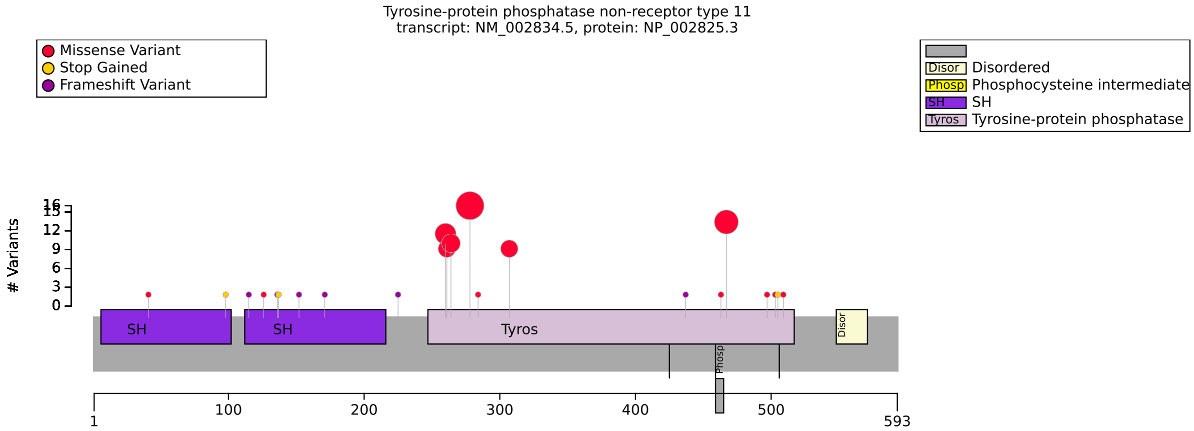


(a) Distribution of variants in PTPN11

| HPO term | Missense | Other | p-value | adj. p-value |
|---|---|---|---|---|
| Hypertelorism [HP:0000316] | 37/41 (90%) | 0/12 (0%) | $6.82 \times 10^{-9}$ | $2.73 \times 10^{-8}$ |
| Intellectual disability, mild [HP:0001256] | 8/23 (35%) | 0/12 (0%) | 0.032 | 0.032 |
| Pulmonic stenosis [HP:0001642] | 18/34 (53%) | 0/12 (0%) | 0.001 | 0.002 |
| Webbed neck [HP:0000465] | 15/20 (75%) | 0/12 (0%) | $2.94 \times 10^{-5}$ | $5.88 \times 10^{-5}$ |

**(b)** Fisher Exact Test performed to compare HPO annotation frequency with respect to Missense and Other. Total of 4 tests were performed.

| Genotype (A) | Genotype (B) | total tests performed | significant results |
|---|---|---|---|
| Tyr279Cys | Other | 8 | 0 |
| TK domain N term | TK domain C term | 5 | 0 |
| FEMALE | MALE | 6 | 0 |

**(c)** Fisher Exact Test performed to compare HPO annotation frequency with respect to genotypes.

**Figure S48:** The cohort comprised 70 individuals (27 females, 33 males, 10 with unknown sex). A total of 69 HPO terms were used to annotate the cohort. Disease diagnoses: LEOPARD syndrome 1 (OMIM:151100) (31 individuals), Noonan syndrome 1 (OMIM:163950) (27 individuals), Metachondromatosis (OMIM:156250) (12 individuals). No previous statistical analysis of correlations with PTPN11 missense variants was identified in the medical literature. A total of 27 unique variant alleles were found in *PTPN11* (transcript: `NM_002834.5`, protein id: `NP_002825.3`).

### RERE

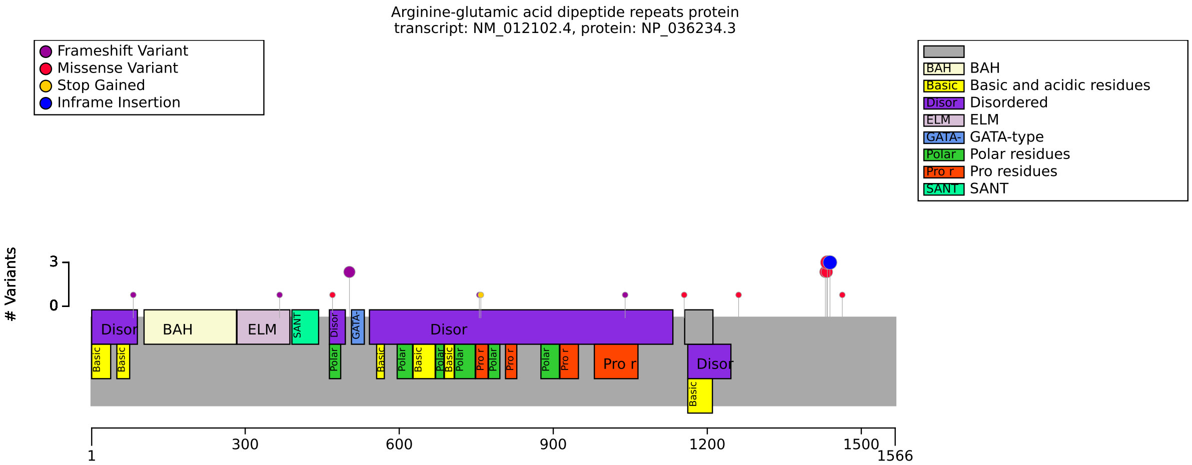


**(a)** Distribution of variants in RERE

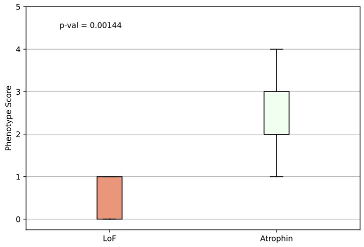


**(b)** Phenotype score adapted from Jordan et al. [45]. Variants in atrophin domain (residues 1425-1445) vs. others. Mann-Whitney U test.

| Genotype (A) | Genotype (B) | total tests performed | significant results |
|---|---|---|---|
| LoF | Atrophin | 56 | 0 |

**(c)** Fisher Exact Test performed to compare HPO annotation frequency with respect to genotypes.

| Description | Variable | Genotype (A) | Genotype (B) | p-value | xrefs |
|---|---|---|---|---|---|
| Phenotype score (see notebook) | HPO group count | LoF | Atrophin | 0.001 | [45] |

**(d)** HPO Group Count to compare LoF and Atrophin with respect to HPO group count (HP:0012443, HP:0012372,HP:0001627,HP:0012210, HP:0000407).

**Figure S49:** The cohort comprised 22 individuals (9 females, 13 males). A total of 115 HPO terms were used to annotate the cohort. Disease diagnosis: Neurodevelopmental disorder with or without anomalies of the brain, eye, or heart (OMIM:616975). Our results recapitulate the results of Jordan et al. [45] that The total number of structural defects and sensorineural hearing loss diagnoses seen in individuals with point mutations in the Atrophin-1 domain is significantly higher than expected based on the number of similar defects seen in individuals with putative loss-of-function variants. A total of 18 unique variant alleles were found in *RERE* (transcript: `NM_012102.4`, protein id: `NP_036234.3`).

#### RNU4-2

| Genotype (A) | Genotype (B) | total tests performed | significant results |
|---|---|---|---|
| n.64_65insT | other | 176 | 0 |
| insertion | other | 176 | 0 |
| FEMALE | MALE | 176 | 0 |

**(a)** Fisher Exact Test performed to compare HPO annotation frequency with respect to genotypes.

**Figure S50:** The cohort comprised 61 individuals (28 females, 33 males). A total of 209 HPO terms were used to annotate the cohort. Disease diagnosis: ReNU syndrome (OMIM:620851). No previous statistical analysis of correlations with RNU4-2 missense variants identified in the medical literature. A total of 7 unique variant alleles were found in *RNU4-2* (transcript: `NR_003137.3`, protein id: ).

### Robinow syndrome

| HPO term | OMIM:268310 | OMIM:616331 | p-value | adj. p-value |
|---|---|---|---|---|
| Mesomelia [HP:0003027] | 31/31 (100%) | 10/15 (67%) | 0.002 | 0.048 |
| Hearing impairment [HP:0000365] | 3/22 (14%) | 7/7 (100%) | $7.69 \times 10^{-5}$ | 0.003 |
| Short stature [HP:0004322] | 29/29 (100%) | 3/11 (27%) | $2.15 \times 10^{-6}$ | $1.89 \times 10^{-4}$ |
| Cleft palate [HP:0000175] | 0/17 (0%) | 5/8 (62%) | 0.001 | 0.031 |

**(a)** Fisher Exact Test performed to compare HPO annotation frequency with respect to Robinow syndrome, autosomal recessive (OMIM:268310) and Robinow syndrome, autosomal dominant 2 (OMIM:616331). Total of 88 tests were performed.

**Figure S51:** The cohort comprised 48 individuals (10 females, 14 males, 24 with unknown sex). A total of 103 HPO terms were used to annotate the cohort. Disease diagnoses: Robinow syndrome, autosomal recessive (OMIM:268310) (32 individuals), Robinow syndrome, autosomal dominant 2 (OMIM:616331) (16 individuals). Robinow syndrome is a skeletal dysplasia characterized by dysmorphic facial features, short-limbed dwarfism, vertebral segmentation, and genital hypoplasia. A total of 44 unique variant alleles were found.

### ROR2

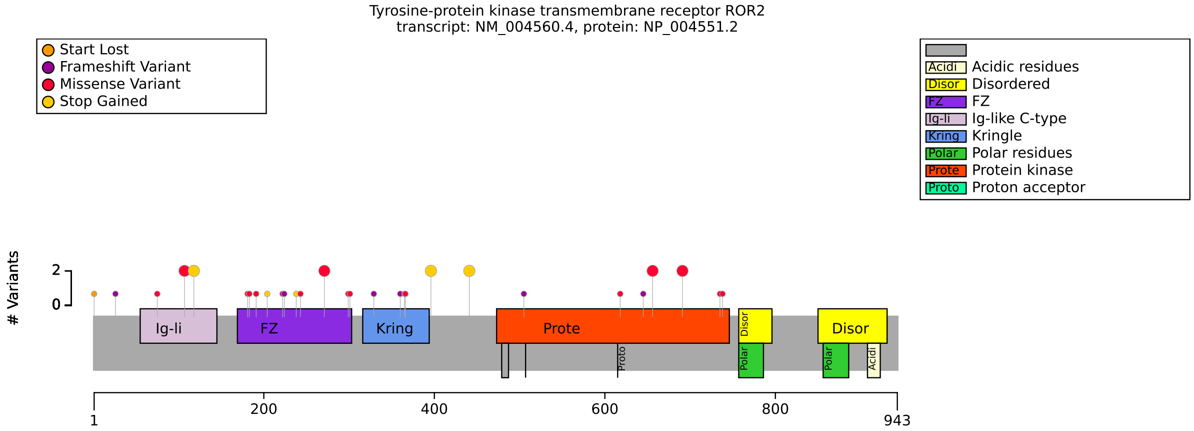


**(a)** Distribution of variants in ROR2

| Genotype (A) | Genotype (B) | total tests performed | significant results |
|---|---|---|---|
| missense/missense OR missense/other | other/other | 116 | 0 |
| FZ domain/FZ domain OR FZ domain/other | other/other | 116 | 0 |

**(b)** Fisher Exact Test performed to compare HPO annotation frequency with respect to genotypes.

**Figure S52:** The cohort comprised 32 individuals (3 females, 7 males, 22 with unknown sex). A total of 81 HPO terms were used to annotate the cohort. Disease diagnosis: Robinow syndrome, autosomal recessive (OMIM:268310). No statistically significant genotype-phenotype correlation was identified. A total of 32 unique variant alleles were found in *ROR2* (transcript: `NM_004560.4`, protein id: `NP_004551.2`).

### RPGRIP1

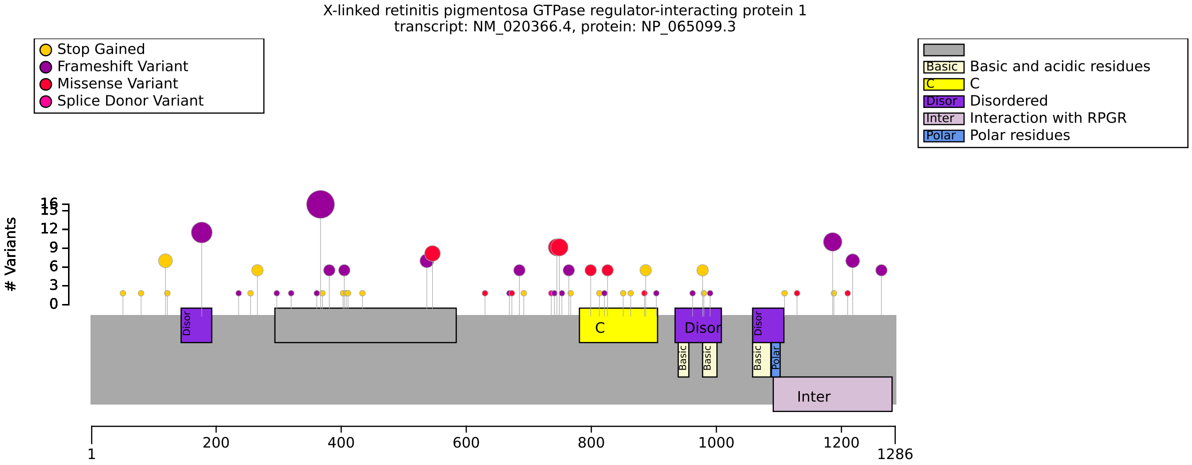


**(a)** Distribution of variants in RPGRIP1

| HPO term | 1107del/1107del OR 1107del/other | other/other | p-value | adj. p-value |
|---|---|---|---|---|
| Eye poking [HP:0001483] | 16/16 (100%) | 19/41 (46%) | $1.30 \times 10^{-4}$ | 0.002 |

**(b)** Fisher Exact Test performed to compare HPO annotation frequency with respect to 1107del/1107del OR 1107del/other and other/other. Total of 16 tests were performed.

| HPO term | OMIM:613826 | OMIM:608194 | p-value | adj. p-value |
|---|---|---|---|---|
| Nystagmus [HP:0000639] | 64/66 (97%) | 11/16 (69%) | 0.003 | 0.020 |
| Very low visual acuity [HP:0032122] | 35/39 (90%) | 4/16 (25%) | $5.21 \times 10^{-6}$ | $7.81 \times 10^{-5}$ |

**(c)** Fisher Exact Test performed to compare HPO annotation frequency with respect to OMIM:613826 and OMIM:608194. Total of 15 tests were performed.

| Genotype (A) | Genotype (B) | total tests performed | significant results |
|---|---|---|---|
| missense/missense | missense/other OR other/other | 17 | 0 |
| frameshift/frameshift | frameshift/other OR other/other | 16 | 0 |
| FEMALE | MALE | 16 | 0 |

**(d)** Fisher Exact Test performed to compare HPO annotation frequency with respect to genotypes.

**Figure S53:** The cohort comprised 100 individuals (45 females, 49 males, 6 with unknown sex). A total of 45 HPO terms were used to annotate the cohort. Disease diagnoses: Leber congenital amaurosis 6 (OMIM:613826) (79 individuals), Cone-rod dystrophy 13 (OMIM:608194) (21 individuals). It was reported that Patients with a double null genotype may develop symptoms earlier and have worse vision [46]. We did not observe a corresponding significant correlation in our dataset. A total of 71 unique variant alleles were found in *RPGRIP1* (transcript: `NM_020366.4`, protein id: `NP_065099.3`).

### SAMD9L

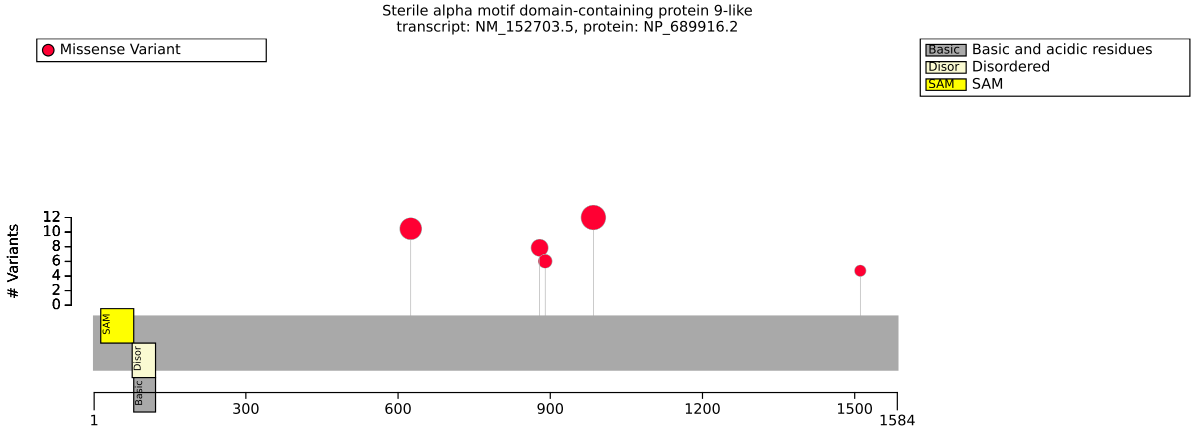


**(a)** Distribution of variants in SAMD9L

| HPO term | Arg986Cys | Ser626Leu | p-value | adj. p-value |
|---|---|---|---|---|
| Pancytopenia [HP:0001876] | 4/6 (67%) | 0/9 (0%) | 0.011 | 0.026 |
| Thrombocytopenia [HP:0001873] | 7/9 (78%) | 0/9 (0%) | 0.002 | 0.008 |
| Neutropenia [HP:0001875] | 7/9 (78%) | 0/9 (0%) | 0.002 | 0.008 |

**(b)** Fisher Exact Test performed to compare HPO annotation frequency with respect to Arg986Cys and Ser626Leu. Total of 7 tests were performed.

| Genotype (A) | Genotype (B) | total tests performed | significant results |
|---|---|---|---|
| N Term | other | 16 | 0 |

**(c)** Fisher Exact Test performed to compare HPO annotation frequency with respect to genotypes.

**Figure S54:** The cohort comprised 31 individuals (15 females, 16 males). 5 of these individuals were reported to be deceased. A total of 41 HPO terms were used to annotate the cohort. Disease diagnoses: Ataxia-pancytopenia syndrome (OMIM:159550) (22 individuals), Spinocerebellar ataxia 49 (OMIM:619806) (9 individuals). A recent summary of SAMD9L variants stated there was no evidence of phenotype–genotype correlation [47]. A total of 6 unique variant alleles were found in *SAMD9L* (transcript: `NM_152703.5`, protein id: `NP_689916.2`).

### SCN2A

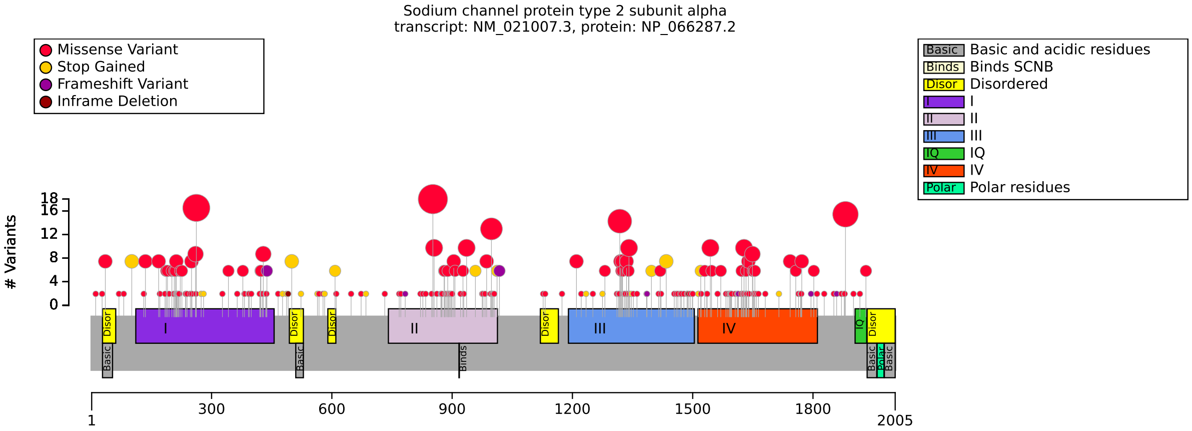


**(a)** Distribution of variants in SCN2A

| HPO term | Missense | Other | p-value | adj. p-value |
|---|---|---|---|---|
| Neurodevelopmental abnormality [HP:0012759] | 201/238 (84%) | 45/45 (100%) | 0.001 | 0.003 |
| Motor seizure [HP:0020219] | 146/175 (83%) | 6/31 (19%) | $4.51 \times 10^{-12}$ | $7.22 \times 10^{-11}$ |
| Seizure [HP:0001250] | 298/327 (91%) | 28/53 (53%) | $1.57 \times 10^{-10}$ | $8.38 \times 10^{-10}$ |
| Autism [HP:0000717] | 59/146 (40%) | 33/43 (77%) | $2.69 \times 10^{-5}$ | $8.61 \times 10^{-5}$ |
| Focal-onset seizure [HP:0007359] | 141/170 (83%) | 8/33 (24%) | $8.66 \times 10^{-11}$ | $6.93 \times 10^{-10}$ |
| Generalized-onset seizure [HP:0002197] | 104/133 (78%) | 6/31 (19%) | $1.43 \times 10^{-9}$ | $5.72 \times 10^{-9}$ |
| Intellectual disability [HP:0001249] | 144/198 (73%) | 34/34 (100%) | $9.39 \times 10^{-5}$ | $2.50 \times 10^{-4}$ |

**(b)** Fisher Exact Test performed to compare HPO annotation frequency with respect to Missense and Other. Total of 16 tests were performed.

| HPO term | I repeat | Other | p-value | adj. p-value |
|---|---|---|---|---|
| Neurodevelopmental abnormality [HP:0012759] | 48/65 (74%) | 198/218 (91%) | 0.001 | 0.009 |
| Intellectual disability [HP:0001249] | 21/42 (50%) | 157/190 (83%) | $2.59 \times 10^{-5}$ | $4.15 \times 10^{-4}$ |

**(c)** Fisher Exact Test performed to compare HPO annotation frequency with respect to I repeat and Other. Total of 16 tests were performed.

| Genotype (A) | Genotype (B) | total tests performed | significant results |
|---|---|---|---|
| Arg853Gln | Other | 16 | 0 |
| Exon 27 | Other | 16 | 0 |

**(d)** Fisher Exact Test performed to compare HPO annotation frequency with respect to genotypes.

**Figure S55:** The cohort comprised 393 individuals (0 females, 0 males, 393 with unknown sex). A total of 289 HPO terms were used to annotate the cohort. Disease diagnoses: Developmental and epileptic encephalopathy 11 (OMIM:613721) (342 individuals), Seizures, benign familial infantile, 3 (OMIM:607745) (51 individuals). Similar genotype-phenotype correlations have been previously published [48]. A total of 264 unique variant alleles were found in *SCN2A* (transcript: `NM_021007.3`, protein id: `NP_066287.2`).

### SCO2

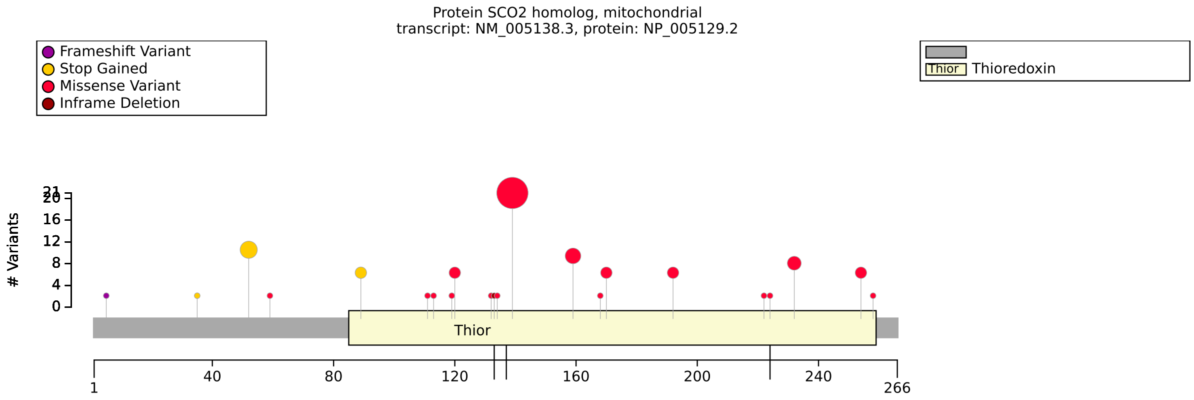


**(a)** Distribution of variants in SCO2

| HPO term | Glu140Lys/Glu140Lys | other/other OR Glu140Lys/other | p-value | adj. p-value |
|---|---|---|---|---|
| Hypertrophic cardiomyopathy [HP:0001639] | 2/6 (33%) | 13/13 (100%) | 0.004 | 0.027 |

**(b)** Fisher Exact Test performed to compare HPO annotation frequency with respect to Glu140Lys/Glu140Lys and other/other OR Glu140Lys/other. Total of 7 tests were performed.

| Description | Variable | Genotype (A) | Genotype (B) | p-value | xrefs |
|---|---|---|---|---|---|
| Survival analysis: Hypertrophic cardiomyopathy | Onset of HP:0001639 | Glu140Lys/Glu140Lys | other/other OR Glu140Lys/other | 0.219 | - |

**(c)** Onset of Hypertrophic cardiomyopathy to compare Glu140Lys/Glu140Lys and other/other OR Glu140Lys/other with respect to Onset of HP:0001639.

| Description | Variable | Genotype (A) | Genotype (B) | p-value | xrefs |
|---|---|---|---|---|---|
| Compute time until OMIM:604377 onset | Onset of OMIM:604377 | Glu140Lys/Glu140Lys | other/other OR Glu140Lys/other | 0.415 | - |

**(d)** Onset of OMIM:604377 to compare Glu140Lys/Glu140Lys and other/other OR Glu140Lys/other with respect to Onset of OMIM:604377.

**Figure S56:** The cohort comprised 37 individuals (10 females, 23 males, 4 with unknown sex). 24 of these individuals were reported to be deceased. A total of 107 HPO terms were used to annotate the cohort. Disease diagnoses: Mitochondrial complex IV deficiency, nuclear type 2 (OMIM:604377) (31 individuals), Myopia 6 (OMIM:608908) (6 individuals). The glu140lys correlation is driven by 5 patients found to be homozygous for the Glu140Lys variant [49]. The children were between the age of 8 months and 1 year and 8 months. Because of the relatively small cohort and the fact that other members of the cohort were not as young, caution is advised in the interpretation of this finding. A total of 25 unique variant alleles were found in *SCO2* (transcript: `NM_005138.3`, protein id: `NP_005129.2`).

### SEC61A1

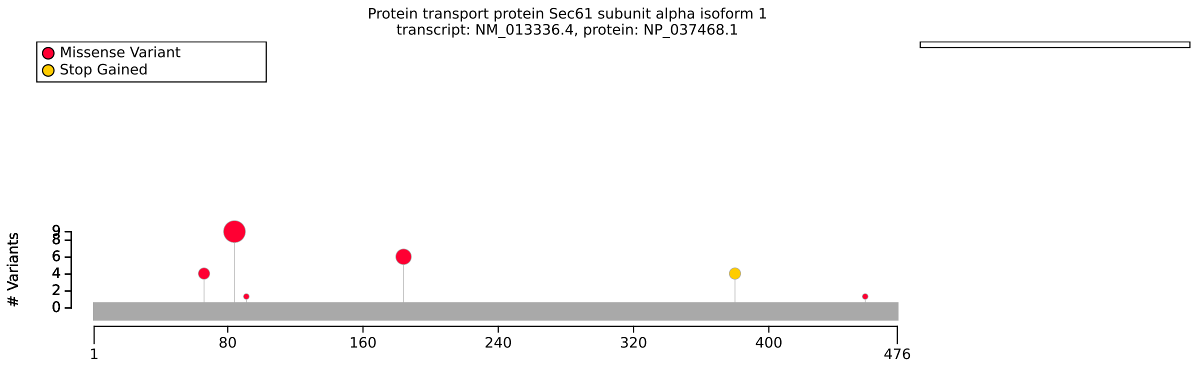


**(a)** Distribution of variants in SEC61A1

| Genotype (A) | Genotype (B) | total tests performed | significant results |
|---|---|---|---|
| p.Val85Asp | Other variant | 24 | 0 |
| FEMALE | MALE | 30 | 0 |

**(b)** Fisher Exact Test performed to compare HPO annotation frequency with respect to genotypes.

**Figure S57:** The cohort comprised 19 individuals (8 females, 11 males). 1 of these individuals were reported to be deceased. A total of 76 HPO terms were used to annotate the cohort. Disease diagnoses: Immunodeficiency, common variable, 15 (OMIM:620670) (11 individuals), Tubulointerstitial kidney disease, autosomal dominant, 5 (OMIM:617056) (7 individuals), Neutropenia, severe congenital, 11, autosomal dominant (OMIM:620674) (1 individuals). The origin of clinical diversity in patients with SEC61A1 mutation is currently unclear. With our present patient set, a particular phenotype cannot be predicted on the basis of location or nature of the mutation [50]. A total of 6 unique variant alleles were found in *SEC61A1* (transcript: `NM_013336.4`, protein id: `NP_037468.1`).

### SETD2

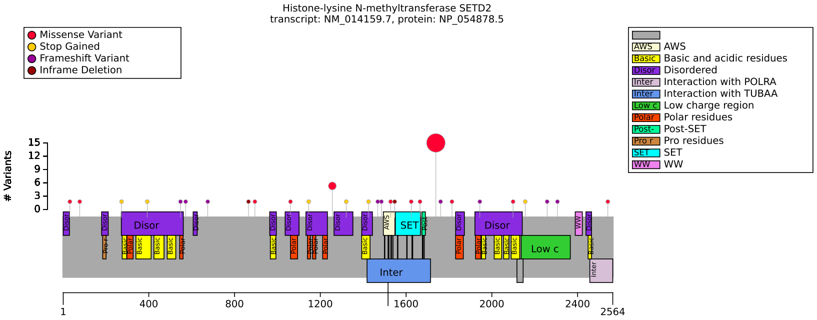


**(a)** Distribution of variants in SETD2

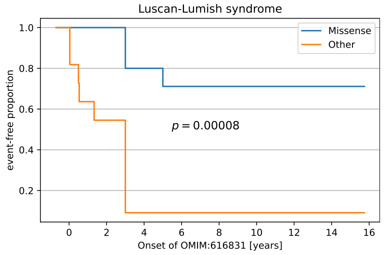


**(b)** Luscan-Lumish syndrome onset.

| HPO term | Arg1740Trp | Other | p-value | adj. p-value |
|---|---|---|---|---|
| Macrocephaly [HP:0000256] | 0/11 (0%) | 19/28 (68%) | $1.45 \times 10^{-4}$ | 0.002 |
| Hypertelorism [HP:0000316] | 11/11 (100%) | 5/23 (22%) | $1.53 \times 10^{-5}$ | $2.60 \times 10^{-4}$ |
| Wide nasal bridge [HP:0000431] | 9/9 (100%) | 2/9 (22%) | 0.002 | 0.012 |
| Ventriculomegaly [HP:0002119] | 4/4 (100%) | 2/17 (12%) | 0.003 | 0.012 |
| Severe global developmental delay [HP:0011344] | 9/9 (100%) | 0/12 (0%) | $3.40 \times 10^{-6}$ | $1.16 \times 10^{-4}$ |
| Delayed ability to walk [HP:0031936] | 8/8 (100%) | 1/10 (10%) | $4.11 \times 10^{-4}$ | 0.003 |
| Scoliosis [HP:0002650] | 6/6 (100%) | 2/14 (14%) | $7.22 \times 10^{-4}$ | 0.005 |

**(c)** Fisher Exact Test:HPO annotation frequency vs. Arg1740Trp / Other. Total of 34 tests were performed.

| HPO term | Missense | Other | p-value | adj. p-value |
|---|---|---|---|---|
| Macrocephaly [HP:0000256] | 4/24 (17%) | 15/15 (100%) | $1.54 \times 10^{-7}$ | $6.32 \times 10^{-6}$ |

**(d)** Fisher Exact Test: HPO annotation frequency vs. Missense / Other. Total of 41 tests were performed.

| Genotype (A) | Genotype (B) | total tests performed | significant results |
|---|---|---|---|
| FEMALE | MALE | 41 | 0 |

**(e)** Fisher Exact Test performed to compare HPO annotation frequency with respect to genotypes.

| Description | Variable | Genotype (A) | Genotype (B) | p-value | xrefs |
|---|---|---|---|---|---|
| Luscan-Lumish syndrome (OMIM:616831) onset | Onset of OMIM:616831 | Missense | Other | $8.47 \times 10^{-5}$ | - |

**(f)** Onset of OMIM:616831 to compare Missense and Other variant.

**Figure S58:** The cohort comprised 45 individuals (18 females, 27 males). 1 of these individuals was reported to be deceased. A total of 202 HPO terms were used to annotate the cohort. Disease diagnoses: Luscan-Lumish syndrome (OMIM:616831) (28 individuals), Rabin-Pappas syndrome (OMIM:620155) (14 individuals), Intellectual developmental disorder, autosomal dominant 70 (OMIM:620157) (3 individuals). Van Nieuwenhove et al. (2020) stated that the origin of clinical diversity in patients with SEC61A1 mutation is currently unclear [50]. Rabin et al. (2023) found that variants in codon 1740 of SETD2 whose features differ from those with LLS [51]. A total of 31 unique variant alleles were found in *SETD2* (transcript: `NM_014159.7`, protein id: `NP_054878.5`).

# SF3B4

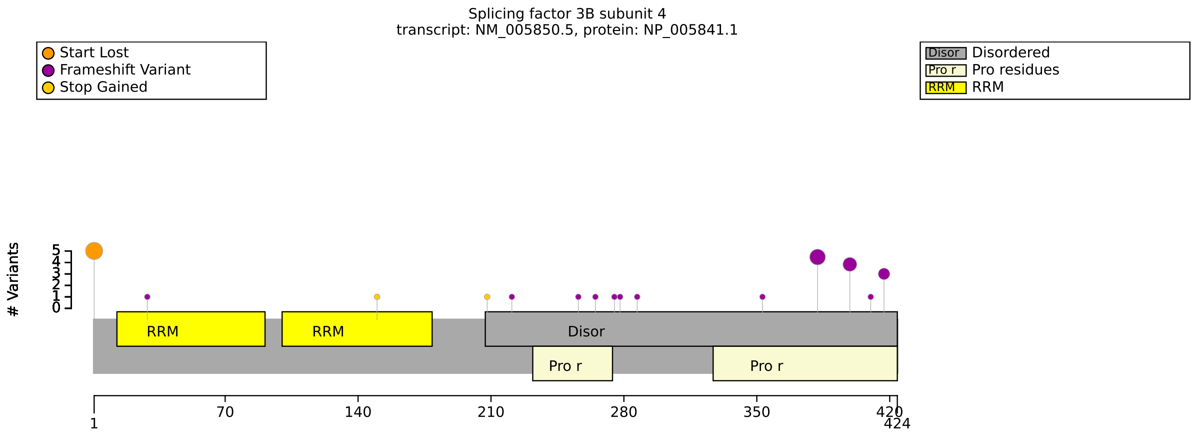


**(a)** Distribution of variants in SF3B4

| Genotype (A) | Genotype (B) | total tests performed | significant results |
|---|---|---|---|
| p.Met1? | Other | 46 | 0 |
| FEMALE | MALE | 46 | 0 |

**(b)** Fisher Exact Test performed to compare HPO annotation frequency with respect to genotypes.

**Figure S59:** The cohort comprised 26 individuals (18 females, 8 males). A total of 41 HPO terms were used to annotate the cohort. Disease diagnosis: Acrofacial dysostosis 1, Nager type (OMIM:154400). In one published analysis, it was stated that “although no significant genotype–phenotype association was found, it is notable that patients with frameshift SF3B4 variants and predicted to lead to nonsense-mediated RNA decay (NMD) of the transcripts tended to have a more severe clinical manifestation”. The authors did not correct for multiple testing and did not find nominally significant associations [52]. No significant correlation found in our study. A total of 18 unique variant alleles were found in *SF3B4* (transcript: `NM_005850.5`, protein id: `NP_005841.1`).

### SLC4A1

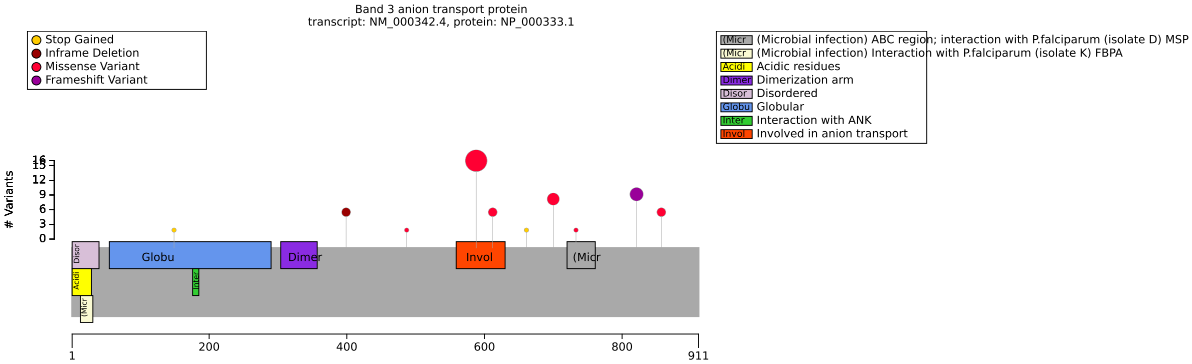


**(a)** Distribution of variants in SLC4A1

| Genotype (A) | Genotype (B) | total tests performed | significant results |
|---|---|---|---|
| r149w | other | 14 | 0 |

**(b)** Fisher Exact Test performed to compare HPO annotation frequency with respect to genotypes.

**Figure S60:** The cohort comprised 33 individuals (16 females, 16 males, 1 with unknown sex). A total of 49 HPO terms were used to annotate the cohort. Disease diagnoses: Distal renal tubular acidosis 1 (OMIM:179800) (18 individuals), Spherocytosis, type 4 (OMIM:612653) (7 individuals), Distal renal tubular acidosis 4 with hemolytic anemia (OMIM:611590) (7 individuals), Cryohydrocytosis (OMIM:185020) (1 individuals). No statistically significant results identified. A total of 11 unique variant alleles were found in *SLC4A1* (transcript: `NM_000342.4`, protein id: `NP_000333.1`).

### SLC32A1

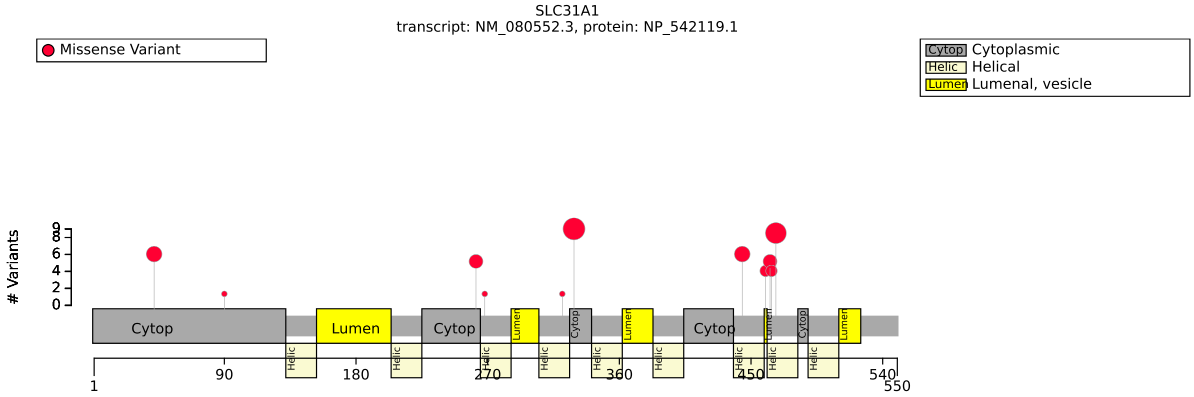


**(a)** Distribution of variants in SLC32A1

| Genotype (A) | Genotype (B) | total tests performed | significant results |
|---|---|---|---|
| N term | other | 11 | 0 |
| p.Met330Thr | Other variant | 11 | 0 |
| FEMALE | MALE | 11 | 0 |

**(b)** Fisher Exact Test performed to compare HPO annotation frequency with respect to genotypes.

**Figure S61:** The cohort comprised 38 individuals (19 females, 19 males). A total of 44 HPO terms were used to annotate the cohort. Disease diagnoses: Generalized epilepsy with febrile seizures plus, type 12 (OMIM:620755) (34 individuals), Developmental and epileptic encephalopathy 114 (OMIM:620774) (4 individuals). No significant correlation identified. A total of 12 unique variant alleles were found in *SLC32A1* (transcript: `NM_080552.3`, protein id: `NP_542119.1`).

### SLC45A2

| Genotype (A) | Genotype (B) | total tests performed | significant results |
|---|---|---|---|
| missense/missense OR missense/other | other/other | 24 | 0 |
| FEMALE | MALE | 25 | 0 |

**(a)** Fisher Exact Test performed to compare HPO annotation frequency with respect to genotypes.

**Figure S62:** The cohort comprised 30 individuals (17 females, 13 males). A total of 16 HPO terms were used to annotate the cohort. Disease diagnosis: Albinism, oculocutaneous, type IV (OMIM:606574). No significant association identified. A total of 28 unique variant alleles were found in *SLC45A2* (transcript: `NM_016180.5`, protein id: `NP_057264.4`).

### SMAD2

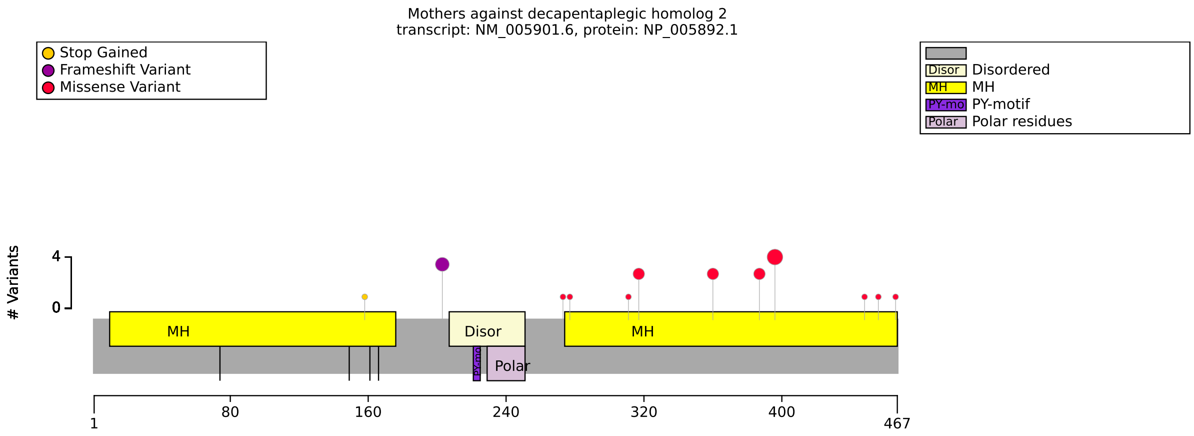


**(a)** Distribution of variants in SMAD2

| Genotype (A) | Genotype (B) | total tests performed | significant results |
|---|---|---|---|
| Ser397Tyr | Other | 26 | 0 |
| N Term | Other | 27 | 0 |
| FEMALE | MALE | 27 | 0 |

**(b)** Fisher Exact Test performed to compare HPO annotation frequency with respect to genotypes.

**Figure S63:** The cohort comprised 23 individuals (13 females, 8 males, 2 with unknown sex). A total of 89 HPO terms were used to annotate the cohort. Disease diagnoses: Loeys-Dietz syndrome 6 (OMIM:619656) (18 individuals), Congenital heart defects, multiple types, 8, with or without heterotaxy (OMIM:619657) (5 individuals). No significant correlations with specific SMAD2 residues or variant categories were identified. A total of 15 unique variant alleles were found in *SMAD2* (transcript: `NM_005901.6`, protein id: `NP_005892.1`).

### SMAD3

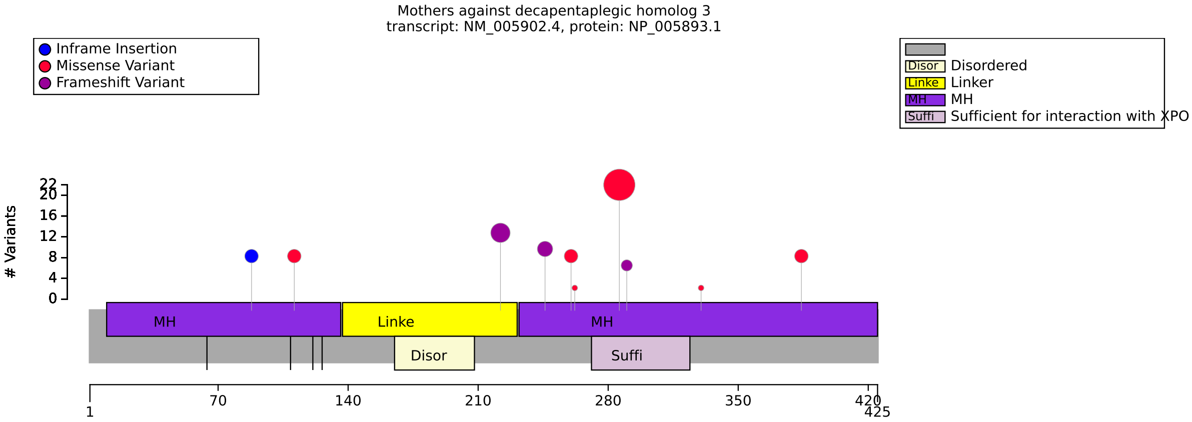


**(a)** Distribution of variants in SMAD3

| HPO term | p.Arg287Trp | Other | p-value | adj. p-value |
|---|---|---|---|---|
| Osteoarthritis [HP:0002758] | 19/19 (100%) | 7/19 (37%) | $3.72 \times 10^{-5}$ | $8.56 \times 10^{-4}$ |

**(b)** Fisher Exact Test performed to compare HPO annotation frequency with respect to p.Arg287Trp and Other. Total of 23 tests were performed.

| Genotype (A) | Genotype (B) | total tests performed | significant results |
|---|---|---|---|
| missense | other | 31 | 0 |
| FEMALE | MALE | 31 | 0 |

**(c)** Fisher Exact Test performed to compare HPO annotation frequency with respect to genotypes.

**Figure S64:** The cohort comprised 49 individuals (22 females, 27 males). A total of 30 HPO terms were used to annotate the cohort. Disease diagnosis: Loeys-Dietz syndrome 3 (OMIM:613795). There was no evidence of a correlation between missense variants and specific phenotypic abnormalities. In contrast, there was a statistically significant correlation with Arg287Trp and Osteoarthritis. The residue Arg287 is located in the Mad homology 2 (MH2) domain in a region that mediates interaction with exportin 4 [53]. Chesneau et al stated that there is an absence of correlation between the SMAD3 variant type and the occurrence of aortic phenotypes [54]. To the best of our knowledge an association between Arg287Trp and Osteoarthritis has not been previously noted. A total of 10 unique variant alleles were found in *SMAD3* (transcript: `NM_005902.4`, protein id: `NP_005893.1`).

### SMARCB1

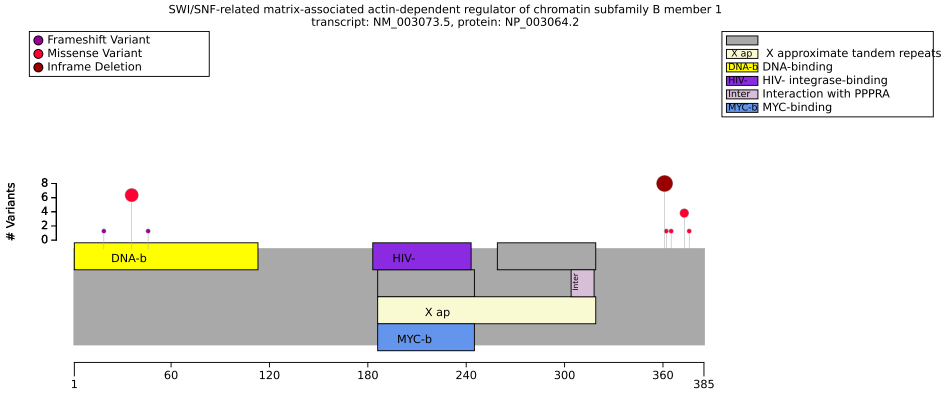


**(a)** Distribution of variants in SMARCB1

| HPO term | Structural variant | Other | p-value | adj. p-value |
|---|---|---|---|---|
| Atypical teratoid/rhabdoid tumor [HP:0034401] | 8/9 (89%) | 2/19 (11%) | $1.19 \times 10^{-4}$ | $5.94 \times 10^{-4}$ |
| Embryonal neoplasm [HP:0002898] | 8/8 (100%) | 2/19 (11%) | $2.03 \times 10^{-5}$ | $1.52 \times 10^{-4}$ |
| Neoplasm by histology [HP:0011792] | 11/11 (100%) | 4/21 (19%) | $1.06 \times 10^{-5}$ | $1.52 \times 10^{-4}$ |
| Rhabdoid tumor [HP:0034557] | 4/4 (100%) | 2/19 (11%) | 0.002 | 0.006 |
| Neoplasm by anatomical site [HP:0011793] | 3/3 (100%) | 3/20 (15%) | 0.011 | 0.028 |
| Neuroepithelial neoplasm [HP:0030063] | 2/2 (100%) | 0/17 (0%) | 0.006 | 0.018 |

**(b)** Fisher Exact Test. Total of 15 tests were performed.

| Genotype (A) | Genotype (B) | total tests performed | significant results |
|---|---|---|---|
| Lys364del | Other | 52 | 0 |
| DNA binding | Other | 51 | 0 |

**(c)** Fisher Exact Test performed to compare HPO annotation frequency with respect to genotypes.

**Figure S65:** The cohort comprised 32 individuals (12 females, 8 males, 12 with unknown sex). 9 of these individuals were reported to be deceased. A total of 110 HPO terms were used to annotate the cohort. Disease diagnoses: Coffin-Siris syndrome 3 (OMIM:614608) (18 individuals), Rhabdoid tumor predisposition syndrome 1 (OMIM:609322) (14 individuals). Our analysis of SMARCB1 included variants associated with Coffin-Siris syndrome 3 (OMIM:614608). A total of 11 unique variant alleles were found in *SMARCB1* (transcript: `NM_003073.5`, protein id: `NP_003064.2`). Presumably the result of embryonal neoplasm being signficantly associated with structural variants relates to a different distribution of variants in rhabdoid tumor predisposition syndrome-1 than in Coffin-Siris syndrome 3. A published analysis of GPCs in Coffin-Siris syndrome did not present a statistical analysis and did not note the current association [55].

### SMARCC2

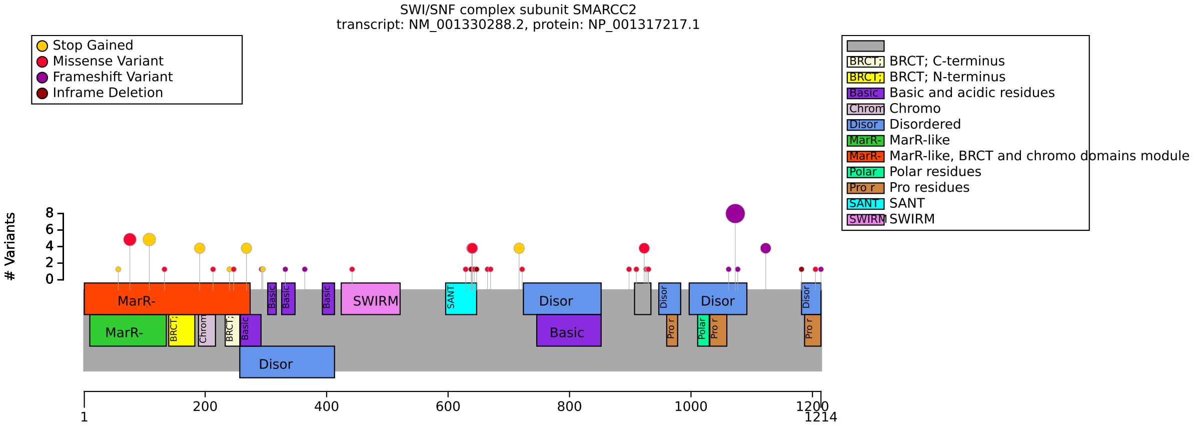


**(a)** Distribution of variants in SMARCC2

| HPO term | c.3222del | other | p-value | adj. p-value |
|---|---|---|---|---|
| Intellectual disability [HP:0001249] | 1/6 (17%) | 49/52 (94%) | $6.99 \times 10^{-5}$ | 0.006 |

**(b)** Fisher Exact Test performed to compare HPO annotation frequency with respect to c.3222del and other. Total of 83 tests were performed.

| Genotype (A) | Genotype (B) | total tests performed | significant results |
|---|---|---|---|
| Missense | Truncating | 72 | 0 |
| N term | Other | 83 | 0 |
| FEMALE | MALE | 83 | 0 |

**(c)** Fisher Exact Test performed to compare HPO annotation frequency with respect to genotypes.

**Figure S66:** The cohort comprised 65 individuals (24 females, 37 males, 4 with unknown sex). A total of 118 HPO terms were used to annotate the cohort. Disease diagnosis: Coffin-Siris syndrome 8 (OMIM:618362). Bosch et al. (2023) reported correlations for missense versus truncating variant cohorts for Global developmental delay; Intellectual disability (HP:0001263;HP:0001249): p = 0.005 (FDR-corr: 0.033); Muscular hypotonia (HP:0001252): p = 0.004 (FDR-corr: 0.033); mild GDD/ID (HP:0011342;HP:0001256): p = 0.012 (FDR-corr: 0.051); Abnormality of the outer ear (HP:0000356): p = 0.013 (FDR-corr: 0.051); Decreased body weight (HP:0004325): p = 0.002 (FDR-corr: 0.024); Abnormality of the eye (HP:0000478): p = 0.000 (FDR-corr: 0.008); Short stature (HP:0004322): p = 0.006 (FDR-corr: 0.035); Feeding difficulties/failure to thrive (HP:0011968;HP:0001508): p = 0.013 (FDR-corr: 0.051) [56]. The authors did not specifically test c.3222del. Differences in results may be due to different assumptions of the analysis procedure. Code to reproduce the results in Bosch et al. was not available. A total of 46 unique variant alleles were found in *SMARCC2* (transcript: `NM_001330288.2`, protein id: `NP_001317217.1`).

### SON

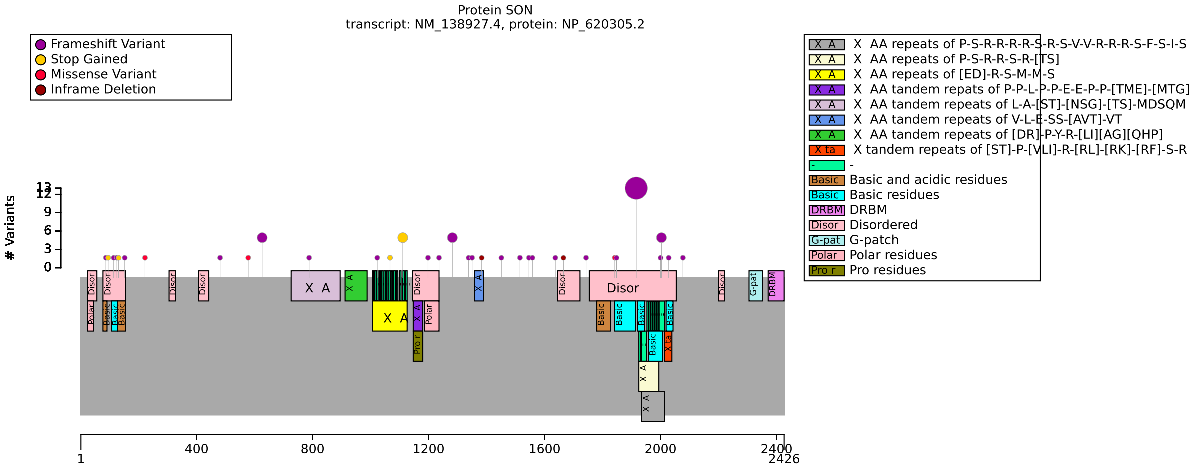


**(a)** Distribution of variants in SON

| Genotype (A) | Genotype (B) | total tests performed | significant results |
|---|---|---|---|
| missense | Other | 48 | 0 |
| c.5753_5756del | Other | 51 | 0 |
| FEMALE | MALE | 51 | 0 |

**(b)** Fisher Exact Test performed to compare HPO annotation frequency with respect to genotypes.

**Figure S67:** The cohort comprised 52 individuals (26 females, 26 males). A total of 47 HPO terms were used to annotate the cohort. Disease diagnosis: ZTTK SYNDROME (OMIM:617140). Dingemans et al (2020) suggested a different pathomechanism for missense variants, but our cohort only contains 3 individuals with missense variants, so there is no statistical power [2]. A total of 35 unique variant alleles were found in *SON* (transcript: `NM_138927.4`, protein id: `NP_620305.2`).

### SPTAN1 (Part 1/2)

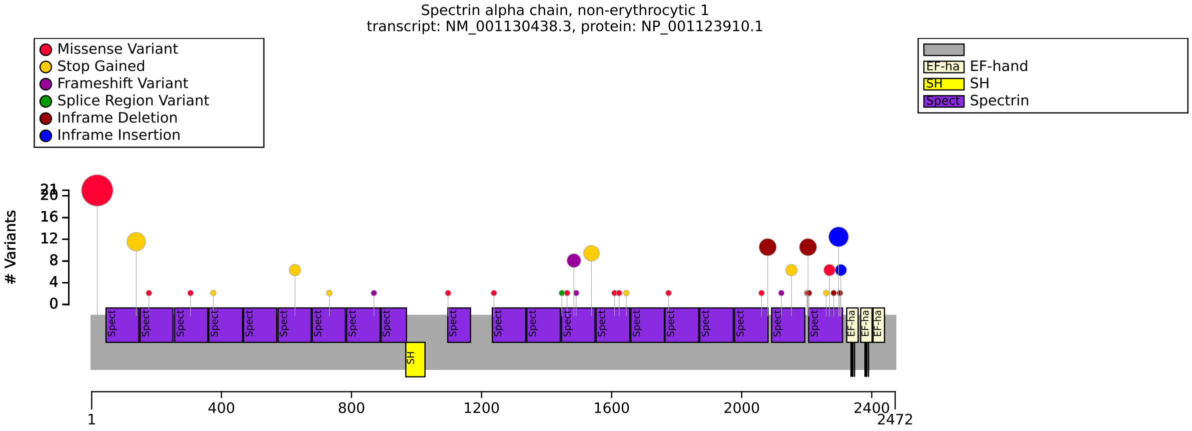


(**a**) Distribution of variants in SPTAN1

| HPO term | missense | other | p-value | adj. p-value |
|---|---|---|---|---|
| Spastic paraplegia [HP:0001258] | 21/25 (84%) | 0/16 (0%) | $4.70 \times 10^{-8}$ | $6.58 \times 10^{-7}$ |
| Lower limb spasticity [HP:0002061] | 21/22 (95%) | 0/12 (0%) | $2.37 \times 10^{-8}$ | $6.58 \times 10^{-7}$ |
| Appendicular spasticity [HP:0034353] | 21/22 (95%) | 2/14 (14%) | $8.72 \times 10^{-7}$ | $8.14 \times 10^{-6}$ |
| Spasticity [HP:0001257] | 22/23 (96%) | 4/16 (25%) | $5.22 \times 10^{-6}$ | $2.92 \times 10^{-5}$ |
| Motor axonal neuropathy [HP:0007002] | 0/19 (0%) | 12/16 (75%) | $2.18 \times 10^{-6}$ | $1.53 \times 10^{-5}$ |
| Peripheral axonal neuropathy [HP:0003477] | 4/23 (17%) | 12/16 (75%) | $6.69 \times 10^{-4}$ | 0.002 |
| Motor seizure [HP:0020219] | 7/27 (26%) | 13/17 (76%) | 0.002 | 0.004 |
| Seizure [HP:0001250] | 13/33 (39%) | 24/28 (86%) | $2.48 \times 10^{-4}$ | $9.92 \times 10^{-4}$ |
| Intellectual disability [HP:0001249] | 9/32 (28%) | 18/23 (78%) | $3.43 \times 10^{-4}$ | 0.001 |
| Lower limb muscle weakness [HP:0007340] | 15/16 (94%) | 16/28 (57%) | 0.015 | 0.030 |
| Distal lower limb muscle weakness [HP:0009053] | 9/19 (47%) | 0/16 (0%) | 0.001 | 0.004 |
| Infantile spasms [HP:0012469] | 2/29 (7%) | 13/31 (42%) | 0.002 | 0.005 |
| Epileptic spasm [HP:0011097] | 2/22 (9%) | 13/17 (76%) | $2.93 \times 10^{-5}$ | $1.37 \times 10^{-4}$ |
| Microcephaly [HP:0000252] | 4/34 (12%) | 15/32 (47%) | 0.002 | 0.005 |

(**b**) Fisher Exact Test performed to compare HPO annotation frequency with respect to missense and other. Total of 28 tests were performed.

| HPO term | truncating | other | p-value | adj. p-value |
|---|---|---|---|---|
| Spastic paraplegia [HP:0001258] | 0/8 (0%) | 21/33 (64%) | 0.001 | 0.005 |
| Lower limb spasticity [HP:0002061] | 0/8 (0%) | 21/26 (81%) | $7.09 \times 10^{-5}$ | $3.54 \times 10^{-4}$ |
| Appendicular spasticity [HP:0034353] | 0/8 (0%) | 23/28 (82%) | $4.25 \times 10^{-5}$ | $2.66 \times 10^{-4}$ |
| Spasticity [HP:0001257] | 0/8 (0%) | 26/31 (84%) | $2.09 \times 10^{-5}$ | $1.74 \times 10^{-4}$ |
| Hypotonia [HP:0001252] | 1/11 (9%) | 15/24 (62%) | 0.004 | 0.015 |
| Motor axonal neuropathy [HP:0007002] | 12/12 (100%) | 0/23 (0%) | $1.20 \times 10^{-9}$ | $3.00 \times 10^{-8}$ |
| Peripheral axonal neuropathy [HP:0003477] | 12/12 (100%) | 4/27 (15%) | $4.65 \times 10^{-7}$ | $5.82 \times 10^{-6}$ |

(**c**) Fisher Exact Test performed to compare HPO annotation frequency with respect to truncating and other. Total of 25 tests were performed.

**Figure S68:** See next page for caption

### SPTAN1 (Part 2/2)

| HPO term | Arg19Trp | Other | p-value | adj. p-value |
|---|---|---|---|---|
| Spastic paraplegia [HP:0001258] | 21/21 (100%) | 0/20 (0%) | $3.72 \times 10^{-12}$ | $9.29 \times 10^{-11}$ |
| Lower limb spasticity [HP:0002061] | 21/21 (100%) | 0/13 (0%) | $1.08 \times 10^{-9}$ | $1.35 \times 10^{-8}$ |
| Appendicular spasticity [HP:0034353] | 21/21 (100%) | 2/15 (13%) | $4.54 \times 10^{-8}$ | $1.89 \times 10^{-7}$ |
| Spasticity [HP:0001257] | 21/21 (100%) | 5/18 (28%) | $1.05 \times 10^{-6}$ | $3.30 \times 10^{-6}$ |
| Motor axonal neuropathy [HP:0007002] | 0/16 (0%) | 12/19 (63%) | $6.26 \times 10^{-5}$ | $1.74 \times 10^{-4}$ |
| Peripheral axonal neuropathy [HP:0003477] | 4/20 (20%) | 12/19 (63%) | 0.010 | 0.017 |
| Motor seizure [HP:0020219] | 0/19 (0%) | 20/25 (80%) | $3.34 \times 10^{-8}$ | $1.67 \times 10^{-7}$ |
| Seizure [HP:0001250] | 2/21 (10%) | 35/40 (88%) | $2.36 \times 10^{-9}$ | $1.47 \times 10^{-8}$ |
| Intellectual disability [HP:0001249] | 0/21 (0%) | 27/34 (79%) | $1.76 \times 10^{-9}$ | $1.47 \times 10^{-8}$ |
| Lower limb muscle weakness [HP:0007340] | 14/14 (100%) | 17/30 (57%) | 0.003 | 0.007 |
| Distal lower limb muscle weakness [HP:0009053] | 8/15 (53%) | 1/20 (5%) | 0.002 | 0.004 |
| Infantile spasms [HP:0012469] | 0/19 (0%) | 15/41 (37%) | 0.001 | 0.003 |
| Epileptic spasm [HP:0011097] | 0/19 (0%) | 15/20 (75%) | $7.71 \times 10^{-7}$ | $2.75 \times 10^{-6}$ |
| Microcephaly [HP:0000252] | 0/21 (0%) | 19/45 (42%) | $2.45 \times 10^{-4}$ | $6.14 \times 10^{-4}$ |

**(a)** Fisher Exact Test performed to compare HPO annotation frequency with respect to Arg19Trp and Other. Total of 25 tests were performed.

| Genotype (A) | Genotype (B) | total tests performed | significant results |
|---|---|---|---|
| Lys2083del | Other | 21 | 0 |
| FEMALE | MALE | 28 | 0 |

**(b)** Fisher Exact Test performed to compare HPO annotation frequency with respect to genotypes.

**Figure S69:** The cohort comprised 85 individuals (40 females, 45 males). 3 of these individuals were reported to be deceased. A total of 96 HPO terms were used to annotate the cohort. Disease diagnoses: Spastic paraplegia 91, autosomal dominant, with or without cerebellar ataxia (OMIM:620538) (28 individuals), Developmental and epileptic encephalopathy 5 (OMIM:613477) (22 individuals), Developmental delay with or without epilepsy (OMIM:620540) (21 individuals), Neuronopathy, distal hereditary motor, autosomal dominant 11 (OMIM:620528) (14 individuals). Several publications have identified genotype-phenotype correlations in SPTAN1 that are comparable to those identified here [57, 58, 59]. A total of 36 unique variant alleles were found in *SPTAN1* (transcript: `NM_001130438.3`, protein id: `NP_001123910.1`).

### STXBP1

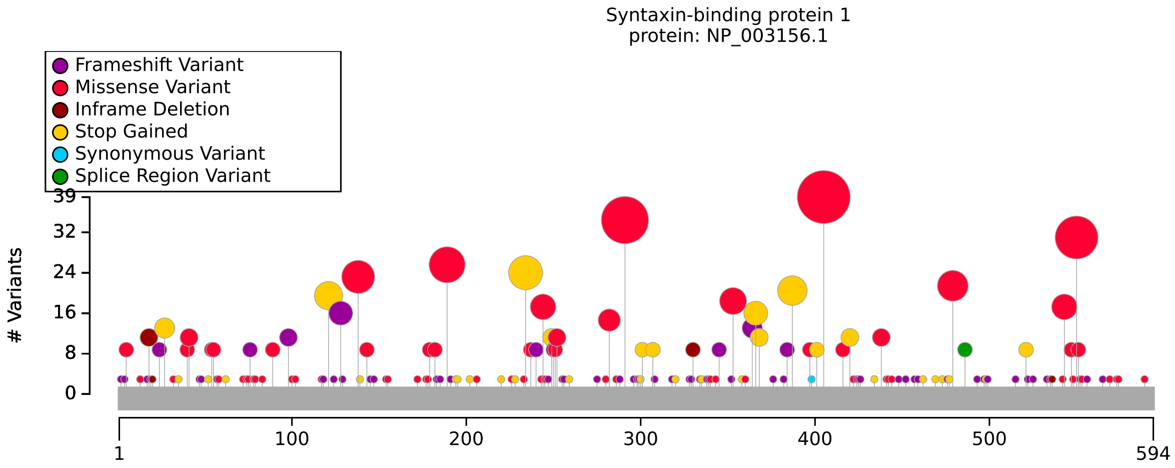


**(a)** Distribution of variants in STXBP1

| Genotype (A) | Genotype (B) | total tests performed | significant results |
|---|---|---|---|
| Missense | Other | 18 | 0 |
| Arg406 Variants | Other | 18 | 0 |
| Exon 14 | Other | 18 | 0 |

**(b)** Fisher Exact Test performed to compare HPO annotation frequency with respect to genotypes.

**Figure S70:** The cohort comprised 462 individuals (206 females, 220 males, 36 with unknown sex). A total of 516 HPO terms were used to annotate the cohort. Disease diagnosis: Developmental and epileptic encephalopathy 4 (OMIM:612164). No significant genotype-phenotype correlations identified. A total of 259 unique variant alleles were found in *STXBP1* (transcript: `NM_003165.6`, protein id: `NP_003156.1`).

### SUOX

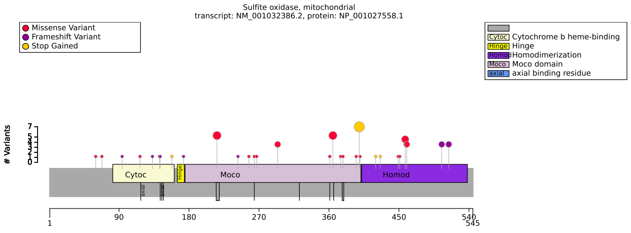


**(a)** Distribution of variants in SUOX

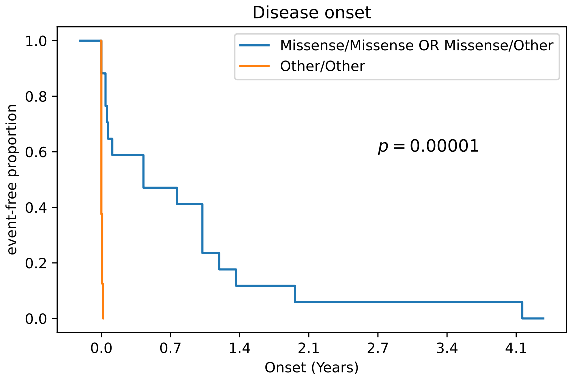


**(b)** Survival analysis: SUOX missense variants and disease onset

| HPO term | homodimerization/homodimerization OR homodimerization/Other | Other/Other | p-value | adj. p-value |
|---|---|---|---|---|
| Microcephaly [HP:0000252] | 0/9 (0%) | 10/12 (83%) | $2.21 \times 10^{-4}$ | 0.003 |

**(c)** Fisher Exact Test performed to compare HPO annotation frequency with respect to homodimerization/homodimerization OR homodimerization/Other and Other/Other. Total of 15 tests were performed.

| Genotype (A) | Genotype (B) | total tests performed | significant results |
|---|---|---|---|
| Missense/Missense OR Missense/Other | Other/Other | 15 | 0 |
| Moco domain/Moco domain OR Moco domain/Other | Other/Other | 15 | 0 |

**(d)** Fisher Exact Test performed to compare HPO annotation frequency with respect to genotypes.

| Description | Variable | Genotype (A) | Genotype (B) | p-value | xrefs |
|---|---|---|---|---|---|
| Onset of Sulfite oxidase deficiency (OMIM:272300) | Onset of OMIM:272300 | Missense/Missense OR Missense/Other | Other/Other | $9.18 \times 10^{-6}$ | - |

**(e)** Onset of OMIM:272300 to compare Missense/Missense OR Missense/Other and Other/Other with respect to Onset of OMIM:272300.

| Description | Variable | Genotype (A) | Genotype (B) | p-value | xrefs |
|---|---|---|---|---|---|
| Onset of Sulfite oxidase deficiency (OMIM:272300) | Onset of OMIM:272300 | homodimerization/homodimerization OR homodimerization/Other | Other/Other | 0.853 | [60] |

**(f)** Onset of OMIM:272300 to compare homodimerization/homodimerization OR homodimerization/Other and Other/Other with respect to Onset of OMIM:272300.

**Figure S71:** The cohort comprised 35 individuals (11 females, 14 males, 10 with unknown sex). 8 of these individuals were reported to be deceased. A total of 20 HPO terms were used to annotate the cohort. Disease diagnosis: Sulfite oxidase deficiency (OMIM:272300). A published Genotype-phenotype analysis demonstrated patients with biallelic missense mutations had milder symptoms (P = 0.023), later age of onset (P < 0.001), and a higher incidence of regression (P = 0.017) than other genotypes [60]. A total of 32 unique variant alleles were found in *SUOX* (transcript: `NM_001032386.2`, protein id: `NP_001027558.1`).

### TBCK

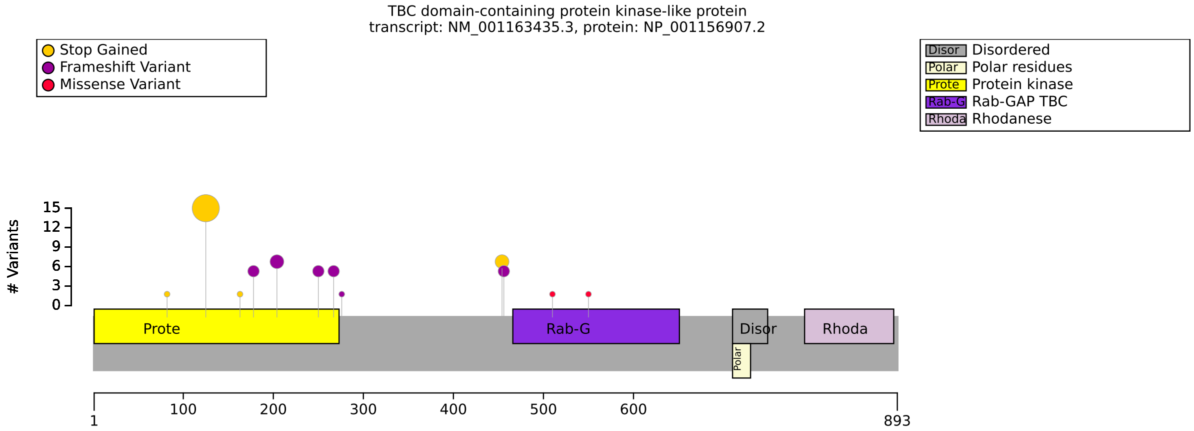


(**a**) Distribution of variants in TBCK

| HPO term | R126*/R126* | R126*/other OR other/other | p-value | adj. p-value |
|---|---|---|---|---|
| Macroglossia [HP:0000158] | 11/12 (92%) | 3/22 (14%) | $1.34 \times 10^{-5}$ | $4.30 \times 10^{-4}$ |
| Developmental regression [HP:0002376] | 9/12 (75%) | 2/22 (9%) | $1.83 \times 10^{-4}$ | 0.003 |

(**b**) Fisher Exact Test performed to compare HPO annotation frequency with respect to R126*/R126* and R126*/other OR other/other. Total of 32 tests were performed.

| Genotype (A) | Genotype (B) | total tests performed | significant results |
|---|---|---|---|
| missense/missense OR missense/other | other/other | 30 | 0 |
| FEMALE | MALE | 32 | 0 |

(**c**) Fisher Exact Test performed to compare HPO annotation frequency with respect to genotypes.

**Figure S72:** The cohort comprised 41 individuals (17 females, 24 males). 3 of these individuals were reported to be deceased. A total of 97 HPO terms were used to annotate the cohort. Disease diagnosis: Hypotonia, infantile, with psychomotor retardation and characteristic facies 3 (OMIM:616900). Durham et al (2023) stated that several studies have touched on a genotype-phenotype correlation of TBCK syndrome; however, more data are required for statistically significant conclusions [61]. A total of 18 unique variant alleles were found in *TBCK* (transcript: `NM_001163435.3`, protein id: `NP_001156907.2`).

### TBX1

| HPO term | Tyr418PhefsTer42 | Other | p-value | adj. p-value |
|---|---|---|---|---|
| Global developmental delay [HP:0001263] | 5/5 (100%) | 3/20 (15%) | 0.001 | 0.023 |
| Narrow nose [HP:0000460] | 5/5 (100%) | 0/6 (0%) | 0.002 | 0.024 |

**(a)** Fisher Exact Test performed to compare HPO annotation frequency with respect to Tyr418PhefsTer42 and Other. Total of 22 tests were performed.

| Genotype (A) | Genotype (B) | total tests performed | significant results |
|---|---|---|---|
| Missense | Other | 21 | 0 |

**(b)** Fisher Exact Test performed to compare HPO annotation frequency with respect to genotypes.

**Figure S73:** The cohort comprised 26 individuals (10 females, 15 males, 1 with unknown sex). A total of 25 HPO terms were used to annotate the cohort. Disease diagnosis: DiGeorge syndrome (OMIM:188400). A total of 12 unique variant alleles were found in *TBX1* (transcript: `NM_001379200.1`, protein id: `NP_001366129.1`).

### TBX5

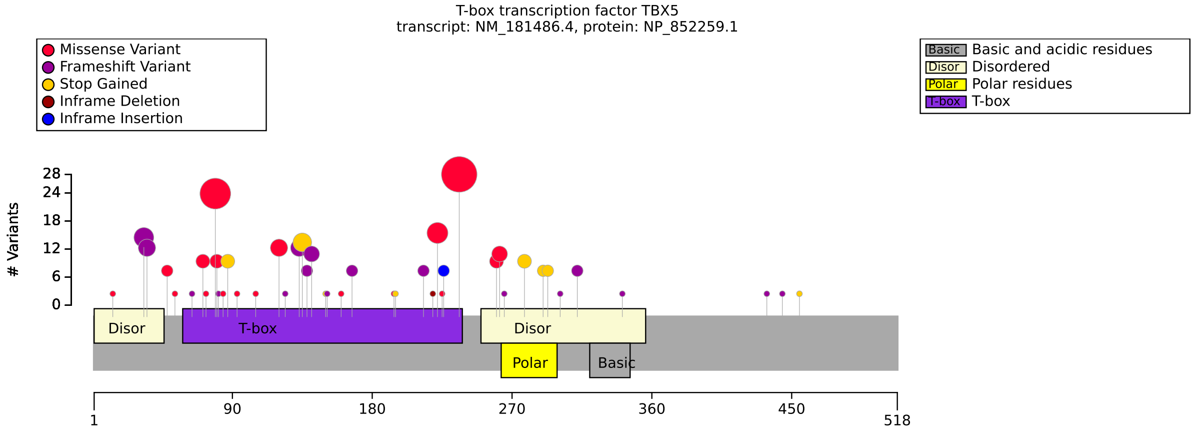


**(a)** Distribution of variants in TBX5

| HPO term | missense | other | p-value | adj. p-value |
|---|---|---|---|---|
| Ventricular septal defect [HP:0001629] | 31/60 (52%) | 30/30 (100%) | $4.63 \times 10^{-7}$ | $1.48 \times 10^{-5}$ |

**(b)** Fisher Exact Test performed to compare HPO annotation frequency with respect to missense and other. Total of 32 tests were performed.

| HPO term | Arg237Gln | other | p-value | adj. p-value |
|---|---|---|---|---|
| Ventricular septal defect [HP:0001629] | 0/17 (0%) | 61/73 (84%) | $5.55 \times 10^{-11}$ | $1.78 \times 10^{-9}$ |
| Upper limb phocomelia [HP:0009813] | 7/22 (32%) | 3/131 (2%) | $4.55 \times 10^{-5}$ | $7.28 \times 10^{-4}$ |

**(c)** Fisher Exact Test performed to compare HPO annotation frequency with respect to Arg237Gln and other. Total of 32 tests were performed.

| Genotype (A) | Genotype (B) | total tests performed | significant results |
|---|---|---|---|
| Gly80Arg | other | 28 | 0 |
| FEMALE | MALE | 13 | 0 |

**(d)** Fisher Exact Test performed to compare HPO annotation frequency with respect to genotypes.

**Figure S74:** The cohort comprised 156 individuals (56 females, 46 males, 54 with unknown sex). A total of 90 HPO terms were used to annotate the cohort. Disease diagnosis: Holt-Oram syndrome (OMIM:142900). Vanlerberghe et al. (2019) observed that isolated septal CHD are more common in the truncating than in the missense variants (p=0.02) [62]. A total of 53 unique variant alleles were found in *TBX5* (transcript: `NM_181486.4`, protein id: `NP_852259.1`).

### TGFB2

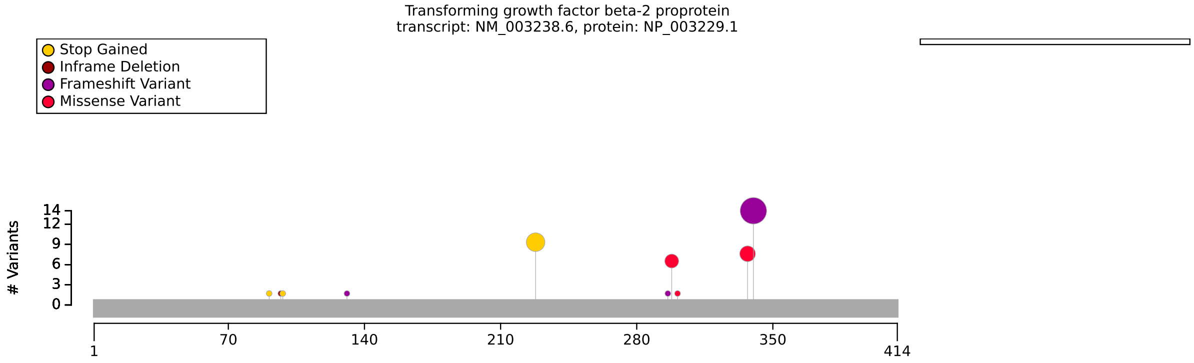


**(a)** Distribution of variants in TGFB2

| Genotype (A) | Genotype (B) | total tests performed | significant results |
|---|---|---|---|
| missense | other | 37 | 0 |
| FEMALE | MALE | 38 | 0 |

**(b)** Fisher Exact Test performed to compare HPO annotation frequency with respect to genotypes.

**Figure S75:** The cohort comprised 36 individuals (10 females, 26 males). 1 of these individuals were reported to be deceased. A total of 68 HPO terms were used to annotate the cohort. Disease diagnosis: Loeys-Dietz syndrome 4 (OMIM:614816). No significant association found. A total of 12 unique variant alleles were found in *TGFB2* (transcript: `NM_003238.6`, protein id: `NP_003229.1`).

### TGFB3

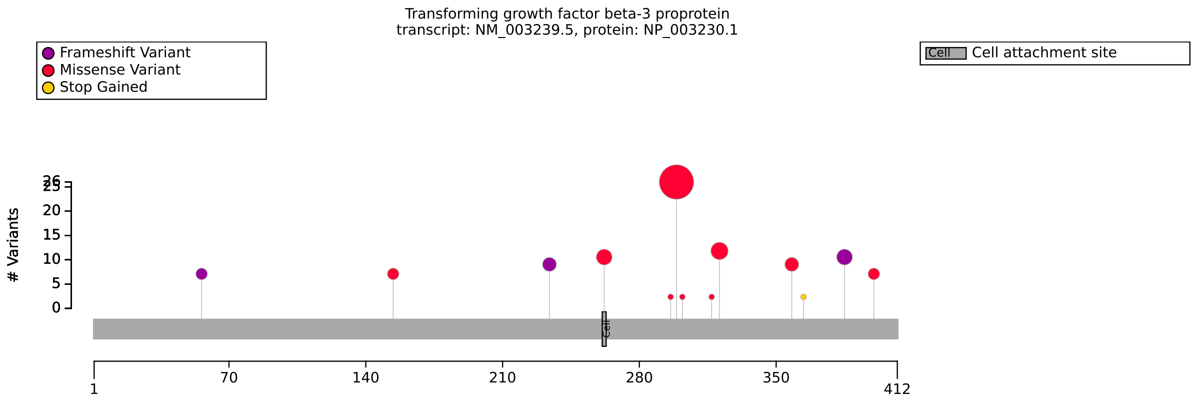


**(a)** Distribution of variants in TGFB3

| Genotype (A) | Genotype (B) | total tests performed | significant results |
|---|---|---|---|
| Missense | Other | 57 | 0 |
| Asp263His | Other | 18 | 0 |
| FEMALE | MALE | 57 | 0 |

**(b)** Fisher Exact Test performed to compare HPO annotation frequency with respect to genotypes.

**Figure S76:** The cohort comprised 75 individuals (34 females, 41 males). 7 of these individuals were reported to be deceased. A total of 73 HPO terms were used to annotate the cohort. Disease diagnosis: Loeys-Dietz syndrome 5 (OMIM:615582). No significant correlations identified. A total of 18 unique variant alleles were found in *TGFB3* (transcript: `NM_003239.5`, protein id: `NP_003230.1`).

### TGFBR1

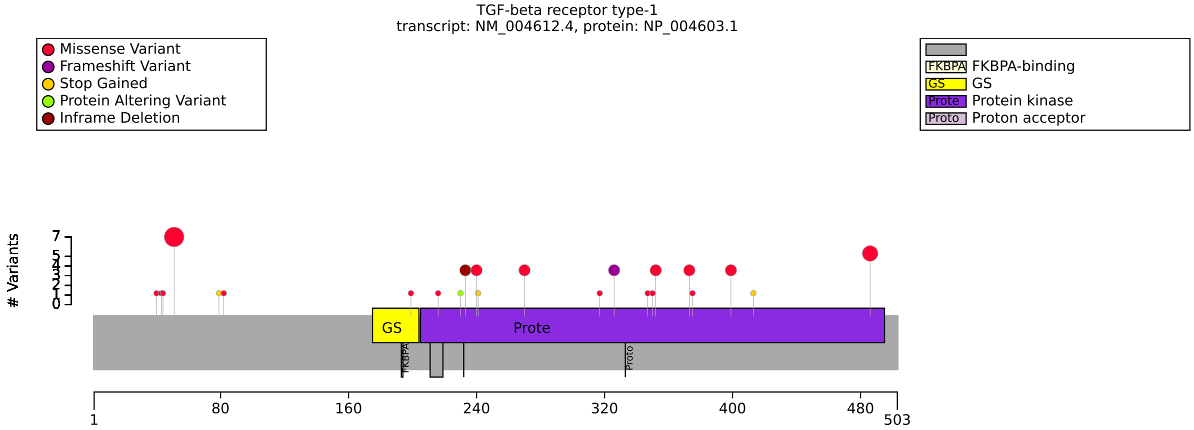


**(a)** Distribution of variants in TGFBR1

| HPO term | MSSE var | other | p-value | adj. p-value |
|---|---|---|---|---|
| Self-healing squamous epithelioma [HP:0034720] | 18/19 (95%) | 0/21 (0%) | $1.68 \times 10^{-10}$ | $2.68 \times 10^{-9}$ |
| Hypertelorism [HP:0000316] | 0/18 (0%) | 15/19 (79%) | $5.01 \times 10^{-7}$ | $4.01 \times 10^{-6}$ |
| Arterial tortuosity [HP:0005116] | 1/19 (5%) | 8/16 (50%) | 0.005 | 0.026 |

**(b)** Fisher Exact Test performed to compare HPO annotation frequency with respect to MSSE var and other. Total of 16 tests were performed.

| HPO term | Gly52Arg | Other | p-value | adj. p-value |
|---|---|---|---|---|
| Self-healing squamous epithelioma [HP:0034720] | 7/7 (100%) | 11/33 (33%) | 0.002 | 0.012 |

**(c)** Fisher Exact Test performed to compare HPO annotation frequency with respect to Gly52Arg and Other. Total of 7 tests were performed.

| Genotype (A) | Genotype (B) | total tests performed | significant results |
|---|---|---|---|
| FEMALE | MALE | 21 | 0 |

**(d)** Fisher Exact Test performed to compare HPO annotation frequency with respect to genotypes.

**Figure S77:** The cohort comprised 41 individuals (7 females, 16 males, 18 with unknown sex). 2 of these individuals were reported to be deceased. A total of 78 HPO terms were used to annotate the cohort. Disease diagnoses: Loeys-Dietz syndrome 1 (OMIM:609192) (23 individuals), Multiple self-healing squamous epithelioma, susceptibility to (OMIM:132800) (18 individuals). A total of 27 unique variant alleles were found in *TGFBR1* (transcript: `NM_004612.4`, protein id: `NP_004603.1`). Goudie et al. (2011) reported that the nature of the sequence variants, which include mutations in the extracellular ligand-binding domain and a series of truncating mutations in the kinase domain, indicates a clear genotype-phenotype correlation between loss-of-function TGFBR1 mutations and MSSE [63].

### TGFBR2

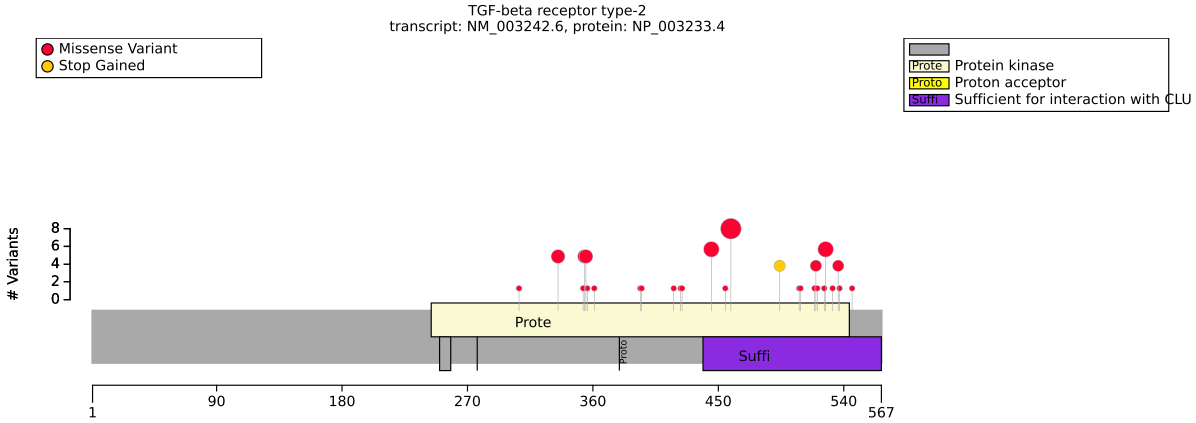


**(a)** Distribution of variants in TGFBR2

| Genotype (A) | Genotype (B) | total tests performed | significant results |
|---|---|---|---|
| CLU interaction region | Other | 42 | 0 |
| Missense | Other | 42 | 0 |
| FEMALE | MALE | 42 | 0 |

**(b)** Fisher Exact Test performed to compare HPO annotation frequency with respect to genotypes.

**Figure S78:** The cohort comprised 53 individuals (23 females, 30 males). 4 of these individuals were reported to be deceased. A total of 96 HPO terms were used to annotate the cohort. Disease diagnosis: Loeys-Dietz syndrome 2 (OMIM:610168). No statistically significant results identified. A total of 34 unique variant alleles were found in *TGFBR2* (transcript: `NM_003242.6`, protein id: `NP_003233.4`).

### TRAF7

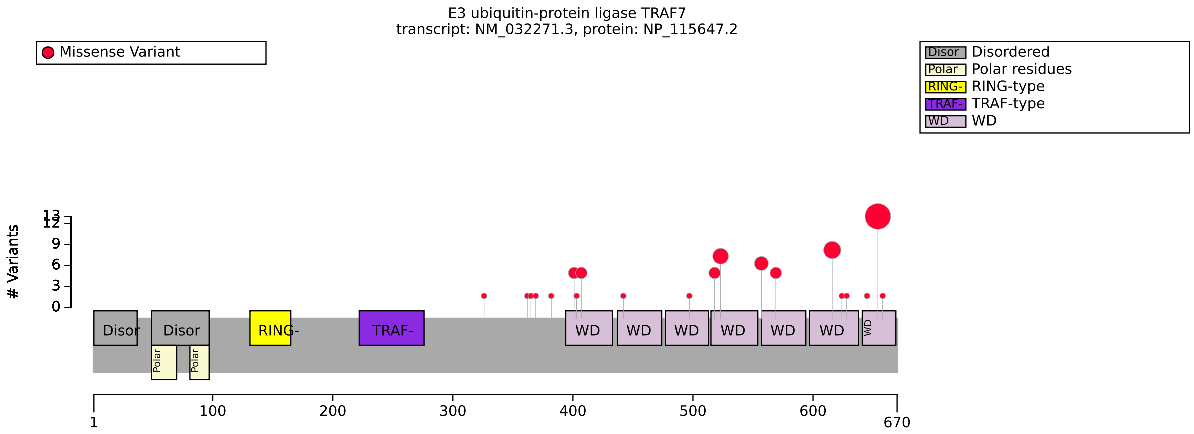


**(a)** Distribution of variants in TRAF7

| Genotype (A) | Genotype (B) | total tests performed | significant results |
|---|---|---|---|
| WD7 | other | 40 | 0 |
| Arg655Gln | Other variant | 40 | 0 |
| FEMALE | MALE | 40 | 0 |

**(b)** Fisher Exact Test performed to compare HPO annotation frequency with respect to genotypes.

**Figure S79:** The cohort comprised 45 individuals (17 females, 28 males). A total of 360 HPO terms were used to annotate the cohort. Disease diagnosis: Cardiac, facial, and digital anomalies with developmental delay (OMIM:618164). No significant correlations identified. A total of 23 unique variant alleles were found in *TRAF7* (transcript: NM_032271.3, protein id: NP_115647.2).

# U2AF2

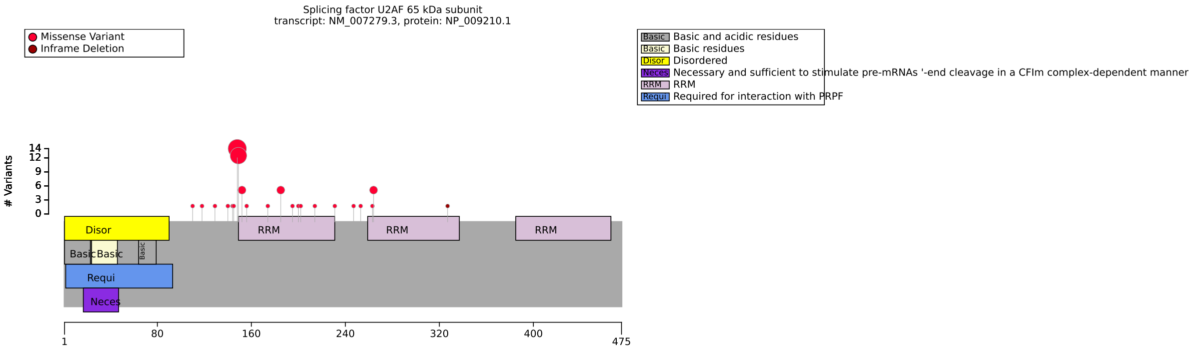


**(a)** Distribution of variants in U2AF2

| Genotype (A) | Genotype (B) | total tests performed | significant results |
|---|---|---|---|
| r149W | other | 177 | 0 |
| R149,R150 variants | other | 177 | 0 |
| RRM 1 | other | 177 | 0 |
| FEMALE | MALE | 177 | 0 |

**(b)** Fisher Exact Test performed to compare HPO annotation frequency with respect to genotypes.

**Figure S80:** The cohort comprised 48 individuals (28 females, 20 males). 2 of these individuals were reported to be deceased. A total of 178 HPO terms were used to annotate the cohort. Disease diagnosis: Developmental delay, dysmorphic facies, and brain anomalies (OMIM:620535). No statistical analysis of GPCs in U2AF2 identified in published literature. A total of 24 unique variant alleles were found in *U2AF2* (transcript: `NM_007279.3`, protein id: `NP_009210.1`).

### UMOD

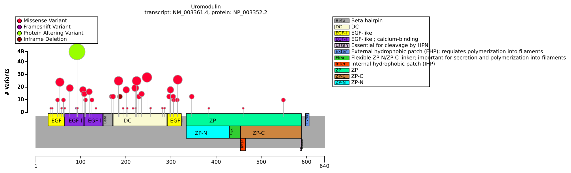

**(a)** Distribution of variants in UMOD

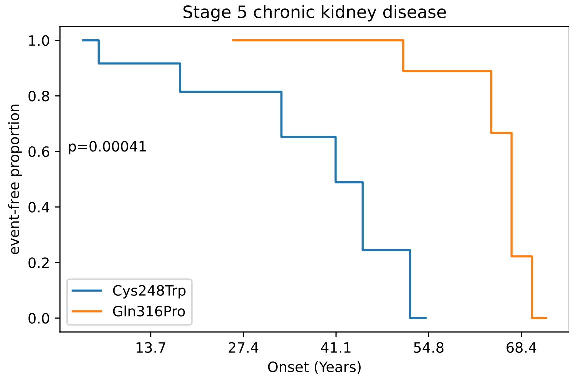


**(b)** Onset of Stage 5 chronic kidney disease (HP:0003774)

| HPO term | EGF | other | p-value | adj. p-value |
|---|---|---|---|---|
| Hyperuricemia [HP:0002149] | 14/32 (44%) | 50/57 (88%) | $1.77 \times 10^{-5}$ | $1.06 \times 10^{-4}$ |

**(c)** Fisher Exact Test: HPO annotation frequency and EGF vs. other. Total of 6 tests were performed.

| HPO term | cysteine | other | p-value | adj. p-value |
|---|---|---|---|---|
| Hyperuricemia [HP:0002149] | 38/41 (93%) | 26/48 (54%) | $4.48 \times 10^{-5}$ | $2.69 \times 10^{-4}$ |

**(d)** Fisher Exact Test: HPO annotation frequency and cysteine vs other. Total of 6 tests were performed.

| Genotype (A) | Genotype (B) | total tests performed | significant results |
|---|---|---|---|
| FEMALE | MALE | 5 | 0 |

**(e)** Fisher Exact Test performed to compare HPO annotation frequency with respect to genotypes.

| Description | Variable | Genotype (A) | Genotype (B) | p-value | xrefs |
|---|---|---|---|---|---|
| Survival analysis: Stage 5 chronic kidney disease | Onset of HP:0003774 | EGF | other | 0.284 | - |

**(f)** Onset of Stage 5 chronic kidney disease to compare EGF and other with respect to Onset of HP:0003774.

| Description | Variable | Genotype (A) | Genotype (B) | p-value | xrefs |
|---|---|---|---|---|---|
| Survival analysis: Stage 5 chronic kidney disease | Onset of HP:0003774 | 278_289delins | other | 0.835 | - |

**(g)** Onset of Stage 5 chronic kidney disease to compare 278_289delins and other with respect to Onset of HP:0003774.

| Description | Variable | Genotype (A) | Genotype (B) | p-value | xrefs |
|---|---|---|---|---|---|
| Survival analysis: Stage 5 chronic kidney disease | Onset of HP:0003774 | Cys248Trp | Gln316Pro | $4.10 \times 10^{-4}$ | - |

**(h)** Onset of Stage 5 chronic kidney disease to compare Cys248Trp and Gln316Pro with respect to Onset of Stage 5 chronic kidney disease (HP:0003774).

**Figure S81:** The cohort comprised 207 individuals (80 females, 88 males, 39 with unknown sex). 30 HPO terms were used to annotate the cohort. Disease diagnoses: Tubulointerstitial kidney disease, autosomal dominant, 1 (OMIM:162000) (183 individuals), Tubulointerstitial kidney disease, autosomal dominant, 3 (OMIM:162002) (12 individuals), Tubulointerstitial kidney disease, autosomal dominant, 2 (OMIM:162001) (12 individuals). One study on UMOD variants showed that median ages at ESRD development were lowest with Cys77Tyr and highest with Gln316Pro [64], which is compatible with our finding here. Another showed that indel mutation p.Val93_Gly97delinsAlaAlaSerCys is associated with a relatively mild clinical UAKD phenotype [65]. We did not observe a significant association between this variant and age of onset with our dataset. A total of 53 unique variant alleles were found in *UMOD* (transcript: `NM_003361.4`, protein id: `NP_003352.2`).

### WWOX

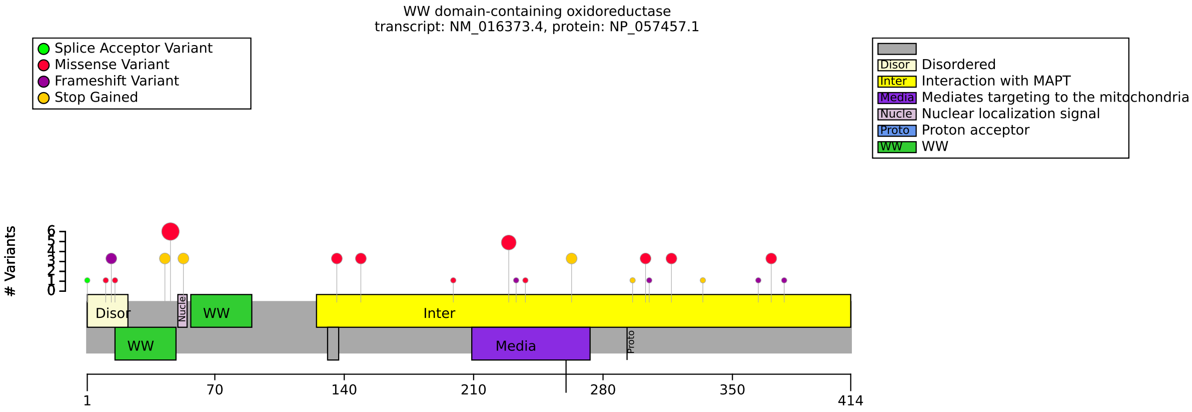


**(a)** Distribution of variants in WWOX

| HPO term | MAPT Interaction/MAPT Interaction OR MAPT Interaction/Other | Other/Other | p-value | adj. p-value |
|---|---|---|---|---|
| Bilateral tonic-clonic seizure with focal onset [HP:0007334] | 0/18 (0%) | 7/9 (78%) | $4.05 \times 10^{-5}$ | 0.002 |

**(b)** Fisher Exact Test performed to compare HPO annotation frequency with respect to MAPT Interaction/MAPT Interaction OR MAPT Interaction/Other and Other/Other. Total of 44 tests were performed.

| Genotype (A) | Genotype (B) | total tests performed | significant results |
|---|---|---|---|
| Missense/Missense OR Missense/Other | Other/Other | 44 | 0 |
| FEMALE | MALE | 44 | 0 |

**(c)** Fisher Exact Test performed to compare HPO annotation frequency with respect to genotypes.

**Figure S82:** The cohort comprised 38 individuals (25 females, 13 males). 11 of these individuals were reported to be deceased. A total of 72 HPO terms were used to annotate the cohort. Disease diagnoses: Developmental and epileptic encephalopathy 28 (OMIM:616211) (32 individuals), Spinocerebellar ataxia, autosomal recessive 12 (OMIM:614322) (6 individuals). Phenotype/genotype correlations were recently suggested for WWOX-related neurodevelopmental disorders with a classification of WWOX genotypes into three groups. According to the tentative classification, patients carrying two predicted null alleles were more likely to present with the most severe WOREE phenotype whereas hypomorphic genotypes with two missense variants would instead result in spinocerebellar ataxia (SCAR12) [66, 67, 68]. A total of 33 unique variant alleles were found in *WWOX* (transcript: `NM_016373.4`, protein id: `NP_057457.1`).

### ZFX

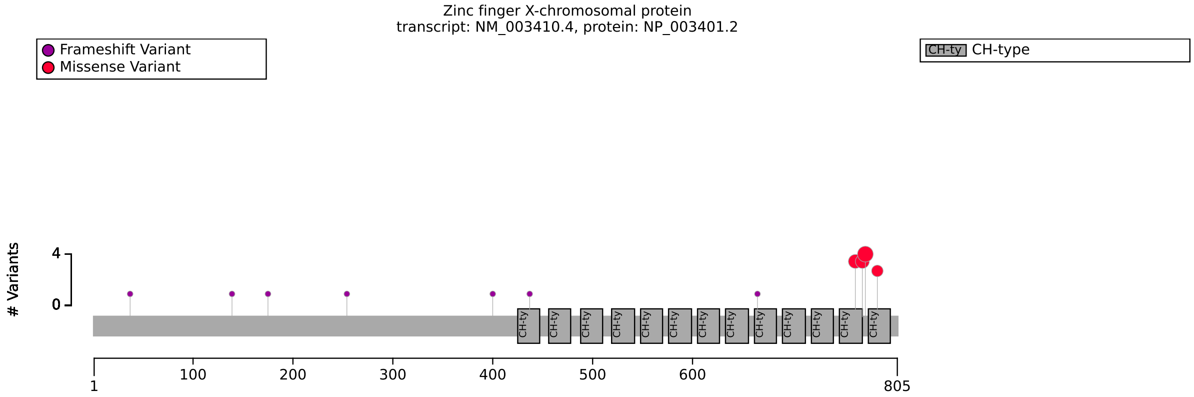


**(a)** Distribution of variants in ZFX

| HPO term | missense | other | p-value | adj. p-value |
|---|---|---|---|---|
| Hyperparathyroidism [HP:0000843] | 7/9 (78%) | 0/5 (0%) | 0.021 | 0.021 |

**(b)** Fisher Exact Test performed to compare HPO annotation frequency with respect to missense and other. Total of 1 test was performed.

| Genotype (A) | Genotype (B) | total tests performed | significant results |
|---|---|---|---|
| missense | other | 225 | 0 |

**(c)** Fisher Exact Test performed to compare HPO annotation frequency with respect to genotypes.

**Figure S83:** The cohort comprised 19 individuals (4 females, 14 males, 1 with unknown sex). A total of 203 HPO terms were used to annotate the cohort. Disease diagnosis: Intellectual developmental disorder, X-linked syndromic 37 (OMIM:301118). The small cohort size and the fact that the majority of the ZFX variants were private to each proband or family made the assessment of genotype-phenotype correlation difficult. A recent report of a germline ZFX missense variant in a patient With primary hyperparathyroidism suggested the hypothesis of testing for correlation between missense variants and hyperparathyroidism [69, 70]. A total of 11 unique variant alleles were found in *ZFX* (transcript: `NM_003410.4`, protein id: `NP_003401.2`).

### ZMYM3

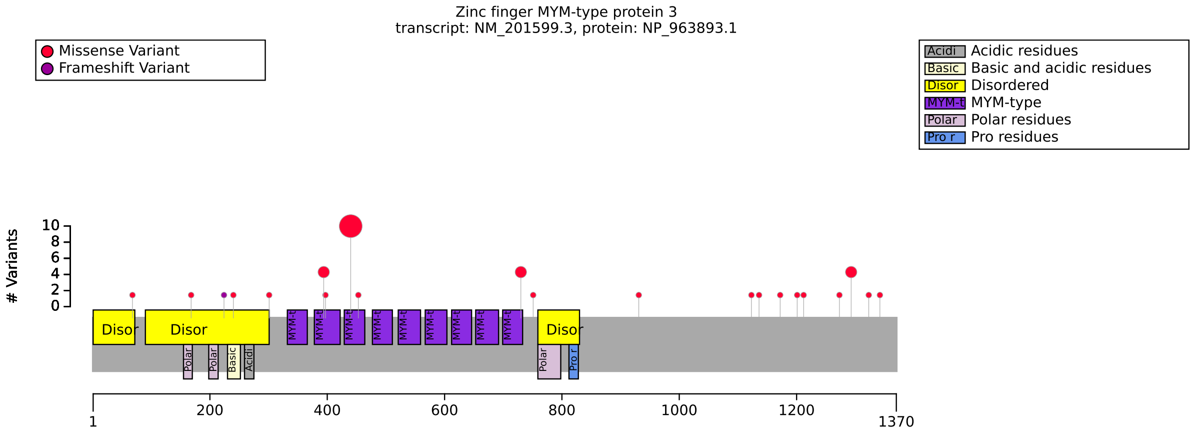


**(a)** Distribution of variants in ZMYM3

| HPO term | R441 | other | p-value | adj. p-value |
|---|---|---|---|---|
| Cupped ear [HP:0000378] | 7/10 (70%) | 1/23 (4%) | $2.02 \times 10^{-4}$ | 0.012 |

**(b)** Fisher Exact Test performed to compare HPO annotation frequency with respect to R441 and other. Total of 60 tests were performed.

| Genotype (A) | Genotype (B) | total tests performed | significant results |
|---|---|---|---|
| N term | other | 60 | 0 |

**(c)** Fisher Exact Test performed to compare HPO annotation frequency with respect to genotypes.

**Figure S84:** The cohort comprised 33 individuals (3 females, 30 males). 1 of these individuals were reported to be deceased. A total of 78 HPO terms were used to annotate the cohort. Disease diagnosis: Intellectual developmental disorder, X-linked 112 (OMIM:301111). A total of 22 unique variant alleles were found in *ZMYM3* (transcript: `NM_201599.3`, protein id: `NP_963893.1`). No analysis of ZMYM3 genotype phgenotype correlation was presented in the two studies on ZMYM3 variants study to be published to date [71, 72].

### ZNF462

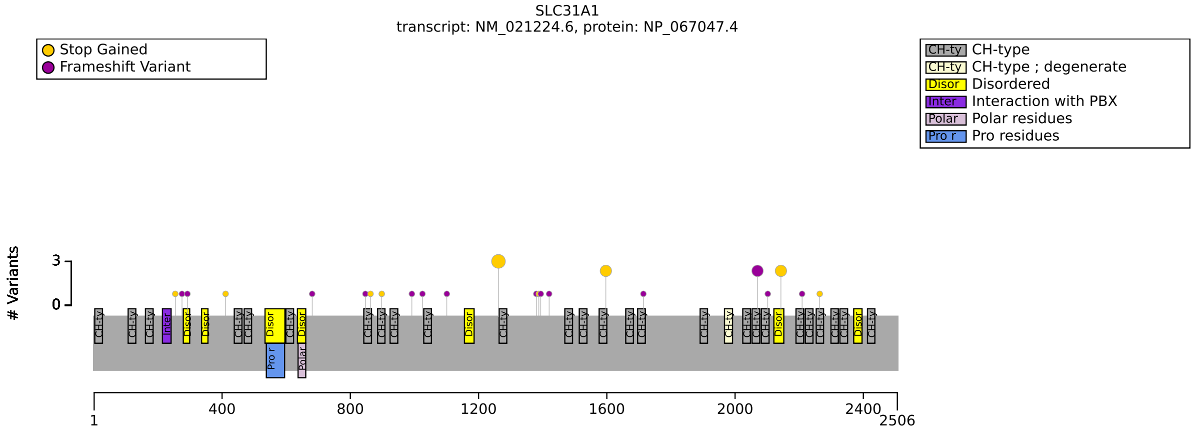


**(a)** Distribution of variants in ZNF462

| Genotype (A) | Genotype (B) | total tests performed | significant results |
|---|---|---|---|
| Ablation | Other | 43 | 0 |
| FEMALE | MALE | 41 | 0 |

**(b)** Fisher Exact Test performed to compare HPO annotation frequency with respect to genotypes.

**Figure S85:** The cohort comprised 39 individuals (11 females, 25 males, 3 with unknown sex). A total of 32 HPO terms were used to annotate the cohort. Disease diagnosis: Weiss-Kruszka syndrome (OMIM:618619). No statistically significant genotype phenotype association was identified. A total of 26 unique variant alleles were found in *ZNF462* (transcript: `NM_021224.6`, protein id: `NP_067047.4`).

### ZSWIM6

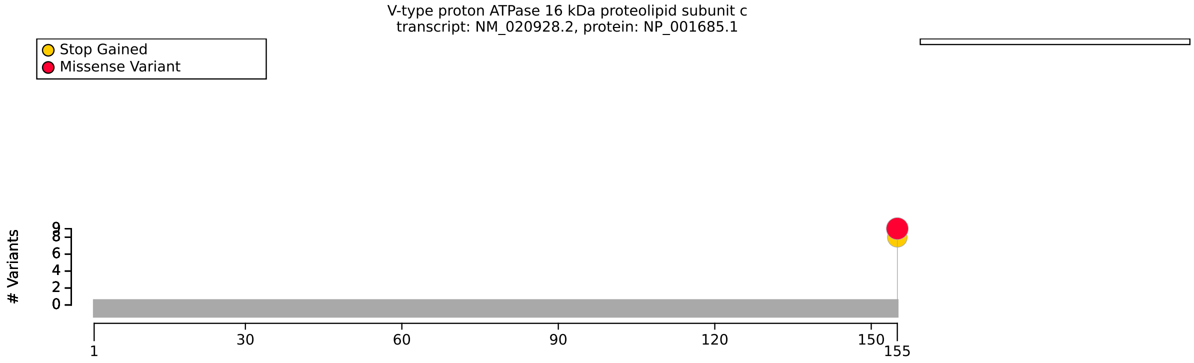


**(a)** Distribution of variants in ZSWIM6

| Genotype (A) | Genotype (B) | total tests performed | significant results |
|---|---|---|---|
| p.Arg913Ter | Arg1163Trp | 12 | 0 |

**(b)** Fisher Exact Test performed to compare HPO annotation frequency with respect to genotypes.

**Figure S86:** The cohort comprised 16 individuals (8 females, 8 males). A total of 85 HPO terms were used to annotate the cohort. Disease diagnoses: Acromelic frontonasal dysostosis (OMIM:603671) (9 individuals), Neurodevelopmental disorder with movement abnormalities, abnormal gait, and autistic features (OMIM:617865) (7 individuals). No statistically significant results identified. A total of 2 unique variant alleles were found in *ZSWIM6* (transcript: `NM_020928.2`, protein id: `NP_001685.1`).

### References


[1] Elena Martinez-Cayuelas, Fiona Blanco-Kelly, Fermina Lopez-Grondona, Saoud Tahsin Swafiri, Rosario Lopez-Rodriguez, Rebeca Losada-Del Pozo, Ignacio Mahillo-Fernandez, Beatriz Moreno, Maria Rodrigo-Moreno, Didac Casas-Alba, Aitor Lopez-Gonzalez, Sixto García-Miñaúr, Maria Ángeles Mori, Marta Pacio-Minguez, Emi Rikeros-Orozco, Fernando Santos-Simarro, Jaime Cruz-Rojo, Juan Francisco Quesada-Espinosa, Maria Teresa Sanchez-Calvin, Jaime Sanchez-Del Pozo, Raquel Bernado Fonz, Maria Isidoro-Garcia, Irene Ruiz-Ayucar, Maria Isabel Alvarez-Mora, Raquel Blanco-Lago, Begoña De Azua, Jesus Eiris, Juan Jose Garcia-Peñas, Belen Gil-Fournier, Carmen Gomez-Lado, Nadia Irazabal, Vanessa Lopez-Gonzalez, Irene Madrigal, Ignacio Malaga, Beatriz Martinez-Menendez, Soraya Ramiro-Leon, Maria Garcia-Hoyos, Pablo Prieto-Matos, Javier Lopez-Pison, Sergio Aguilera-Albesa, Sara Alvarez, Alberto Fernández-Jaén, Isabel Llano-Rivas, Blanca Gener-Querol, Carmen Ayuso, Ana Arteche-Lopez, Maria Palomares-Bralo, Anna Cueto-González, Irene Valenzuela, Antonio Martinez-Monseny, Isabel Lorda-Sanchez, and Berta Almoguera. Clinical description, molecular delineation and genotype-phenotype correlation in 340 patients with KBG syndrome: addition of 67 new patients. *J. Med. Genet.*, 60(7):644–654, July 2023.

[2] Alexander J M Dingemans, Kim M G Truijen, Jung-Hyun Kim, Zahide Alaçam, Laurence Faivre, Kathleen M Collins, Erica H Gerkes, Mieke van Haelst, Ingrid M B H van de Laar, Kristin Lindstrom, Mathilde Nizon, James Pauling, Edyta Heropolitańska-Pliszka, Astrid S Plomp, Caroline Racine, Rani Sachdev, Margje Sinnema, Jon Skranes, Hermine E Veenstra-Knol, Eline A Verberne, Anneke T Vulto-van Silfhout, Marlon E F Wilsterman, Eun-Young Erin Ahn, Bert B A de Vries, and Lisenka E L M Vissers. Establishing the phenotypic spectrum of ZTTK syndrome by analysis of 52 individuals with variants in SON. *Eur. J. Hum. Genet.*, 30(3):271–281, March 2022.

[3] Sara Tucci, Christine Wagner, Sarah C Grünert, Uta Matysiak, Natalie Weinhold, Jeannette Klein, Francesco Porta, Marco Spada, Andrea Bordugo, Giulia Rodella, Francesca Furlan, Anna Sajeva, Francesca Menni, and Ute Spiekerkoetter. Genotype and residual enzyme activity in medium-chain acyl-CoA dehydrogenase (MCAD) deficiency: Are predictions possible? *J. Inherit. Metab. Dis.*, 44(4):916–925, July 2021.

[4] Maria Halonen, Petra Eskelin, Anne-Grethe Myhre, Jaakko Perheentupa, Eystein S Husebye, Olle Kämpe, Fredrik Rorsman, Leena Peltonen, Ismo Ulmanen, and Jukka Partanen. AIRE mutations and human leukocyte antigen genotypes as determinants of the autoimmune polyendocrinopathy-candidiasis-ectodermal dystrophy phenotype. *J. Clin. Endocrinol. Metab.*, 87(6):2568–2574, June 2002.

[5] Zain Awamleh, Sanaa Choufani, Cheryl Cytrynbaum, Fowzan S. Alkuraya, Stephen Scherer, Sofia Fernandes, Catarina Rosas, Pedro Louro, Patricia Dias, Mariana Tomásio Neves, Sérgio B. Sousa, and Rosanna Weksberg. Ankrd11 pathogenic variants and 16q24.3 microdeletions share an altered dna methylation signature in patients with kbg syndrome. *Human molecular genetics*, 32:1429–1438, April 2023.

[6] Alexander J M Dingemans, Kim M G Truijen, Sam van de Ven, Raphael Bernier, Ernie M H F Bongers, Arjan Bouman, Laura de Graaff-Herder, Evan E Eichler, Erica H Gerkes, Christa M De Geus, Johanna M van Hagen, Philip R Jansen, Jennifer Kerkhof, Anneke J A Kievit, Tjitske Kleefstra, Saskia M Maas, Stella A de Man, Haley McConkey, Wesley G Patterson, Amy T Dobson, Eloise J Prijoles, Bekim Sadikovic, Raissa Relator, Roger E Stevenson, Connie T R M Stumpel, Malou Heijligers, Kyra E Stuurman, Katharina Löhner, Shimriet Zeidler, Jennifer A Lee, Amanda Lindy, Fanggeng Zou, Matthew L Tedder, Lisenka E L M Vissers, and Bert B A de Vries. The phenotypic spectrum and genotype-phenotype correlations in 106 patients with variants in major autism gene CHD8. *Transl. Psychiatry*, 12(1):421, October 2022.

[7] Martin Konrad, Jianghui Hou, Stefanie Weber, Jörg Dötsch, Jameela A Kari, Tomas Seeman, Eberhard Kuwertz-Bröking, Amira Peco-Antic, Velibor Tasic, Katalin Dittrich, Hammad O Alshaya, Rodo O von Vigier, Sabina Gallati, Daniel A Goodenough, and André Schaller. CLDN16 genotype predicts renal decline in familial hypomagnesemia with hypercalciuria and nephrocalcinosis. *J. Am. Soc. Nephrol.*, 19(1):171–181, January 2008.

[8] Gianluca D'Onofrio, Andrea Accogli, Mariasavina Severino, Haluk Caliskan, Tomislav Kokotović, Antonela Blazekovic, Kristina Gotovac Jercic, Silvana Markovic, Tamara Zigman, Krnjak Goran, Nina Barišić, Vlasta Duranovic, Ana Ban, Fran Borovecki, Danijela Petković Ramadža, Ivo Barić, Walid Fazeli, Peter Herkenrath, Carla Marini, Roberta Vittorini, Vykuntaraju Gowda, Arjan Bouman, Clarissa Rocca, Issam Azmi Alkhawaja, Bibi Nazia Murtaza, Malik Mujaddad Ur Rehman, Chadi Al Alam, Gisele Nader, Maria Margherita Mancardi, Thea Giacomini, Siddharth Srivastava, Javeria Raza Alvi, Hoda Tomoum, Sara Matricardi, Michele Iacomino, Antonella Riva, Marcello Scala, Francesca Madia, Angela Pistorio, Vincenzo Salpietro, Carlo Minetti, Jean-Baptiste Rivière, Myriam Srour, Stephanie Efthymiou, Reza Maroofian, Henry Houlden, Sonja Catherine Vernes, Federico Zara, Pasquale Striano, and Vanja Nagy. Genotype-phenotype correlation in contactin-associated protein-like 2 (CNTNAP-2) developmental disorder. *Hum. Genet.*, 142(7):909–925, July 2023.

[9] Jieqiong Xie, Jiayang Jiang, and Qiwei Guo. Primary coenzyme q10 deficiency-7 and pathogenic coq4 variants: Clinical presentation, biochemical analyses, and treatment. *Frontiers in genetics*, 12:776807, 2021.

[10] Hannah Gabriela Valverde de Morales, Hsiao-Lin V Wang, Kathryn Garber, Xiaodong Cheng, Victor G Corces, and Hong Li. Expansion of the genotypic and phenotypic spectrum of CTCF-related disorder guides clinical management: 43 new subjects and a comprehensive literature review. *Am. J. Med. Genet. A*, 191(3):718–729, March 2023.

[11] Chao Xu, Wenyu Jia, Xiangdeng Cheng, Hui Ying, Jing Chen, Jin Xu, Qingbo Guan, Xinli Zhou, Dongmei Zheng, Guimei Li, and Jiajun Zhao. Genotype-phenotype correlation study and mutational and hormonal analysis in a chinese cohort with 21-hydroxylase deficiency. *Molecular genetics & genomic medicine*, 7:e671, June 2019.

[12] Dmitrijs Rots, Arianne Bouman, Ayumi Yamada, Michael Levy, Alexander J. M. Dingemans, Bert B. A. de Vries, Martina Ruiterkamp-Versteeg, Nicole de Leeuw, Charlotte W. Ockeloen, Rolph Pfundt, Elke de Boer, Joost Kummeling, Bregje van Bon, Hans van Bokhoven, Nael Nadif Kasri, Hanka Venselaar, Marielle Alders, Jennifer Kerkhof, Haley McConkey, Alma Kuechler, Bart Elffers, Rixje van Beeck Calkoen, Susanna Hofman, Audrey Smith, Maria Irene Valenzuela, Siddharth Srivastava, Zoe Frazier, Isabelle Maystadt, Carmelo Piscopo, Giuseppe Merla, Meena Balasubramanian, Gijs W. E. Santen, Kay Metcalfe, Soo-Mi Park, Laurent Pasquier, Siddharth Banka, Dian Donnai, Daniel Weisberg, Gertrud Strobl-Wildemann, Annemieke Wagemans, Maaike Vreeburg, Diana Baralle, Nicola Foulds, Ingrid Scurr, Nicola Brunetti-Pierri, Johanna M. van Hagen, Emilia K. Bijlsma, Anna H. Hakonen, Carolina Courage, David Genevieve, Lucile Pinson, Francesca Forzano, Charu Deshpande, Maria L. Kluskens, Lindsey Welling, Astrid S. Plomp, Els K. Vanhoutte, Louisa Kalsner, Janna A. Hol, Audrey Putoux, Johanna Lazier, Pradeep Vasudevan, Elizabeth Ames, Jessica O'Shea, Damien Lederer, Julie Fleischer, Mary O'Connor, Melissa Pauly, Georgia Vasileiou, André Reis, Catherine Kiraly-Borri, Arjan Bouman, Chris Barnett, Marjan Nezarati, Lauren Borch, Gea Beunders, Kübra Özcan, Stéphanie Miot, Catharina M. L. Volker-Touw, Koen L. I. van Gassen, Gerarda Cappuccio, Katrien Janssens, Nofar Mor, Inna Shomer, Dan Dominissini, Matthew L. Tedder, Alison M. Muir, Bekim Sadikovic, Han G. Brunner, Lisenka E. L. M. Vissers, Yoichi Shinkai, and Tjitske Kleefstra. Comprehensive ehmt1 variants analysis broadens genotype-phenotype associations and molecular mechanisms in kleefstra syndrome. *American journal of human genetics*, 111:1605–1625, August 2024.

[13] Zoë J. Frazier, Seyda Kilic, Hailey Osika, Alisa Mo, Meg Quinn, Sonia Ballal, Tamar Katz, A. Eliot Shearer, Max A. Horlbeck, Lynn S. Pais, Kira A. Dies, Anne O'Donnell-Luria, Joe Kossowsky, Jonathan O. Lipton, Tjitske Kleefstra, and Siddharth Srivastava. Novel phenotypes and genotype-phenotype correlations in a large clinical cohort of patients with kleefstra syndrome. *Clinical genetics*, January 2025.

[14] Carolina Gracia-Diaz, Yijing Zhou, Qian Yang, Reza Maroofian, Paula Espana-Bonilla, Chul-Hwan Lee, Shuo Zhang, Natàlia Padilla, Raquel Fueyo, Elisa A Waxman, Sunyimeng Lei, Garrett Otrimski, Dong Li, Sarah E Sheppard, Paul Mark, Margaret H Harr, Hakon Hakonarson, Lance Rodan, Adam Jackson, Pradeep Vasudevan, Corrina Powel, Shehla Mohammed, Sateesh Maddirevula, Hamad Alzaidan, Eissa A Faqeih, Stephanie Efthymiou, Valentina Turchetti, Fatima Rahman, Shazia Maqbool, Vincenzo Salpietro, Shahnaz H Ibrahim, Gabriella di Rosa, Henry Houlden, Maha Nasser Alharbi, Nouriya Abbas Al-Sannaa, Peter Bauer, Giovanni Zifarelli, Conchi Estaras, Anna C E Hurst, Michelle L Thompson, Anna Chassevent, Constance L Smith-Hicks, Xavier de la Cruz, Alexander M Holtz, Houda Zghal Elloumi, M J Hajianpour, Claudine Rieubland, Dominique Braun, Siddharth Banka, Genomic England Research Consortium, Deborah L French, Elizabeth A Heller, Murielle Saade, Hongjun Song, Guo-Li Ming, Fowzan S Alkuraya, Pankaj B Agrawal, Danny Reinberg, Elizabeth J Bhoj, Marian A Martínez-Balbás, and Naiara Akizu. Gain and loss of function variants in EZH1 disrupt neurogenesis and cause dominant and recessive neurodevelopmental disorders. *Nat. Commun.*, 14(1):4109, July 2023.

[15] Víctor Manuel Becerra-Muñoz, Juan José Gómez-Doblas, Carlos Porras-Martín, Miguel Such-Martínez, María Generosa Crespo-Leiro, Roberto Barriales-Villa, Eduardo de Teresa-Galván, Manuel Jiménez-Navarro, and Fernando Cabrera-Bueno. The importance of genotype-phenotype correlation in the clinical management of marfan syndrome. *Orphanet journal of rare diseases*, 13:16, January 2018.

[16] Pauline Arnaud, Olivier Milleron, Nadine Hanna, Jacques Ropers, Nadia Ould Ouali, Amel Affoune, Maud Langeois, Ludivine Eliahou, Florence Arnoult, Philippe Renard, Marlène Michelon-Jouneaux, Marie Cotillon, Laurent Gouya, Catherine Boileau, and Guillaume Jondeau. Clinical relevance of genotype-phenotype correlations beyond vascular events in a cohort study of 1500 marfan syndrome patients with fbn1 pathogenic variants. *Genetics in medicine : official journal of the American College of Medical Genetics*, 23:1296–1304, July 2021.

[17] Amparo Hernándiz, Angel Zúñiga, Francisco Valera, Diana Domingo, Imelda Ontoria-Oviedo, Jose F. Marí, Jose A. Román, Inmaculada Calvo, Beatriz Insa, Rosa Gómez, José V. Cervera, Manuel Miralles, Jose A. Montero, Luis Martínez-Dolz, and Pilar Sepúlveda. Genotype fbn1/phenotype relationship in a cohort of patients with marfan syndrome. *Clinical genetics*, 99:269–280, February 2021.

[18] Kristian A. Groth, Yskert Von Kodolitsch, Kerstin Kutsche, Mette Gaustadnes, Kasper Thorsen, Niels H. Andersen, and Claus H. Gravholt. Evaluating the quality of marfan genotype-phenotype correlations in existing fbn1 databases. *Genetics in medicine : official journal of the American College of Medical Genetics*, 19:772–777, July 2017.

[19] Ayman W El-Hattab, Hongzheng Dai, Mohammed Almannai, Julia Wang, Eissa A Faqeih, Ali Al Asmari, Mohammed A M Saleh, Mohammed A O Elamin, Majid Alfadhel, Fowzan S Alkuraya, Mais Hashem, Mazhor S Aldosary, Rawan Almass, Faten B Almutairi, Maysoon Alsagob, Mohammed Al-Owain, Shirin Al-Sharfa, Zuhair N Al-Hassnan, Zuhair Rahbeeni, Mohammed A Al-Muhaizea, Nawal Makhseed, Gretchen K Foskett, David A Stevenson, Natalia Gomez-Ospina, Chung Lee, Richard G Boles, Samantha A Schrier Vergano, Saskia B Wortmann, Wolfgang Sperl, Thomas Opladen, Georg F Hoffmann, Maja Hempel, Holger Prokisch, Bader Alhaddad, Johannes A Mayr, Wenyaw Chan, Namik Kaya, and Lee-Jun C Wong. Molecular and clinical spectra of FBXL4 deficiency. *Hum. Mutat.*, 38(12):1649–1659, December 2017.

[20] Sujuan Li, Anran Tian, Yu Wen, Wei Gu, Wei Li, Xiaohong Qiao, Cai Zhang, and Xiaoping Luo. Fgd1-related aarskog-scott syndrome: Identification of four novel variations and a literature review of clinical and molecular aspects. *European journal of pediatrics*, 183:2257–2272, May 2024.

[21] Victor Zanetti Drumond, Lucas Sousa Salgado, Camila Sousa Salgado, Vitor Augusto de Lima Oliveira, Eliene Magda de Assis, Michel Campos Ribeiro, Analina Furtado Valadão, and Alfredo Orrico. The prevalence of clinical features in patients with aarskog-scott syndrome and assessment of genotype-phenotype correlation: A systematic review. *Genetics research*, 2021:6652957, 2021.

[22] Jennifer J. Johnston, Isabelle Olivos-Glander, Christina Killoran, Emma Elson, Joyce T. Turner, Kathryn F. Peters, Margaret H. Abbott, David J. Aughton, Arthur S. Aylsworth, Michael J. Bamshad, Carol Booth, Cynthia J. Curry, Albert David, Mary Beth Dinulos, David B. Flannery, Michelle A. Fox, John M. Graham, Dorothy K. Grange, Alan E. Guttmacher, Mark C. Hannibal, Wolfram Henn, Raoul C. M. Hennekam, Lewis B. Holmes, H. Eugene Hoyme, Kathleen A. Leppig, Angela E. Lin, Patrick Macleod, David K. Manchester, Carlo Marcelis, Laura Mazzanti, Emma McCann, Marie T. McDonald, Nancy J. Mendelsohn, John B. Moeschler, Billur Moghaddam, Giovanni Neri, Ruth Newbury-Ecob, Roberta A. Pagon, John A. Phillips, Laurie S. Sadler, Joan M. Stoler, David Tilstra, Catherine M. Walsh Vockley, Elaine H. Zackai, Touran M. Zadeh, Louise Brueton, Graeme Charles M. Black, and Leslie G. Biesecker. Molecular and clinical analyses of greig cephalopolysyndactyly and pallister-hall syndromes: robust phenotype prediction from the type and position of gli3 mutations. *American journal of human genetics*, 76:609–622, April 2005.

[23] Aleksander Jamsheer, Anna Sowińska, Tomasz Trzeciak, Małgorzata Jamsheer-Bratkowska, Anita Geppert, and Anna Latos-Bieleńska. Expanded mutational spectrum of the gli3 gene substantiates genotype-phenotype correlations. *Journal of applied genetics*, 53:415–422, November 2012.

[24] Rashmi Patel, Subodh Kumar Singh, Visweswar Bhattacharya, and Akhtar Ali. Novel gli3 pathogenic variants in complex pre- and postaxial polysyndactyly and greig cephalopolysyndactyly syndrome. *American journal of medical genetics. Part A*, 185:97–104, January 2021.

[25] Florence Démurger, Amale Ichkou, Soumaya Mougou-Zerelli, Martine Le Merrer, Géraldine Goudefroye, Anne-Lise Delezoide, Chloé Quélin, Sylvie Manouvrier, Geneviève Baujat, Mélanie Fradin, Laurent Pasquier, André Megarbané, Laurence Faivre, Clarisse Baumann, Sheela Nampoothiri, Joëlle Roume, Bertrand Isidor, Didier Lacombe, Marie-Ange Delrue, Sandra Mercier, Nicole Philip, Elise Schaefer, Muriel Holder, Amanda Krause, Fanny Laffargue, Martine Sinico, Daniel Amram, Gwenaelle André, Alain Liquier, Massimiliano Rossi, Jeanne Amiel, Fabienne Giuliano, Odile Boute, Anne Dieux-Coeslier, Marie-Line Jacquemont, Alexandra Afenjar, Lionel Van Maldergem, Marylin Lackmy-Port-Lis, Catherine Vincent-Delorme, Marie-Liesse Chauvet, Valérie Cormier-Daire, Louise Devisme, David Geneviève, Arnold Munnich, Géraldine Viot, Odile Raoul, Serge Romana, Marie Gonzales, Ferechte Encha-Razavi, Sylvie Odent, Michel Vekemans, and Tania Attie-Bitach. New insights into genotype-phenotype correlation for gli3 mutations. *European journal of human genetics : EJHG*, 23:92–102, January 2015.

[26] L. G. Biesecker. Strike three for gli3. *Nature genetics*, 17:259–260, November 1997.

[27] M. M. Al-Qattan, H. E. Shamseldin, M. A. Salih, and F. S. Alkuraya. Gli3-related polydactyly: a review. *Clinical genetics*, 92:457–466, November 2017.

[28] Henrike Lisa Sczakiel, Wiebke Hülsemann, Manuel Holtgrewe, Angela Teresa Abad-Perez, Jonas Elsner, Sarina Schwartzmann, Denise Horn, Malte Spielmann, Stefan Mundlos, and Martin Atta Mensah. Gli3 variants causing isolated polysyndactyly are not restricted to the protein's c-terminal third. *Clinical genetics*, 100:758–765, December 2021.

[29] David Boutboul, Hye Sun Kuehn, Zoé Van de Wyngaert, Julie E. Niemela, Isabelle Callebaut, Jennifer Stoddard, Christelle Lenoir, Vincent Barlogis, Catherine Farnarier, Frédéric Vely, Nao Yoshida, Seiji Kojima, Hirokazu Kanegane, Akihiro Hoshino, Fabian Hauck, Ludovic Lhermitte, Vahid Asnafi, Philip Roehrs, Shaoying Chen, James W. Verbsky, Katherine R. Calvo, Ammar Husami, Kejian Zhang, Joseph Roberts, David Amrol, John Sleaseman, Amy P. Hsu, Steven M. Holland, Rebecca Marsh, Alain Fischer, Thomas A. Fleisher, Capucine Picard, Sylvain Latour, and Sergio D. Rosenzweig. Dominant-negative ikzf1 mutations cause a t, b, and myeloid cell combined immunodeficiency. *The Journal of clinical investigation*, 128:3071–3087, July 2018.

[30] Margaret P. Adam, Siddharth Banka, Hans T. Bjornsson, Olaf Bodamer, Albert E. Chudley, Jaqueline Harris, Hiroshi Kawame, Brendan C. Lanpher, Andrew W. Lindsley, Giuseppe Merla, Noriko Miyake, Nobuhiko Okamoto, Constanze T. Stumpel, Norio Niikawa, and Kabuki Syndrome Medical Advisory Board. Kabuki syndrome: international consensus diagnostic criteria. *Journal of medical genetics*, 56:89–95, February 2019.

[31] P. Makrythanasis, B. W. van Bon, M. Steehouwer, B. Rodríguez-Santiago, M. Simpson, P. Dias, B. M. Anderlid, P. Arts, M. Bhat, B. Augello, E. Biamino, E. M. H. F. Bongers, M. Del Campo, I. Cordeiro, A. M. Cueto-González, I. Cuscó, C. Deshpande, E. Frysira, L. Izatt, R. Flores, E. Galán, B. Gener, C. Gilissen, S. M. Granneman, J. Hoyer, H. G. Yntema, C. M. Kets, D. A. Koolen, C. l Marcelis, A. Medeira, L. Micale, S. Mohammed, S. A. de Munnik, A. Nordgren, S. Psoni, W. Reardon, N. Revencu, T. Roscioli, M. Ruiterkamp-Versteeg, H. G. Santos, J. Schoumans, J. H. M. Schuurs-Hoeijmakers, M. C. Silengo, L. Toledo, T. Vendrell, I. van der Burgt, B. van Lier, C. Zweier, A. Reymond, R. C. Trembath, L. Perez-Jurado, J. Dupont, B. B. A. de Vries, H. G. Brunner, J. A. Veltman, G. Merla, S. E. Antonarakis, and A. Hoischen. Mll2 mutation detection in 86 patients with kabuki syndrome: a genotype-phenotype study. *Clinical genetics*, 84:539–545, December 2013.

[32] Yirou Wang, Yufei Xu, Yao Chen, Yabin Hu, Qun Li, Shijian Liu, Jian Wang, and Xiumin Wang. Sex-specific difference in phenotype of kabuki syndrome type 2 patients: a matched case-control study. *BMC pediatrics*, 24:133, February 2024.

[33] Hannah C. Happ, Lynette G. Sadleir, Matthew Zemel, Guillem de Valles-Ibáñez, Michael S. Hildebrand, Allyn McConkie-Rosell, Marie McDonald, Halie May, Tristan Sands, Vimla Aggarwal, Christopher Elder, Timothy Feyma, Allan Bayat, Rikke S. Møller, Christina D. Fenger, Jens Erik Klint Nielsen, Anita N. Datta, Kathleen M. Gorman, Mary D. King, Natalia D. Linhares, Barbara K. Burton, Andrea Paras, Sian Ellard, Julia Rankin, Anju Shukla, Purvi Majethia, Rory J. Olson, Karthik Muthusamy, Lisa A. Schimmenti, Keith Starnes, Lucie Sedláčková, Katalin Štěrbová, Markéta Vlčková, Petra Laššuthová, Alena Jahodová, Brenda E. Porter, Nathalie Couque, Estelle Colin, Clément Prouteau, Corinne Collet, Thomas Smol, Roseline Caumes, Fleur Vansenne, Francesca Bisulli, Laura Licchetta, Richard Person, Erin Torti, Kirsty McWalter, Richard Webster, Elizabeth E. Gerard, Gaetan Lesca, Pierre Szepetowski, Ingrid E. Scheffer, Heather C. Mefford, and Gemma L. Carvill. Neurodevelopmental and epilepsy phenotypes in individuals with missense variants in the voltage-sensing and pore domains of kcnh5. *Neurology*, 100:e603–e615, February 2023.

[34] Víctor Faundes, Stephanie Goh, Rhoda Akilapa, Heidre Bezuidenhout, Hans T. Bjornsson, Lisa Bradley, Angela F. Brady, Elise Brischoux-Boucher, Han Brunner, Saskia Bulk, Natalie Canham, Declan Cody, Maria Lisa Dentici, Maria Cristina Digilio, Frances Elmslie, Andrew E. Fry, Harinder Gill, Jane Hurst, Diana Johnson, Sophie Julia, Katherine Lachlan, Robert Roger Lebel, Melissa Byler, Eric Gershon, Edmond Lemire, Maria Gnazzo, Francesca Romana Lepri, Antonia Marchese, Meriel McEntagart, Julie McGaughran, Seiji Mizuno, Nobuhiko Okamoto, Claudine Rieubland, Jonathan Rodgers, Erina Sasaki, Emmanuel Scalais, Ingrid Scurr, Mohnish Suri, Ineke van der Burgt, Naomichi Matsumoto, Noriko Miyake, Valérie Benoit, Damien Lederer, and Siddharth Banka. Clinical delineation, sex differences, and genotype-phenotype correlation in pathogenic kdm6a variants causing x-linked kabuki syndrome type 2. *Genetics in medicine : official journal of the American College of Medical Genetics*, 23:1202–1210, July 2021.

[35] Elliot S. Stolerman, Elizabeth Francisco, Jennifer L. Stallworth, Julie R. Jones, Kristin G. Monaghan, Jennifer Keller-Ramey, Richard Person, Ingrid M. Wentzensen, Kirsty McWalter, Boris Keren, Benedicte Heron, Caroline Nava, Delphine Heron, Katherine Kim, Barbara Burton, Fatima Al-Musafri, Lauren O'Grady, Inderneel Sahai, Luis F. Escobar, Marije Meuwissen, Edwin Reyniers, Frank Kooy, Yves Lacassie, Meral Gunay-Aygun, Krista Sondergaard Schatz, Ron Hochstenbach, Petra J. G. Zwijnenburg, Quinten Waisfisz, Marjon van Slegtenhorst, Grazia M. S. Mancini, and Raymond J. Louie. Genetic variants in the kdm6b gene are associated with neurodevelopmental delays and dysmorphic features. *American journal of medical genetics. Part A*, 179:1276–1286, July 2019.

[36] Gabriella Captur, Eloisa Arbustini, Petros Syrris, Dina Radenkovic, Ben O'Brien, William J. Mckenna, and James C. Moon. Lamin mutation location predicts cardiac phenotype severity: combined analysis of the published literature. *Open heart*, 5:e000915, 2018.

[37] Eric W. Lin, Graham F. Brady, Raymond Kwan, Alexey I. Nesvizhskii, and M. Bishr Omary. Genotype-phenotype analysis of lmna-related diseases predicts phenotype-selective alterations in lamin phosphorylation. *FASEB journal : official publication of the Federation of American Societies for Experimental Biology*, 34:9051–9073, July 2020.

[38] Ayman W. El-Hattab, Fang-Yuan Li, Eric Schmitt, Shulin Zhang, William J. Craigen, and Lee-Jun C. Wong. Mpv17-associated hepatocerebral mitochondrial dna depletion syndrome: new patients and novel mutations. *Molecular genetics and metabolism*, 99:300–308, March 2010.

[39] Margaret P. Adam, Jerry Feldman, Ghayda M. Mirzaa, Roberta A. Pagon, Stephanie E. Wallace, and Anne Amemiya. Genereviews (®). 1993.

[40] Nicole Hammann, Dominic Lenz, Ivo Baric, Ellen Crushell, Carlo Dionisi Vici, Felix Distelmaier, Francois Feillet, Peter Freisinger, Maja Hempel, Anna L. Khoreva, Martin W. Laass, Yves Lacassie, Elke Lainka, Catherine Larson-Nath, Zhongdie Li, Patryk Lipiński, Eberhard Lurz, André Mégarbané, Susana Nobre, Giorgia Olivieri, Bianca Peters, Paolo Prontera, Lea D. Schlieben, Christine M. Seroogy, Cristina Sobacchi, Shigeru Suzuki, Christel Tran, Jerry Vockley, Jian-She Wang, Matias Wagner, Holger Prokisch, Sven F. Garbade, Stefan Kölker, Georg F. Hoffmann, and Christian Staufner. Impact of genetic and non-genetic factors on phenotypic diversity in nbas-associated disease. *Molecular genetics and metabolism*, 141:108118, March 2024.

[41] Kitiwan Rojnueangnit, Jing Xie, Alicia Gomes, Angela Sharp, Tom Callens, Yunjia Chen, Ying Liu, Meagan Cochran, Mary-Alice Abbott, Joan Atkin, Dusica Babovic-Vuksanovic, Christopher P. Barnett, Melissa Crenshaw, Dennis W. Bartholomew, Lina Basel, Gary Bellus, Shay Ben-Shachar, Martin G. Bialer, David Bick, Bruce Blumberg, Fanny Cortes, Karen L. David, Anne Destree, Anna Duat-Rodriguez, Dawn Earl, Luis Escobar, Marthanda Eswara, Begona Ezquieta, Ian M. Frayling, Moshe Frydman, Kathy Gardner, Karen W. Gripp, Concepcion Hernández-Chico, Kurt Heyrman, Jennifer Ibrahim, Sandra Janssens, Beth A. Keena, Isabel Llano-Rivas, Kathy Leppig, Marie McDonald, Vinod K. Misra, Jennifer Mulbury, Vinodh Narayanan, Naama Orenstein, Patricia Galvin-Parton, Helio Pedro, Eniko K. Pivnick, Cynthia M. Powell, Linda Randolph, Salmo Raskin, Jordi Rosell, Karol Rubin, Margretta Seashore, Christian P. Schaaf, Angela Scheuerle, Meredith Schultz, Elizabeth Schorry, Rhonda Schnur, Elizabeth Siqveland, Amanda Tkachuk, James Tonsgard, Meena Upadhyaya, Ishwar C. Verma, Stephanie Wallace, Charles Williams, Elaine Zackai, Jonathan Zonana, Conxi Lazaro, Kathleen Claes, Bruce Korf, Yolanda Martin, Eric Legius, and Ludwine Messiaen. High incidence of noonan syndrome features including short stature and pulmonic stenosis in patients carrying nf1 missense mutations affecting p.arg1809: Genotype-phenotype correlation. *Human mutation*, 36:1052–1063, November 2015.

[42] M. Upadhyaya, S. M. Huson, M. Davies, N. Thomas, N. Chuzhanova, S. Giovannini, D. G. Evans, E. Howard, B. Kerr, S. Griffiths, C. Consoli, L. Side, D. Adams, M. Pierpont, R. Hachen, A. Barnicoat, H. Li, P. Wallace, J. P. Van Biervliet, D. Stevenson, D. Viskochil, D. Baralle, E. Haan, V. Riccardi, P. Turnpenny, C. Lazaro, and L. Messiaen. An absence of cutaneous neurofibromas associated with a 3-bp inframe deletion in exon 17 of the nf1 gene (c.2970-2972 delaat): evidence of a clinically significant nf1 genotype-phenotype correlation. *American journal of human genetics*, 80:140–151, January 2007.

[43] Eric Pasmant, Audrey Sabbagh, Gill Spurlock, Ingrid Laurendeau, Elisa Grillo, Marie-José Hamel, Ludovic Martin, Sébastien Barbarot, Bruno Leheup, Diana Rodriguez, Didier Lacombe, Hélène Dollfus, Laurent Pasquier, Bertrand Isidor, Salah Ferkal, Jean Soulier, Marc Sanson, Anne Dieux-Coeslier, Ivan Bièche, Béatrice Parfait, Michel Vidaud, Pierre Wolkenstein, Meena Upadhyaya, Dominique Vidaud, and members of the NF France Network. Nf1 microdeletions in neurofibromatosis type 1: from genotype to phenotype. *Human mutation*, 31:E1506–E1518, June 2010.

[44] Dóra Nagy, Sarah Verheyen, Kristen M. Wigby, Artem Borovikov, Artem Sharkov, Valerie Slegesky, Austin Larson, Christina Fagerberg, Charlotte Brasch-Andersen, Maria Kibæk, Ingrid Bader, Rebecca Hernan, Frances A. High, Wendy K. Chung, Jolanda H. Schieving, Jana Behunova, Mateja Smogavec, Franco Laccone, Martina Witsch-Baumgartner, Joachim Zobel, Hans-Christoph Duba, and Denisa Weis. Genotype-phenotype comparison in pogz-related neurodevelopmental disorders by using clinical scoring. *Genes*, 13, January 2022.

[45] Valerie K. Jordan, Brieana Fregeau, Xiaoyan Ge, Jessica Giordano, Ronald J. Wapner, Tugce B. Balci, Melissa T. Carter, John A. Bernat, Amanda N. Moccia, Anshika Srivastava, Donna M. Martin, Stephanie L. Bielas, John Pappas, Melissa D. Svoboda, Marlène Rio, Nathalie Boddaert, Vincent Cantagrel, Andrea M. Lewis, Fernando Scaglia, Undiagnosed Diseases Network, Jennefer N. Kohler, Jonathan A. Bernstein, Annika M. Dries, Jill A. Rosenfeld, Colette DeFilippo, Willa Thorson, Yaping Yang, Elliott H. Sherr, Weimin Bi, and Daryl A. Scott. Genotype-phenotype correlations in individuals with pathogenic rere variants. *Human mutation*, 39:666–675, May 2018.

[46] Malena Daich Varela, Mrunmayi Jeste, Thales A. C. de Guimaraes, Omar A. Mahroo, Gavin Arno, Andrew R. Webster, and Michel Michaelides. Clinical, ophthalmic, and genetic characterization of rpgrip1-associated leber congenital amaurosis/early-onset severe retinal dystrophy. *American journal of ophthalmology*, 266:255–263, October 2024.

[47] Carla D. Zingariello, Dong-Hui Chen, Wendy H. Raskind, William B. Slayton, Sub Subramony, Joyce Severance, Megan Feagle, and Sonja A. Rasmussen. Assessing long-term neurologic outcomes in samd9l-related ataxia-pancytopenia syndrome. *Movement disorders clinical practice*, 11:728–733, June 2024.

[48] Markus Wolff, Andreas Brunklaus, and Sameer M. Zuberi. Phenotypic spectrum and genetics of scn2a-related disorders, treatment options, and outcomes in epilepsy and beyond. *Epilepsia*, 60 Suppl 3:S59–S67, December 2019.

[49] K. Vesela, H. Hansikova, M. Tesarova, P. Martasek, M. Elleder, J. Houstek, and J. Zeman. Clinical, biochemical and molecular analyses of six patients with isolated cytochrome c oxidase deficiency due to mutations in the sco2 gene. *Acta paediatrica (Oslo, Norway : 1992)*, 93:1312–1317, October 2004.

[50] Erika Van Nieuwenhove, John S Barber, Julika Neumann, Elien Smeets, Mathijs Willemsen, Emanuela Pasciuto, Teresa Prezzemolo, Vasiliki Lagou, Laura Seldeslachts, Bert Malengier-Devlies, Mieke Metzemaekers, Sarah Haßdenteufel, Axelle Kerstens, Rob van der Kant, Frederic Rousseau, Joost Schymkowitz, Daniele Di Marino, Sven Lang, Richard Zimmermann, Susan Schlenner, Sebastian Munck, Paul Proost, Patrick Matthys, Christine Devalck, Nancy Boeckx, Frank Claessens, Carine Wouters, Stephanie Humblet-Baron, Isabelle Meyts, and Adrian Liston. Defective Sec61$\alpha$1 underlies a novel cause of autosomal dominant severe congenital neutropenia. *J. Allergy Clin. Immunol.*, 146(5):1180–1193, November 2020.

[56] Elisabeth Bosch, Bernt Popp, Esther Güse, Cindy Skinner, Pleuntje J van der Sluijs, Isabelle Maystadt, Anna Maria Pinto, Alessandra Renieri, Lucia Pia Bruno, Stefania Granata, Carlo Marcelis, Özlem Baysal, Dewi Hartwich, Laura Holthöfer, Bertrand Isidor, Benjamin Cogne, Dagmar Wieczorek, Valeria Capra, Marcello Scala, Patrizia De Marco, Marzia Ognibene, Rami Abou Jamra, Konrad Platzer, Lauren B Carter, Outi Kuismin, Arie van Haeringen, Reza Maroofian, Irene Valenzuela, Ivon Cuscó, Julian A Martinez-Agosto, Ahna M Rabani, Heather C Mefford, Elaine M Pereira, Charlotte Close, Kwame Anyane-Yeboa, Mallory Wagner, Mark C Hannibal, Pia Zacher, Isabelle Thiffault, Gea Beunders, Muhammad Umair, Priya T Bhola, Erin McGinnis, John Millichap, Jiddeke M van de Kamp, Eloise J Prijoles, Amy Dobson, Amelle Shillington, Brett H Graham, Evan-Jacob Garcia, Maureen Kelly Galindo, Fabienne G Ropers, Esther A R Nibbeling, Gail Hubbard, Catherine Karimov, Guido Goj, Renee Bend, Julie Rath, Michelle M Morrow, Francisca Millan, Vincenzo Salpietro, Annalaura Torella, Vincenzo Nigro, Mitja Kurki, Roger E Stevenson, Gijs W E Santen, Markus Zweier, Philippe M Campeau, Mariasavina Severino, André Reis, Andrea Accogli, and Georgia Vasileiou. Elucidating the clinical and molecular spectrum of SMARCC2-associated NDD in a cohort of 65 affected individuals. *Genet. Med.*, 25(11):100950, November 2023.

[57] Jun Tohyama, Mitsuko Nakashima, Shin Nabatame, Ch'ng Gaik-Siew, Rie Miyata, Zvonka Rener-Primec, Mitsuhiro Kato, Naomichi Matsumoto, and Hirotomo Saitsu. Sptan1 encephalopathy: distinct phenotypes and genotypes. *Journal of human genetics*, 60:167–173, April 2015.

[58] Heba Morsy, Mehdi Benkirane, Elisa Cali, Clarissa Rocca, Kristina Zhelcheska, Valentina Cipriani, Evangelia Galanaki, Reza Maroofian, Stephanie Efthymiou, David Murphy, Mary O'Driscoll, Mohnish Suri, Siddharth Banka, Jill Clayton-Smith, Thomas Wright, Melody Redman, Jennifer A. Bassetti, Mathilde Nizon, Benjamin Cogne, Rami Abu Jamra, Tobias Bartolomaeus, Marion Heruth, Ilona Krey, Janina Gburek-Augustat, Dagmar Wieczorek, Felix Gattermann, Meriel Mcentagart, Alice Goldenberg, Lucie Guyant-Marechal, Hector Garcia-Moreno, Paola Giunti, Brigitte Chabrol, Severine Bacrot, Roger Buissonnière, Virginie Magry, Vykuntaraju K. Gowda, Varunvenkat M. Srinivasan, Béla Melegh, András Szabó, Katalin Sümegi, Mireille Cossée, Monica Ziff, Russell Butterfield, David Hunt, Georgina Bird-Lieberman, Michael Hanna, Michel Koenig, Michael Stankewich, Jana Vandrovcova, Henry Houlden, and Genomics England Research Consortium. Expanding sptan1 monoallelic variant associated disorders: From epileptic encephalopathy to pure spastic paraplegia and ataxia. *Genetics in medicine : official journal of the American College of Medical Genetics*, 25:76–89, January 2023.

[59] Liedewei Van de Vondel, Jonathan De Winter, Danique Beijer, Giulia Coarelli, Melanie Wayand, Robin Palvadeau, Martje G. Pauly, Katrin Klein, Maren Rautenberg, Léna Guillot-Noël, Tine Deconinck, Atay Vural, Sibel Ertan, Okan Dogu, Hilmi Uysal, Vesna Brankovic, Rebecca Herzog, Alexis Brice, Alexandra Durr, Stephan Klebe, Friedrich Stock, Almut Turid Bischoff, Tim W. Rattay, María-Jesús Sobrido, Giovanna De Michele, Peter De Jonghe, Thomas Klopstock, Katja Lohmann, Ginevra Zanni, Filippo M. Santorelli, Vincent Timmerman, Tobias B. Haack, Stephan Züchner, P. R. E. P. A. R. E. Consortium, Rebecca Schüle, Giovanni Stevanin, Matthis Synofzik, A. Nazli Basak, and Jonathan Baets. De novo and dominantly inherited sptan1 mutations cause spastic paraplegia and cerebellar ataxia. *Movement disorders : official journal of the Movement Disorder Society*, 37:1175–1186, June 2022.

[66] Juliette Piard, Lara Hawkes, Mathieu Milh, Laurent Villard, Renato Borgatti, Romina Romaniello, Melanie Fradin, Yline Capri, Delphine Héron, Marie-Christine Nougues, Caroline Nava, Oana Tarta Arsene, Debbie Shears, John Taylor, Alistair Pagnamenta, Jenny C. Taylor, Yoshimi Sogawa, Diana Johnson, Helen Firth, Pradeep Vasudevan, Gabriela Jones, Marie-Ange Nguyen-Morel, Tiffany Busa, Agathe Roubertie, Myrthe van den Born, Elise Brischoux-Boucher, Michel Koenig, Cyril Mignot, D. D. D. Study, Usha Kini, and Christophe Philippe. The phenotypic spectrum of wwox-related disorders: 20 additional cases of woree syndrome and review of the literature. *Genetics in medicine : official journal of the American College of Medical Genetics*, 21:1308–1318, June 2019.

[67] Mylène Valduga, Christophe Philippe, Laetitia Lambert, Pascale Bach-Segura, Emmanuelle Schmitt, Jean Pierre Masutti, Bénédicte François, Patrick Pinaud, Mireille Vibert, and Philippe Jonveaux. Wwox and severe autosomal recessive epileptic encephalopathy: first case in the prenatal period. *Journal of human genetics*, 60:267–271, May 2015.

[68] Cyril Mignot, Laetitia Lambert, Laurent Pasquier, Thierry Bienvenu, Andrée Delahaye-Duriez, Boris Keren, Jérémie Lefranc, Aline Saunier, Lila Allou, Virginie Roth, Mylène Valduga, Aissa Moustaïne, Stéphane Auvin, Catherine Barrey, Sandra Chantot-Bastaraud, Nicolas Lebrun, Marie-Laure Moutard, Marie-Christine Nougues, Anne-Isabelle Vermersch, Bénédicte Héron, Eva Pipiras, Delphine Héron, Laurence Olivier-Faivre, Jean-Louis Guéant, Philippe Jonveaux, and Christophe Philippe. Wwox-related encephalopathies: delineation of the phenotypical spectrum and emerging genotype-phenotype correlation. *Journal of medical genetics*, 52:61–70, January 2015.

[69] James L. Shepherdson, Katie Hutchison, Dilan Wellalage Don, George McGillivray, Tae-Ik Choi, Carolyn A. Allan, David J. Amor, Siddharth Banka, Donald G. Basel, Laura D. Buch, Deanna Alexis Carere, Renée Carroll, Jill Clayton-Smith, Ali Crawford, Morten Dunø, Laurence Faivre, Christopher P. Gilfillan, Nina B. Gold, Karen W. Gripp, Emma Hobson, Alexander M. Holtz, A. Micheil Innes, Bertrand Isidor, Adam Jackson, Panagiotis Katsonis, Leila Amel Riazat Kesh, Genomics England Research Consortium, Sébastien Küry, François Lecoquierre, Paul Lockhart, Julien Maraval, Naomichi Matsumoto, Julie McCarrier, Josephine McCarthy, Noriko Miyake, Lip Hen Moey, Andrea H. Németh, Elsebet Østergaard, Rushina Patel, Kate Pope, Jennifer E. Posey, Rhonda E. Schnur, Marie Shaw, Elliot Stolerman, Julie P. Taylor, Erin Wadman, Emma Wakeling, Susan M. White, Lawrence C. Wong, James R. Lupski, Olivier Lichtarge, Mark A. Corbett, Jozef Gecz, Charles M. Nicolet, Peggy J. Farnham, Cheol-Hee Kim, and Marwan Shinawi. Variants in zfx are associated with an x-linked neurodevelopmental disorder with recurrent facial gestalt. *American journal of human genetics*, 111:487–508, March 2024.

[70] Bin Guan, Sunita K. Agarwal, James M. Welch, Smita Jha, Lee S. Weinstein, and William F. Simonds. A germline zfx missense variant in a patient with primary hyperparathyroidism. *JCEM case reports*, 2:luae115, August 2024.

[71] Anju K. Philips, Auli Sirén, Kristiina Avela, Mirja Somer, Maarit Peippo, Minna Ahvenainen, Fatma Doagu, Maria Arvio, Helena Kääriäinen, Hilde Van Esch, Guy Froyen, Stefan A. Haas, Hao Hu, Vera M. Kalscheuer, and Irma Järvelä. X-exome sequencing in finnish families with intellectual disability–four novel mutations and two novel syndromic phenotypes. *Orphanet journal of rare diseases*, 9:49, April 2014.

[72] Susan M. Hiatt, Slavica Trajkova, Matteo Rossi Sebastiano, E. Christopher Partridge, Fatima E. Abidi, Ashlyn Anderson, Muhammad Ansar, Stylianos E. Antonarakis, Azadeh Azadi, Ruxandra Bachmann-Gagescu, Andrea Bartuli, Caroline Benech, Jennifer L. Berkowitz, Michael J. Betti, Alfredo Brusco, Ashley Cannon, Giulia Caron, Yanmin Chen, Meagan E. Cochran, Tanner F. Coleman, Molly M. Crenshaw, Laurence Cuisset, Cynthia J. Curry, Hossein Darvish, Serwet Demirdas, Maria Descartes, Jessica Douglas, David A. Dyment, Houda Zghal Elloumi, Giuseppe Ermondi, Marie Faoucher, Emily G. Farrow, Stephanie A. Felker, Heather Fisher, Anna C. E. Hurst, Pascal Joset, Melissa A. Kelly, Stanislav Kmoch, Benjamin R. Leadem, Michael J. Lyons, Marina Macchiaiolo, Martin Magner, Giorgia Mandrile, Francesca Mattioli, Megan McEown, Sarah K. Meadows, Livija Medne, Naomi J. L. Meeks, Sarah Montgomery, Melanie P. Napier, Marvin Natowicz, Kimberly M. Newberry, Marcello Niceta, Lenka Noskova, Catherine B. Nowak, Amanda G. Noyes, Matthew Osmond, Eloise J. Prijoles, Jada Pugh, Verdiana Pullano, Chloé Quélin, Simin Rahimi-Aliabadi, Anita Rauch, Sylvia Redon, Alexandre Reymond, Caitlin R. Schwager, Elizabeth A. Sellars, Angela E. Scheuerle, Elena Shukarova-Angelovska, Cara Skraban, Elliot Stolerman, Bonnie R. Sullivan, Marco Tartaglia, Isabelle Thiffault, Kevin Uguen, Luis A. Umaña, Yolande van Bever, Saskia N. van der Crabben, Marjon A. van Slegtenhorst, Quinten Waisfisz, Camerun Washington, Lance H. Rodan, Richard M. Myers, and Gregory M. Cooper. Deleterious, protein-altering variants in the transcriptional coregulator zmym3 in 27 individuals with a neurodevelopmental delay phenotype. *American journal of human genetics*, 110:215–227, February 2023.
